# The Genetic Architecture of Biological Age in Nine Human Organ Systems

**DOI:** 10.1101/2023.06.08.23291168

**Authors:** Junhao Wen, Ye Ella Tian, Ioanna Skampardoni, Zhijian Yang, Yuhan Cui, Filippos Anagnostakis, Elizabeth Mamourian, Bingxin Zhao, Arthur W. Toga, Andrew Zaleskey, Christos Davatzikos

**Affiliations:** Laboratory of AI and Biomedical Science (LABS), Keck School of Medicine of USC, University of Southern California, Los Angeles, California, USA; Melbourne Neuropsychiatry Centre, Department of Psychiatry, Melbourne Medical School, The University of Melbourne, Melbourne, Victoria, Australia; Artificial Intelligence in Biomedical Imaging Laboratory (AIBIL), Center for AI and Data Science for Integrated Diagnostics (AI^2^D), Perelman School of Medicine, University of Pennsylvania, Philadelphia, USA; Department of Medical and Surgical Sciences, University of Bologna, 40126 Bologna, Italy; Department of Statistics and Data Science, University of Pennsylvania, Philadelphia, PA, USA; Laboratory of Neuro Imaging (LONI), Stevens Neuroimaging and Informatics Institute, Keck School of Medicine of USC, University of Southern California, Los Angeles, California, USA

## Abstract

Understanding the genetic basis of biological aging in multi-organ systems is vital for elucidating age-related disease mechanisms and identifying therapeutic interventions. This study characterized the genetic architecture of the biological age gap (BAG) across nine human organ systems in 377,028 individuals of European ancestry from the UK Biobank. We discovered 393 genomic loci-BAG pairs (P-value<5×10^-8^) linked to the brain, eye, cardiovascular, hepatic, immune, metabolic, musculoskeletal, pulmonary, and renal systems. We observed BAG-organ specificity and inter-organ connections. Genetic variants associated with the nine BAGs are predominantly specific to the respective organ system while exerting pleiotropic effects on traits linked to multiple organ systems. A gene-drug-disease network confirmed the involvement of the metabolic BAG-associated genes in drugs targeting various metabolic disorders. Genetic correlation analyses supported Cheverud’s Conjecture^1^ – the genetic correlation between BAGs mirrors their phenotypic correlation. A causal network revealed potential causal effects linking chronic diseases (e.g., Alzheimer’s disease), body weight, and sleep duration to the BAG of multiple organ systems. Our findings shed light on promising therapeutic interventions to enhance human organ health within a complex multi-organ network, including lifestyle modifications and potential drug repositioning strategies for treating chronic diseases. All results are publicly available at https://labs-laboratory.com/medicine.

## Main

Biological aging is complex and influenced by many factors, including genetics^2^, environmental exposures^3^, and modifiable lifestyle factors^4^ across multiple organ systems. A comprehensive understanding of the phenotypic landscape and genetic architecture underlying biological aging in multiple human organ systems is paramount in forging the path toward precision medicine^5^, including identifying vulnerability (e.g., smoking) and resilience factors (e.g., physical activities). This knowledge can improve our understanding of the underlying mechanisms driving age-related diseases, identify novel therapeutic targets, and develop personalized interventions for maintaining health and functional independence in the aging population.

Previous research efforts have made progress in studying the interconnectedness of multi-organ systems in human health^3,6–13^. In a recent study by McCracken et al., a heart-brain-liver axis was studied, highlighting direct and indirect associations among the three organs and their interconnectivity and shared biological pathways^11^. A recent review highlighted the role of inter-organ signals in metabolic control, including the secretion of peptides, small molecules, and lipid mediators by metabolic tissues and the involvement of the central nervous system in coordinating peripheral metabolic functions^9^. Riding the crest of the wave of artificial intelligence (AI), the biomedical community has increasingly adopted the biological age gap (BAG) as a comprehensive biomarker of human aging in multiple human organ systems.

Specifically, BAG serves as a quantitative phenotype to capture the disparity between an individual’s AI-derived age and chronological age, which can be used to model aging-related normative trajectory at the individual level and holds potential for application in disease populations to capture pertinent pathological processes. For instance, Nie et al. derived the biological age in nine organ systems to predict the possibility of becoming centenarian^13^. In our previous study, Tian et al. derived eight BAGs in eight organ systems, correlating them with cognition, chronic disease, lifestyle factors, and mortality^3^. We employed a support vector machine in cross-validation to predict BAGs for multiple organ systems (**Method 1** for details).

However, genetic determinants and biological pathways that underlie the observed heterogeneity of organ-specific BAGs remain elusive. Furthermore, whether chronic diseases and lifestyle factors causally impact the divergence between predicted age and chronological age in these organ systems remains to be established, manifesting as either a younger or older biological age. Our previous genome-wide association study (GWAS) uncovered the genetic heterogeneity of the multimodal brain BAGs using magnetic resonance imaging (MRI) data^14^.

Expanding on prior research, the current study sought to comprehensively depict the genetic architecture underlying biological aging across nine human organ systems, including the brain, cardiovascular, eye, hepatic, immune, metabolic, musculoskeletal, pulmonary, and renal BAGs. Our overarching hypothesis postulates that the genetic determinants associated with the nine BAGs are not only specific to individual organ systems (i.e., BAG-organ specificity) but also directly or indirectly interconnected with other organ systems (i.e., inter-organ connection).

In the current study, we analyzed multimodal data from 377,028 individuals of European ancestry in the UK Biobank study^15^ (UKBB) to comprehensively capture the genetic architecture of the nine organ systems (**Method 2**). First, we used data from 154,774 participants to perform GWAS, gene-level, partitioned heritability, and genetic correlation analyses (**Method 3**). In our Mendelian randomization analyses, we used 222,254 UKBB participants that did not overlap with the individuals used to compute BAG to avoid potential bias^16^. We *i*) identified both previously reported and newly identified genomic loci, *ii*) demonstrated a greater genetic heritability estimate for the brain BAG compared to other organ systems, *iii*) constructed a network linking genes, drugs, and diseases for potential drug repurposing, *iv*) confirmed that BAG-associated variants and genes exhibit BAG-organ specificity and inter-organ connection, and *v*) established both genetic correlations and causal networks among the nine BAGs, chronic diseases, and lifestyle factors. All results, including the GWAS summary statistics, are publicly accessible through the MEDICINE (**M**ulti-organ biom**EDI**cal s**CI**e**N**c**E**) knowledge portal: https://labs-laboratory.com/medicine.

## Results

### Genome-wide associations identify 393 genomic loci associated with the nine biological age gaps

In the European populations, GWAS (**Method 3a**) identified 11, 44, 17, 41, 61, 76, 24, 67, and 52 genomic loci (P-value<5×10^-8^) significantly associated with the brain, cardiovascular, eye, hepatic, immune, metabolic, musculoskeletal, pulmonary, and renal BAGs, respectively (**Fig. 1**). All details of the identified loci are presented in **Supplementary eFile 1**. Manhattan and QQ plots are presented in **Supplementary eFigures 1-9** and available in the MEDICINE knowledge portal (https://labs-laboratory.com/medicine).

**Figure 1:**
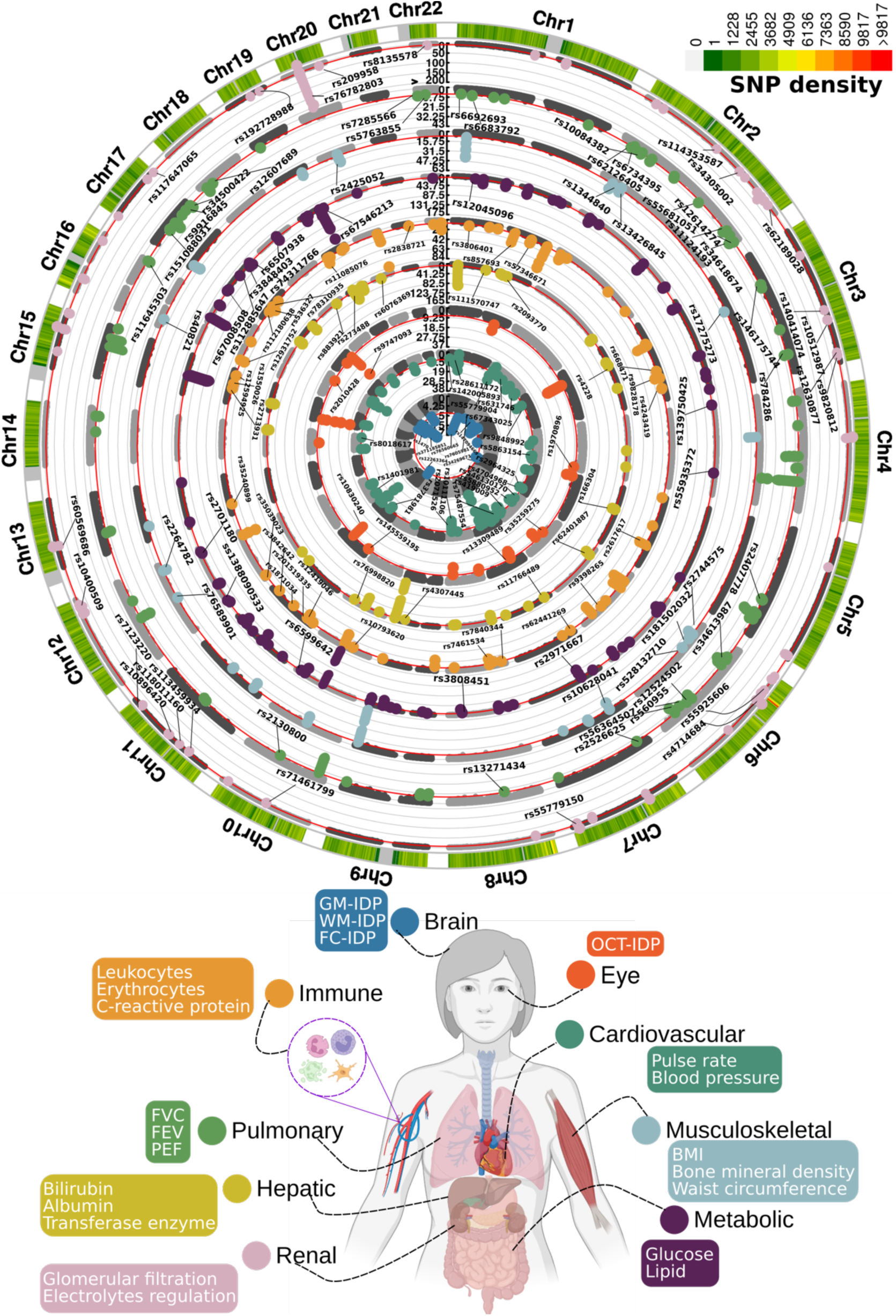
Genomic loci associated with the nine biological age gaps. Organ-specific biological age gap (BAG) was derived from a large cohort of 30,108 to 111,543 European ancestry participants from the UK Biobank cohort. The nine organ systems include the brain (*N*=30,108), cardiovascular (*N*=111,543), eye (*N*=36,004), hepatic (*N*=111,543), immune (*N*=111,543), metabolic (*N*=111,543), musculoskeletal (*N*=111,543), pulmonary (*N*=111,543), and renal (*N*=111,543) BAGs. 393 genomic loci-BAG pairs were identified using a genome-wide P-value threshold [–log_10_(P-value) > 7.30]. For visualization purposes, we denoted the genomic loci using their top lead SNPs that are not associated with any clinical traits in the EMBL-EBI GWAS Catalog. The anatomical illustration of the human body was created using <u>BioRender.com</u>. All analyses used the Genome Reference Consortium Human Build 37 (GRCh37). We present representative features employed in the calculation of each organ organ’s BAG. BMI: body mass index; IDP: imaging-derived phenotype; GM: gray matter; WM: white matter; FC: functional connectivity; OCT: optical coherence tomography; FVC: forced vital capacity; FEV: forced expiratory volume; PEF: peak expiratory flow.

We estimated the intercept of linkage disequilibrium score regression (LDSC)^17^ for the nine main GWAS and obtained intercepts of 0.9989±0.009, 1.0185±0.0099, 0.9926±0.0106, 1.0416±0.0113, 1.0293±0.0107, 1.0308±0.0124, 1.0282±0.0099, 1.0442±0.0104, and 1.0257±0.0112 for the nine BAGs. All the intercepts were close to 1, indicating no substantial genomic inflation in the primary GWAS. Furthermore, we conducted four sensitivity analyses (**Method 3a**) to assess the robustness of the primary nine GWASs on individuals of European ancestry (**Supplementary eText 1**). Our GWASs demonstrated robustness in split-sample GWAS, with a perfect concordance rate for the sign (+/-) of *β* values (*C-β*=1) between the split1 and split2 GWASs. The two sets of *β* values were highly correlated (0.90<*r-β*<0.99 for Pearson’s *r*) and did not significantly differ (*P-β*>0.48 for two-sample t-test). We compared the GWAS results with linear models in PLINK and linear mixed-effect models in fastGWA^18^, resulting in a perfect concordance for the two sets of *β* values, as well as very similar LDSC intercept values. These findings further support the absence of cryptic population stratification in our primary GWASs. Sex-stratified GWASs unveiled distinctive genetic patterns specific to each sex, with noteworthy disparities observed in the genetic architecture of the immune BAG (*r-β*=0.29; *P-β*=0.01; *C-β*=0.55). Immune responses exhibit sex differences that vary across the lifespan and are influenced by age and reproductive status^19^. Detailed quantitative information regarding these observations can be found in **Supplementary eText 1**, while visual representations of these patterns are available in **Supplementary eFigures 5 and 7**. Finally, the genetic signals identified within non-European populations were less prominent compared to the European GWAS due to the limited sample size, but we found a high concordance between the two sets of *β* values using the three proposed metrics (0.85<*r-β*<0.95; 0.89<*C-β*<1; *P-β*>0.12). This underscores the necessity of expanding sample sizes within underrepresented ethnic groups in future GWAS studies. Detailed statistics can be found in **Supplementary eFiles 2-5**.

Certain genomic loci exhibited unique associations with individual organs, whereas others displayed connections to multiple organ BAGs in close genomic proximity based on their cytogenetic position. For instance, the locus on chromosome 6 associated with the hepatic (rs62401887, position: 24416482 at 6p22.3), immune (rs80215559, position: 25918225 at 6p22.3), metabolic (rs79220007, position: 26098474 at 6p22.2), musculoskeletal (rs2744575, position: 24494975 at 6p22.3), pulmonary (rs411535, position: 22061040 at 6p22.3), and renal BAGs (rs55925606, position: 25878848 at 6p22.2) was close with each other on the human genome. Bayesian colocalization^20^ analyses (**Method 3h**) supported two distinct causal SNP within this locus with the liver and musculoskeletal BAGs. Our results showed a posterior possibility (PP) of two distinct causal variants (PP.H3.ABF=0.744) or one shared causal variant (PP.H4.ABF=0.256) associated with both traits in the *GPLD1* gene, although the PP.H4.ABF hypothesis did not achieve the suggested threshold (>0.8)^20^. Detailed results are presented in **Supplementary eFigure 10**. However, note that these loci on chromosome 6 are near the major histocompatibility complex (MHC) region; further dedicated analyses are needed to understand the underlying genetics across different BAGs (e.g., pleiotropy).

Many of these loci were mapped to protein-encoding genes and provided functional insights. For example, the top lead SNP (rs62401887 at 6p22.3) within the locus of the hepatic BAG was mapped to the *MRS2* gene by position (with a deleterious score of 14.89) and expression quantitative trait loci (eQTL, P-value=1.09×10^-10^) (**Method 3c**), which enables magnesium ion transmembrane transporter activity. We illustrate the regional Manhattan plot for the genomic locus with the highest significance for each organ BAG in **Supplementary eFigure 11**. For instance, the brain BAG exhibited a highly significant locus (top lead SNP: rs371185851 at 17q21.31) with multiple protein-encoding genes, including the widely recognized *MAPT* gene encoding tau protein associated with neurodegenerative diseases, such as Alzheimer’s disease (AD)^21^. Moreover, the SNPs within this locus included enhancers and transcription start sites specific to brain tissue chromatin states, highlighting their functional relevance in brain-related processes (**Supplementary eFigure11a**).

### Phenome-wide associations demonstrate organ system specificity and inter-organ connection

We aimed to investigate the agreement of the identified genomic loci in existing GWAS literature. To this end, we performed a phenome-wide association query in the EMBL-EBI GWAS Catalog^22^ for independent significant SNPs within each locus, considering linkage disequilibrium and redundant associations (**Method 3d**).

This pheno-wide associations query identified 11,709 significant associations between the identified loci in our GWAS and clinical traits in the literature linked to each organ system (i.e., BAG-organ specificity) (**Fig. 2a**). The genomic loci associated with the brain BAG exhibited the highest proportion of associations (74 out of 173) with traits related to the brain, including imaging-derived phenotypes such as brain volume metrics and white matter microstructure, demonstrated in the keyword cloud presented in **Fig. 2a**. The brain BAG loci were also largely linked to many other traits related to other organ systems and chronic diseases, evidencing inter-organ connections, including metabolic (*N*=43/173, e.g., cholesterol levels), lifestyle factor (*N*=1/173, i.e., alcohol consumption), neurodegenerative traits (*N*=16/173, e.g., AD), and immune (*N*=7/173, e.g., lymphocyte count). For the eye BAG loci, most associations were found in the eye (*N*=31/128, e.g., retinal nerve fiber layer thickness) and brain traits (*N*=6/128, e.g., brain morphology), among others.

**Figure 2:**
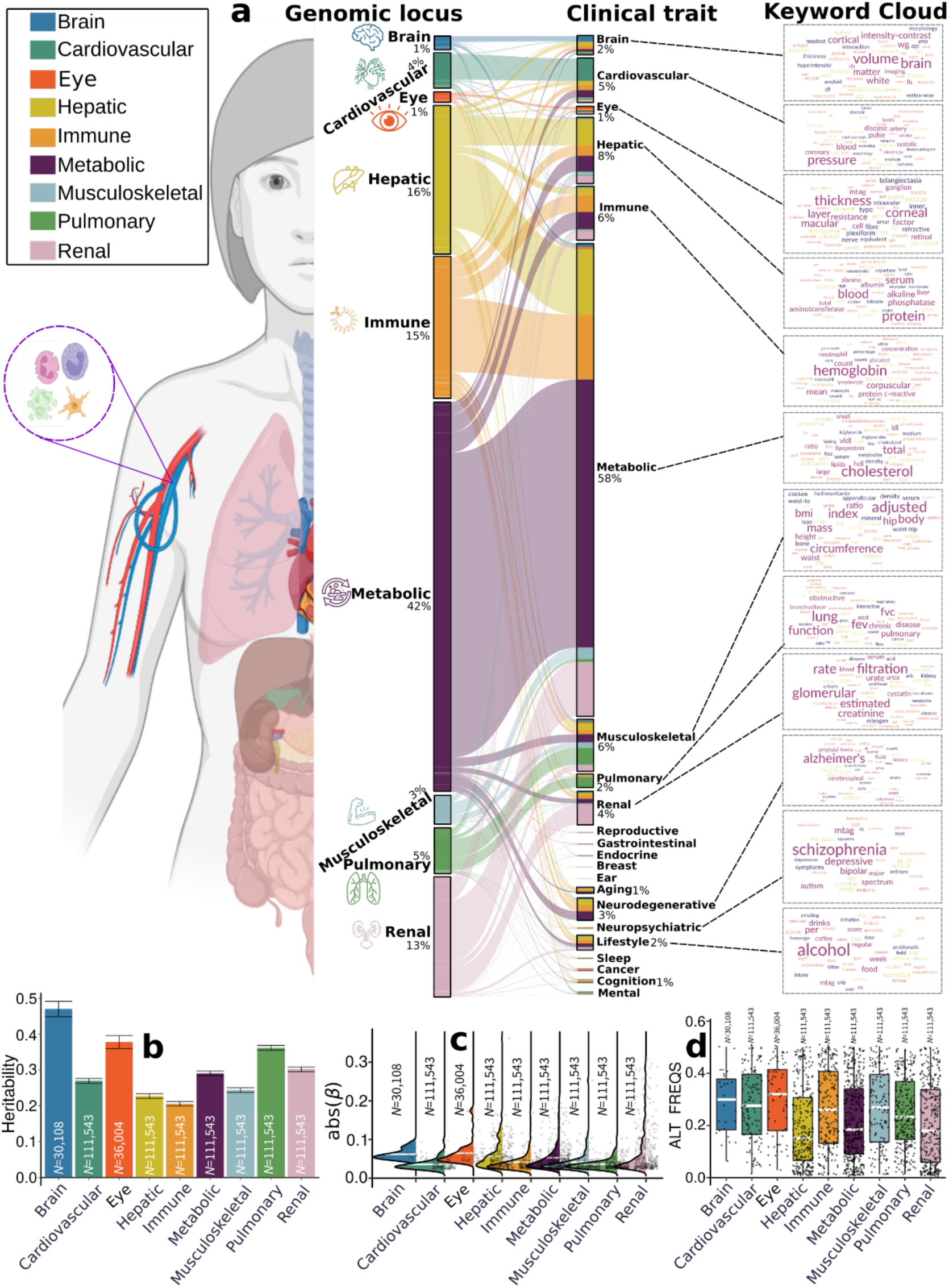
Phenome-wide associations of the identified genomic loci and SNP-wide heritability estimates of the nine biological age gap. **a**) Phenome-wide association query of the identified genomic loci in the EMBL-EBI GWAS Catalog (query date: 24^th^ April 2023, via FUMA version: v1.5.4) showed an organ-specific and inter-organ landscape. By examining the independent significant SNPs considering linkage disequilibrium (**Method 3d**) within each genomic locus, we linked them to various clinical traits.

For the seven body organ systems, among the loci associated with the cardiovascular BAG, most associations were observed with cardiovascular traits (319 out of 439), such as systolic/diastolic blood pressure and coronary artery disease. Other associations were found with musculoskeletal (*N*=30/439), metabolic (*N*=14/439), immune (*N*=6/439), renal (*N*=18/1890), and brain (*N*=9/439) traits. 376 out of 1853 associations were related to hepatic traits (e.g., blood protein, cirrhosis, and bilirubin) for the hepatic BAG loci. Among the loci associated with the immune BAG, abundant associations were found in metabolic (929 out of 1773), immune (*N*=244/1773), hepatic (*N*=149/1853), musculoskeletal (*N*=57/1853), and cardiovascular traits (*N*=72/1853). For the metabolic BAG loci, most associations were observed in metabolic traits (3841 out of 4907). We found a significant intertwining of metabolic systems with other organ systems, highlighting inter-organ connections in human metabolic activities. Details of the phenome-wide associations are presented in **Supplementary eFile 6**. Furthermore, we reported the complementary results of this phenome-wide association query using the GWAS Atalas^23^ platform (**Supplementary eText 2** and **Supplementary eFile 7**).

### The SNP-based heritability estimates of the nine biological age gaps

We estimated the SNP-based heritability (*h^2^*) across the nine organ systems using the full sample sizes (**Fig. 2b**) of the nine BAGs. Additionally, the distributions of the magnitude of the *β* coefficient in GWAS and the allele frequency of the alternative allele (effect allele) are shown in Fig. **2c** and **d**. Notably, the sample sizes of the brain and eye BAGs were much smaller than that of the seven body organ BAGs; the body organ BAGs had the same populations.

Upon analyzing the full sample sizes, the estimated *h^2^* for the brain BAG (0.47±0.02) outperformed all other organ systems, followed by the eye (0.38±0.02), pulmonary (0.36±0.006), renal (0.31±0.006), metabolic (0.29±0.006), cardiovascular (0.27±0.006), musculoskeletal (0.24±0.006), hepatic (0.23±0.006), and immune BAGs (0.21±0.006) (**Fig. 2b**). All heritability estimates were statistically significant after controlling for multiple comparisons using the Bonferroni correction. This trend persisted when subsampling the population of other BAGs to match that of the brain BAG, with comparable distributions in sex and age (**Supplementary eFigure 12a**). Detailed results of the *h^2^* estimate are presented in **Supplementary eTable 1a-b**. Of note, we employed the GCTA^24^ software to estimate *h^2^*, acknowledging that previous research^25,26^ has demonstrated variations in the magnitude of *h^2^* estimates based on the choice of methods.

To gain deeper insights into the significant genetic signals in the brain and eye, we conducted a detailed examination of the effect sizes (*β* coefficient) in the GWAS of the nine BAGs, as the effect size is independent of the sample size. The independent significant SNPs of the brain (|*β|*=0.062±0.013; [0.0470, 0.093]) and eye (|*β|*=0.0645±0.030) BAG showed larger mean magnitudes than the seven body organ systems (**Fig. 2c**). Among the body organ BAGs with the same sample size, the renal BAG showed the largest effect size (0.023<|*β|*<0.306). This pattern persisted with the results using the subsampled populations to the brain BAGs, presented in **Supplementary eFigure 12b**. The full set of statistics (e.g., *β* coefficient) of the independent significant SNPs is detailed in **Supplementary eFile 5** for the European ancestry GWAS.

It is widely recognized that the effect size of common genetic variants tends to increase as the allele frequency decreases^27,28^. This “inverse relationship” was evidenced by our data using independent significant SNPs from the 9 BAGs (**Supplementary eFigure 13**); the SNP with a lower allele frequency requires a larger sample size to achieve statistical significance. We then hypothesized that the smaller sample sizes of the brain and eye BAGs enabled us to detect significant variants with a relatively higher allele frequency but could not identify the SNPs with a relatively lower allele frequency associated with the body organ BAGs. As shown in **Fig. 2d**, we observed that the alternative (effect) allele frequency of the independent significant SNPs associated with the brain and eye BAGs was relatively higher than that of the body organ BAGs. This indicates that larger samples are required for the brain and eye to detect SNP effects with a relatively lower allele frequency. This relationship persisted by subsampling the population of other BAGs to that of the brain BAGs, which is presented in **Supplementary eFigure 12c**. As expected, the *β* coefficients derived from the whole samples (*N*>10k for body organ BAGs) were not significantly different from the results using the brain-BAG comparable down-sampled samples (*N*=30,108) (**Supplementary eTable 2**).

Another hypothesis is that the features used to compute the brain and eye BAGs – *in vivo* imaging features – are more heritable than those of the body-organ systems. We compared the genetic structure of the nine BAGs and the individual features used to compute the BAGs. This comparison is crucial for gaining insights into how the choice of predictors impacts the results of BAG GWAS, which, in turn, is fundamental for subsequent analyses related to pleiotropy and trait associations. We first estimated the SNP-based heritability for four pulmonary features and compared these with a set of multimodal brain imaging-derived phenotypes from our previous studies^14,29–32^ using the same GCTA software. We hypothesized that the brain imaging features would exhibit a higher degree of heritability than the 4 pulmonary features of the pulmonary BAG (i.e., forced vital capacity, forced expiratory volume, peak expiratory flow, and the ratio of forced expiratory volume to forced vital capacity), supported by the results in **Supplementary eTable 1c**. We then performed GWAS for the four pulmonary features within the European ancestry populations. The Manhattan and QQ plots are presented in **Supplementary eFigure 14**. The pulmonary BAG showed high genetic correlations using LDSC with the four pulmonary features (-0.79<g_c_<0.83, **Supplementary eTable 3**). Using Bayesian colocalization analysis (**Method 3h**), we identified 99 potential causal variants (PP.H4.ABF>0.80) between the pulmonary BAG and the four underlying features (**Supplementary eFile 8**). We showcased one causal variant evidenced at one locus (4q24) between the pulmonary BAG and the FEV/FCV feature (**Supplementary eFigure 15**). The PP.H4.ABF (0.99) denotes the posterior probability of hypothesis H4, which suggests that both traits share the same causal SNP (rs7664805, mapped gene: *NPNT*). SNPs in linkage disequilibrium with the causal SNP were previously linked to chronic obstructive pulmonary disease in the GWAS Catalog. To elucidate the genetic overlap at the individual SNP level, we showed the *β* coefficient of the 48 potential causal variants that passed the genome-wide significance for the pulmonary BAG and at least one pulmonary feature in **Supplementary eFigure 16**.

These traits were categorized into high-level groups encompassing different organ systems, neurodegenerative and neuropsychiatric disorders, and lifestyle factors. To visually represent the findings, we generated keyword cloud plots based on the frequency of these clinical traits within each BAG. The length of each rectangle block indicates the number of associations concerning the genomic loci in our analysis and clinical traits in the literature. The individual disease traits were categorized within their respective organ systems. However, this categorization doesn’t imply that the sum of these diseases exclusively represents the entirety of the organ system or that these diseases are solely associated with one specific organ system. Additional searches on alternative public GWAS platforms, such as the GWAS Atlas, are provided in **Supplementary eText 2**. **b**) Brain BAG is more heritable than other organ systems using GCTA^24^. **c**) Brain BAG showed larger effect sizes of the independent significant SNPs than other organ systems. The kernel density estimate plot shows the distribution of the effect sizes (i.e., the magnitude of the linear regression *β* coefficients) in the nine GWAS. The white horizontal lines represent the mean effect sizes. **d**) The distribution of the alternative allele frequency (effect allele) for the nine BAGs. Of note, only independent significant SNPs were shown for each BAG in Figures **c**-**d**. All results in Figures **b-d** used the original full sample sizes of the nine BAGs; the brain, eye, and other body organ BAGs have different sample sizes. Error bars represent the standard error of the estimated parameters. Results for Figure **b-d** using the down-sampled sample sizes (*N*=30,108 of the brain BAG) are shown in **Supplementary eFigure 12**. ALT FREQS: allele frequency of the alternative (effective) allele.

### Genes linked to the nine biological age gaps are implicated in organ system-specific biological pathways

To biologically validate our GWAS findings at the gene level, we performed gene-based associations using the MAGMA^33^ software based on the full P-value distribution from the GWAS of the nine BAGs. The significantly associated genes (**Supplementary eFile 9**) were used for the gene set enrichment analysis (GSEA, **Method 3e**) to annotate relevant biological pathways underlying each organ system (**Fig. 3a**).

**Figure 3:**
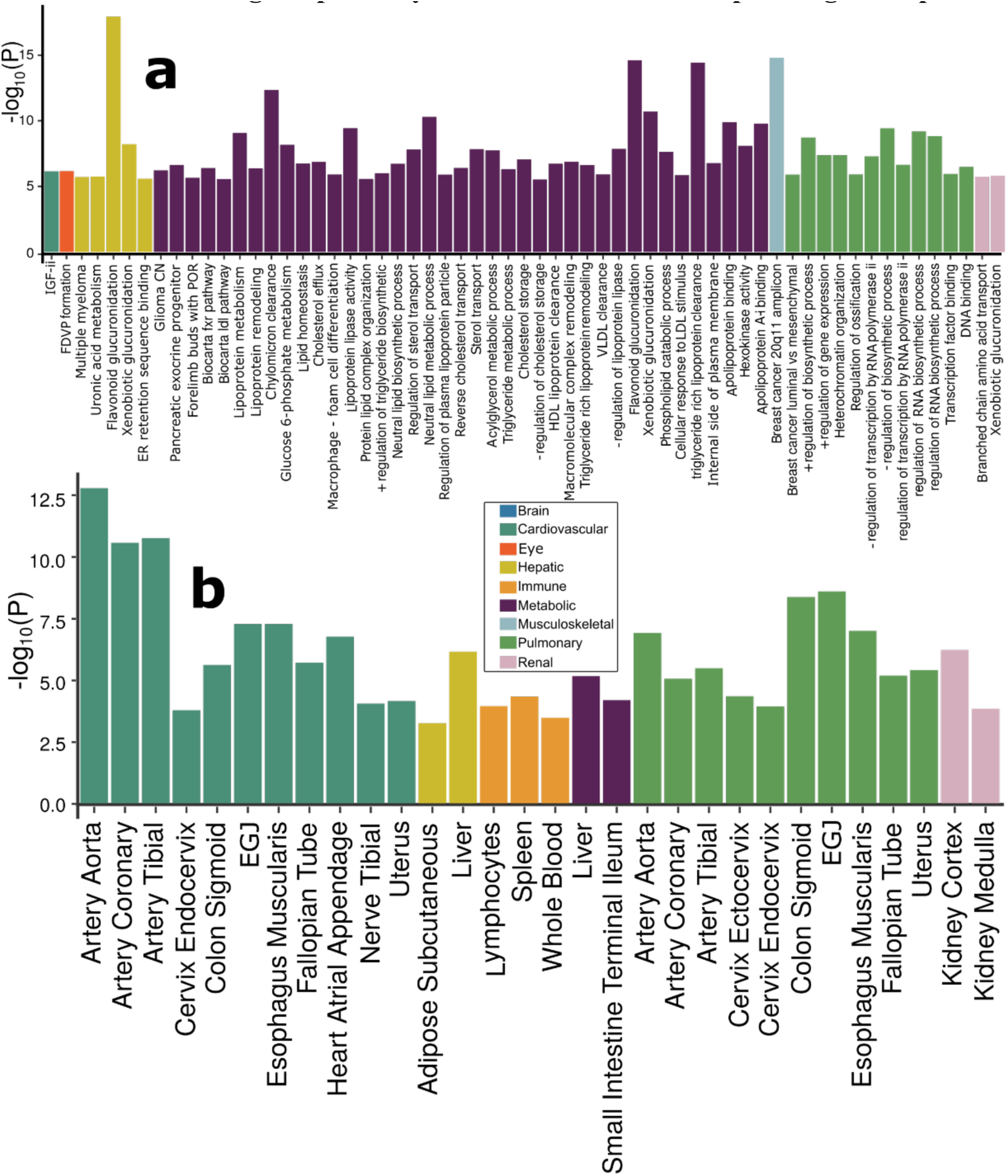
Gene-level biological pathway annotation and tissue-specific gene expression. **a**) Validation of the nine BAGs in gene set enrichment analyses. Gene set enrichment analyses were performed using curated gene sets and GO terms from the MsigDB database. **b**) Validation of the nine BAGs in gene-property analyses. Gene-property analyses evaluate tissue-specific gene expressions for the nine BAG-related genes using the full SNP P-values distribution. Only significant gene sets are presented after adjusting for multiple comparisons using the Bonferroni correction. Abbreviation: EGJ: esophagus gastroesophageal junction.

Genes associated with the cardiovascular BAG were implicated in the insulin-like growth factor II binding (IGF-II) pathway (P-value=7.08×10^-7^). Genes associated with the eye BAG were enriched in the pathway of forebrain dorsal-ventral pattern (FDVP) formation (P-value=6.46×10^-7^). Among others, the most significant enrichment result shown in the hepatic BAG was the flavonoid glucuronidation pathway (P-value=1.71×10^-8^). Genes linked to the metabolic BAG displayed enrichment in several pathways, including the flavonoid glucuronidation pathway (P-value=2.46×10^-15^) and triglyceride-rich lipoprotein particle clearance pathway (P-value=3.72×10^-15^), both of which are implicated in liver function. In addition, the neutral lipid metabolic process, regulated by complex pathways featuring lipid metabolism enzymes and structural proteins, was also identified. Genes associated with the musculoskeletal BAG exhibited enrichment in the gene set in an amplicon at 20q11 (P-value=1.54×10^-15^), defined by a study of copy number alterations conducted on 191 patients with breast tumors^34^. Genes associated with the pulmonary BAG displayed significant enrichment in the pathways of the negative regulation biosynthetic process (P-value=3.72×10^-10^), consistent with a previous DNA methylation analysis of pulmonary function using old-aged Chinese monozygotic twins^35^. Genes associated with the renal BAG were implicated in the xenobiotic glucuronidation pathway (P-value=1.56×10^-6^). Given that the kidney contains most enzymes metabolizing foreign substances (i.e., xenobiotics), it plays a crucial role in the overall metabolism of drugs and other foreign compounds within the body (**Fig. 3a**). Detailed results of GSEA are presented in **Supplementary eFile 10**. Sex-stratified results are presented in **Supplementary eFigure 17**.

### Genes linked to the nine biological age gaps display organ system-specific gene expression patterns

To investigate the gene expression patterns of the significant genes associated with the nine BAGs, we performed a tissue-specific gene expression analysis^33^ using MAGMA and the GTEx RNA-seq dataset^36^ (**Method 3f**).

Across 54 human organ tissues (**Fig. 3b**), genes associated with the cardiovascular BAG exhibited significant overexpression in various heart-related tissues (e.g., the aorta and tibial artery) and other organs (e.g., the uterus and colon sigmoid). Genes associated with the hepatic BAG were overexpressed in the liver and adipose subcutaneous. Several immune system-related tissues showed a high average expression of the genes related to the immune BAG, including the spleen, blood, and lymphocytes. Likewise, the genes associated with the metabolic BAG showed a high expression level in the liver and intestine – critical organs in the metabolic system. The genes related to the pulmonary BAG displayed significant overexpression in the esophagus gastroesophageal junction, artery, and others. The genes associated with the renal BAG were overexpressed in the kidney. Detailed results are presented in **Supplementary eFile 11**. Sex-stratified results are presented in **Supplementary eFigure 18**.

### Gene-drug-disease network substantiates potentially repositionable drugs for aging-related diseases

We performed a drug target enrichment analysis^37^ for the genes linked to the nine BAGs in the targeted gene sets of drug categories using the DrugBank database^38^, thereby constructing a gene-drug-disease network of potentially repositionable drugs (**Method 3g**).

The constructed gene-drug-disease network (**Fig. 4**) identified significant interactions between 12 metabolic BAG-linked genes, 46 drugs, and many metabolic disorders encoded in the ICD10 code (E70-E90). For instance, the *PPARD* gene was the target gene of the PPAR-δ agonist (SAR 351034, denoted in **Fig. 4**), which aimed to improve insulin sensitivity and lipid-related activities and battle against inflammation and oxidative stress, serving as actionable drugs for metabolic disorders, diabetes, and kidney and liver injury-related diseases^39^. Our results showed that genes associated with the metabolic BAG were used to develop drugs treating various other diseases – beyond metabolic disorders – related to multiple organ systems (**Fig. 4**). These included heart-related diseases (e.g., chronic rheumatic heart diseases for I05-I09) and cerebrovascular disease (I60-I69), although the enrichment did not survive correction for multiple comparisons (**Fig. 4**). For instance, the drug MPSK3169A (clinical trial number: NCT01609140; metabolic BAG linked gene: *PCSK9*) is used to treat cerebrovascular disease and coronary heart disease; T3D-959 (clinical trial number: NCT04251182; pulmonary BAG linked gene: *PPARD*), was a candidate drug targeting AD. Detailed results are presented in **Supplementary eFile 12**.

**Figure 4:**
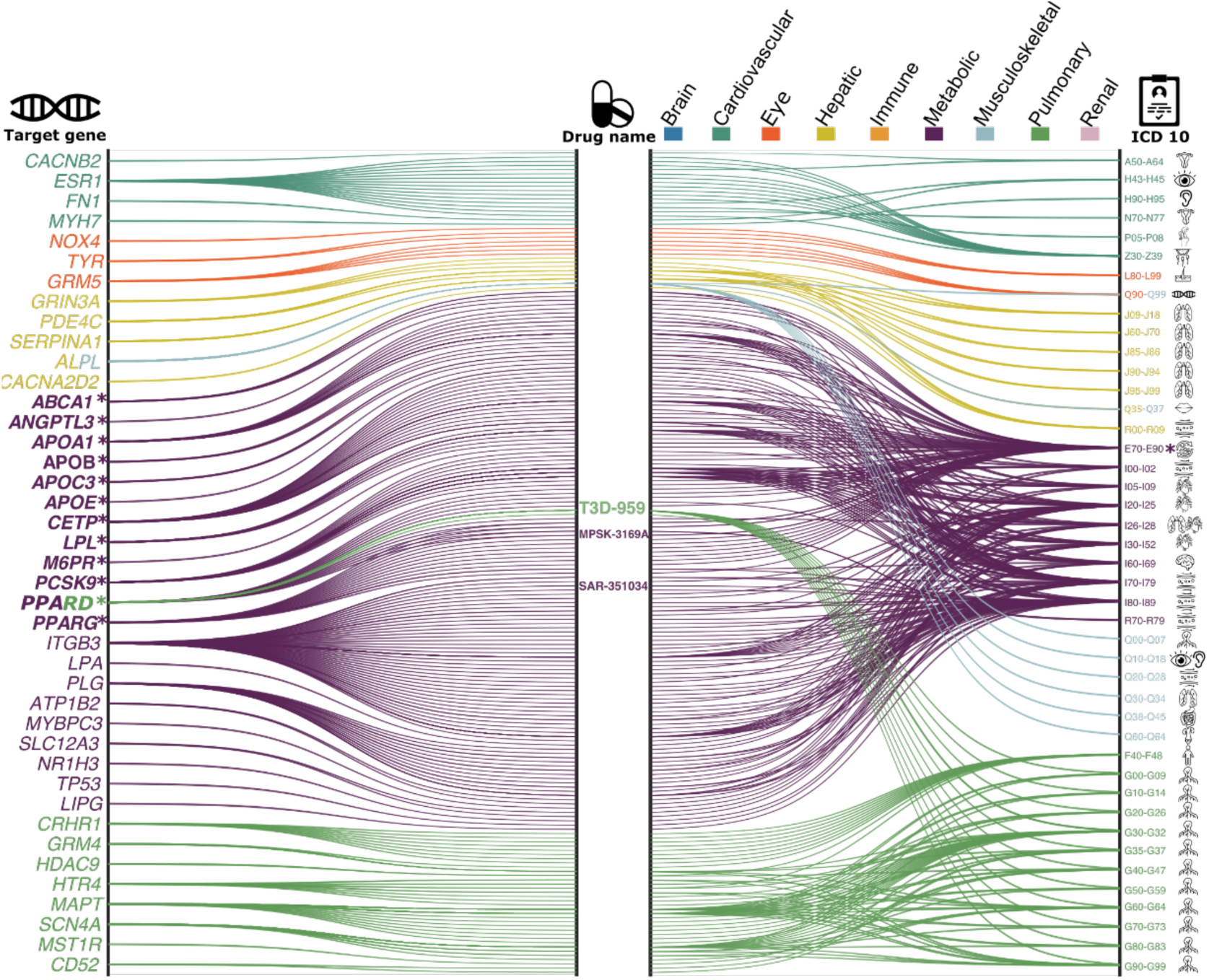
Gene-drug-disease network of the nine biological age gaps. The gene-drug-disease network reveals a broad spectrum of gene, drug, and disease interactions across the nine BAGs, highlighting the metabolic-related genes. The ICD-10 code icons symbolize disease categories linked to the primary organ systems (e.g., G30 for Alzheimer’s disease in the CNS). All presented genes passed the nominal P-value threshold (<0.05) and were pharmaco-genetically associated with drug categories in the DrugBank database; the symbol * indicates gene-drug-disease interactions that survived the Bonferroni correction. Abbreviation: ICD: International Classification of Diseases; EGJ: esophagus gastroesophageal junction.

The drug-gene-disease network reveals the association between genes related to the metabolic BAG and drugs targeting various chronic diseases. It highlights the importance of the metabolic system in the overall functioning of the human body and the potentials of repositioning existing drugs to tackle biological aging.

### Heritability enrichment in different cell types, functional categories, tissue-specific gene expression, and chromatin states

To further biologically validate our GWAS findings at the SNP level, we performed partitioned heritability analyses^40^ (**Method 3i**) to estimate the heritability enrichment of genetic variants related to the nine BAGs concerning three different cell types^41^ (i.e., neurons, oligodendrocytes, and astrocytes, **Fig. 5a**), 53 non-tissue-specific functional categories^40^ (**Fig. 5b**), 205 tissue-specific gene expression data^36^ (**Fig. 5c**) and 489 tissue-specific chromatin states^42,43^ (**Fig. 5d**).

**Figure 5:**
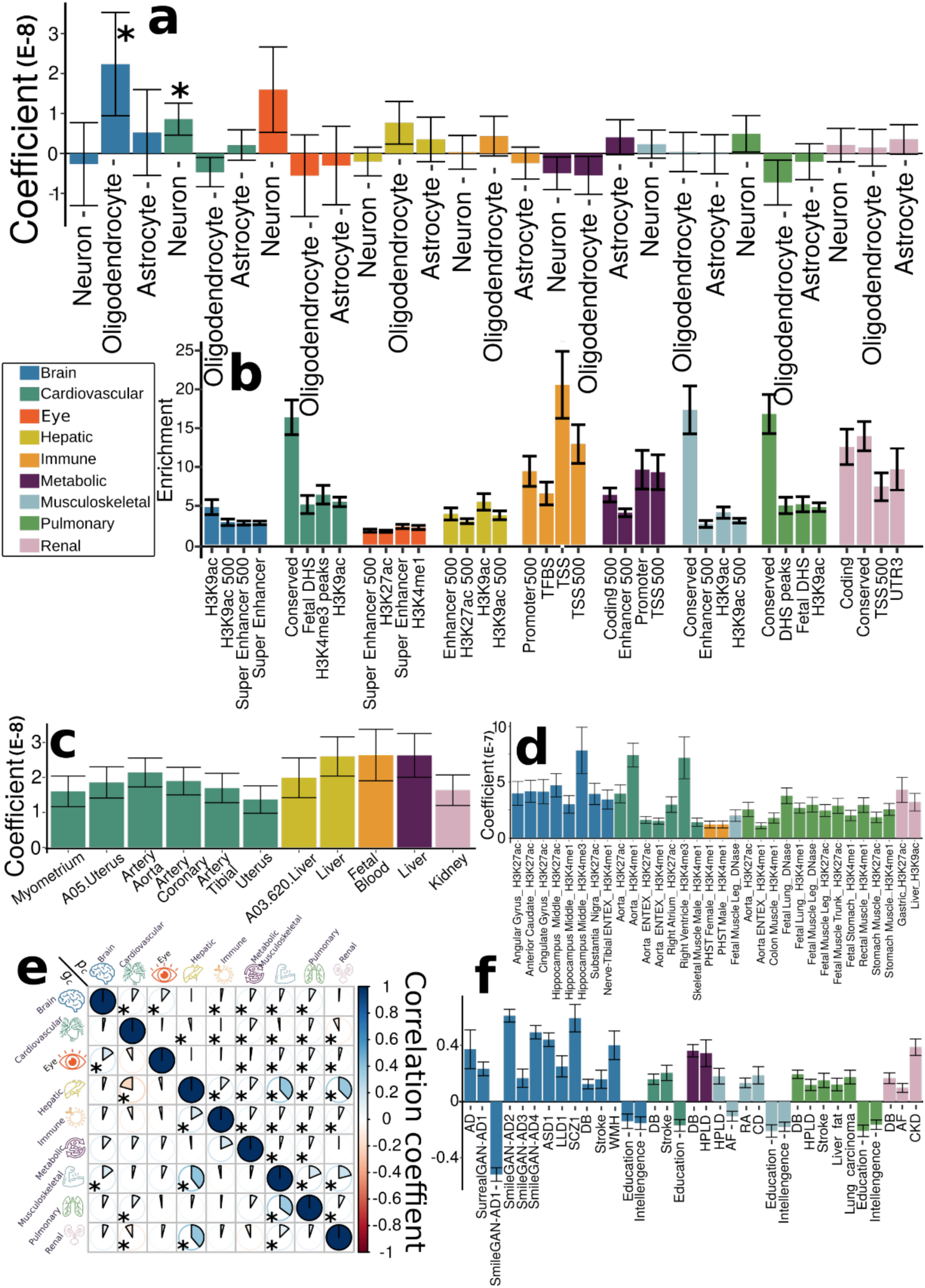
Partitioned heritability enrichment and genetic correlation of the nine biological age gaps. **a**) Cell type-specific partitioned heritability estimates for neurons, oligodendrocytes, and astrocytes. **b**) Partitioned heritability estimates for the general 53 functional categories. For visualization purposes, we only showed the four categories with the highest significant estimates for each BAG. The label for 500 denotes a 500-bp window around each of the 24 main annotations in the full baseline model, which prevents a biased estimate inflated by heritability in flanking regions^52^. **c**) Tissue-specific partitioned heritability estimates using gene sets from multi-tissue gene expression data. **d**) Tissue and chromatin-specific partitioned heritability estimates using multi-tissue chromatin data. **e**) Cheverud’s Conjecture: the genetic correlation between two BAGs (*g_c_*, lower triangle) mirrors their phenotypic correlation (*p_c_*, upper triangle). **f**) Genetic correlations between the nine BAGs and 41 clinical traits, including chronic diseases and their subtypes involving multiple human organ systems, education, intelligence, and reaction time. The symbol * denotes Bonferroni-corrected significance; the absence of * indicates all results remain significant after correction. The standard error of the estimated parameters is presented using error bars. Abbreviation: AD: Alzheimer’s disease; ASD: autism spectrum disorder; LLD: late-life depression; SCZ: schizophrenia; DB: type 2 diabetes; WMH: white matter hyperintensity; HPLD: hyperlipidemia; AF: atrial fibrillation; RA: rheumatoid arthritis; CD: Crohn’s disease; CKD: chronic kidney disease.

We found significant heritability enrichment in oligodendrocytes (P-value=0.03), a specific type of neuroglial cells, for the brain BAG. The cardiovascular BAG also exhibited significant heritability enrichment in neurons (P-value=0.01) (**Fig. 5a**, **Supplementary eFile 13**). Concerning the heritability enrichment in non-tissue-specific functional categories, we exemplified the four highest significant partitioned heritability estimates for each BAG in **Fig. 5b**. For the brain BAG, the super-enhancer regions employed 17.16% of SNPs to explain 0.47±0.04 of SNP heritability (P-value=1.80×10^-11^), and the histone H3 at lysine 9

(H3K9ac) regions used 12.61% of SNPs to explain 0.61±0.12 of SNP heritability (P-value=2.96×10^-4^). For the eye BAG, the super-enhancer regions explained 0.39±0.05 of SNP heritability (P-value=2.12×10^-6^) using 16.84% of SNPs. For the hepatic BAG, the H3K9ac regions explained 0.69±0.13 of SNP heritability (P-value=3.60×10^-5^) using 12.61% of SNPs. For the immune BAG, the TSS regions (i.e., core promoters) explained 0.37±0.08 of SNP heritability (P-value=1.48×10^-6^) using 1.82% of SNPs. The 3.11% of SNPs annotated by the promoter regions explained 0.30±0.08 of SNP heritability (P-value=7.64×10^-4^) for the metabolic BAG. For the cardiovascular (enrichment=16.39±2.23; P-value=4.70×10^-11^), musculoskeletal (enrichment=17.34±4.08; P-value=1.65×10^-6^), pulmonary (enrichment=16.82±2.51; P-value=7.58×10^-9^), and renal (enrichment=13.96±1.88; P-value=7.25×10^-9^) BAGs, the highest heritability enrichment was found in the regions conserved across mammals (**Fig. 5b**, **Supplementary eFile 14**). These results suggested disproportionate genomic contributions to the heritability of BAGs from multiple functional categories.

In addition, the nine BAGs showed high heritability enrichment in specific tissues corresponding to their organ systems. For example, the cardiovascular BAG showed significant heritability enrichment in multiple tissue types, including the artery (e.g., the aorta: P-value=1.03×10^-7^), myometrium (P-value=1.35×10^-4^), and uterus (P-value=2.43×10^-4^). Significant heritability enrichment was found in the liver for the hepatic (P-value=5.60×10^-9^) and metabolic BAGs (P-value=6.24×10^-9^). For the immune BAG, significant heritability enrichment was found in fetal blood tissues (P-value=7.36×10^-9^) (**Fig. 5c**, **Supplementary eFile 15**). These findings were aligned with the tissue-specific gene expression patterns observed at the gene level (**Fig. 3b**).

The results from multi-tissue chromatin states-specific data further provide the proof-of-concept for the organ-specific heritability enrichment among these nine BAGs. For the brain BAG, significant heritability enrichment was found in multiple brain tissues in the H3K4me3 (e.g., P-value=9.06×10^-5^ for the hippocampus), H3K4me1 (e.g., P-value=6.94×10^-5^ for the hippocampus), and H3K27ac (e.g., P-value=1.15×10^-5^ for the anterior caudate) regions. For the cardiovascular BAG, significant heritability enrichment was shown in the right ventricle in the H3K4me3 region (P-value=6.36×10^-5^) and the artery aorta in the H3K27ac region (P-value=5.81×10^-7^). Significant heritability enrichment was found in primary hematopoietic stem cells in the H3K4me1 region for the immune BAG for both females (P-value=5.61×10^-5^) and males (P-value=9.50×10^-5^). The fetal leg muscle tissue in the DNase regions (P-value=6.54×10^-5^) for the musculoskeletal BAG showed significant heritability enrichment. For the pulmonary BAG, significant heritability enrichment was found in the fetal lung in the H3K4me1 (P-value=1.33×10^-9^) and DNase regions (P-value=3.80×10^-8^), among other tissues from the stomach, artery, and muscle. For the renal BAG, significant enrichment was shown in the liver in the H3K9ac region (P-value=2.46×10^-5^) and the gastric tissues in the H3K27ac region (P-value=6.24×10^-5^) (**Fig. 5d, Supplementary eFile 16**).

### Cheverud’s Conjecture: genetic correlations between the nine biological age gaps mirror their phenotypic correlations

We estimated the genetic correlation (*g_c_*) (**Method 3h**) and the phenotypic correlation (*p_c_* for Pearson’s correlation coefficient) between each pair of the nine BAGs. Our results supported the long-standing Cheverud’s Conjecture^1^ – the genetic correlation between two clinical traits reflects their phenotypic correlation (**Fig. 5e**).

The musculoskeletal and hepatic BAGs showed the highest genetic correlation (*g_c_*=0.40) and phenotypic correlation (*p_c_*=0.38). Similarly, the hepatic and renal BAGs showed a high genetic correlation (*g_c_*=0.39) and phenotypic correlation (*p_c_*=0.37). The musculoskeletal BAG also showed significant genetic and phenotypic correlations with pulmonary (*g_c_*=0.35, *p_c_* =0.19) and renal BAGs (*g_c_*=0.13, *p_c_* =0.21). In addition, the eye BAG showed small genetic and phenotypic correlations with the brain BAG (*g_c_*=0.15, *p_c_* =0.11). The correlations between the brain and eye BAGs and other organ BAGs were relatively weaker than those observed among other organ pairs. These findings indicate the presence of shared genetic underpinnings that collectively contribute to the biological aging processes captured by these organ BAGs. Most of the genetic correlations showed consistency between females and males, albeit sex differences were evident in certain BAGs, particularly in the cardiovascular BAG results. Specifically, males exhibited dominant correlations between cardiovascular BAGs and hepatic and renal BAGs, while females demonstrated unique correlations with musculoskeletal and pulmonary BAGs (**Supplementary eFigure 19**). Sex differences in cardiovascular diseases have been explored in prior literature^44^, highlighting the divergent effects of factors associated with both sex and gender on the clinical presentations and outcomes of cardiovascular disease. Detailed results are presented in **Supplementary eFile 17**.

### Genetic correlations between the nine biological age gaps and 41 clinical traits of chronic diseases, cognition, and lifestyle factors

We also estimated *g_c_* between the nine BAGs and 41 clinical traits to examine their genetic correlations. The 41 clinical traits encompassed many common chronic diseases and conditions and their disease subtypes^7,45–48^, cognition (e.g., general intelligence and reaction time, and lifestyle factors (e.g., computer use) (**Fig. 5f** and **Supplementary eTable 4**).

The brain BAG was genetically associated with several brain diseases of the central nervous system (CNS) and their subtypes, including AD (*g_c_*=0.37±0.14) and late-life depression (LLD, *g_c_*=0.25±0.07). Furthermore, we observed significant genetic correlations between the brain BAG and years of education (*g_c_*=-0.14±0.05) and intelligence (*g_c_* =-0.15±0.05). The cardiovascular BAG was positively correlated with stroke (*g_c_*=0.20±0.05), a significant cardiovascular disease, and was negatively correlated with years of education (*g_c_*=-0.17±0.05). The musculoskeletal BAG was positively correlated with hyperlipidemia (*g_c_*=0.18±0.06), rheumatoid arthritis (*g_c_*=0.13±0.03), and Crohn’s disease (*g_c_*=0.19±0.06) and was negatively correlated with atrial fibrillation (*g_c_*=-0.11±0.04), years of education (*g_c_*=-0.21±0.04), and intelligence (*g_c_*=-0.18±0.03). The pulmonary BAG was positively associated with hyperlipidemia (*g_c_*=0.12±0.04), stroke (*g_c_*=0.15±0.05), liver fat (*g_c_*=0.12±0.04), and lung carcinoma (*g_c_*=0.17±0.05). Finally, the renal BAG was positively correlated with chronic kidney disease (*g_c_*=0.39±0.06) and atrial fibrillation (*g_c_*=0.09±0.03). Notably, type 2 diabetes showed abundant positive genetic correlations with multiple BAGs, including the brain, cardiovascular, metabolic, pulmonary, and renal. Detailed results are presented in **Supplementary eFile 18**.

Furthermore, we calculated the genetic correlation between the nine BAGs and longevity^49^ and household income^50^. Our findings indicated that the cardiovascular (*g_c_*=-0.16±0.09) and pulmonary BAG (*g_c_*=-0.12±0.07) exhibited negative associations with longevity, defined as cases surviving at or beyond the age corresponding to the 90th survival percentile; the brain BAG (*g_c_*=-0.21±0.04), musculoskeletal (*g_c_*=-0.29±0.03), and pulmonary BAG (*g_c_*=-0.16±0.03) were negatively genetically correlated with household income. We used GWAS summary statistics from a prior study^51^ to detect a significant genetic correlation between the immune BAG (*g_c_*=-0.13±0.03), pulmonary BAG (*g_c_*=-0.09±0.03), and telomere length (**Supplementary eTable 5**).

These genetic correlations yield insights into potential shared mechanisms underlying the nine BAGs, their relationships with chronic diseases, particularly AD and type 2 diabetes, and cognition. These compelling results prompted us to explore the potential causal effects of these traits on the nine BAGs. In the subsequent section, we unbiasedly selected 17 clinical traits encompassing chronic diseases, cognition, and lifestyle factors to perform Mendelian randomization (**Method 3g**).

### Hepatic and musculoskeletal biological age gaps are causally associated with each other

We performed two-sample bi-directional Mendelian randomization for each pair of BAGs by excluding overlapping populations to avoid bias^16^ (**Method 3j**). We found that the hepatic and musculoskeletal BAGs showed a bi-directional causal relationship [from the hepatic BAG to the musculoskeletal BAG: P-value=9.85×10^-7^, OR (95% CI) = 1.47 (1.26, 1.71); from the musculoskeletal BAG to the hepatic BAG: P-value=1.54×10^-8^, OR (95% CI) = 2.78 (1.95, 3.97)] (**Fig. 6**). This causal relationship echoes our genetic correlation results: the musculoskeletal and hepatic BAGs showed the highest genetic correlation compared to other organ systems (**Fig. 5e**). Detailed results and sensitivity check results are presented in **Supplementary eFile 19** and **Supplementary eFigure 20 and 21**.

**Figure 6:**
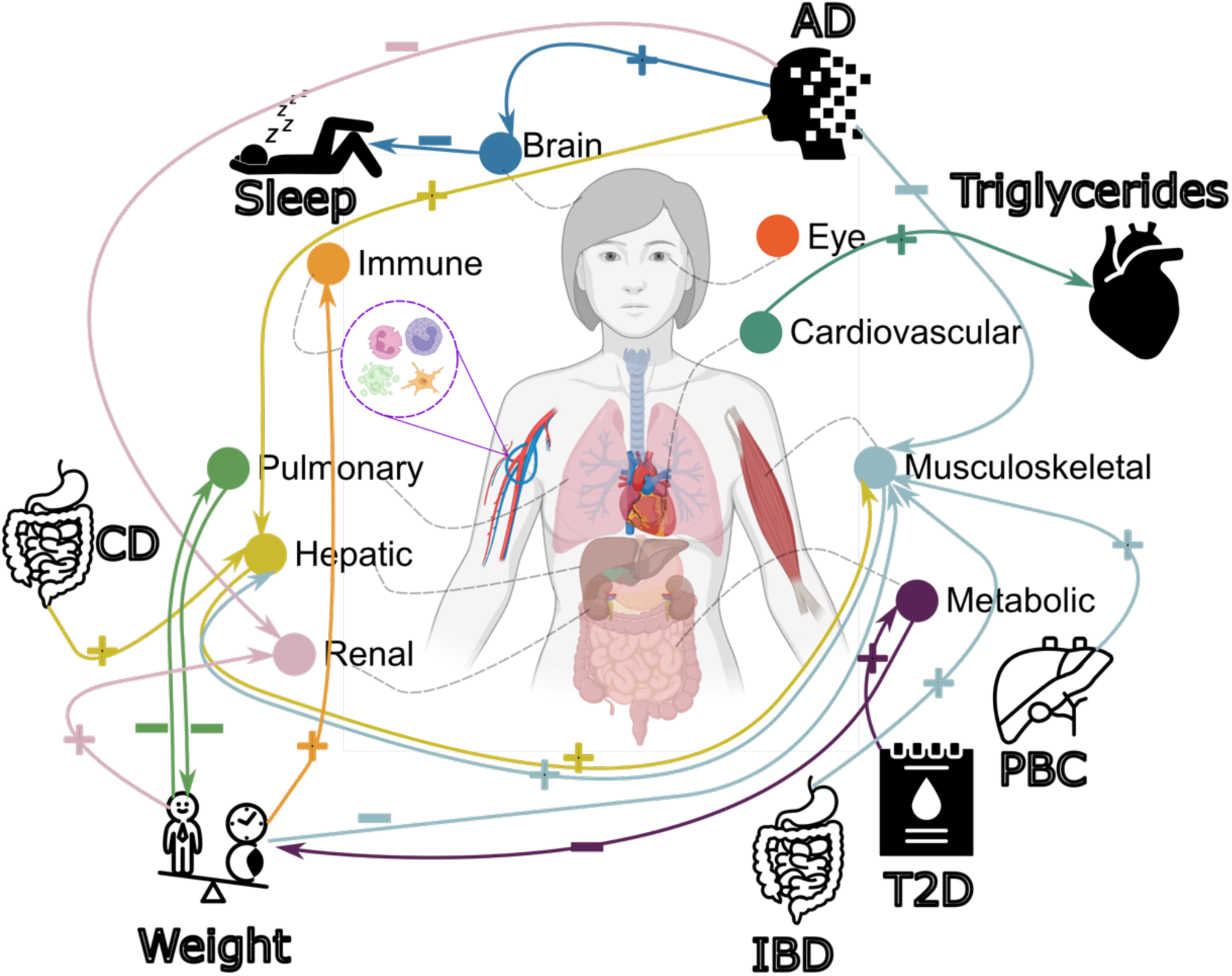
Causal multi-organ network between the 9 biological age gaps and 17 clinical traits of chronic diseases, lifestyle factors, and cognition. We conducted two sets of Mendelian randomization analyses. Firstly, we examined the causal relationships between each pair of BAGs, excluding overlapping populations. Secondly, we investigated the causal associations between the 9 BAGs and the 17 unbiasedly selected clinical traits. Bi-directional analyses, including forward and inverse analyses on the exposure and outcome variables, were performed in all experiments. Significant tests were adjusted for multiple comparisons using the Bonferroni correction. Each colored arrow represents a potential causal effect connecting the exposure variable to the outcome variable. The symbol “+” denotes an OR larger than 1, while “-” represents an OR smaller than 1. Detailed OR and 95%CI information can be found in **Supplementary eFigure 38 and eFile 19-20**. It’s crucial to approach the interpretation of these potential causal relationships with caution despite our thorough efforts in conducting multiple sensitivity checks to assess any potential violations of underlying assumptions. Abbreviation: AD: Alzheimer’s disease; T2D: type 2 diabetes; PBC: primary biliary cholangitis; CD: Crohn’s disease; IBD: inflammatory bowel disease; CI: confidence interval; OR: odds ratio.

We performed three additional sensitivity check analyses for this bi-directional causal relationship. First, we reperformed the GWAS for hepatic BAG and musculoskeletal BAG, incorporating weight as a covariate due to its established causal associations with several organ systems (**Fig. 6**). This analysis reaffirmed this bi-directional causal relationship (**Supplementary eText 3A)**. Furthermore, we performed Mendelian randomization by excluding the common SNP within the *APOE* gene (rs429358) due to its pleiotropic effects. This analysis underscored the robustness of the potential causal relationship from the hepatic BAG to the musculoskeletal BAG, both with and without including this SNP as an instrumental variable, as elaborated in **Supplementary eText 3B**. Finally, the latent causal variable (LCV^53^, **Method 3j**) model confirmed a partially genetically causal effect from the hepatic BAG to the musculoskeletal BAG [genetic causality proportion =0.75±0.14, -log_10_(P-value)=11.0, *g_c_*=0.41±0.06] (**Supplementary eTable 6**).

### Biological age gaps are causally associated with several chronic diseases, body weight, and sleep duration

We investigated the bi-directional causal effects between chronic diseases (e.g., AD) and lifestyle factors (e.g., sleep duration) and the nine BAGs. We unbiasedly and systematically included 17 clinical traits (**Method 3j**) guided by our genetic correlation results (**Fig. 5f**). The 17 clinical traits included chronic diseases linked to the brain, cardiovascular, metabolic, digestive, renal, and musculoskeletal systems, cognition, and lifestyle factors (**Supplementary eTable 7**).

In the forward Mendelian randomization, we found potential causal effects of AD on the brain [P-value=3.99×10^-8^, OR (95% CI) = 1.05 (1.03, 1.06), number of SNPs=10], hepatic [P-value=7.53×10^-7^, OR (95% CI) = 1.03 (1.02, 1.04), number of SNPs=10], musculoskeletal [P-value=1.73×10^-5^, OR (95% CI) = 0.98 (0.97, 0.99), number of SNPs=10], and renal [P-value=5.71×10^-4^, OR (95% CI) = 0.98 (0.97, 0.99), number of SNPs=10] BAGs. Body weight showed causal effects on multiple organ systems, including the immune [P-value=8.96×10^-5^, OR (95% CI) = 1.08 (1.04, 1.11), number of SNPs=160], musculoskeletal [P-value=4.32×10^-15^, OR (95% CI) = 0.83 (0.79, 0.86), number of SNPs=160], pulmonary [P-value=3.50×10^-7^, OR (95% CI) = 0.84 (0.79, 0.90), number of SNPs=160], and renal BAGs [P-value=4.53×10^-13^, OR (95% CI) = 1.18 (1.13, 1.23), number of SNPs=160]. In addition, we also found that Crohn’s disease had causal effects on the hepatic BAG [P-value=3.00×10^-3^, OR (95% CI) = 1.02 (1.00, 1.03), number of SNPs=77], type 2 diabetes on the metabolic BAG [P-value=9.92×10^-12^, OR (95% CI) =1.16 (1.09, 1.24), number of SNPs=8], inflammatory bowel disease [P-value=1.42×10^-3^, OR (95% CI) = 1.02 (1.00, 1.03), number of SNPs=80] and primary biliary cholangitis [P-value=7.41×10^-4^, OR (95% CI) = 1.02 (1.00, 1.03), number of SNPs=16] on the musculoskeletal BAG (**Fig. 6**).

For the inverse Mendelian randomization, we found potential causal effects of the metabolic [P-value=6.85×10^-4^, OR (95% CI) = 0.94 (0.91, 0.97), number of SNPs=71] and pulmonary [P-value=3.79×10^-5^, OR (95% CI) = 0.84 (0.79, 0.91), number of SNPs=62] BAGs on body weight, the cardiovascular BAG on triglycerides versus lipid ratio in very large very-low-density lipoprotein (VLDL) [P-value=2.14×10^-4^, OR (95% CI) = 1.09 (1.04, 1.14), number of SNPs=39], and the brain BAG on sleep duration [P-value=2.61×10^-3^, OR (95% CI) = 1.09 (1.04, 1.14), number of SNPs=10] (**Fig. 6**). Detailed results are presented in **Supplementary eFile 20**.

We performed several sensitivity analyses (**Method 3j**) to test the robustness of our findings. Based on these sensitivity checks, we identified potential outlier instrumental variables (IVs, i.e., SNPs) for four Mendelian randomization tests (AD and body weight on musculoskeletal BAG, Crohn’s disease on hepatic BAG, and type 2 diabetes on metabolic BAG) in the forward Mendelian randomization and one Mendelian randomization test (metabolic BAG on body weight) in the inverse Mendelian randomization. Detailed results of the sensitivity check are presented in **Supplementary eFigure 22-37** for all significant results. We showcased a detailed analysis of the sensitivity results for the metabolic BAG on body weight in **Supplementary eText 3C**. In summary, the potential causal link from the metabolic BAG to body weight remained robust across several sensitivity checks despite the identification of two potential outlier instrumental variables, namely, rs117233107 and rs33959228.

In addition, we used the LCV method and found a partially genetically causal effect from longevity (99th survival percentile) to the brain BAG (genetic causality proportion =0.45±0.20, P-value=0.04). Importantly, we selected the LCV method over Mendelian randomization because of the partial population overlap between the longevity GWAS summary statistics and our BAG GWAS summary statistics. The LCV analysis also detected a partially genetically causal effect from telomere length to the immune BAG (genetic causality proportion =0.33±0.12, P-value=0.0002) and the pulmonary BAG (genetic causality proportion =0.67±0.20, P-value=3.57×10^-16^) (**Supplementary eTable 6**).

## Discussion

The current study comprehensively depicts the genetic architecture of common genetic variants on biological aging of nine human organ systems using multimodal data from 377,028 European ancestry participants. We identified many genomic loci for the BAGs of nine human organ systems, which exhibited significant associations with a wide range of clinical traits documented in the GWAS Catalog. These associations were observed within a phenotypic landscape characterized by BAG-organ specificity and inter-organ connections. The brain BAG showed the highest SNP-based heritability estimate among all nine organ systems. GSEA, tissue-specific gene expression patterns, and heritability enrichment results provided additional evidence supporting biological validation for BAG-organ specificity and inter-organ connections. The phenotypic correlation between BAGs was a proxy for their genetic correlation, thereby supporting the long-standing Cheverud’s Conjecture. Mendelian randomization demonstrated potential causal relationships between chronic diseases, particularly AD and type 2 diabetes, body weight, sleep duration, and the nine BAGs.

Our large-scale multi-organ GWAS significantly expands the current catalog of genetic variants associated with health-related traits. The discovery of these identified genomic loci has significant clinical implications. These findings provide an invaluable foundation to validate genes or regulatory elements, molecular pathways, and biological processes related to the clinical traits and diseases of interest in the current study and future GWAS analyses. Previous GWAS mainly focused on the BAG in one organ system, such as the brain BAG^54–57^ from imaging-derived phenotypes. These investigations have largely overlooked the inherent interconnectedness of human organ systems, which are intricately intertwined with distinct axes. Recent studies have identified notable axes, such as the heart-brain-liver^11^, brain-eye^58^, and brain-heart^59^ axes, highlighting the importance of comprehending these intricate relationships to understand human physiology and health.

Our phenome-wide associations validate the pleiotropic effects of the identified genomic loci, influencing various health-related clinical traits in the GWAS Catalog. Our findings also highlight BAG-organ specificity and inter-organ connections, further supporting that biological aging is a complex, multifaceted phenomenon. The human brain regulates various physiological processes and maintains homeostasis throughout the body. Consequently, it is unsurprising that the brain exhibits interconnectedness with clinical traits associated with multiple organ systems. The remarkable enrichment of metabolic traits across various organ systems is unsurprising. As a vital metabolic organ, the liver substantially overlaps genetic variants and loci with both the hepatic and metabolic BAGs. Biologically, the liver’s metabolic functions are intricately regulated by hormones like insulin and other metabolic regulators^12^. Similarly, the interplay between immune and metabolic processes is essential for maintaining overall health and is crucial for the body’s ability to respond to pathogens and regulate metabolic homeostasis^6^.

We highlighted that the brain BAG is the most heritable among the nine organ systems. Determining the genetic heritability of specific organ systems can be complex as no organ system functions independently, and many diseases or traits involve complex interactions between multiple organ systems, as well as genetic and environmental factors. The brain plays a crucial role in developing and functioning various physiological processes across the body. Its intricate structure and diverse cell types render it vulnerable to genetic influences^60^. Therefore, the brain may exhibit higher genetic stability and less environmental variability^61^ than other organs. The human brain’s extensive functional connectivity and intricate networks may also contribute to its higher heritability. These networks facilitate the transmission of genetic information and the propagation of genetic effects across different brain regions^30^. Lastly, genetic variations shaping the human brain are pleiotropic and influence cognitive abilities, behavior, and susceptibility to neurological and psychiatric disorders. Collectively, these factors may contribute to the marked genetic heritability observed in the human brain compared to other organ systems.

Our gene-level and partitioned heritability analyses further validate our GWAS findings, supporting BAG-organ specificity and inter-organ connections. In GSEA, the genes associated with the cardiovascular BAG were implicated in the IGF-II pathway. IGF-II activates two receptors (IGF-1R and IR-A) to promote cell growth and survival. The IGF signaling pathway is essential for cardiac development in the human heart - the first functional organ to develop^62^. In particular, IGF-II promotes fetal cardiomyocyte proliferation through the tyrosine kinase receptors IGF1R and INSR. Previous research provided appealing evidence on IGF signaling in cardiac regeneration in animal models and induced pluripotent stem cells^63^. The flavonoid glucuronidation pathway was the most significant enrichment result shown in the hepatic BAG. A previous study demonstrated that procyanidin C1, a flavonoid in grape seed extract, extended the lifespan of mice^64^. Furthermore, ample evidence indicated that natural flavonoids could be potential therapeutic approaches for non-alcoholic fatty liver disease^65^. The metabolites formed through this pathway can also exert effects beyond the liver and impact other organ systems. Our tissue-specific gene expression analyses provided additional support for the biological relevance of our GWAS findings, as the identified genes exhibited specific expression patterns within tissues from the corresponding organ systems.

The heritability enrichment analysis further validates the BAG-organ specificity and inter-organ connections by highlighting the disproportional heritability enrichment of genetic variants in different functional categories, cell types, tissues, and chromatin states. The cell type-specific enrichment results in the brain (i.e., oligodendrocytes) and cardiovascular (i.e., neurons) BAGs align with previous research. Specifically, Zhao et al. conducted a large-scale GWAS on brain white matter microstructure and found significant heritability enrichment in glial cells, particularly oligodendrocytes^31^, which aligns with our current findings. Our previous multimodal brain BAG GWAS^54^ also confirmed this enrichment in the brain BAG derived from the white matter microstructural features. Similarly, research has revealed the presence of an “intrinsic cardiac nervous system” within the heart, often called the “heart brain.” This system consists of around 40,000 neurons similar to those found in the brain, indicating that the heart possesses a distinct nervous system^66^.

Our genetic correlation results confirmed that the genetic correlation generally mirrors phenotypic correlations in multi-organ biological age. This suggests that environmental factors likely affect the aging of multiple organ systems in the same direction. Providing evidence for Cheverud’s Conjecture can have clinical implications by providing valuable insights into the genetic basis of complex age-related diseases. For instance, by identifying the shared genetic factors underlying multiple age-related diseases, we can target these common pathways to develop novel treatments or repurpose existing drugs^67^ that have proven efficacy in one disease or condition for treating others. Moreover, the validation of Cheverud’s Conjecture emphasizes the importance of considering the genetic covariance of age-related diseases in clinical practice. It underscores the need for comprehensive genetic assessments and genomic analyses to understand disease risk and progression^68^.

We found a bi-directional causal relationship between the hepatic and musculoskeletal BAGs. Abundant research has suggested that liver function and metabolic health, particularly related to glucose and lipid metabolism, can significantly impact musculoskeletal health^69^. This inter-organ connection can cause dysregulation of liver metabolism (e.g., non-alcoholic fatty liver disease) linked to musculoskeletal disorders, including osteoporosis, sarcopenia, and muscle wasting. The musculoskeletal system can also exert an inverse influence on liver function. Regular physical activity and muscle strength have been linked to enhanced liver health and decreased susceptibility to liver diseases. To further support this, causal effects of primary biliary cholangitis, a chronic liver disease, on elevated musculoskeletal BAG were confirmed in our Mendelian randomization results (**Fig. 6**). The absence of direct causal relationships between the remaining BAGs can be attributed to various factors with potential explanations and implications. One possible explanation is that the brain BAG, having the smallest sample size in our GWAS (after removing overlapping participants), may be limited in statistical power. In addition, this may suggest that various factors, including chronic diseases, environmental exposures, and lifestyle choices, influence biological aging in alternative pathways or mediate such changes. Thus, understanding the collective contribution of chronic diseases, environmental factors, and lifestyle choices is crucial for comprehending the overall aging process and its impact on organ health.

We found that several clinical traits collectively cause organ systems to appear older or younger than their chronological age. For instance, body weight was causally associated with the immune, musculoskeletal, metabolic, and pulmonary BAGs. For several reasons, body weight can causally influence multiple organ systems. Excessive body weight (e.g., obesity) has metabolic consequences, including increased inflammation, insulin resistance, and dysregulation of metabolic pathways in adipose tissue^70^. It also leads to mechanical stress on the body, contributing to musculoskeletal strain^71^ and cardiovascular workload^72^. Hormonal imbalances^73^ and lifestyle factors linked to body weight also influence multi-organ function and the development of chronic diseases. Being overweight is also a risk factor for type 2 diabetes, which was positively causally associated with metabolic BAG (**Fig. 6**). AD was causally linked to the brain, hepatic, musculoskeletal, and renal BAGs. AD, a neurodegenerative disorder primarily affecting the brain, can have causal influences on multiple organ systems. For example, it has broader systemic involvement beyond the brain, mediated by mechanisms including protein aggregation (e.g., amyloid-β and tau^74^), vascular dysfunction^75^, inflammation^76^, and other secondary factors. Protein aggregates can spread to other organs; vascular abnormalities can impact blood flow; inflammation can affect distant organ systems; secondary factors, such as medication use and lifestyle changes, also contribute.

### Limitations

This study has several limitations. First, the generalizability of genetic findings from European to non-European ancestry populations is limited. Future studies can extend their scope to encompass a more diverse array of underrepresented ethnicities, a wider range of disease cohorts, and individuals of varying ages throughout their entire lifespan. Secondly, it is essential to approach the causality results cautiously, considering the assumptions underlying Mendelian randomization. In future studies, more advanced multi-response Mendelian randomization methods^77^ should be utilized. Thirdly, despite our efforts of quality check analyses to scrutinize our primary GWAS, it’s essential to acknowledge that potential ascertainment bias^78^ and confounding related to demographic and socioeconomic factors could potentially introduce cryptic population stratification, which may not be entirely resolved in the current study. Finally, the large number of genomic loci identified in our GWAS may have connections to BAGs due to various factors, such as biological processes, potential confounding due to demographics, or specific study design and phenotyping aspects. It’s important to note that the effects at these loci might not be inherently biological but could be influenced by other unmeasured confounding factors.

### Outlook

In conclusion, our study presents compelling genetic evidence to support that *no organ system is an island*^1^ – the collective influence of various chronic diseases on these multi-organ systems and the interconnectedness among these human organ systems. These findings highlight the importance of comprehensively understanding the underlying causes of chronic diseases within the multi-organ network. By shedding light on its comprehensive genetic architecture, our study paves the way for future research to unravel complex disease mechanisms and develop holistic approaches to ameliorate overall organ health.

## Methods

### Method 1: Support vector machines to predict the chronological age of nine organ systems

Our prior study^3^ used support vector machines to predict the chronological age of healthy individuals – defined as no self-reported and healthcare-documented lifetime chronic medical conditions – based on phenotypes from the nine organ systems. Support vector machine regression was preferred over linear regression for its enhanced robustness to outliers and overfitting. We performed a 20-fold cross-validation procedure and developed predictive models for each organ system.

In each of the 20-fold cross-validation iterations, a linear support vector machine was employed to predict chronological age. The training set consisted of 19 folds of individuals, and the fitted regression coefficients (feature weights) were then applied iteratively to the remaining held-out set (test set) to predict the chronological age of each healthy individual. This approach ensured that the prediction model was not trained using the same individuals for which it made predictions, minimizing the risk of overfitting. Before each iteration of model training, all measures (excluding categorical variables) were standardized using the weighted column mean and standard deviation computed within the training set. The SVM box constraint and kernel scale were set to unity, while the half-width of the epsilon-insensitive band was set to a tenth of the standard deviation of the interquartile range of the predicted variable (chronological age). The SVM was solved using sequential minimal optimization with a gap tolerance of 0.001. The mathematical principles of support vector machines are well-established in the field and have been widely recognized^79^. Further details on this topic can be found in our previous study^3^.

The concept of biological age gap derived from artificial intelligence has been widely investigated, especially the brain age^80,81^. The calculation of the nine BAGs were established in our previous works^3,14^. We previously showed that the prediction accuracy of biological age was not influenced by the number of phenotypes, despite variations across different organ systems. While some prior studies^82^ used deep learning for brain BAG and obtained a lower mean absolute error, we have previously demonstrated that lower mean absolute error might compromise sensitivity to disease-related information^83^. In our previous GWAS^14^, which separately examined three multimodal brain BAGs derived from T1-weighted, diffusion, and resting-state fMRI data, we extensively investigated the influence of various brain imaging feature types and study designs on the genetic signals. Our results unveiled both the consistency and distinctions in the genetic foundations across these diverse contexts. Finally, we recognize that ascertainment bias may be present in our GWAS due to variations in sequencing techniques, differences between populations (e.g., disease populations vs. healthy controls), and socioeconomic factors that have not been explicitly modeled in our study.

### Method 2: Study populations

UKBB is a population-based study of approximately 500,000 people recruited between 2006 and 2010 from the United Kingdom. The UKBB study has ethical approval, and the ethics committee is detailed here: https://www.ukbiobank.ac.uk/learn-more-about-uk-biobank/governance/ethics-advisory-committee.

The current study analyzed multimodal data, including imaging-derived phenotypes (IDP) and physical and physiological measures in nine human organ systems from 154,774 UKBB participants. In our previous study, we constructed BAGs for eight organ systems using machine learning, including MRI data for brain BAG from 30,108 participants (European ancestry), pulse rate and blood pressure data for cardiovascular BAG, liver-related blood biomarkers for hepatic BAG, C-reactive protein and blood hematology variables for immune BAG, blood biomarkers for metabolic BAG, physical measurements and vitamin D for musculoskeletal BAG, lung functioning measurements for pulmonary BAG, and glomerular filtration and electrolyte regulation biomarkers for renal BAG from 111,543 participants.

Furthermore, the current study also used 60 optical coherence tomography (OCT)-derived measures from 36,004 participants to derive the BAG of the ninth organ system – the eye BAG. The inclusion criteria for the features used to predict the eight BAGs, the machine learning methods, and cross-validation procedures are detailed in our previous study^3^. We initially used the 88 OCT-derived measures (category ID: 10079) for the additional eye BAG in 67,549 participants. Of these measures, 28 were excluded due to a high missing rate (>20% of participants). Additionally, 4172 participants were excluded due to missing data, and 1798 participants identified as outliers (outside mean +/-6SD) for the 60 remaining measures were discarded. This finally resulted in 41,966 participants (36,004 European ancestry participants). The included 2444 features to derive the BAG of the nine organ systems are presented in **Supplementary eFile 21**.

In addition, we also performed GWAS for seven variables from 222,254 UKBB participants by excluding the 154,774 participants from the BAG populations to avoid bias due to overlapping samples. These variables included six lifestyle factors and one cognitive variable: *N*=219,661 (European ancestry) for coffee intake (Field ID:1498), *N*=221,393 for fresh fruit intake (Field ID:1309), *N*=221,739 for tea intake (Field ID:1488), *N*=220,765 for sleep duration (Field ID:1160), *N*=209,012 for time spent outdoors in summer (Field ID:1050), *N*=221,337 for body weight (Field ID:21002), and *N*=220,624 for reaction time (Field ID:20023).

The current work was jointly performed under application numbers 35148 (i.e., genetic data) and 60698 (i.e., the generation of the nine BAGs). In total, we analyzed data from 377,028 individuals of European ancestry in the current study.

### Method 3: Genetic analyses

We used the imputed genotype data for all genetic analyses, and our quality check pipeline resulted in 487,409 participants and 6,477,810 SNPs. After merging with the population for each BAG, we included 30,108-111,543 European ancestry participants for the nine BAGs (**Fig. 1**). To avoid bias due to overlapping populations^16^, we also used the rest of the UKBB participants of European ancestry (non-overlapping) to derive the GWAS summary statistics for several lifestyle factors (**Method 3j**). We summarize the genetic QC pipeline. First, we excluded related individuals (up to 2^nd^-degree) from the complete UKBB sample using the KING software for family relationship inference.^84^ We then removed duplicated variants from all 22 autosomal chromosomes. Individuals whose genetically identified sex did not match their self-acknowledged sex were removed. Other excluding criteria were: i) individuals with more than 3% of missing genotypes; ii) variants with minor allele frequency (MAF) of less than 1% (dosage mode^85^); iii) variants with larger than 3% missing genotyping rate; iv) variants that failed the Hardy-Weinberg test at 1×10^-10^. To adjust for population stratification,^86^ we derived the first 40 genetic principle components (PC) using the FlashPCA software^87^. Details of the genetic quality check protocol are described elsewhere^14,46,88^. Details of the genetic quality check protocol are described elsewhere^29,46^.

**(a) : Genome-wide association analysis**: For GWAS, we ran a linear regression using Plink^89^ for each BAG, controlling for confounders of age, dataset status (training/validation/test or independent test dataset), age x squared, sex, age x sex interaction, age-squared x sex interaction, and the first 40 genetic principal components; additional covariates for total intracranial volume and the brain position in the scanner were included for brain BAG GWAS. We adopted the genome-wide P-value threshold (5 x 10^-8^) and annotated independent genetic signals considering linkage disequilibrium (see below).

To check the robustness of our GWAS results, we performed several sensitivity check analyses, including *i*) sex-stratified GWAS for males and females, *ii*) split-sample GWAS by randomly dividing the entire population into two splits (sex and age-matched), *iii*) non-European ancestries GWAS, and *iv*) fastGWA for linear mixed effect GWAS, hypothesizing that the main GWASs with European ancestry did not show substantial genomic inflation due to cryptic population stratification. In all our sensitivity check analyses, we considered linkage disequilibrium. We only evaluated the independent significant SNPs of the two sets of *β* coefficients between splits, genders, ancestry groups, and GWAS methods. The definition of the independent significant SNPs used the same parameters as in FUMA (**Supplementary eMethod 1**). We used the raw genotype data and the Plink *clump* command (250 kb) and defined a set of SNPs in linkage disequilibrium with the independent significant SNPs – analogous to the candidate SNPs in FUMA.

**(b) : SNP-based heritability**: We estimated the SNP-based heritability (*h^2^*) using GCTA^24^ with the same covariates in GWAS. We reported results from two experiments for each BAG using *i*) the full sample sizes and *ii*) randomly down-sampled sample sizes to that (*N*=30,108) of the brain BAG with comparable distributions regarding sex and age – the sample size of brain BAGs was smaller than the other BAGs.

**(c) : Annotation of genomic loci**: The annotation of genomic loci and mapped genes was performed via FUMA^90^. For the annotation of genomic loci, FUMA first defined lead SNPs (correlation *r^2^* ≤ 0.1, distance < 250 kb) and assigned them to a genomic locus (non-overlapping); the lead SNP with the lowest P-value (i.e., the top lead SNP) was used to represent the genomic locus in **Fig. 1**. For gene mappings, three different strategies were considered. First, positional mapping assigns the SNP to its physically nearby genes (a 10 kb window by default). Second, eQTL mapping annotates SNPs to genes based on eQTL associations using the GTEx v8 data. Finally, chromatin interaction mapping annotates SNPs to genes when there is a significant chromatin interaction between the disease-associated regions and nearby or distant genes^90^. The definition of top lead SNP, lead SNP, independent significant SNP, and candidate SNP can be found in **Supplementary eMethod 1**.

For the top lead SNP of each identified genomic locus, we showcased whether it was previously associated with any clinical traits considering linkage disequilibrium (5000kb around the top lead SNP) in the EMBL-EBI GWAS Catalog platform (https://www.ebi.ac.uk/gwas/home). For instance, we aimed to query the locus with the top lead SNP (rs60569686) associated with the renal BAG. First, we looked up the chromosomal position (i.e., chromosome 13) and found that the location is chr13:49170160 (GRCh38). We then search the GWAS Catalog for a 5000kb region around this top lead SNP: “chr13:49167660-49172660” (https://www.ebi.ac.uk/gwas/regions/chr13:49167660-49172660; query date: 12th October 2023). In this region, we discovered no prior associations. It’s important to note that this search is not comprehensive, as new GWAS studies continually emerge on various open platforms, such as IEU OpenGWAS^91^ (https://gwas.mrcieu.ac.uk/) and GWAS ATLAS^23^ (https://atlas.ctglab.nl/PheWAS).

**(d) : Phenome-wide association look-up queries**: We first queried the significant independent SNPs within each locus in the EMBL-EBI GWAS Catalog (query date: 24^th^ April 2023, via FUMA version: v1.5.4) to determine their previously identified associations with any other traits (P-value<1×10^-5^ by default in the EMBL-EBI GWAS Catalog). For visualization purposes, we further mapped the associated traits into organ-specific groups and other chronic disease traits and cognition. We performed the following procedure to fully consider LD and remove redundant associations among the independent significant SNPs. If the top lead SNP showed any clinical associations, this would present the current locus; if not, we queried the independent significant SNPs (in high correlation with the top lead SNP), starting with the most significant SNPs, until we identified established associations. In this way, only one genetic variant within each genomic locus was considered. We also conducted a complementary phenome-wide association query on the GWAS Atlas platform. We applied the same P-value threshold search criteria as those used in the EMBL-EBI GWAS Catalog. The same procedure, considering linkage disequilibrium and redundant associations, was applied. These exemplary findings are presented as a supplementary search to complement the results shown in **Fig. 2a**, and are available in **Supplementary eText 2**.

**(e) : Gene set enrichment analysis**: We first performed gene-level association analysis using MAGMA^33^. First, gene annotation was performed to map the SNPs (reference variant location from Phase 3 of 1,000 Genomes for European ancestry) to genes according to their physical positions. Of note, other advanced annotation methods exist that integrate functional insights, such as brain chromatin interaction^92^ and cell-type-specific gene expression^93^. We then performed gene-level associations based on the SNP GWAS summary statistics to obtain gene-level p-values between the nine BAGs and the curated protein-encoding genes containing valid SNPs. We performed GSEA using the gene-level association p-values. Gene sets were obtained from the Molecular Signatures Database (MsigDB, v7.5.1)^94^, including 6366 curated and 10,402 ontology gene sets. All other parameters were set by default for MAGMA. The Bonferroni method was used to correct multiple comparisons for all tested gene sets.

**(f) : Tissue-specific gene expression analysis**: MAGMA performed gene-property analyses to identify tissue-specific gene expression of the nine BAGs. The gene-property analysis converts the gene-level association P-values (above) to Z scores and tests a specific tissue’s gene expression value versus the average expression value across all tissues in a regression model. Bonferroni correction was performed for all tested gene sets. We reported the results from the 54 tissue types using the GTEx V8 data.

**(g) : Gene-drug-disease network**: We tested the enrichment of the nine BAG-linked genes in the targeted gene sets for different drug categories from the DrugBank database^38^. The gene-drug-disease network was constructed to prioritize potentially repositionable drugs. The GREP software^37^ performs Fisher’s exact tests to examine whether the prioritized genes are enriched in gene sets targeted by drugs in a clinical indication category for a certain disease or condition. Bonferroni correction was performed for all tested drugs.

**(h) : Genetic correlation**: We used the LDSC^17^ software to estimate the pairwise genetic correlation (*g_c_*) between each pair of BAGs, as well as between the nine BAG and 41 other clinical traits, including chronic diseases involving multiple organ systems, such as AD for brain and chronic kidney disease for kidney, cognition, and lifestyle factors. We used the precomputed LD scores from the 1000 Genomes of European ancestry. To ensure the suitability of the GWAS summary statistics, we first checked that the selected study’s population was European ancestry; we then guaranteed a moderate SNP-based heritability *h^2^* estimate. Notably, LDSC corrects for sample overlap and provides an unbiased estimate of genetic correlation^68^. The inclusion criteria and finally included traits are detailed in **Supplementary eTable 3**. Bonferroni correction was performed for the 41 clinical traits.

**(i) : Partitioned heritability estimate**: Our objective is to comprehend how distinct functional genome categories play varying roles in contributing to the heritability of the nine BAGs. Therefore, the partitioned heritability analysis via stratified LD score regression calculates the extent to which heritability enrichment can be attributed to predefined and annotated genome regions and categories^40^. Three sets of functional categories and cell and tissue-specific types were considered. First, the partitioned heritability was calculated for 53 general functional categories (one including the entire set of SNPs). The 53 functional categories are not specific to any cell type and include coding, UTR, promoter and intronic regions, etc. The details of the 53 categories are described elsewhere^40^. Subsequently, cell and tissue type-specific partitioned heritability was estimated using gene sets from Cahoy et al.^41^ for three main cell types (i.e., astrocyte, neuron, and oligodendrocyte), multi-tissue chromatin states-specific data (ROADMAP^42^ and ENTEx^43^), and multi-tissue gene expression data (GTEx V8^36^). Bonferroni correction was performed for all tested annotations and categories. The detailed methodologies for the stratified LD score regression are presented in the original work^40^. The LD scores and allele frequencies for the European ancestry were obtained from a predefined version based on data from the 1000 Genomes project.

**(j) : Two-sample bi-directional Mendelian randomization**: We investigated whether one BAG was causally associated with another BAG and whether the 41 clinical traits were causally associated with the nine BAGs (**Fig. 5**). To this end, we employed a bidirectional, two-sample Mendelian randomization using the TwoSampleMR package^95^. Both the forward and inverse Mendelian randomization were performed between each pair of traits by switching the exposure and outcome variables. We applied five different Mendelian randomization methods and reported the results of inverse variance weighted (IVW) in the main text and the four others (i.e., Egger, weighted median, simple mode, and weighted mode estimators) in the supplement.

Mendelian randomization needs to fulfill several instrumental variable assumptions^96^, including:

- the genotype is associated with the exposure
- the genotype is associated with the outcome through the studied exposure only (exclusion restriction assumption)
- the genotype is independent of other factors that affect the outcome (independence assumption^97^)

We followed a systematic procedure guided by the STROBE-MR Statement^98^ in all steps of our causality analyses, including selecting exposure and outcome variables, reporting comprehensive statistics, performing sensitivity checks for potential violations of underlying assumptions, and performing the analyses using alternative methods and software^53,77^. For the causal inference of each pair of BAGs, all GWAS summary statistics were derived from our analyses by excluding overlapping populations of the two BAGs. For example, to test the causal relationship between the brain BAG and cardiovascular BAG, we reran GWAS for the cardiovascular BAG by excluding the partially overlapping population from the brain BAG. For all the seven body organ systems that had entirely overlapping populations, we used the GWAS data from the split-sample analyses (**Method 3a**). For instance, the GWAS for the cardiovascular BAG was from the first-split data, and the pulmonary BAG was from the second-split data. Bonferroni correction was performed for the tested BAGs.

One key challenge in our hypothesis-driven Mendelian randomization is to select these exposure variables unbiasedly. Clinical traits sharing common genetic covariance with nine BAGs are more likely to be causally associated with them. We performed a systematic inclusion procedure using the following criteria to overcome this. We manually queried the 41 clinical traits – used in our genetic correlation analyses – in the IEU GWAS database, specifically curated for Mendelian randomization analyses. We ranked all available studies for a certain trait (e.g., AD) based on the sample sizes. We then chose the study whose populations were of European ancestry and did not include UKBB participants to avoid bias due to overlapping populations^16^. For the traits whose GWAS data were available in the IEU GWAS database, we used the TwoSampleMR package to perform the Mendelian randomization analysis. For the traits whose data were not appropriate in the IEU GWAS database, we then performed another manual query in the EMBL-EBI GWAS Catalog database to download the available GWAS summary statistics with the same filter criteria. For the traits whose GWAS data were dominated by studies using UKBB participants in both databases, we ran GWAS using our own UKBB data by excluding overlapping populations. Finally, after harmonizing their GWAS summary statistics (using the function *harmonise_data* from 2SampleMR), this resulted in 17 clinical traits with at least eight valid IVs (i.e., SNPs). The 17 clinical traits included chronic diseases affecting multiple organ systems, cognition, and lifestyle factors (**Supplementary eTable 7**). Bonferroni correction was performed for all tested clinical traits.

We performed several sensitivity analyses. First, a heterogeneity test was performed to check for violating the IV assumptions. Horizontal pleiotropy was estimated to navigate the violation of the IV’s exclusivity assumption^99^ using a funnel plot, single-SNP Mendelian randomization approaches, and Mendelian randomization Egger estimator^100^. Moreover, the leave-one-out analysis excluded one instrument (SNP) at a time and assessed the sensitivity of the results to individual SNP.

Following these analyses, we performed three supplementary sensitivity checks for some specific significant causal signals: *i*) The exclusion of two common SNPs/IVs (rs429358 and rs7412) in the *APOE* gene, considering their potential pleiotropic effects for the hepatic BAG on musculoskeletal BAG; *ii*) Incorporating body weight as a covariate in the GWAS for the bi-directional causality between the hepatic BAG and musculoskeletal BAG, as body weight displayed causal associations with BAGs in multiple organ systems; *iii*) Re-executing the Mendelian randomization analysis using alternative software. Specifically, we scrutinized the causal relationship between the hepatic BAG and musculoskeletal BAG using the latent causal variance (LCV) model^53^. Employing different modeling assumptions and instrumental variables in contrast to Mendelian randomization, it examined the causal relationship between two interchangeable traits without distinction between the direct and inverse directions. A latent causal variable (L) acted as a mediator for the genetic correlation between the two traits, allowing us to quantify the genetic causality proportion (GCP). A positive GCP value between 0 and 1 indicates that trait 1 is partially genetically causal; a negative GCP value means trait 2 is partially genetically causal.

**(h) : Bayesian colocalization**: The R package (*coloc*) was employed to investigate the genetic colocalization signals between two traits at each genomic locus defined by the pulmonary BAG GWAS. We employed the Fully Bayesian colocalization analysis using Bayes Factors (*coloc.abf*). The method tests five hypotheses, denoted by their posterior probabilities: H0 (no association with either trait), H1 (association with trait 1 but not trait 2), H2 (association with trait 2 but not trait 1), H3 (association with both traits but with separate causal variants), and H4 (association with both traits with a shared causal variant). It examines the posterior probability (PP.H4.ABF: Approximate Bayes Factor) to evaluate hypothesis *H4*, which suggests the presence of a single shared causal variant associated with both traits within a specific genomic locus. To determine the significance of the *H4* hypothesis, we set a threshold of PP.H4.ABF>0.8^20^ and at least 100 SNPs were included within the genomic locus. All other parameters (e.g., the prior probability of p_12_) were set as default.

## Data Availability

The GWAS summary statistics corresponding to this study are publicly available on the MEDICINE knowledge portal (https://labs-laboratory.com/medicine).

## Code Availability

The software and resources used in this study are all publicly available:

- MEDICINE: https://labs-laboratory.com/medicine, knowledge portal for dissemination
- BioAge: https://github.com/yetianmed/BioAge, biological age prediction
- PLINK: https://www.cog-genomics.org/plink/, linear model GWAS
- FUMA: https://fuma.ctglab.nl/, gene mapping, genomic locus annotation
- GCTA: https://yanglab.westlake.edu.cn/software/gcta/#Overview, heritability estimates, mixed effect GWAS
- LDSC: https://github.com/bulik/ldsc, genetic correlation, and partitioned heritability
- TwoSampleMR: https://mrcieu.github.io/TwoSampleMR/index.html, Mendelian randomization
- Coloc: https://github.com/chr1swallace/coloc, Bayesian colocalization
- LCV: https://github.com/lukejoconnor/LCV, Latent causal variable for causal inference

## Competing Interests

None

## Authors’ contributions

Dr. Wen has full access to all the data in the study and takes responsibility for the integrity of the data and the accuracy of the data analysis.

*Study concept and design*: Wen

*Acquisition, analysis, or interpretation of data*: Wen

*Drafting of the manuscript*: Wen, Tian, Zalesky, Davatzikos

*Critical revision of the manuscript for important intellectual content*: all authors

*Statistical analysis*: Wen

*BAG index generation*: Tian

## Acknowledgments

The primary funding support for this present study is from the initial funding package provided by the University of Southern California for WJ. We want to express our sincere gratitude to the UK Biobank team for their invaluable contribution to advancing clinical research in our field. This study used the UK Biobank resource under Application Numbers: 35148 and 60698. Furthermore, we acknowledge the collaborative effort between the University of Southern California, the University of Pennsylvania, and the University of Melbourne in conducting this research. We gratefully acknowledge the support of the iSTAGING consortium, funded by the National Institute on Aging through grant RF1 AG054409 at the University of Pennsylvania (CD). Additionally, we acknowledge the funding program from the Rebecca L. Cooper Foundation at the University of Melbourne (AZ). Lastly, WJ would like to thank Paraskevi Parmpi and Jessica Incmikoski for their valuable administrative support during his postdoctoral research at AIBIL. We thank Dr. Joris Deelen and Dr. Joanne M. Murabito for their generosity in providing the GWAS summary statistics from their research^49^ during the revision process.

## Footnotes

1 We adapt the concept of “No Man Is An Island” from the poem by John Donne, highlighting the interconnectedness of human organ systems.

## Notes

### Competing Interest Statement

The authors have declared no competing interest.

### Funding Statement

The iSTAGING consortium is a multi-institutional effort funded by NIA by RF1 AG054409.

### Author Declarations

We requested and used data only from the UK Biobank. We do not need to provide local IRB at UPenn. UK Biobank has approval from the North West Multi-centre Research Ethics Committee (MREC) as a Research Tissue Bank (RTB) approval. This approval means that researchers do not require separate ethical clearance and can operate under the RTB approval (there are certain exceptions to this which are set out in the Access Procedures, such as re contact applications). UK Biobank seeks to keep the wider public informed of research findings deriving from access to the Resource. For this reason, unless agreed otherwise, details of approved Research Projects identifying the Principal Investigator will be made available on the UK Biobank website. Periodic updates of research that have been conducted, and publications that derive from the Resource, will be linked to the UK Biobank website.

### Summary of Updates

We have corrected the author affiliation and funding information for another round of revision

