## Supplement for "The Genetic Architecture of Biological Age in Nine Human Organ Systems"

**eMethod 1: The definition of genomic loci, independent significant SNP, lead SNP, candidate SNP**
**eText1: Sensitivity check analyses for the main GWAS of the nine BAGs using European ancestry**
**eText2: Phenome-wide association query using the GWAS Atlas platform**
**eText3: Sensitivity check analyses for the causality between the hepatic BAG and musculoskeletal BAG**
**eFigure 1: GWAS Manhattan plots for the brain BAG**
**eFigure 2: GWAS Manhattan plots for the cardiovascular BAG**
**eFigure 3: GWAS Manhattan plots for the eye BAG**
**eFigure 4: GWAS Manhattan plots for the hepatic BAG**
**eFigure 5: GWAS Manhattan plots for the immune BAG**
**eFigure 6: GWAS Manhattan plots for the metabolic BAG**
**eFigure 7: GWAS Manhattan plots for the musculoskeletal BAG**
**eFigure 8: GWAS Manhattan plots for the pulmonary BAG**
**eFigure 9: GWAS Manhattan plots for the renal BAG**
**eFigure 10: Bayesian colocalization analysis for the locus on chromosome 6 between the hepatic and musculoskeletal BAGs**
**eFigure 11: Exemplary genomic locus for each BAG in the nine human organ systems**
**eFigure 12: SNP-based heritability, beta coefficients, and alternative allele frequency using the brain-BAG comparable populations and different inclusion criteria for the SNPs**
**eFigure 13: Trumpet plots of the alternative allele frequency vs. the beta coefficient of the nine BAG GWASs**
**eFigure 14: Manhattan of and QQ plots for the four pulmonary features used to compute the pulmonary BAG**
**eFigure 15: Bayesian colocalization signal between the pulmonary BAG and FEV/FVC**
**eFigure 16: Beta coefficients of the significant colocalization signal between the pulmonary BAG and the four pulmonary features**
**eFigure 17: GSEA using sex-stratified GWAS results**
**eFigure 18: TEA correlations using sex-stratified GWAS results**
**eFigure 19: Genetic correlations using sex-stratified GWAS results**
**eFigure 20: Mendelian randomization sensitivity check for the hepatic BAG on the musculoskeletal BAG**
**eFigure 21: Mendelian randomization sensitivity check for the musculoskeletal BAG on the hepatic BAG**
**eFigure 22: Mendelian randomization sensitivity check for AD on the brain BAG**
**eFigure 23: Mendelian randomization sensitivity check for AD on the hepatic BAG**
**eFigure 24: Mendelian randomization sensitivity check for Crohn's disease on the hepatic BAG**
**eFigure 25: Mendelian randomization sensitivity check for body weight on the immune BAG**
**eFigure 26: Mendelian randomization sensitivity check for type 2 diabetes on the metabolic BAG**
**eFigure 27: Mendelian randomization sensitivity check for AD on the musculoskeletal BAG**
**eFigure 28: Mendelian randomization sensitivity check for IBD on the musculoskeletal BAG**

**eFigure 29: Mendelian randomization sensitivity check for PBC on the musculoskeletal BAG**

**eFigure 30: Mendelian randomization sensitivity check for weight on the musculoskeletal BAG**

**eFigure 31: Mendelian randomization sensitivity check for weight on the pulmonary BAG**

**eFigure 32: Mendelian randomization sensitivity check for AD on the renal BAG**

**eFigure 33: Mendelian randomization sensitivity check for weight on the renal BAG**

**eFigure 34: Mendelian randomization sensitivity check for the brain BAG on sleep duration**

**eFigure 35: Mendelian randomization sensitivity check for the cardiovascular BAG on triglycerides to lipids ratio in very large VLDL**

**eFigure 36: Mendelian randomization sensitivity check for the metabolic BAG on weight**

**eFigure 37: Mendelian randomization sensitivity check for the pulmonary BAG on weight**

**eFigure 38: Causal multi-organ network between the nine biological age gaps and 17 clinical traits of chronic diseases, lifestyle factors, and cognition**

**eTable 1: Heritability estimates using the GCTA software**

**eTable 2: The beta coefficient and its SE estimate from the full sample vs. the down-sampled brain BAG comparable sample**

**eTable 3: Genetic correlation analyses between the pulmonary BAG and the four features used to derive the BAG**.

**eTable 4: Selected 41 clinical traits for genetic correlation analyses**

**eTable 5: Genetic correlations analyses between the nine BAGs and longevity, household income, and telomere length**

**eTable 6: Causal analysis using the LCV method**

**eTable 7: Selected 17 clinical traits for Mendelian randomization analyses**

### eMethod 1: The definition of genomic loci, independent significant SNP, lead SNP, candidate SNP

FUMA defined the significant independent SNPs, lead SNPs, candidate SNPs, and genomic risk loci as follows (https://fuma.ctglab.nl/tutorial#snp2gene):

*Independent significant SNPs*

They are defined as SNPs with $P \leq 5\times10^{-8}$ that are independent of each other at the user-defined $r^2$ (set to 0.6 in the current study). We further describe *candidate SNPs* as those in linkage disequilibrium (LD) with independent significant SNPs. FUMA then queries each candidate SNP in the GWAS Catalog to check whether any clinical traits have been reported to be associated with previous GWAS studies.

*Lead SNPs*

Lead SNPs are defined as independent significant SNPs that are also independent of each other at $r^2<0.1$. If multiple independent significant SNPs are correlated at $r^2 \geq 0.1$, then the one with the lowest individual *P*-value becomes the lead SNP. If $r^2$ threshold is set to 0.1 for the independent significant SNPs, then they would constitute the identical set as the lead SNPs by definition. FUMA thus advises setting $r^2$ to be 0.6 or higher.

*Genomic risk loci*

FUMA defines genomic risk loci to include all independent signals physically close or overlapping in a single locus. First, independent significant SNPs dependent on each other at $r^2 \geq 0.1$ are assigned to the same genomic risk locus. Then, independent significant SNPs with less than the user-defined distance (250 kb by default) away from one another are merged into the same genomic risk locus - the distance between two LD blocks of two independent significant SNPs is the distance between the closest points from each LD block. Each locus is represented by the SNP within the locus with the lowest *P*-value.

**eText 1: Sensitivity check analyses for the main GWAS of the nine BAGs using European ancestry**

We fully considered linkage disequilibrium and only included the independent significant SNPs in this sensitivity check analysis. We exemplified this analysis in the split-sample GWAS. We first used the Plink *clump* command *(--clump-p1 0.00000005 --clump-p2 0.05 --clump-r2 0.60 --clump-kb 250*) to define the independent significant SNPs for the split1 and split2 GWAS. We then included all the unique independent significant SNPs in either of the two split GWASs. We then calculated three statistics to scrutinize the concordance of the two split GWASs:

- *r-β*: Pearson's *r* between the two sets of *β* coefficients from the two splits;
- *C-β*: concordance rate of the sign of the *β* coefficients from the two splits – if the same SNP exerts the same protective/risk effect between the two splits;
- *P-β*: the difference between the two sets of *β* coefficients from the two splits – if the two sets of *β* coefficients (mean) statistically differ.

The two metrics were calculated for sex-stratified, fastGWA, and non-Euroepan GWAS sensitivity check analyses.

**Split-sample GWAS**

**P-values:**

In the split1 GWAS, we found 6, 28, 20, 117, 62, 160, 37, 40, and 127 independent significant SNPs for the brain, cardiovascular, eye, hepatic, immune, metabolic, musculoskeletal, pulmonary, and renal BAGs, and 5, 30, 21, 110, 55, 164, 45, 43, and139 independent significant SNPs in split2 GWAS.

For the brain BAG, we obtained an *r-β* of -0.06 (P-value=0.84; *N*=11), but the two sets of coefficients did not statistically differ (*P-β*=0.70). All the 11 independent significant SNPs showed the same direction of effect (*C-β*=1). The low *r-β* was likely due to small sample sizes in the brain BAG. For all the other 8 BAGs, we obtained significantly h70h *r-β* estimates (0.90<*r-β*<0.99; P-value<$1x10^{-19}$). The two sets of coefficients did not statistically differ (*P-β*>0.48). All independent significant SNPs showed the same direction of effect (*C-β*=1). Detailed results of these SNPs are presented in **Supplementary eFile 2** for split-sample GWAS. The scatter plot of the independent SNPs' *β* coefficients is shown below.

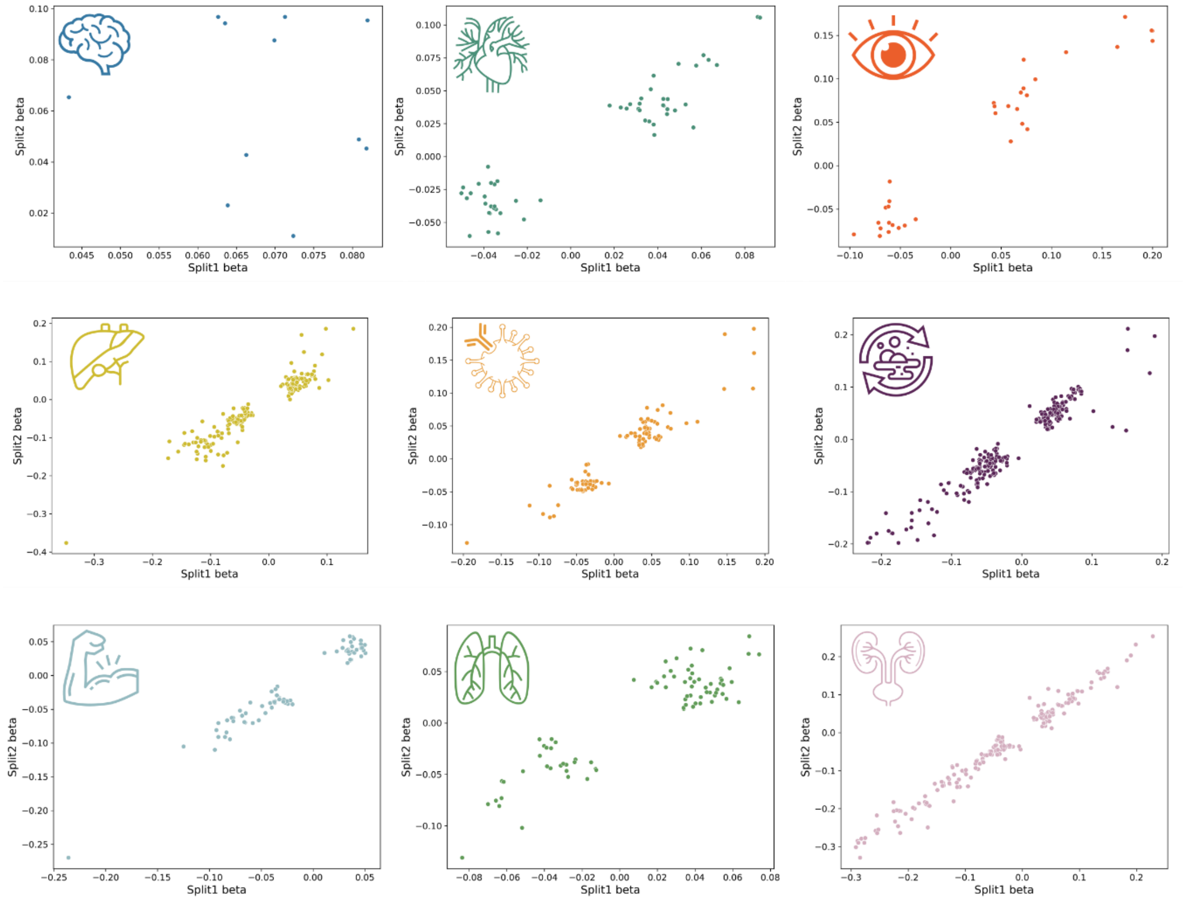

The figures present the scatter plots for the two sets of beta coefficients estimated from different splits.

**Sex-stratified GWAS**

In the female GWAS, we found 7, 24, 23, 286, 116, 142, 153, 30, and 131 independent significant SNPs for the brain, cardiovascular, eye, hepatic, immune, metabolic, musculoskeletal, pulmonary, and renal BAGs, and 7, 38, 22, 126, 275, 286, 42, 71, and 167 independent significant SNPs in the male GWAS.

For the brain BAG, we obtained an *r-β* of -0.869 (P-value=5.29x10$^{-5}$, *N*=14), but the two sets of coefficients did not statistically differ (*P-β*=0.66). 13 out of the 14 independent significant SNPs showed the same direction of effect (*C-β*=0.93). The one independent significant SNP (rs1634777) that had the opposite *β* sign in males compared to females was because the *β* coefficient was close to 0 (*β*=-0.000417162) and was not statistically significant (P-value=0.99). For all the other 8 BAGs, we obtained significantly high *r-β* estimates (0.30<*r-β*<0.96; P-value<2.57x10$^{-7}$). The two sets of coefficients did not statistically differ (*P-β*>0.40), except for the immune BAG (*P-β*=0.013). Most independent significant SNPs showed the same direction of effect (*C-β*>0.89), except for the immune (0.54) and musculoskeletal BAGs (0.70). Detailed results of these SNPs are presented in **Supplementary eFile 3** for sex-stratified GWAS. The scatter plot of the independent SNPs' *β* coefficients is shown below.

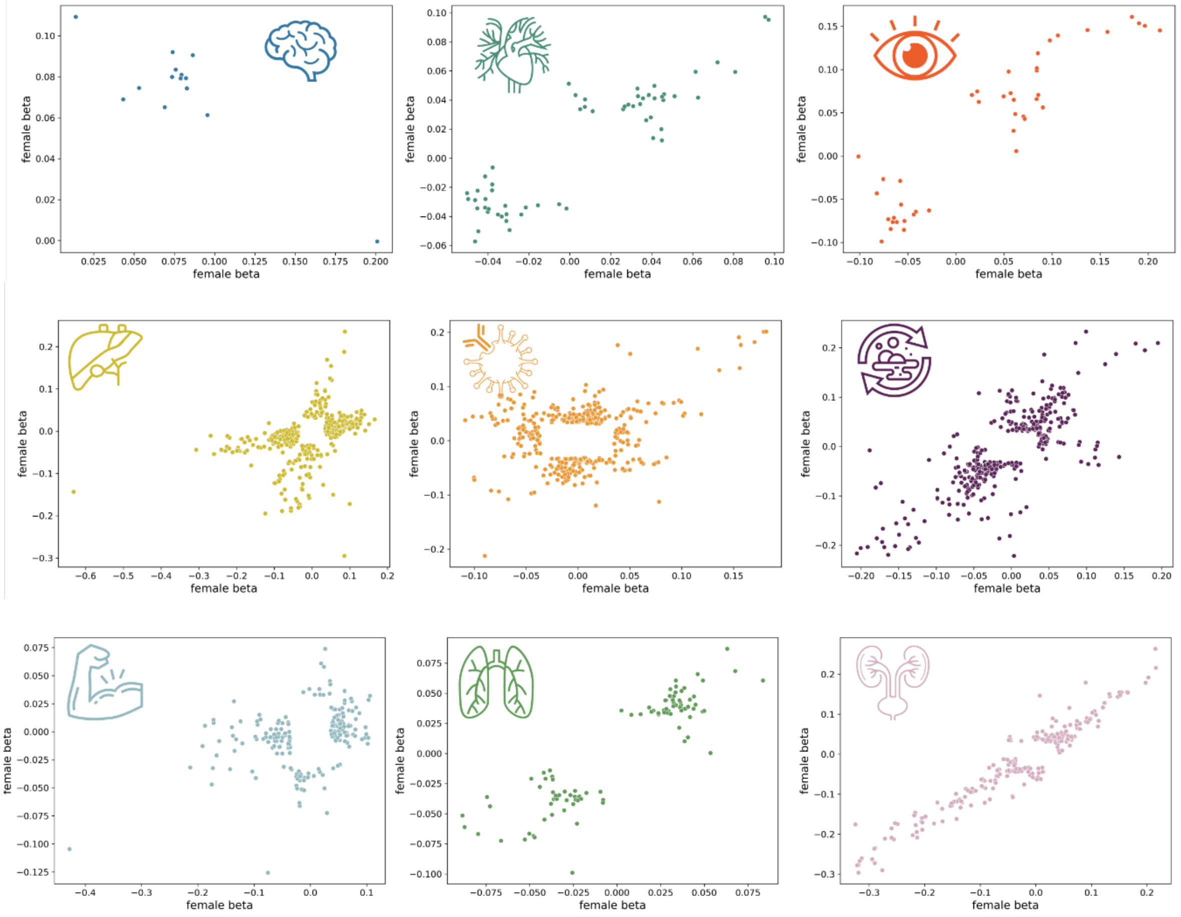

The figures present the scatter plots for the two sets of beta coefficients estimated from different genders.

**fastGWA vs PLINK GWAS**

In the PLINK GWAS, we found 27, 124, 69, 289, 217, 422, 147, 272, and 331 independent significant SNPs for the brain, cardiovascular, eye, hepatic, immune, metabolic, musculoskeletal, pulmonary, and renal BAGs, and 27, 124, 69, 292, 218, 422, 148, 269, and 333 independent significant SNPs in fastGWA GWAS.

For all the nine BAGs, we found almost perfect concordance between the PLINK and fastGWA GWASs using the three proposed metrics ($r$-$\beta$=1; $C$-$\beta$=1; $P$-$\beta$=1). Detailed results of these SNPs are presented in **Supplementary eFile 4** for method-specific GWAS. The scatter plot of the independent SNPs' $\beta$ coefficients is shown below.

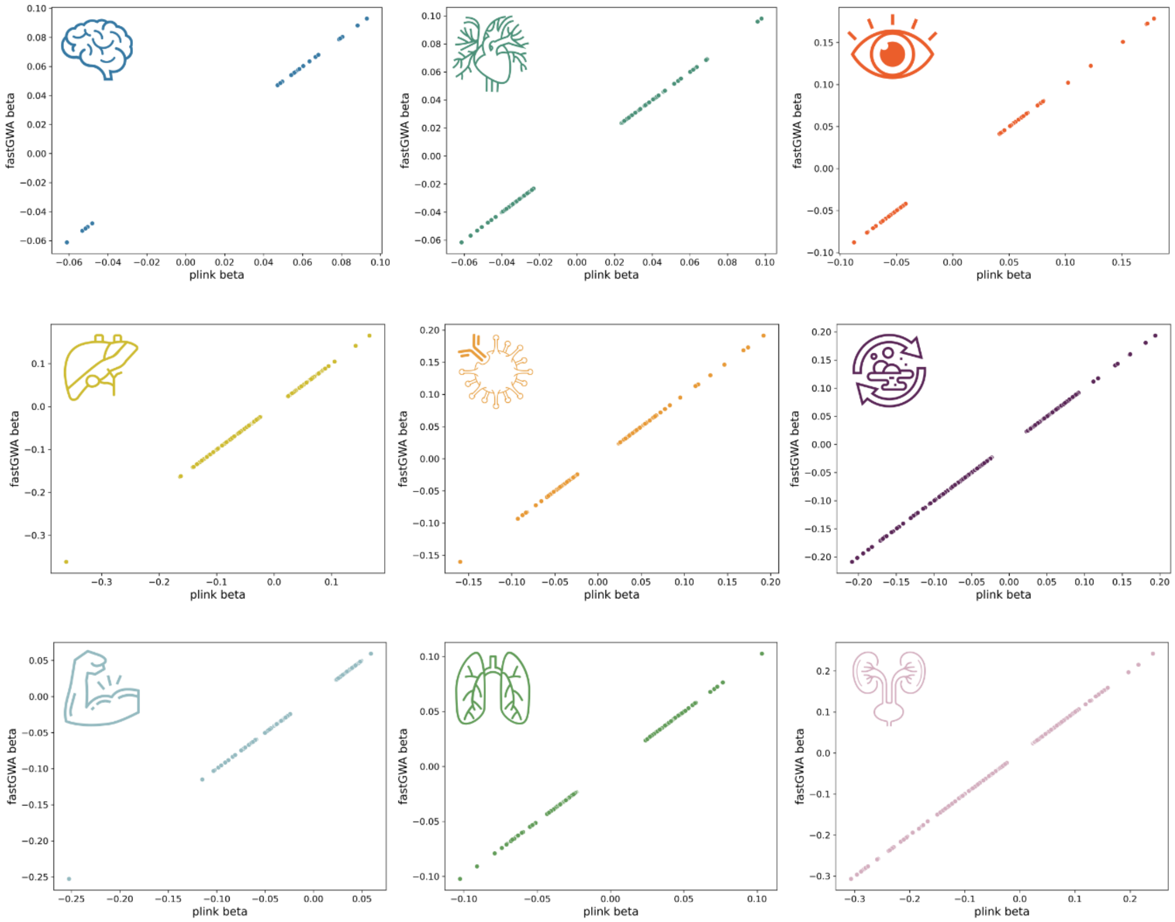

The figures present the scatter plots for the two sets of beta coefficients estimated from different GWAS methods.

**European vs. non-European GWAS**

In the European GWAS, we found 27, 124, 69, 289, 217, 422, 147, 272, and 331 independent significant SNPs for the brain, cardiovascular, eye, hepatic, immune, metabolic, musculoskeletal, pulmonary, and renal BAGs, and 0, 2, 1, 16, 2, 23, 1, 1, and 35 independent significant SNPs in non-European GWAS (with much smaller sample sizes).

For all the nine BAGs, we found a high concordance between the European and non-Eurorpean GWASs using the three proposed metrics (0.85<$r$-$\beta$<0.95; 0.89<$C$-$\beta$<1). The two sets of $\beta$ coefficients did not significantly differ ($P$-$\beta$>0.12). Detailed results of these SNPs are presented in **Supplementary eFile 5** for ancestry-specific GWAS. The scatter plot of the independent SNPs' $\beta$ coefficients is shown below.

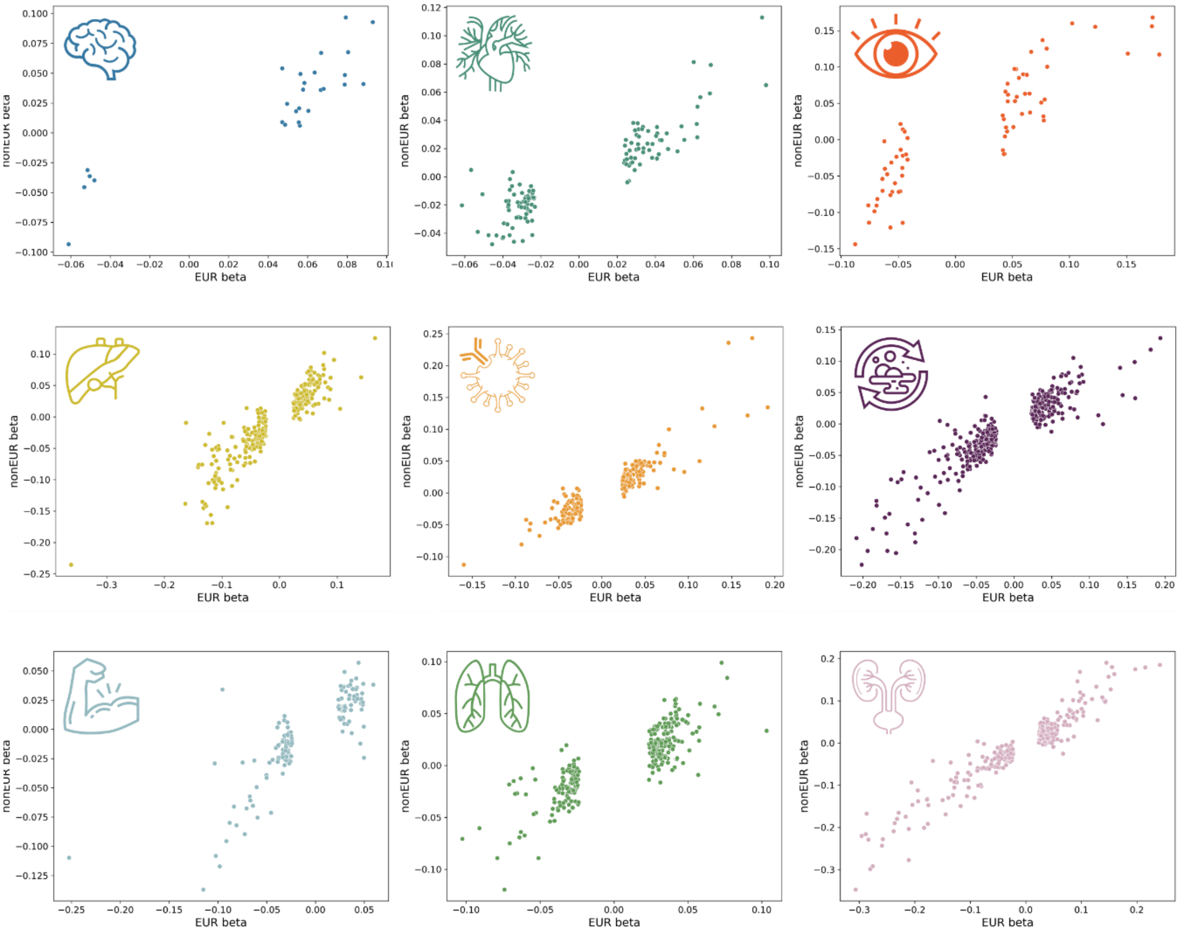

The figures present the scatter plots for the two sets of beta coefficients estimated from different GWAS ancestry groups.

**eText 2: Phenome-wide association query using the GWAS Atlas platform**

To comprehensively encompass the genetic landscape reported in previous literature, we comparatively conducted a phenome-wide association query using the GWAS Atlas platform (https://atlas.ctglab.nl/PheWAS). We applied the same P-value threshold search criteria as those used in the EMBL-EBI GWAS Catalog (P-value<$1x10^{-5}$). These findings are presented as a supplementary search to complement the results shown in **Fig. 2a**. The details of this comparative search are presented in **Supplementary eFile 7**.

It's important to note that the two platforms may exhibit variations in their curated GWAS datasets, the genome build versions utilized, and the specific P-value thresholds set for their search analyses by default. We tried our best to harmonize the query criteria. Hence, this comparative search was not exhaustive, and the results may differ. Rather, we intend to offer a broad overview of the two platforms commonly employed for phenome-wide association studies (PheWAS). Given the rapid updates in GWAS summary statistics in the field, it's worth mentioning that this comparative search was originally conducted on October 23, 2023, and revised on January 13, 2024, based on the reviewer's comments. The results from the GWAS Atlas are shown in the figure below.

In the GWAS Atlas platform, we identified 8,576 significant associations between the identified loci in our GWAS and clinical traits. The genomic loci associated with the brain BAG exhibited the highest proportion of associations (109 out of 308) with traits related to the brain. The brain BAG loci were also largely linked to many other traits related to other organ systems, evidencing inter-organ connections, including metabolic (*N*=78/308), lifestyle factor (*N*=13/308), neurodegenerative traits (*N*=5/308), and immune (*N*=35/308). For the eye BAG loci, most associations were found in the musculoskeletal (*N*=139/279), eye (*N*=14/279), and mental traits (*N*=19/279), among many others.

For the seven body organ systems, among the loci associated with the cardiovascular BAG, most associations were observed with musculoskeletal traits (*N*=249/611) and cardiovascular traits (166/611). 29 out of 1009 associations were related to hepatic traits (e.g., blood protein, cirrhosis, and bilirubin) for the hepatic BAG loci. Among the loci associated with the immune BAG, abundant associations were found enriched in immune (*N*=467/1062) traits. For the metabolic BAG loci, most associations were observed in metabolic traits (*N*=993/1990). We found a significant intertwining of musculoskeletal systems with other organ systems in the GWAS Atlas platform. Details of the phenome-wide associations are presented in **Supplementary eFile 7**.

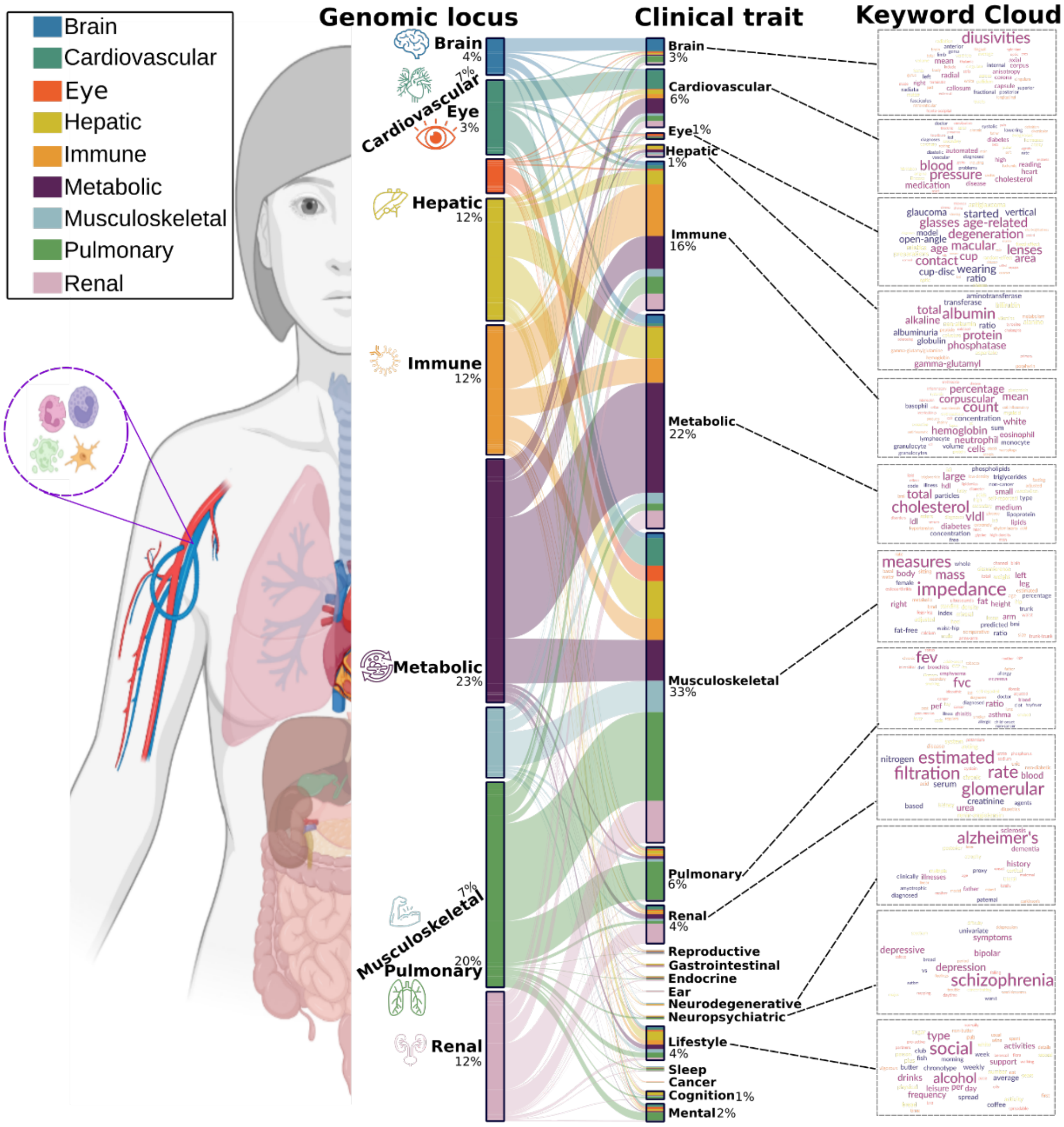

**Figure**. We queried the clumped independent significant SNPs using the PheWAS functionaly provided by the GWAS Atlas platforms.

### eText 3: Sensitivity check analyses for the causality between the hepatic BAG and musculoskeletal BAG

#### A) Sensitivity analyses on body weight for the bi-directional causality between the hepatic and musculoskeletal BAGs

We conducted a revised Mendelian randomization analysis by introducing body weight as a covariate in the split-sample GWASs for hepatic and musculoskeletal BAGs. In this approach, we employed hepatic BAG as the exposure variable in split1 GWAS and musculoskeletal BAG as the outcome variable in split2 GWAS. Likewise, we reversed the roles, using musculoskeletal BAG as the exposure variable in split1 GWAS and hepatic BAG as the outcome variable in split2 GWAS, thus assessing the inverse causal relationship. This methodology ensured the absence of overlapping populations while effectively controlling for the influence of body weight.

Compared to the original results, this bi-directional causality persisted while adjusting the body weight as a covariate, shown in the tables below:

1) **GWAS without and with body weight as a covariate for the causal relationship from the hepatic BAG to the musculoskeletal BAG.**

| Weight | Outcome (split2) | Exposure (split1) | Method | nSNP | BETA | SE | P | OR | CI_low | CI_high |
|---|---|---|---|---|---|---|---|---|---|---|
| N | Musculoskeletal | Hepatic | MR Egger | 19 | 0.51783336 | 0.14070786 | 0.00185593 | 1.67838725 | 1.27385274 | 2.21138886 |
| | Musculoskeletal | Hepatic | Weighted median | 19 | 0.35295633 | 0.06606437 | 9.16E-08 | 1.42326899 | 1.25040832 | 1.62002649 |
| | Musculoskeletal | Hepatic | Inverse variance weighted | 19 | 0.38344296 | 0.07834137 | 9.85E-07 | 1.46732785 | 1.25846644 | 1.71085295 |
| | Musculoskeletal | Hepatic | Simple mode | 19 | 0.15733154 | 0.10700058 | 0.15872332 | 1.17038357 | 0.94895908 | 1.44347395 |
| | Musculoskeletal | Hepatic | Weighted mode | 19 | 0.46614953 | 0.08121762 | 1.93E-05 | 1.59384531 | 1.35929067 | 1.86887391 |
| Y | Musculoskeletal | Hepatic | MR Egger | 18 | 0.51517011 | 0.14245065 | 0.00231711 | 1.67392323 | 1.26613232 | 2.21305384 |
| | Musculoskeletal | Hepatic | Weighted median | 18 | 0.35613857 | 0.06002398 | 2.97E-09 | 1.42780539 | 1.26933301 | 1.60606258 |
| | Musculoskeletal | Hepatic | Inverse variance weighted | 18 | 0.38926537 | 0.0792834 | 9.12E-07 | 1.47589615 | 1.2634801 | 1.72402356 |
| | Musculoskeletal | Hepatic | Simple mode | 18 | 0.24697399 | 0.11293776 | 0.04302518 | 1.28014581 | 1.02594689 | 1.59732761 |
| | Musculoskeletal | Hepatic | Weighted mode | 18 | 0.47542746 | 0.06925444 | 2.74E-06 | 1.60870171 | 1.40451037 | 1.84257891 |

2) **GWAS without and with body weight as a covariate for the causal relationship from the musculoskeletal BAG to the hepatic BAG.**

| Weight | Outcome (split2) | Exposure (split1) | Method | nSNP | BETA | SE | P | OR | CI_low | CI_high |
|---|---|---|---|---|---|---|---|---|---|---|
| N | Hepatic | Musculoskeletal | MR Egger | 9 | 1.8282501 | 0.24293965 | 0.00013439 | 6.22298749 | 3.8654897 | 10.0182839 |
| | Hepatic | Musculoskeletal | Weighted median | 9 | 0.92114305 | 0.13768954 | 2.23E-11 | 2.51216028 | 1.9179781 | 3.2904178 |
| | Hepatic | Musculoskeletal | Inverse variance weighted | 9 | 1.02402966 | 0.18103365 | 1.54E-08 | 2.78439235 | 1.9526818 | 3.97035541 |

| | | | | | | | | | | |
|---|---|---|---|---|---|---|---|---|---|---|
| | Hepatic | Musculoskeletal | Simple mode | 9 | 1.20577311 | 0.18620161 | 0.000193 | 3.33933976 | 2.31826245 | 4.81014995 |
| | Hepatic | Musculoskeletal | Weighted mode | 9 | 1.25833413 | 0.13034769 | 1.10E-05 | 3.51955347 | 2.7260472 | 4.54403601 |
| Y | Hepatic | Musculoskeletal | MR Egger | 9 | 1.69092352 | 0.35916855 | 0.00218827 | 5.42448802 | 2.68304718 | 10.9670342 |
| | Hepatic | Musculoskeletal | Weighted median | 9 | 0.85408009 | 0.13197703 | 9.71E-11 | 2.34921232 | 1.81376558 | 3.04272978 |
| | Hepatic | Musculoskeletal | Inverse variance weighted | 9 | 0.99179962 | 0.19767923 | 5.24E-07 | 2.69608204 | 1.83005923 | 3.97192521 |
| | Hepatic | Musculoskeletal | Simple mode | 9 | 1.23665687 | 0.15851732 | 5.23E-05 | 3.44408019 | 2.52429777 | 4.6990052 |
| | Hepatic | Musculoskeletal | Weighted mode | 9 | 1.27628794 | 0.1538585 | 3.36E-05 | 3.58331353 | 2.65043899 | 4.84453174 |

**B) Sensitivity analysis for the hepatic BAG on musculoskeletal BAG excluding the *APOE* gene**

We conducted a revised Mendelian randomization analysis by excluding SNPs within the *APOE* gene for the causal relationship from the hepatic BAG to the musculoskeletal BAGs; all other significant causality did not involve the two common *APOE* gene SNPs (rs429358 and rs7412). In this approach, we employed hepatic BAG as the exposure variable in split1 GWAS and musculoskeletal BAG as the outcome variable in split2 GWAS.

Compared to the original results, this causality persisted while excluding the SNP (rs429358) as an IV, shown in the tables below:

**GWAS without and with rs429358 as an IV for the causal relationship from the hepatic BAG to the musculoskeletal BAG.**

| rs429358 | Outcome (split2) | Exposure (split1) | Method | nSNP | BETA | SE | P | OR | CI_low | CI_high |
|---|---|---|---|---|---|---|---|---|---|---|
| N | Musculoskeletal | Hepatic | MR Egger | 18 | 0.51522659 | 0.12736616 | 0.00093844 | 1.67401778 | 1.30419881 | 2.14870271 |
| | Musculoskeletal | Hepatic | Weighted median | 18 | 0.36478773 | 0.06339608 | 8.71E-09 | 1.44020827 | 1.27192489 | 1.63075657 |
| | Musculoskeletal | Hepatic | Inverse variance weighted | 18 | 0.41660503 | 0.07146014 | 5.55E-09 | 1.5168033 | 1.31856385 | 1.74484706 |
| | Musculoskeletal | Hepatic | Simple mode | 18 | 0.15924454 | 0.09710274 | 0.11938508 | 1.17262466 | 0.96940109 | 1.41845167 |
| | Musculoskeletal | Hepatic | Weighted mode | 18 | 0.45942325 | 0.07899932 | 2.07E-05 | 1.58316063 | 1.35606155 | 1.84829191 |
| Y | Musculoskeletal | Hepatic | MR Egger | 19 | 0.51783336 | 0.14070786 | 0.00185593 | 1.67838725 | 1.27385274 | 2.21138886 |
| | Musculoskeletal | Hepatic | Weighted median | 19 | 0.35295633 | 0.06606437 | 9.16E-08 | 1.42326899 | 1.25040832 | 1.62002649 |
| | Musculoskeletal | Hepatic | Inverse variance weighted | 19 | 0.38344296 | 0.07834137 | 9.85E-07 | 1.46732785 | 1.25846644 | 1.71085295 |
| | Musculoskeletal | Hepatic | Simple mode | 19 | 0.15733154 | 0.10700058 | 0.15872332 | 1.17038357 | 0.94895908 | 1.44347395 |
| | Musculoskeletal | Hepatic | Weighted mode | 19 | 0.46614953 | 0.08121762 | 1.93E-05 | 1.59384531 | 1.35929067 | 1.86887391 |

**C) Sensitivity analyses for metabolic BAG on body weight**

We showcased sensitivity analyses to investigate potential violations of the three IV assumptions (**Method 3j**). To illustrate this, we showcased the sensitivity analysis results for the causal effect of the metabolic BAG on body weight (**Supplementary eFigure 33**). In a leave-one-out analysis, no single SNP overwhelmingly drove the overall effect. There was evidence for minor heterogeneity[1] of the causal effect amongst SNPs (Cochran's Q value=57.33, P-value<1x10$^{-5}$). Some SNPs exerted opposite causal effects compared to the model using all SNPs. The scatter plot indicated two obvious SNP outliers (rs117233107 and rs33959228), and the funnel plot showed slight asymmetry. Finally, the MR Egger estimator allows for pleiotropic effects independent of the effect on the exposure of interest (i.e., the InSIDE assumption[2]). Our results from the Egger estimator showed a small but not significant positive intercept (3.62x10$^{-4}$±1.67x10$^{-3}$, P-value=0.83), which may indicate that the IVW estimate is not likely biased[2]. We re-analyzed the IVW MR analyses by excluding the two outliers identified in **Supplementary eFigure 33** (rs117233107 and rs33959228), which led to a similar OR [0.94 (0.91, 0.97) vs. 0.95 (0.92, 0.98)] and a less significant P-value [6.9x10$^{-4}$ vs. 1.2x10$^{-3}$].

**eFigure 1: GWAS Manhattan plots for the brain BAG**

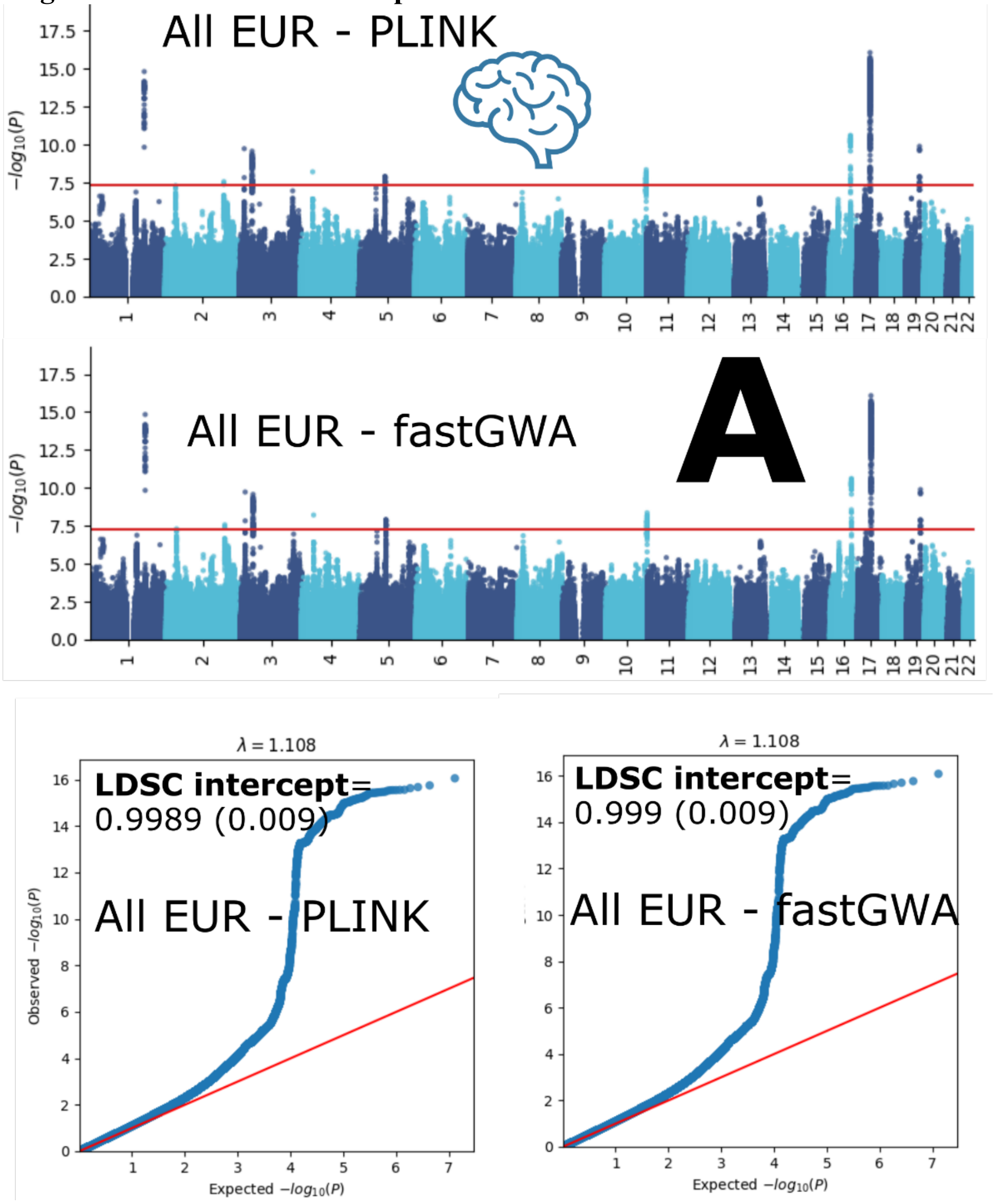

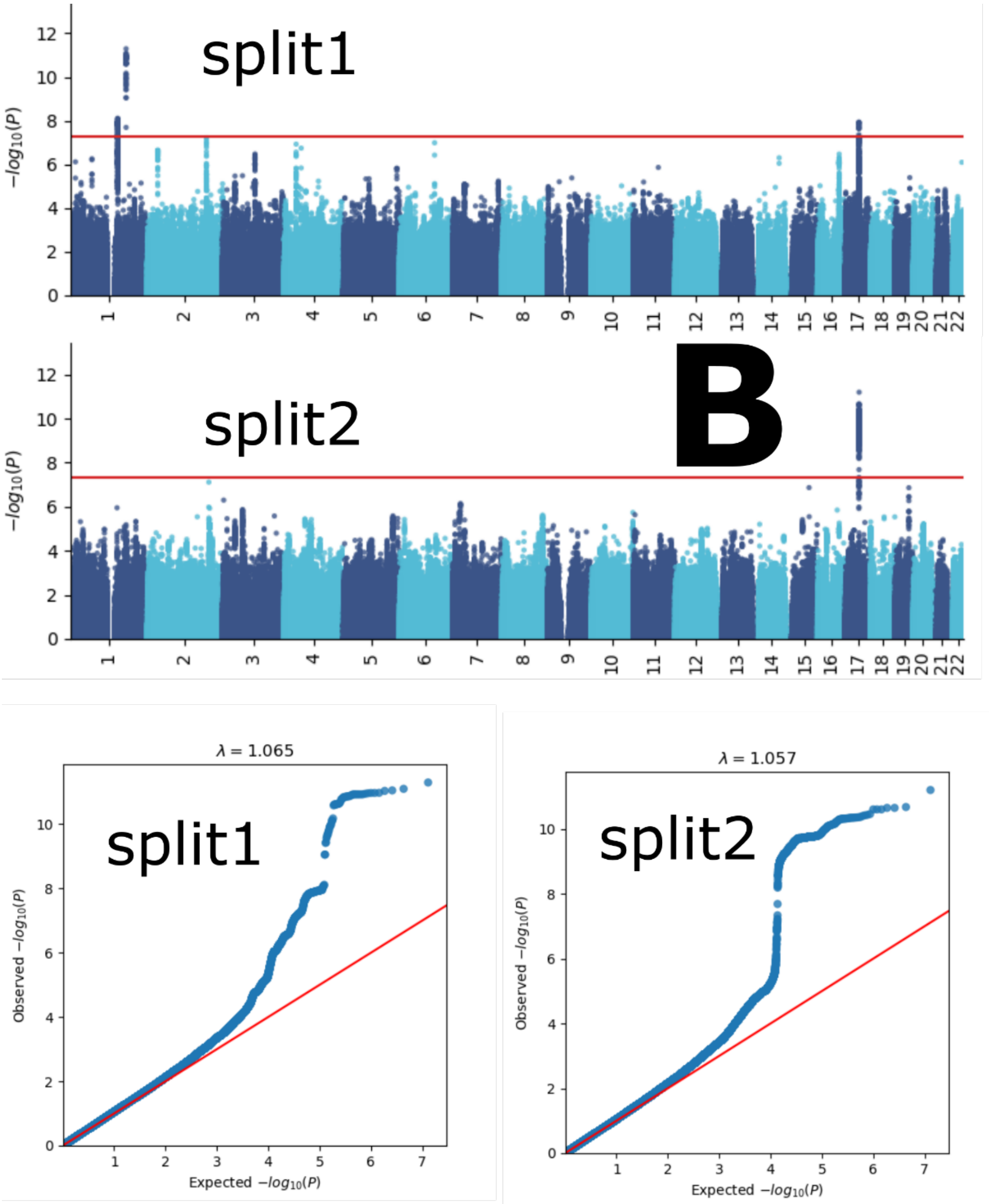
split1
split2
B
$-log_{10}(P)$
1 2 3 4 5 6 7 8 9 10 11 12 13 14 15 16 17 18 19 20 21 22
λ = 1.065
λ = 1.057
Observed $-log_{10}(P)$
Expected $-log_{10}(P)$

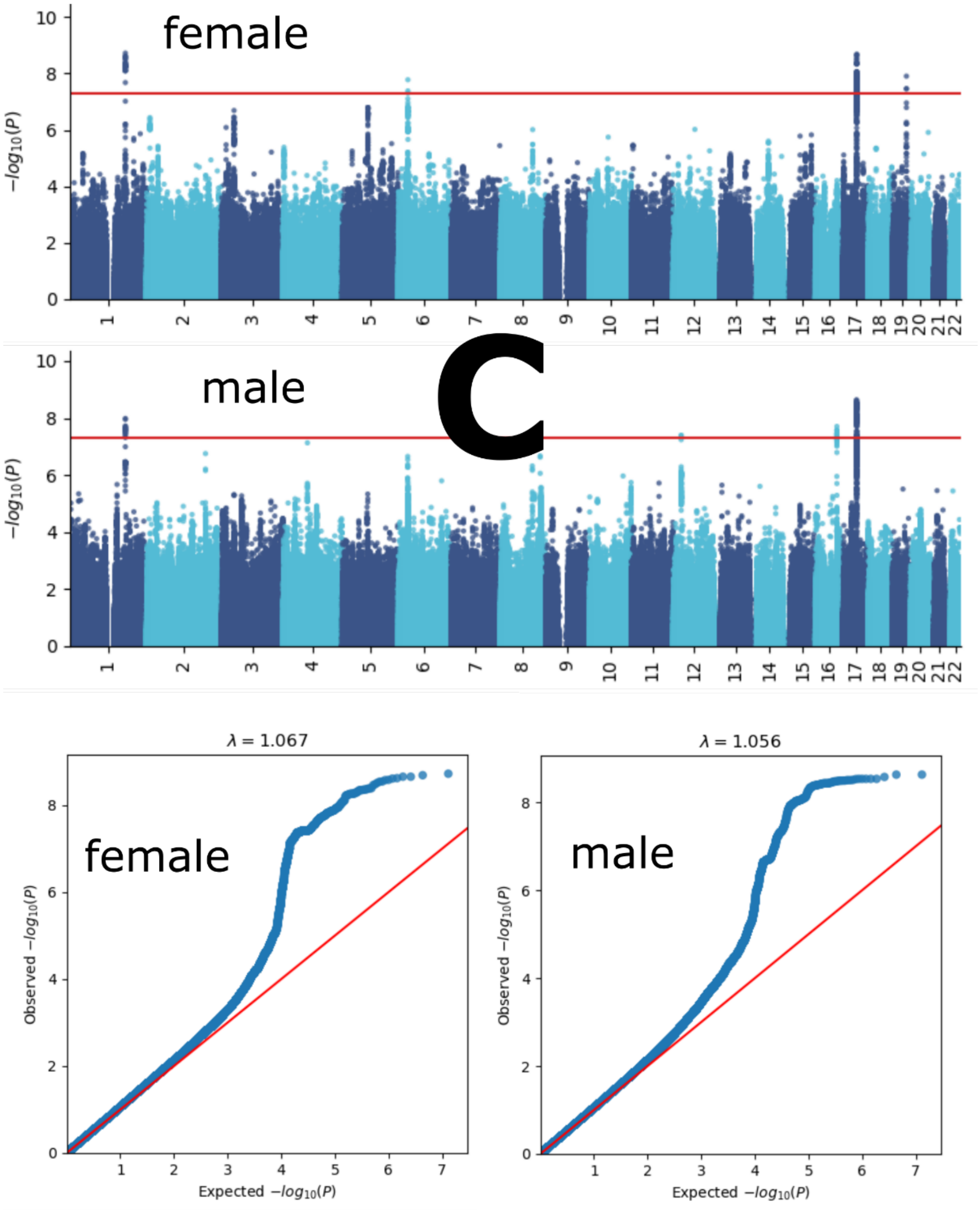
female
male
C
$-log_{10}(P)$
λ = 1.067
λ = 1.056
female
male
Observed $-log_{10}(P)$
Expected $-log_{10}(P)$

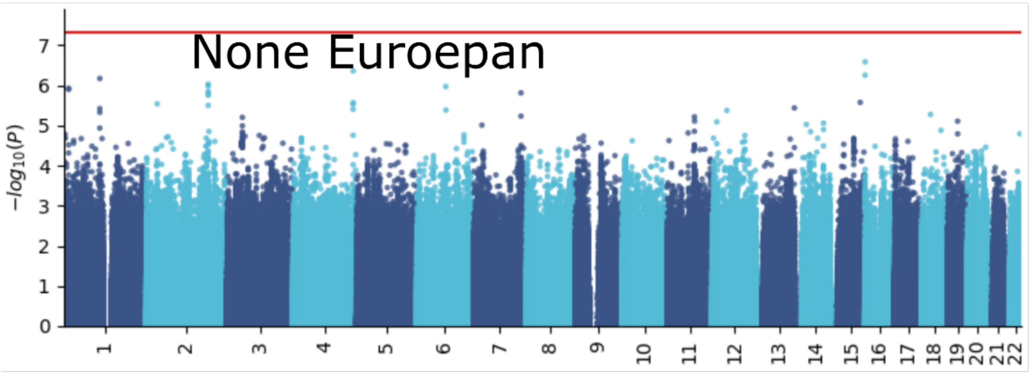

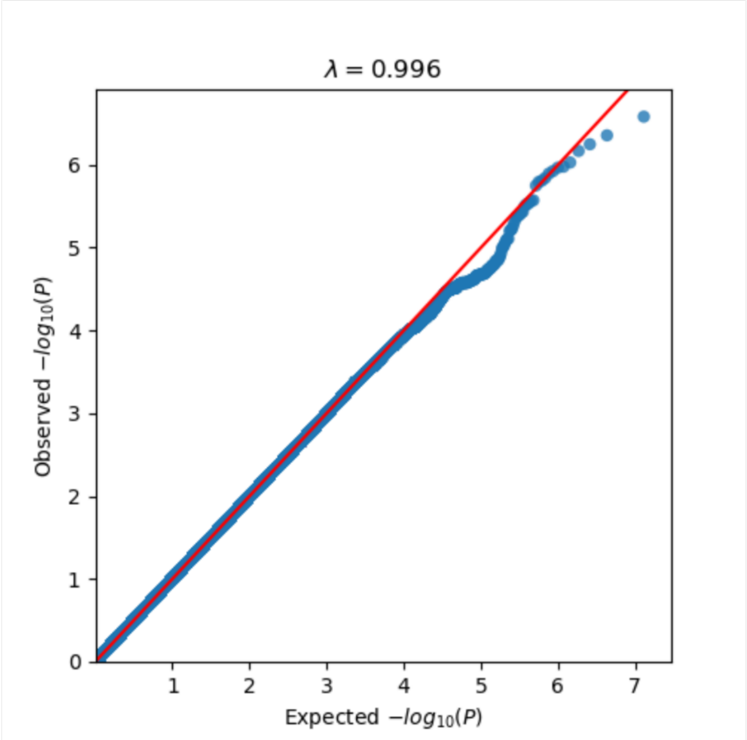

**D**

Manhattan and QQ plots, along with genomic inflation factors and LDSC intercepts, are displayed for the primary GWAS conducted on individuals of European ancestry (*N*=30,062) using PLINK and fastGWA (**A**). Additionally, results are presented for split-sample GWAS (split1 and split2, **B**), sex-stratified GWAS (female and male, **C**), and GWAS involving non-European ancestry populations (*N*=4465, **D**).

**eFigure 2: GWAS Manhattan plots for the cardiovascular BAG**

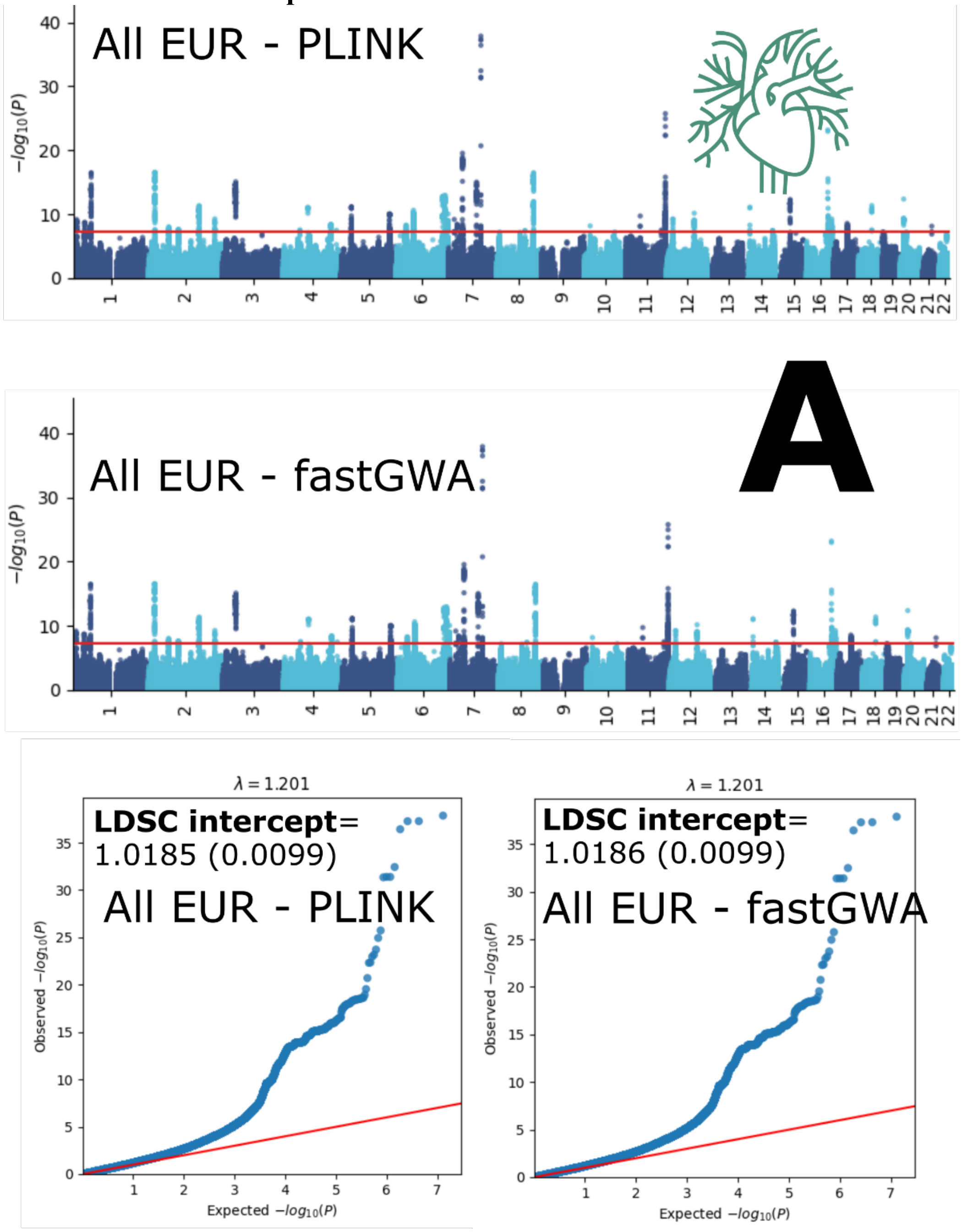

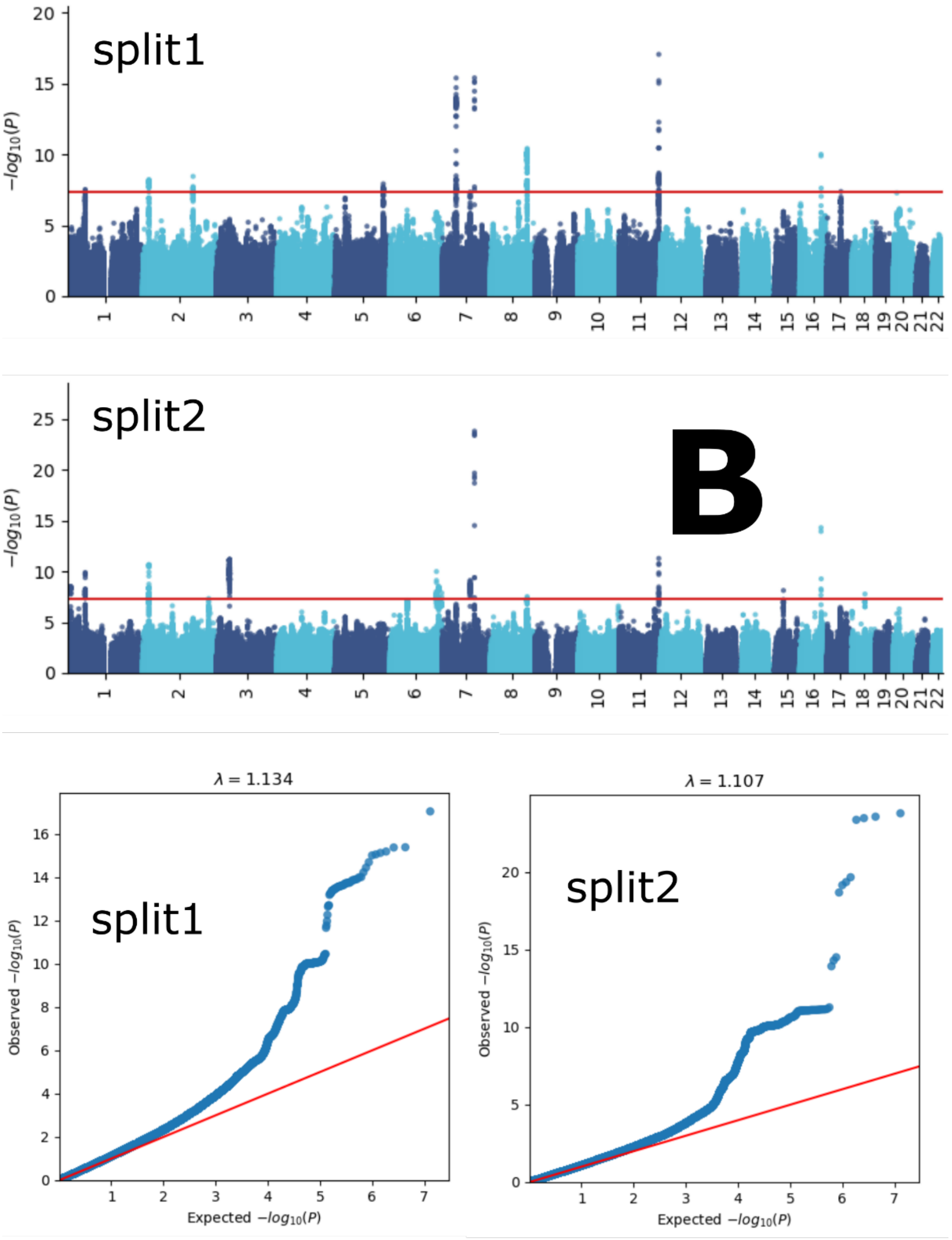

split1
split2
B
$-log_{10}(P)$
λ = 1.134
λ = 1.107
Observed $-log_{10}(P)$
Expected $-log_{10}(P)$

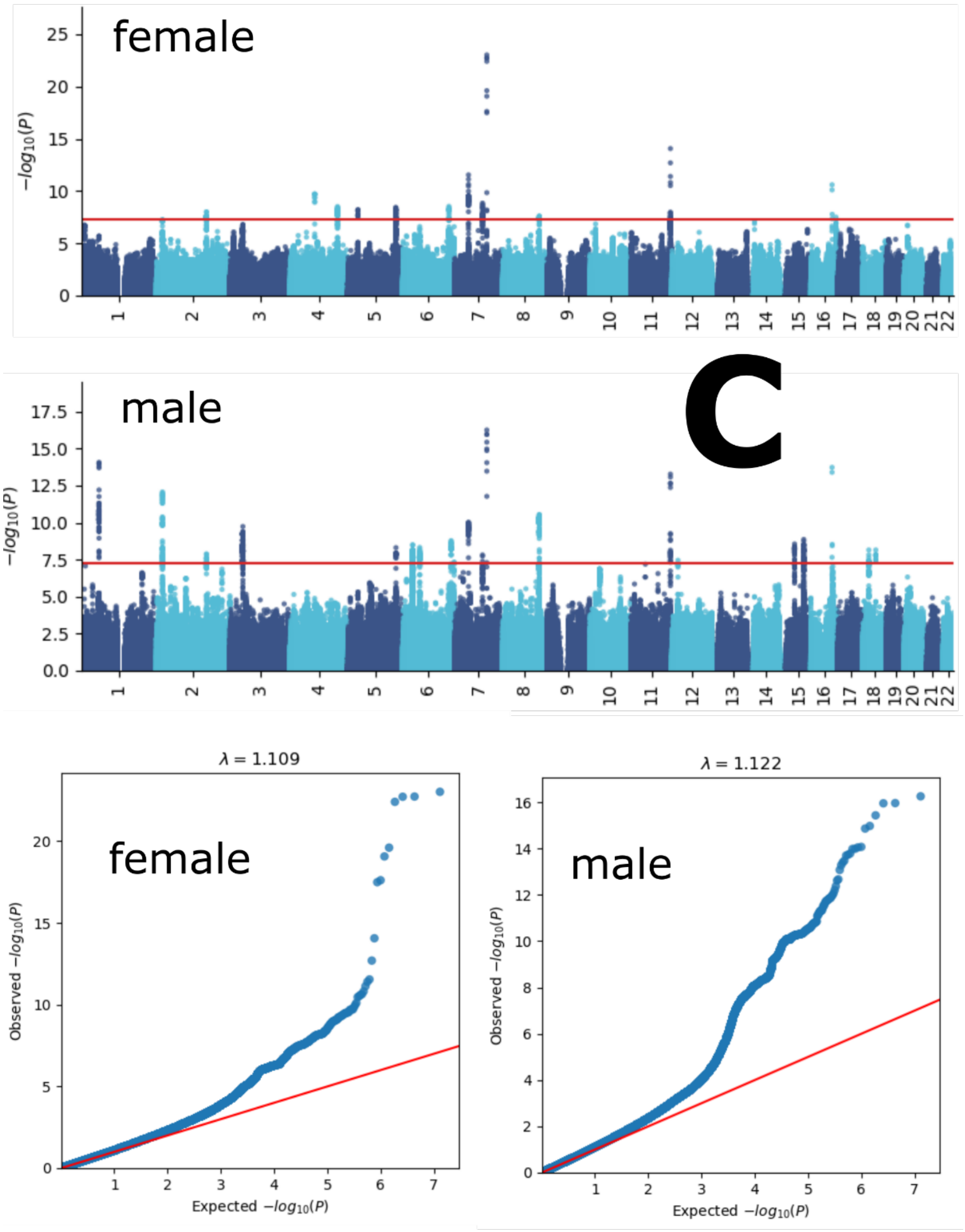
female
−log10(P)
male
C
λ = 1.109
λ = 1.122
Observed −log10(P)
Expected −log10(P)

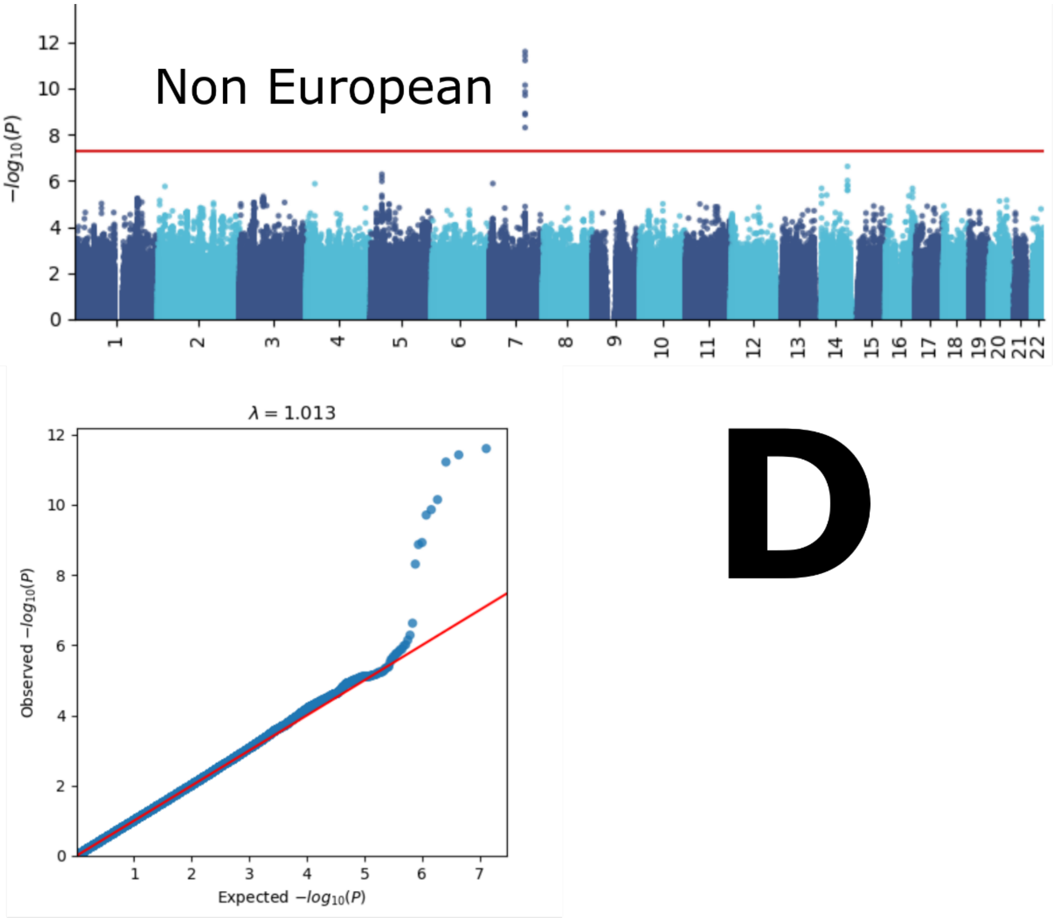

Manhattan and QQ plots, along with genomic inflation factors and LDSC intercepts, are displayed for the primary GWAS conducted on individuals of European ancestry (*N*=111,386) using PLINK and fastGWA (**A**). Additionally, results are presented for split-sample GWAS (split1 and split2, **B**), sex-stratified GWAS (female and male, **C**), and GWAS involving non-European ancestry populations (*N*=20,408, **D**).

**eFigure 3: GWAS Manhattan plots for the eye BAG**

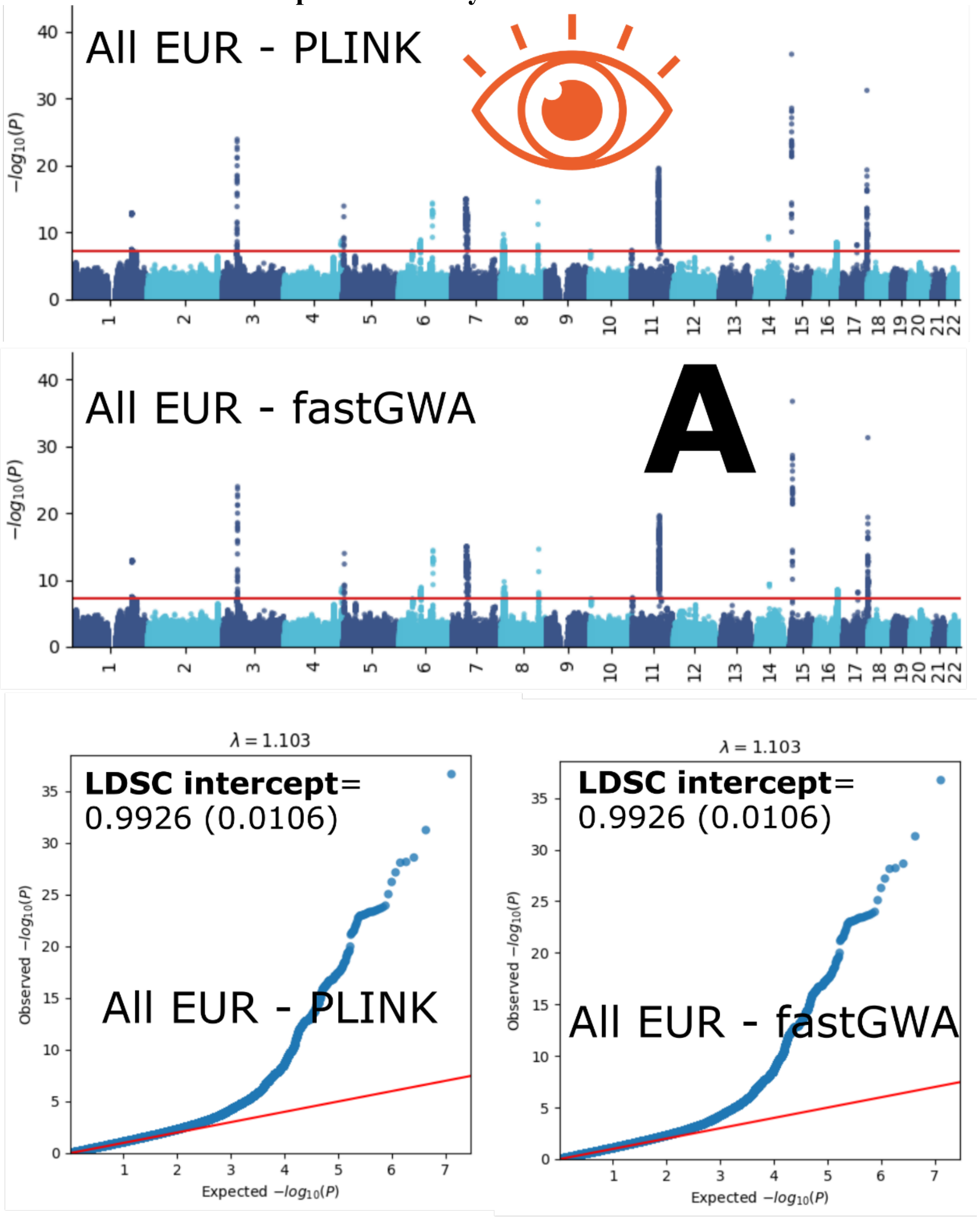

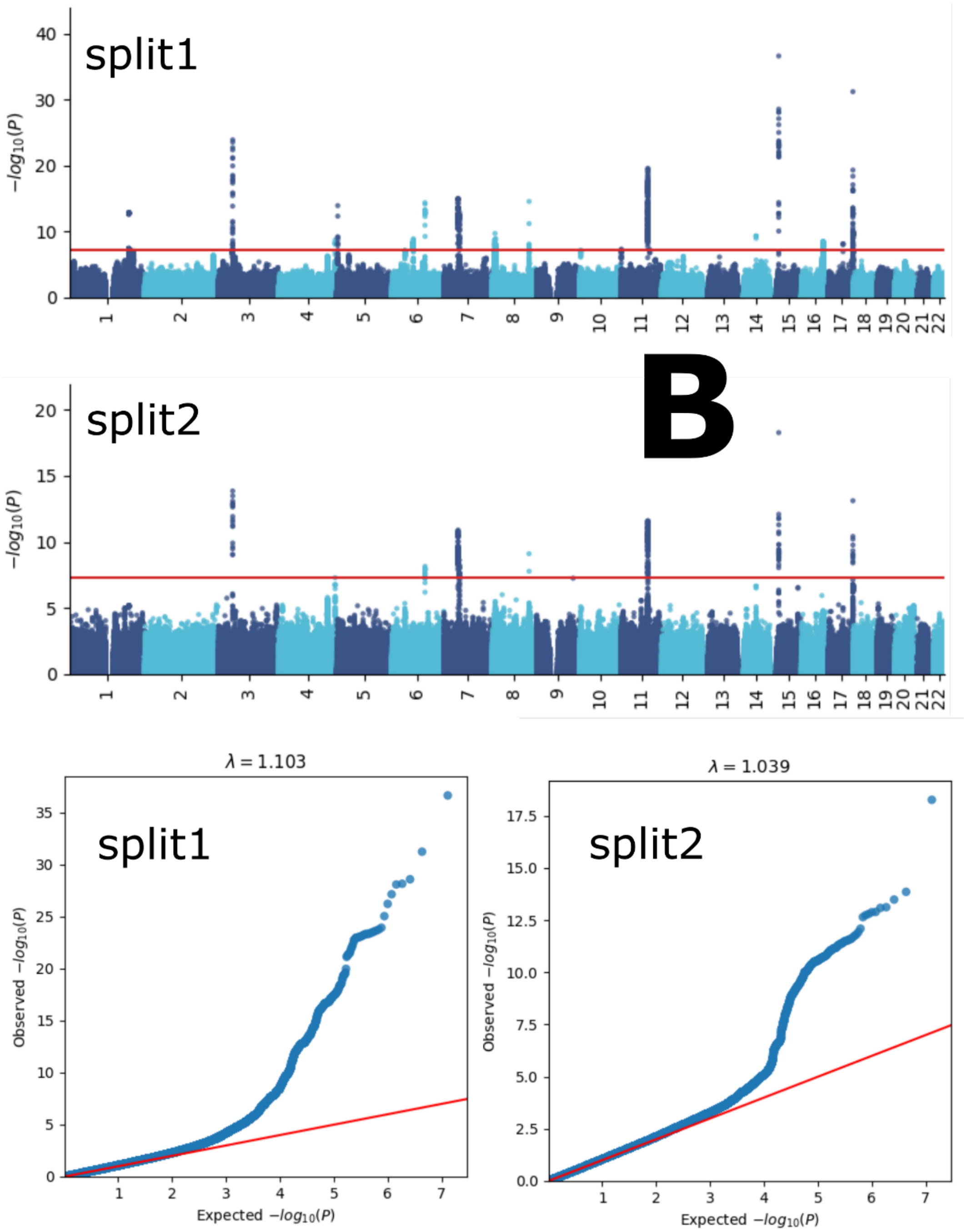
split1
split2
B
$-log_{10}(P)$
$\lambda = 1.103$
$\lambda = 1.039$
Observed $-log_{10}(P)$
Expected $-log_{10}(P)$

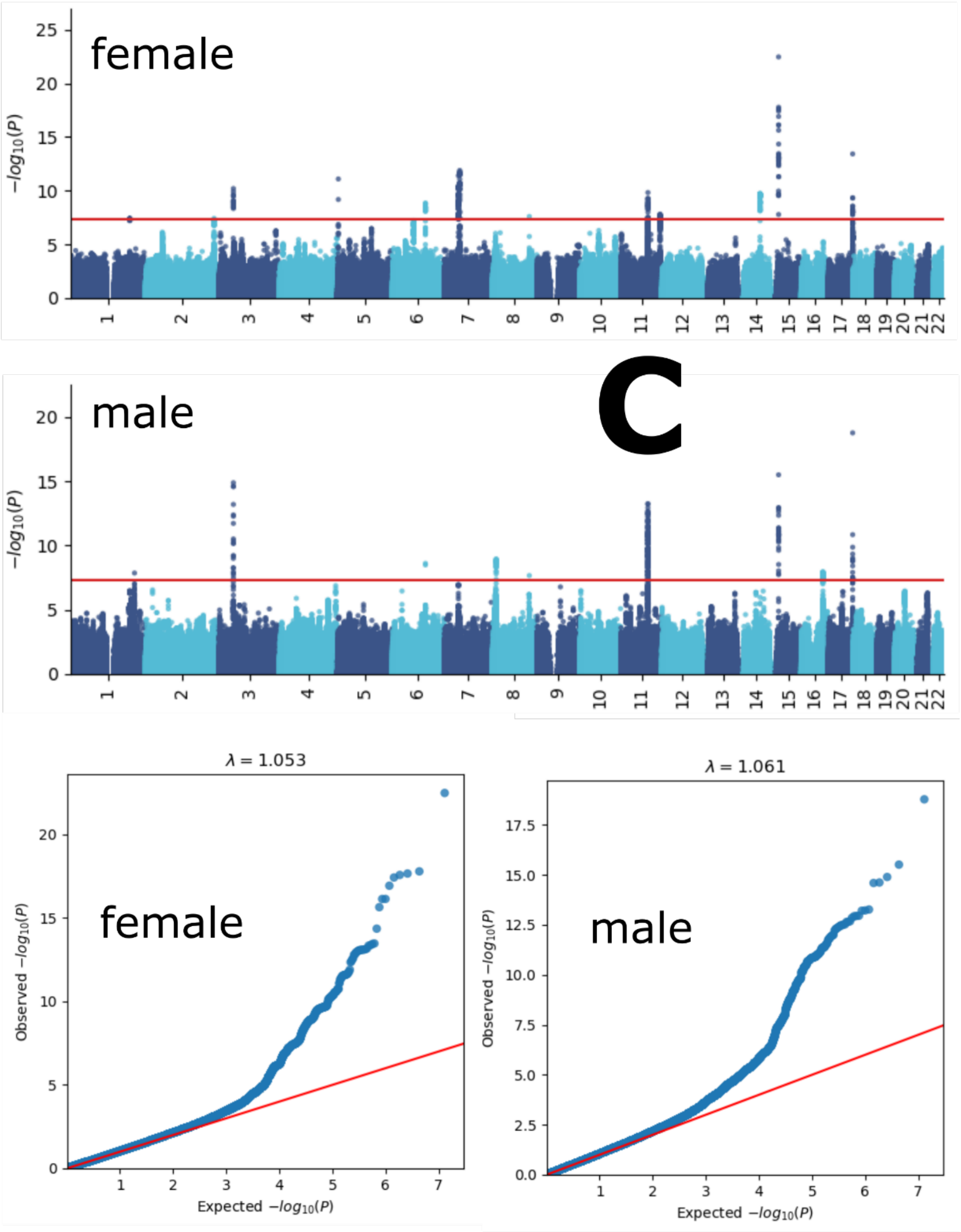
female
male
C
λ = 1.053
λ = 1.061
female
male
Observed −log10(P)
Expected −log10(P)
−log10(P)

307

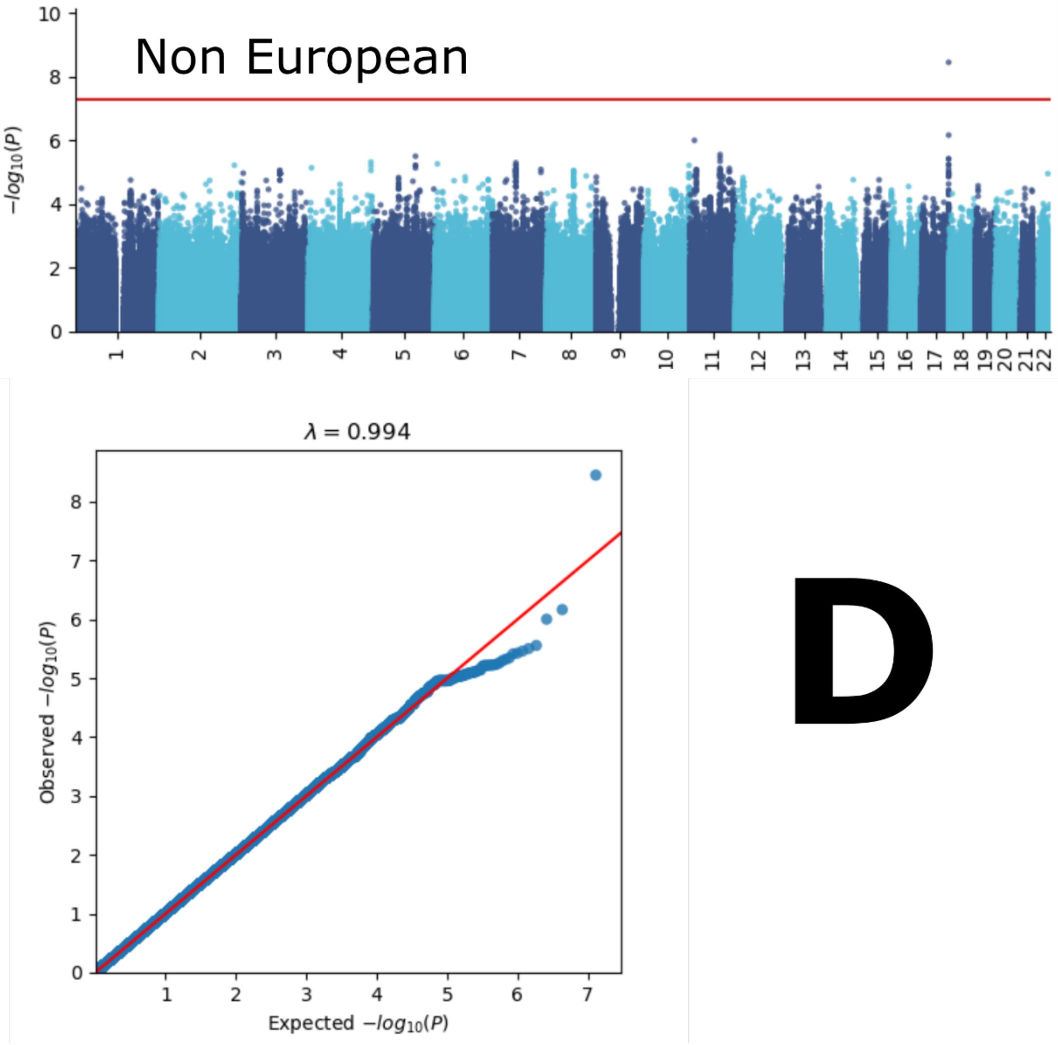

Manhattan and QQ plots, along with genomic inflation factors and LDSC intercepts, are displayed for the primary GWAS conducted on individuals of European ancestry (*N*=36,004) using PLINK and fastGWA (**A**). Additionally, results are presented for split-sample GWAS (split1 and split2, **B**), sex-stratified GWAS (female and male, **C**), and GWAS involving non-European ancestry populations (*N*=3407, **D**).

314 **eFigure 4: GWAS Manhattan plots for the hepatic BAG**

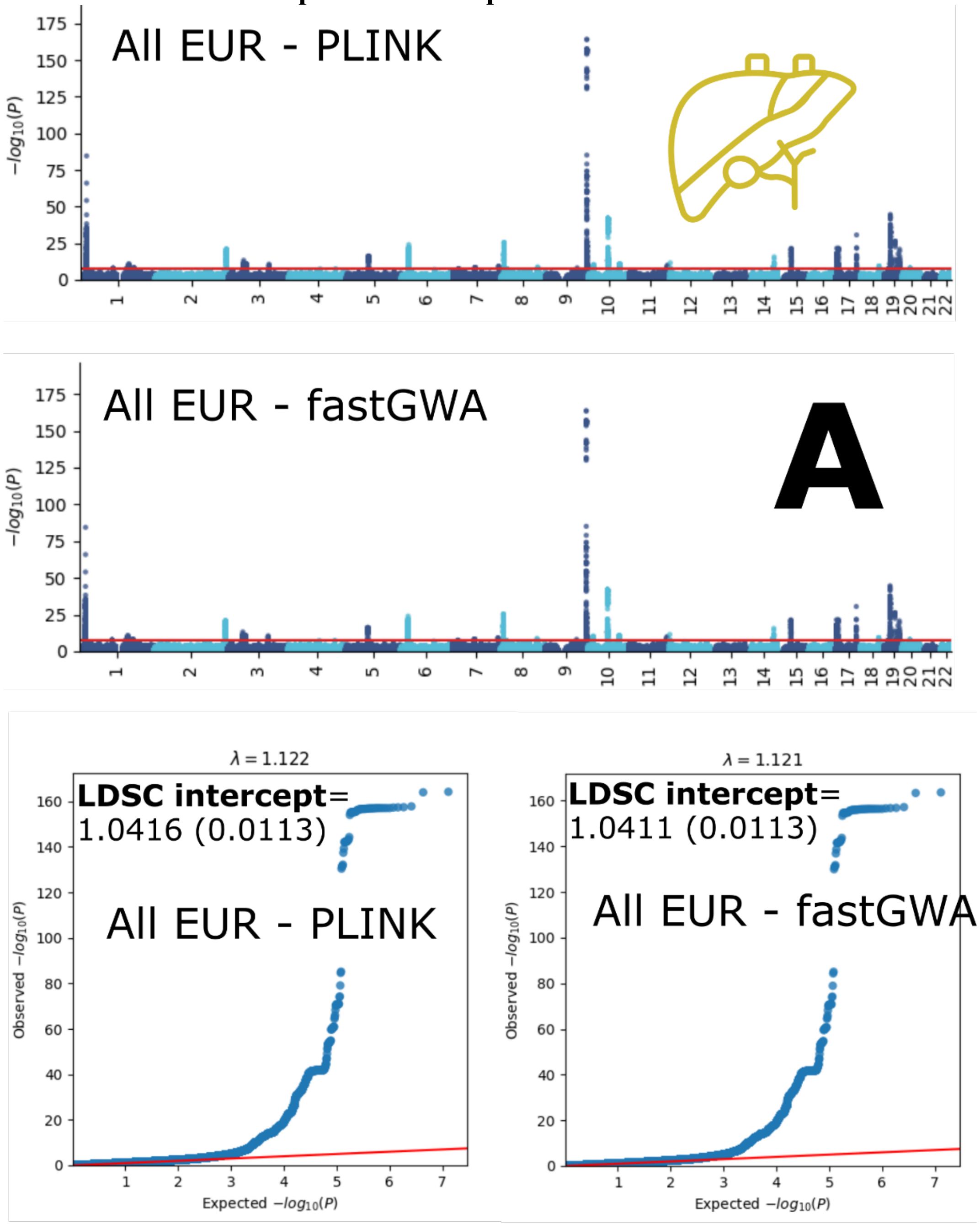

315

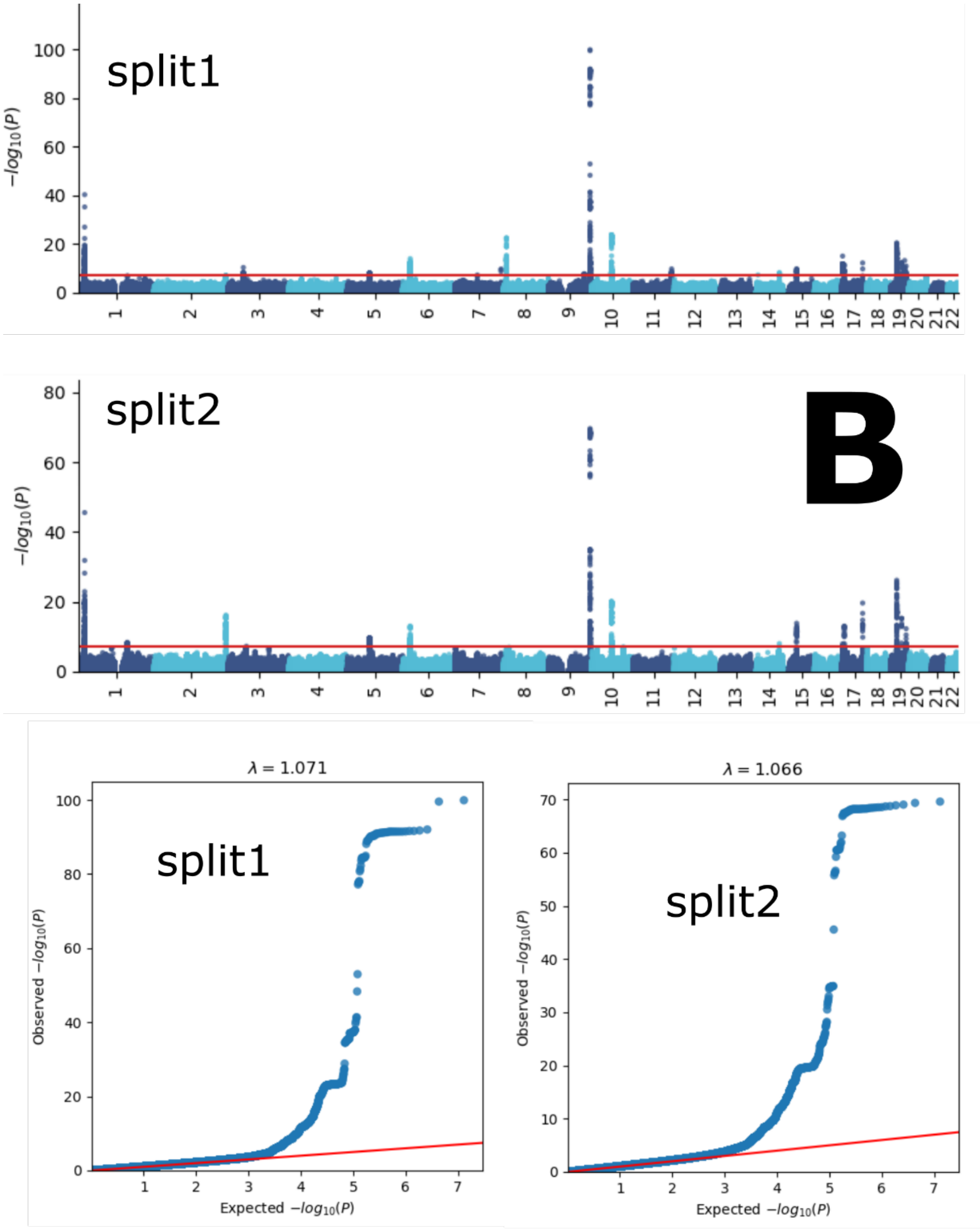
split1
split2
B
$-log_{10}(P)$
λ = 1.071
λ = 1.066
Observed $-log_{10}(P)$
Expected $-log_{10}(P)$

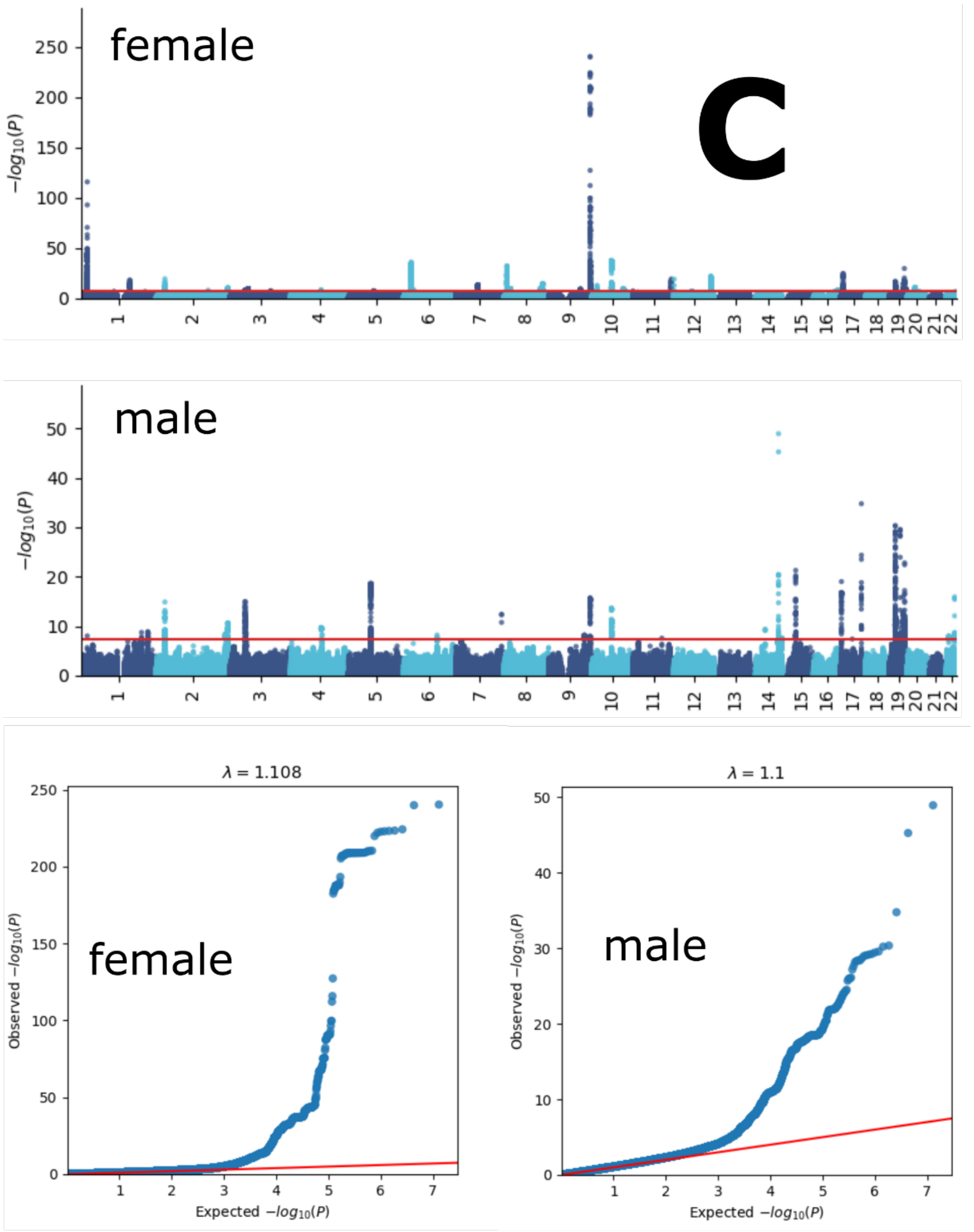
female
C
male
$-log_{10}(P)$
$\lambda = 1.108$
$\lambda = 1.1$
female
male
Observed $-log_{10}(P)$
Expected $-log_{10}(P)$

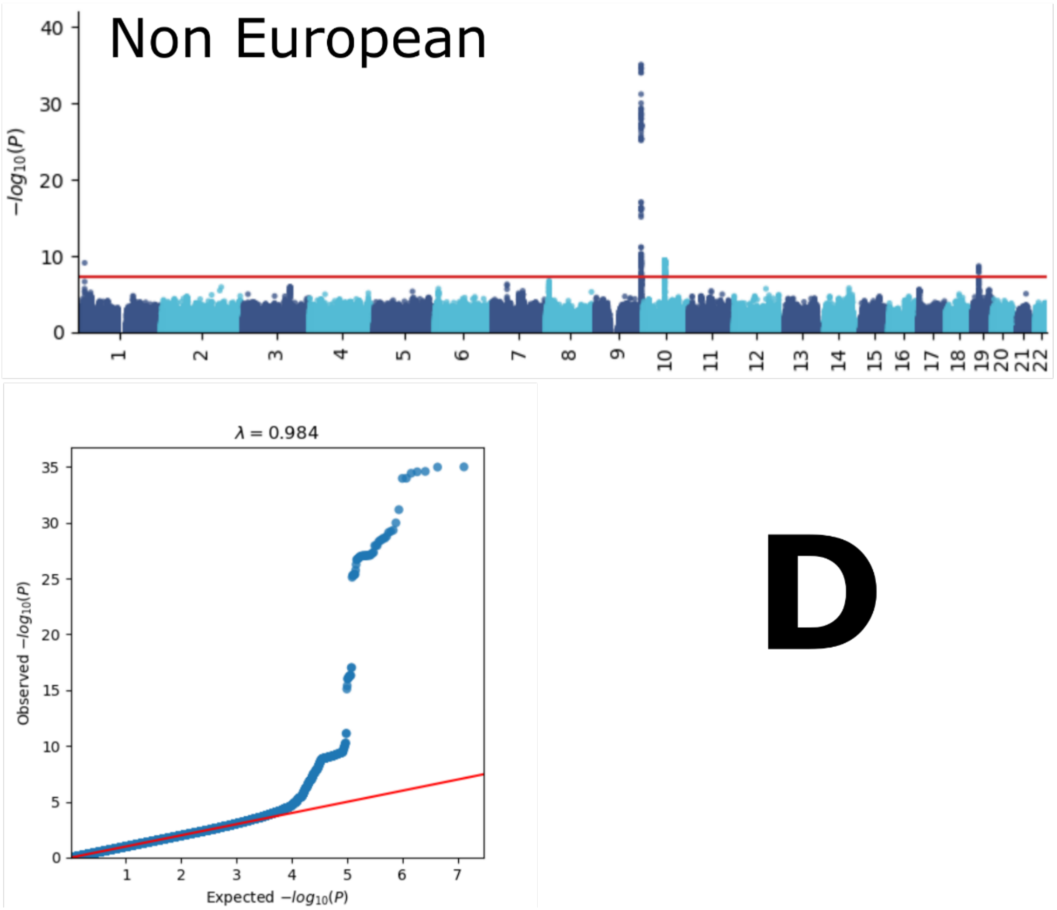

Manhattan and QQ plots, along with genomic inflation factors and LDSC intercepts, are displayed for the primary GWAS conducted on individuals of European ancestry (*N*=111,386) using PLINK and fastGWA (**A**). Additionally, results are presented for split-sample GWAS (split1 and split2, **B**), sex-stratified GWAS (female and male, **C**), and GWAS involving non-European ancestry populations (*N*=20,408, **D**).

**eFigure 5: GWAS Manhattan plots for the immune BAG**

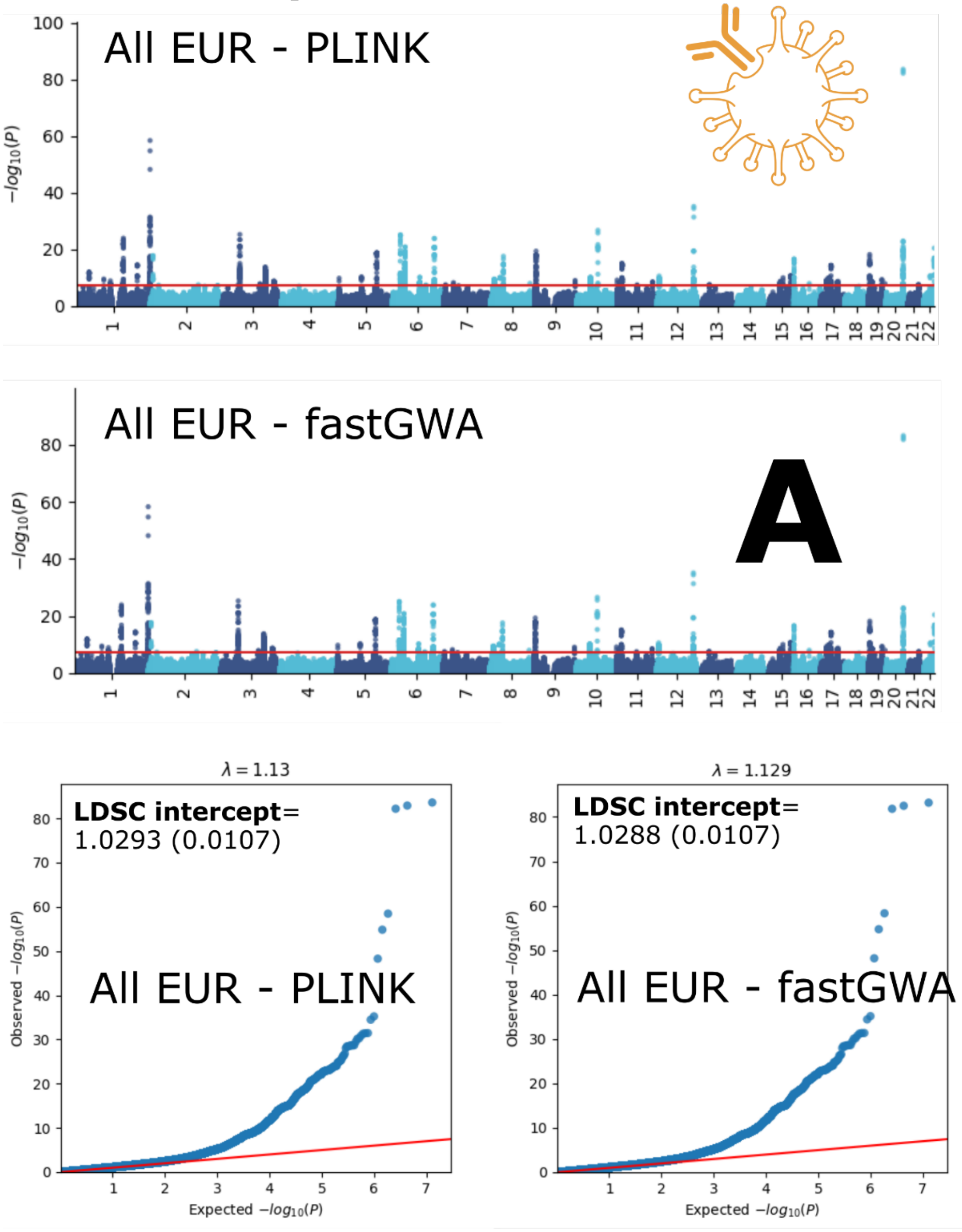

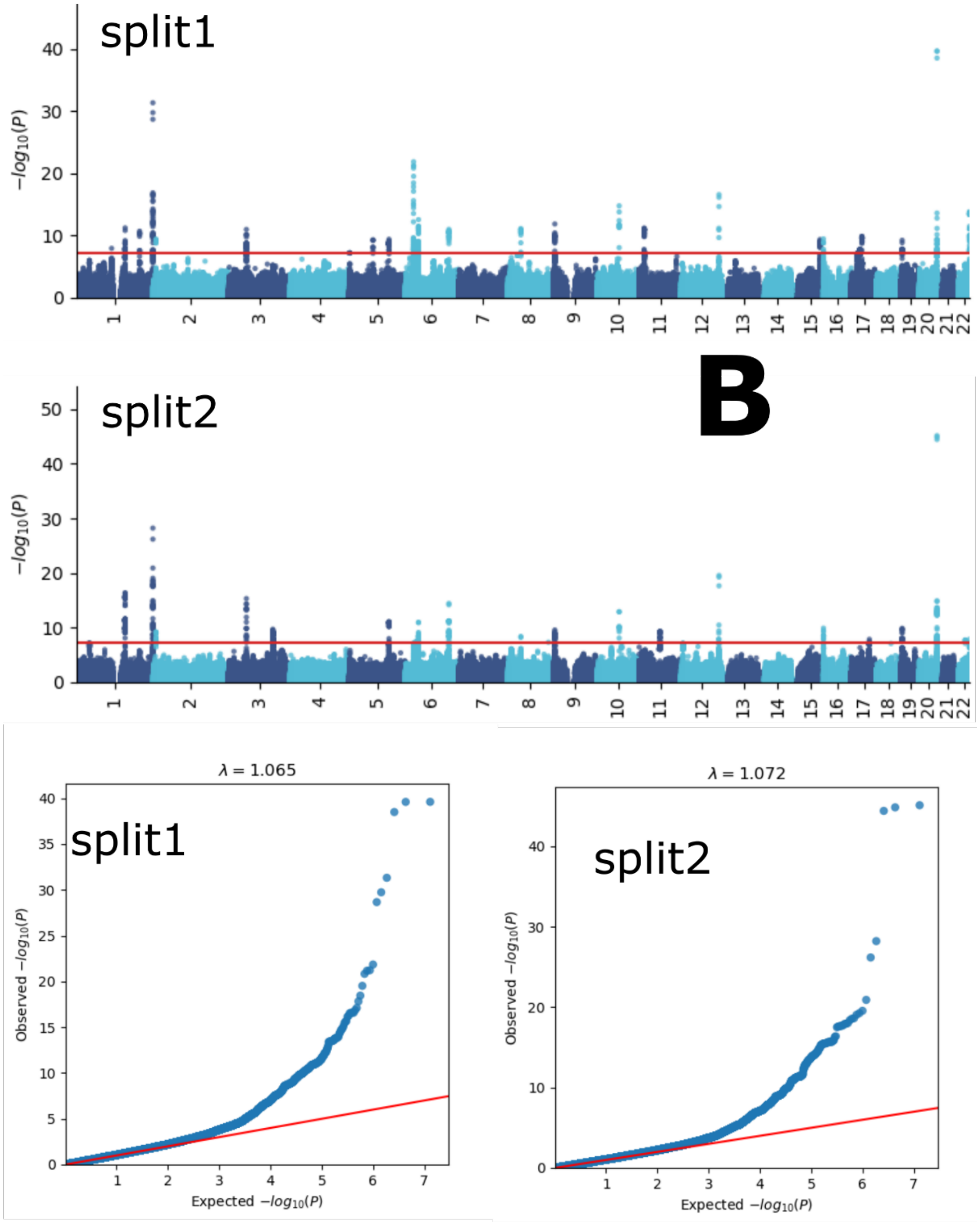
split1
$-log_{10}(P)$
split2
B
$-log_{10}(P)$
$\lambda = 1.065$
split1
Observed $-log_{10}(P)$
Expected $-log_{10}(P)$
$\lambda = 1.072$
split2
Observed $-log_{10}(P)$
Expected $-log_{10}(P)$

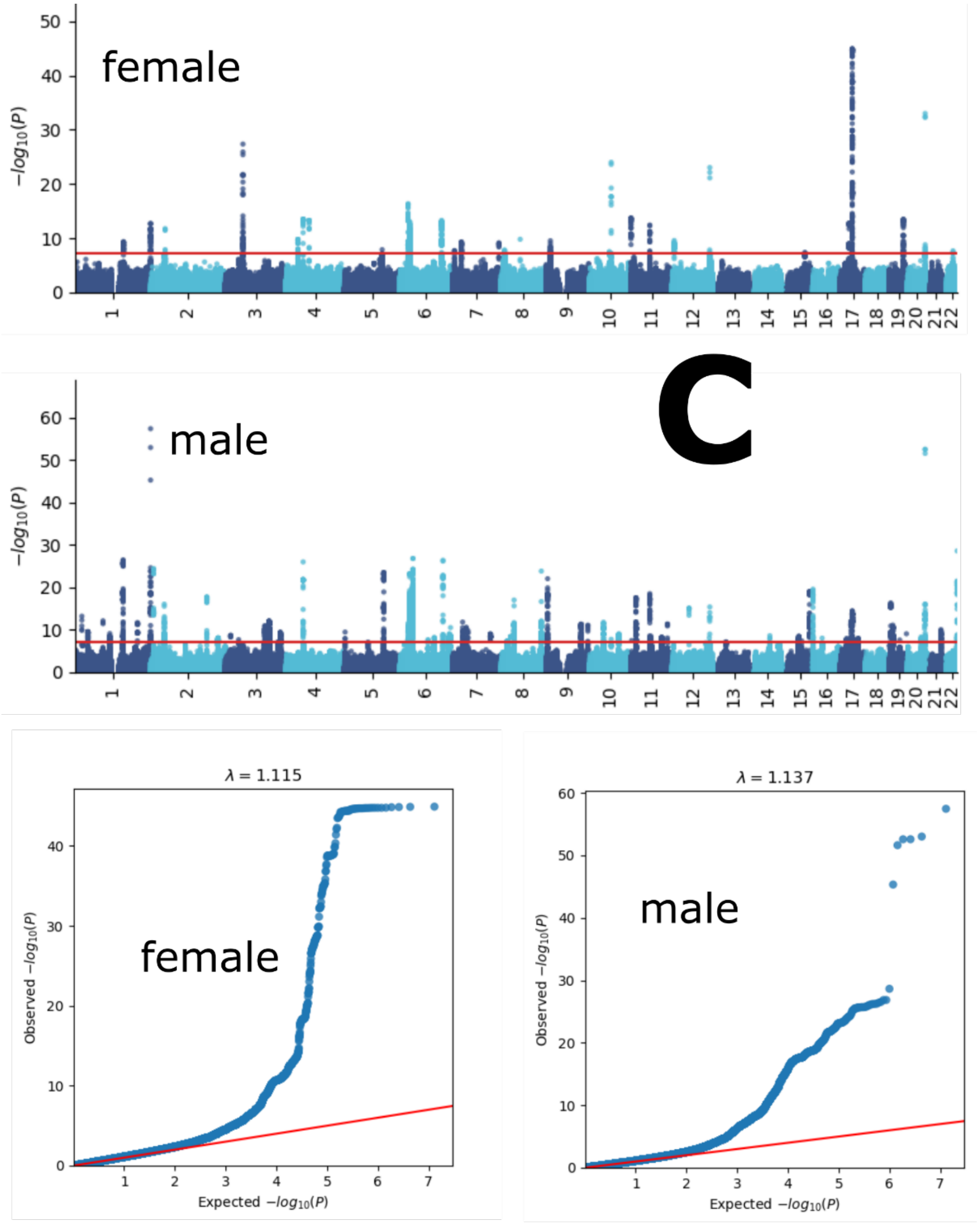
female
male
C
$-log_{10}(P)$
1 2 3 4 5 6 7 8 9 10 11 12 13 14 15 16 17 18 19 20 21 22
λ = 1.115
λ = 1.137
Observed $-log_{10}(P)$
Expected $-log_{10}(P)$

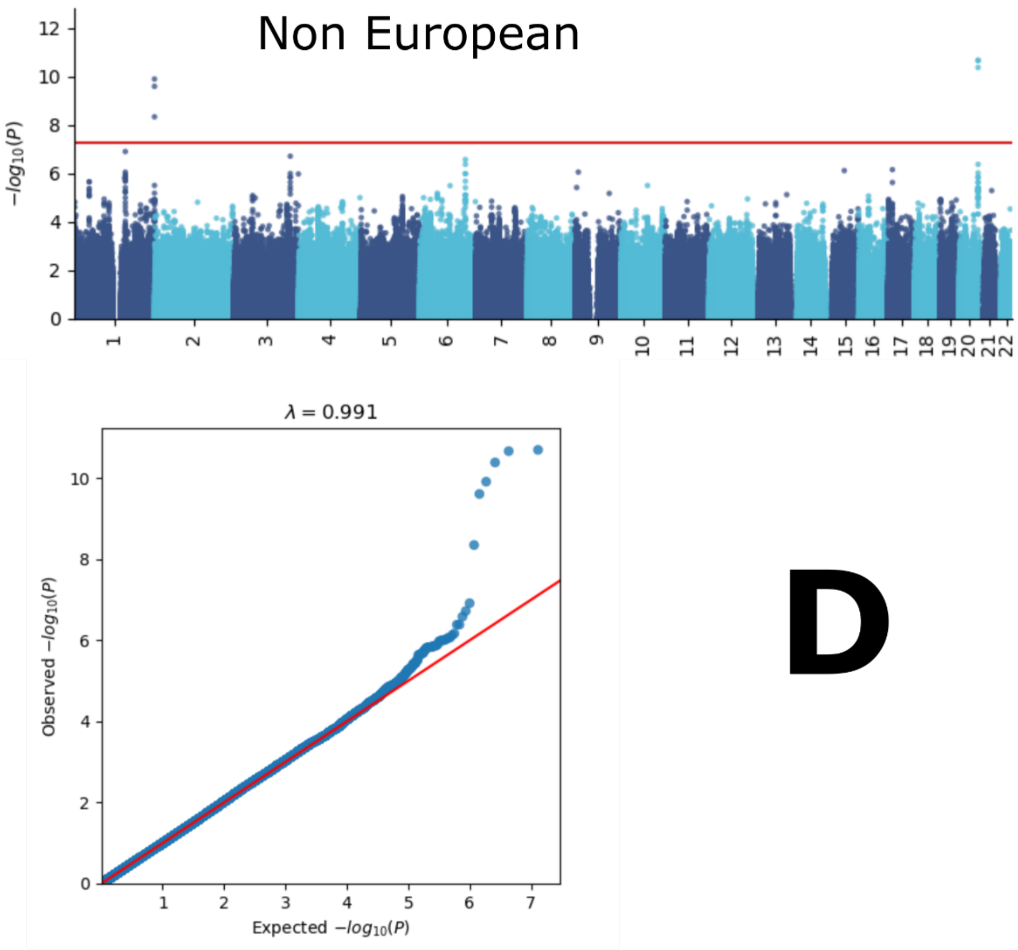

Manhattan and QQ plots, along with genomic inflation factors and LDSC intercepts, are displayed for the primary GWAS conducted on individuals of European ancestry (*N*=111,386) using PLINK and fastGWA (**A**). Additionally, results are presented for split-sample GWAS (split1 and split2, **B**), sex-stratified GWAS (female and male, **C**), and GWAS involving non-European ancestry populations (*N*=20,408, **D**).

**eFigure 6: GWAS Manhattan plots for the metabolic BAG**

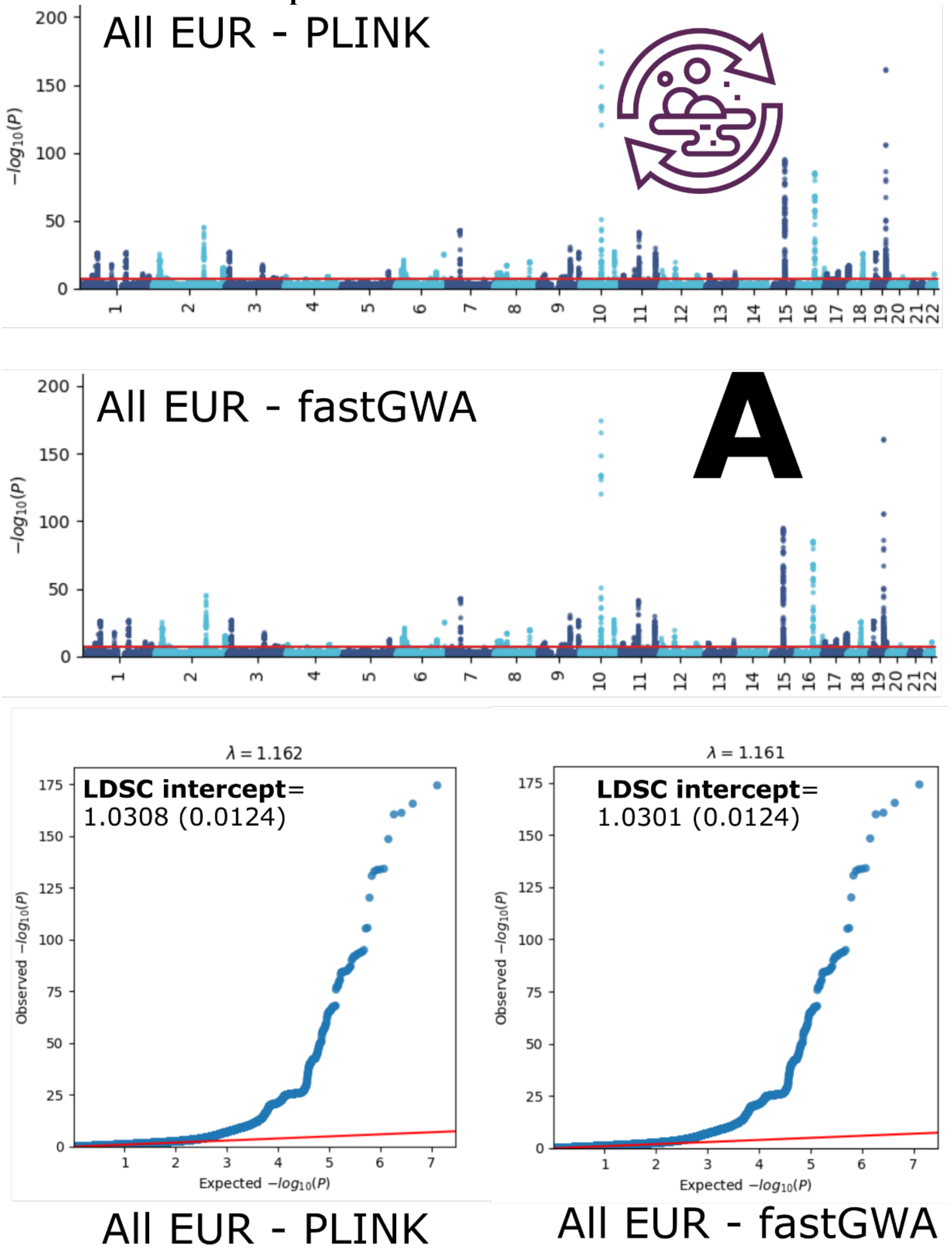

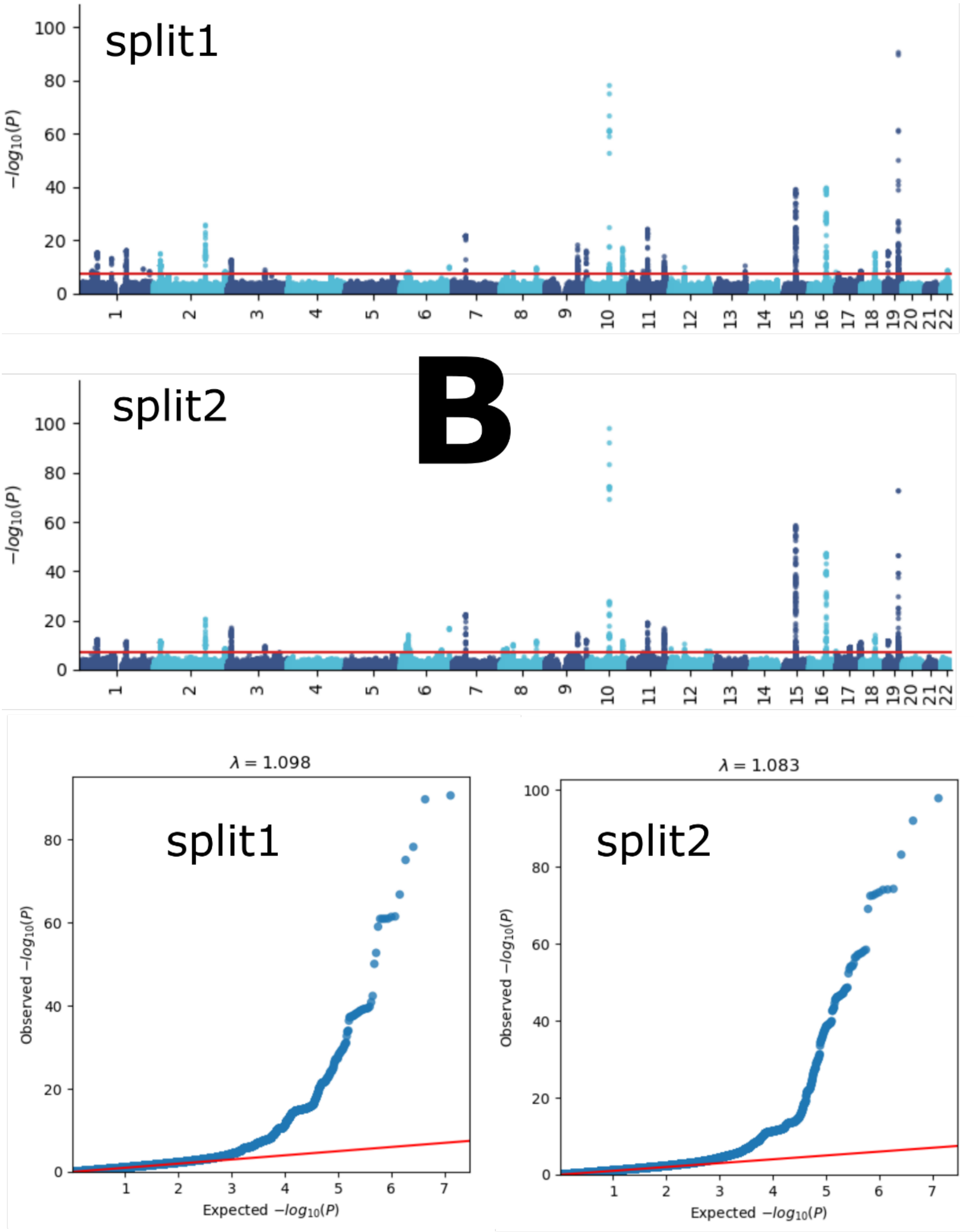
split1
$-log_{10}(P)$
split2
B
$\lambda = 1.098$
$\lambda = 1.083$
Observed $-log_{10}(P)$
Expected $-log_{10}(P)$

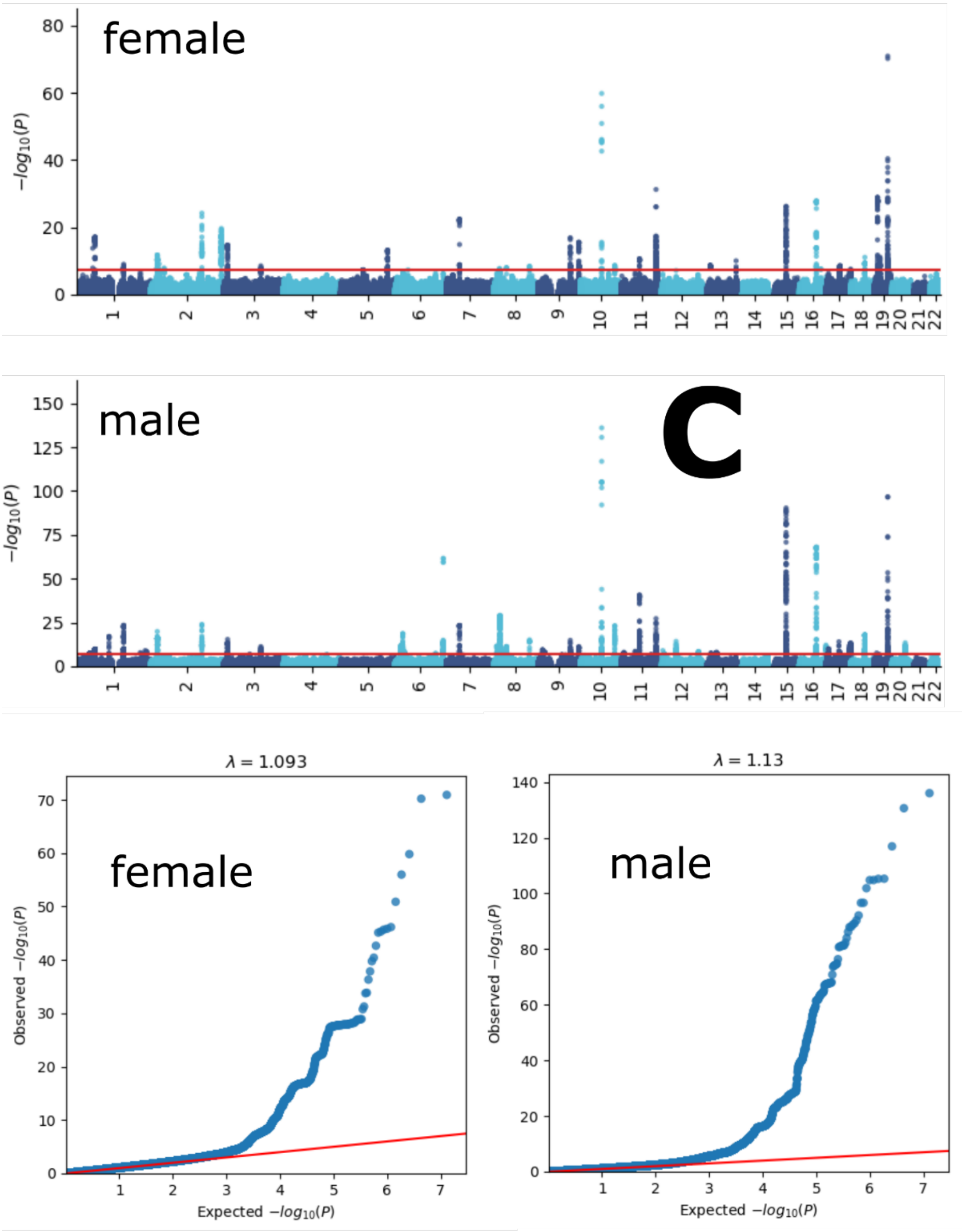
female
male
C
λ = 1.093
female
λ = 1.13
male
Observed −log10(P)
Expected −log10(P)

337

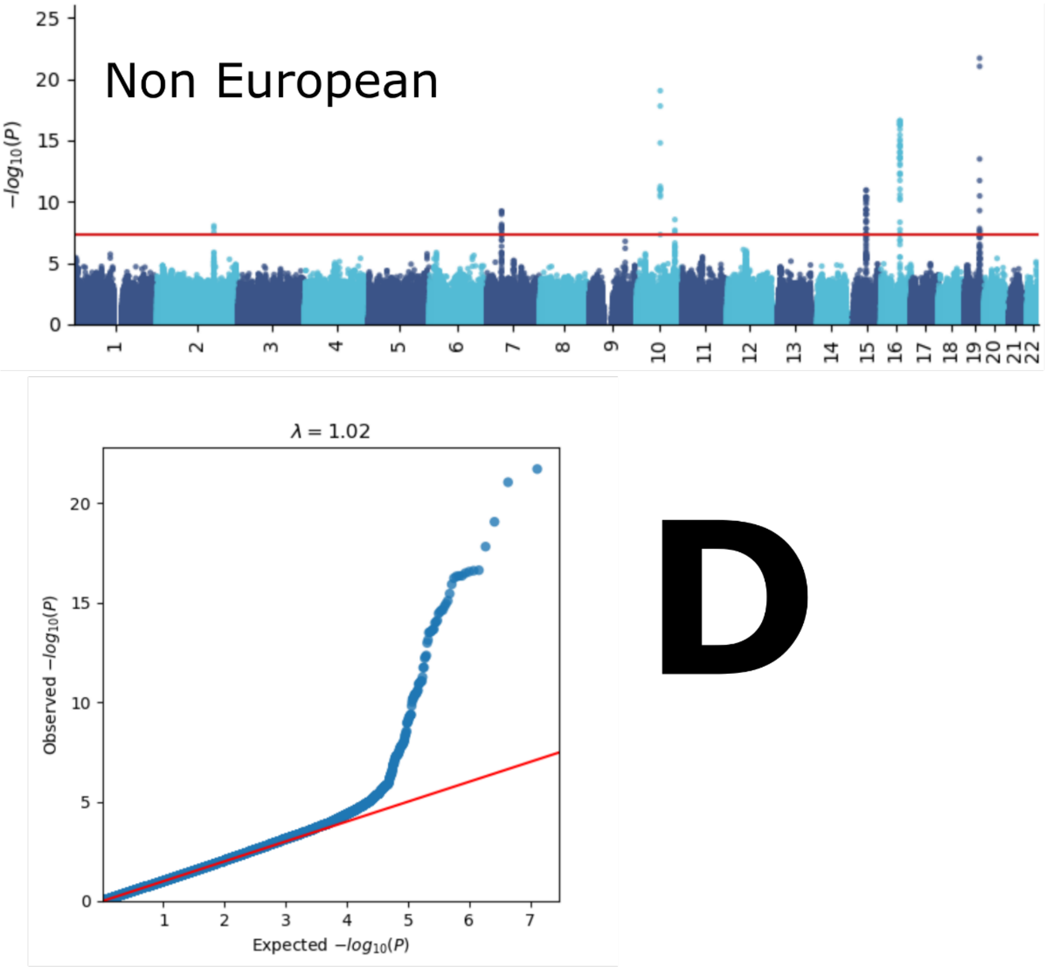

Manhattan and QQ plots, along with genomic inflation factors and LDSC intercepts, are displayed for the primary GWAS conducted on individuals of European ancestry (*N*=111,386) using PLINK and fastGWA (**A**). Additionally, results are presented for split-sample GWAS (split1 and split2, **B**), sex-stratified GWAS (female and male, **C**), and GWAS involving non-European ancestry populations (*N*=20,408, **D**).

**eFigure 7: GWAS Manhattan plots for the musculoskeletal BAG**

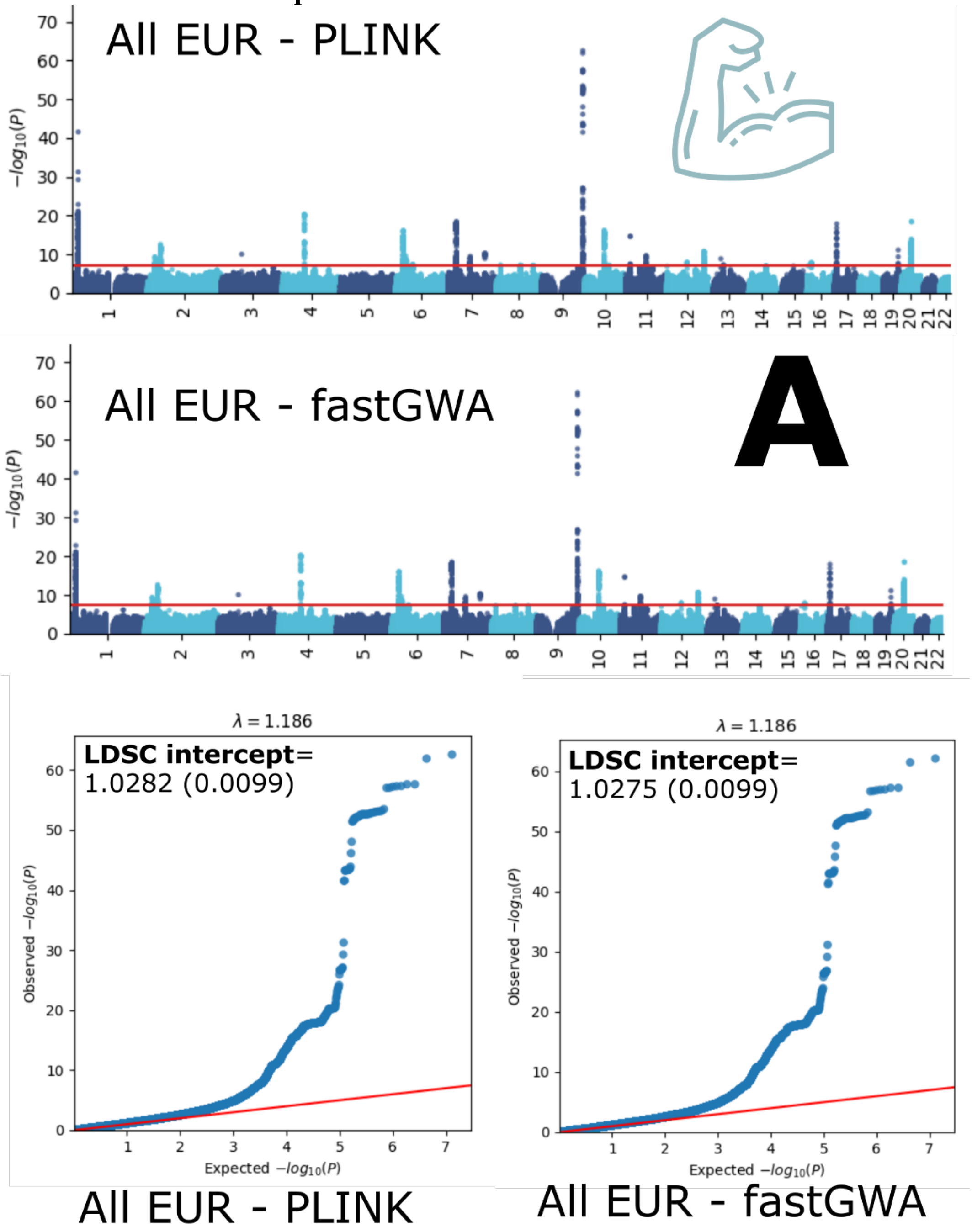

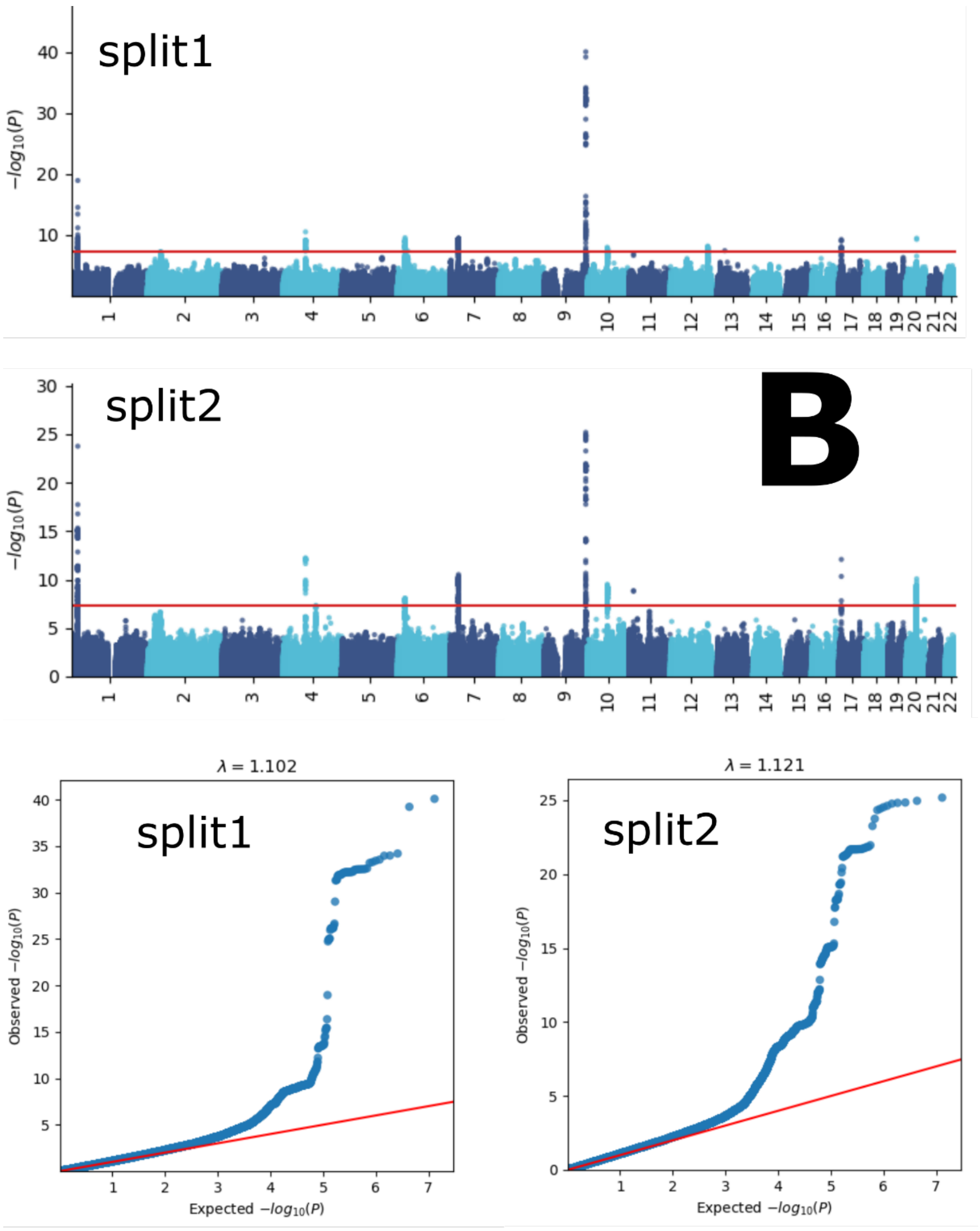
split1
split2
B
$-log_{10}(P)$
$\lambda = 1.102$
$\lambda = 1.121$
Observed $-log_{10}(P)$
Expected $-log_{10}(P)$

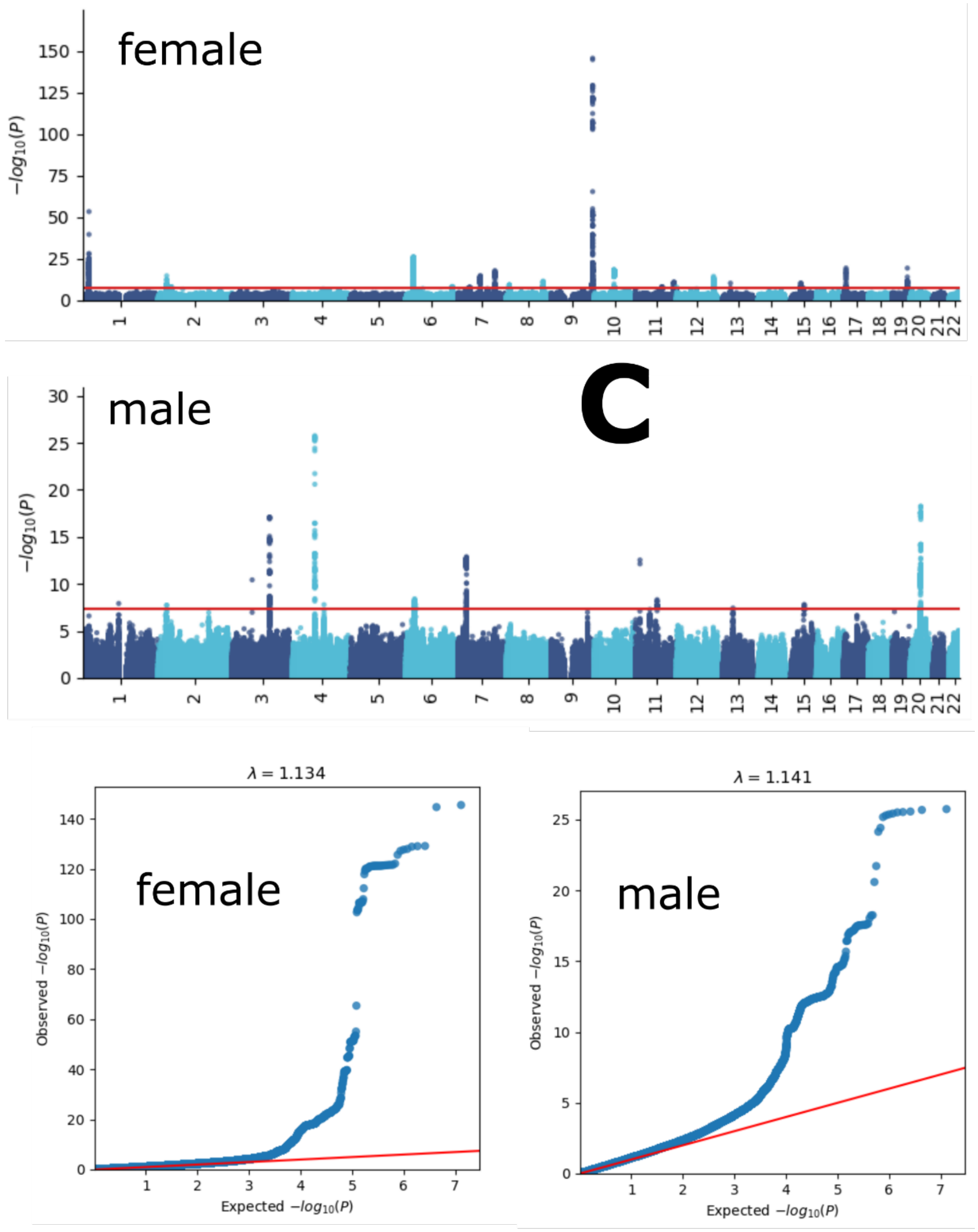
female
−log10(P)
male
C
−log10(P)
λ = 1.134
female
Observed −log10(P)
Expected −log10(P)
λ = 1.141
male
Observed −log10(P)
Expected −log10(P)

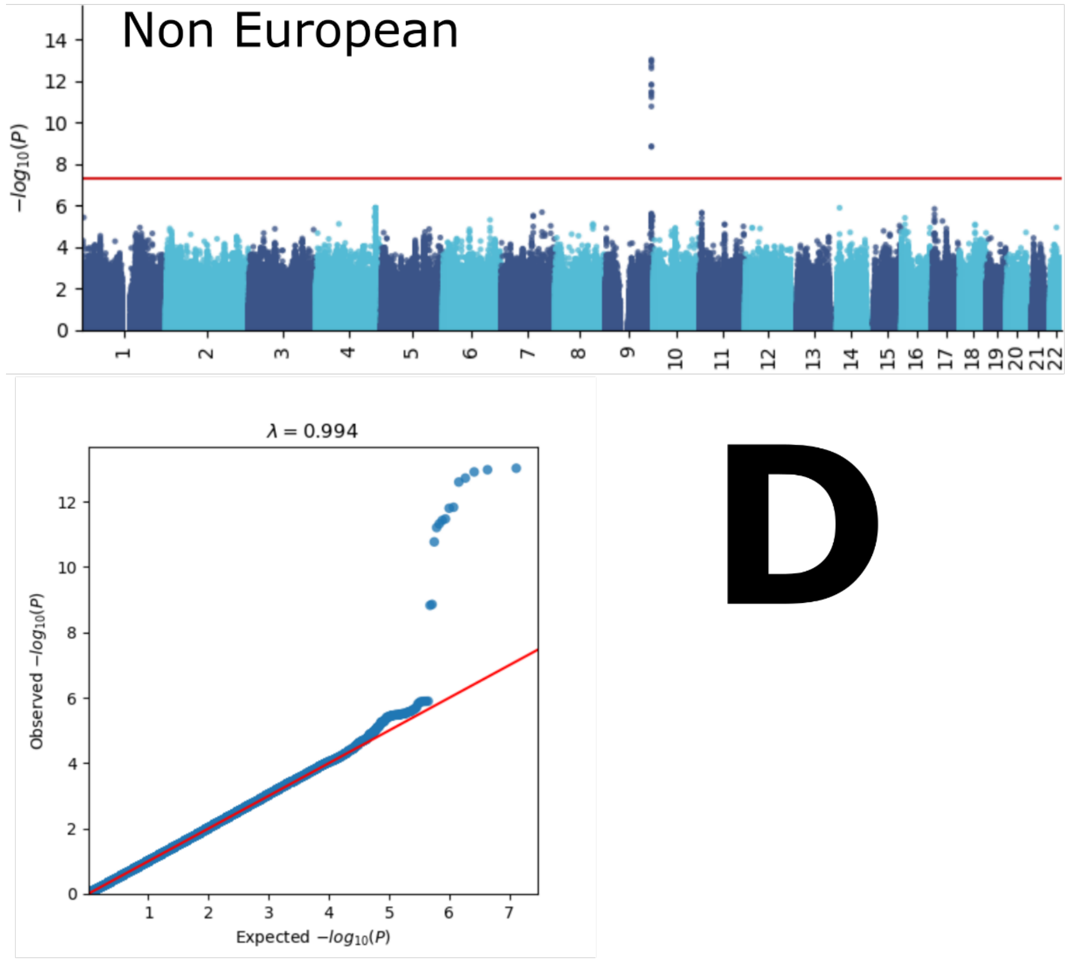

Manhattan and QQ plots, along with genomic inflation factors and LDSC intercepts, are displayed for the primary GWAS conducted on individuals of European ancestry (*N*=111,386) using PLINK and fastGWA (**A**). Additionally, results are presented for split-sample GWAS (split1 and split2, **B**), sex-stratified GWAS (female and male, **C**), and GWAS involving non-European ancestry populations (*N*=20,408, **D**).

**eFigure 8: GWAS Manhattan plots for the pulmonary BAG**

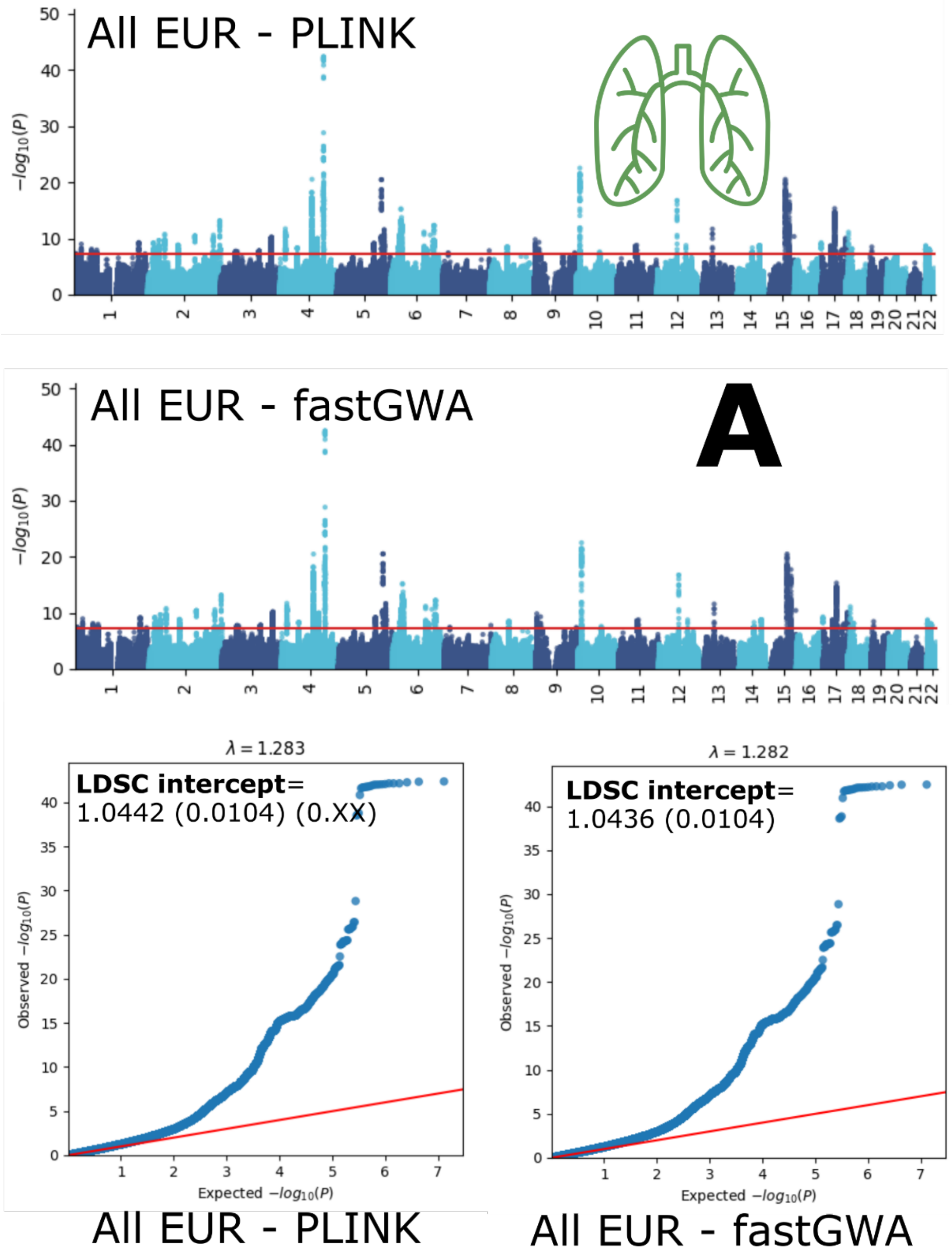

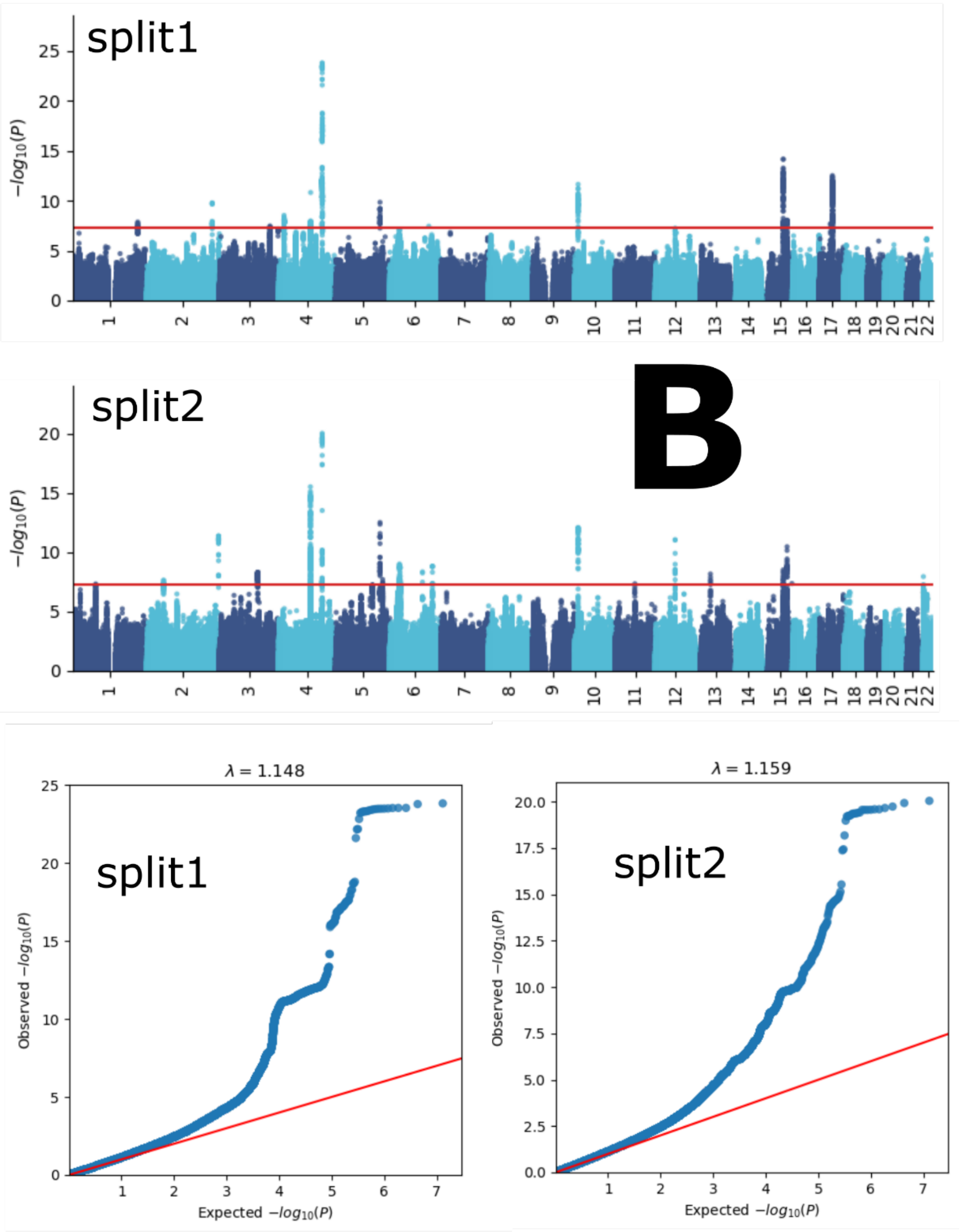
split1
$-log_{10}(P)$
split2
B
$-log_{10}(P)$
$\lambda = 1.148$
split1
Observed $-log_{10}(P)$
Expected $-log_{10}(P)$
$\lambda = 1.159$
split2
Observed $-log_{10}(P)$
Expected $-log_{10}(P)$

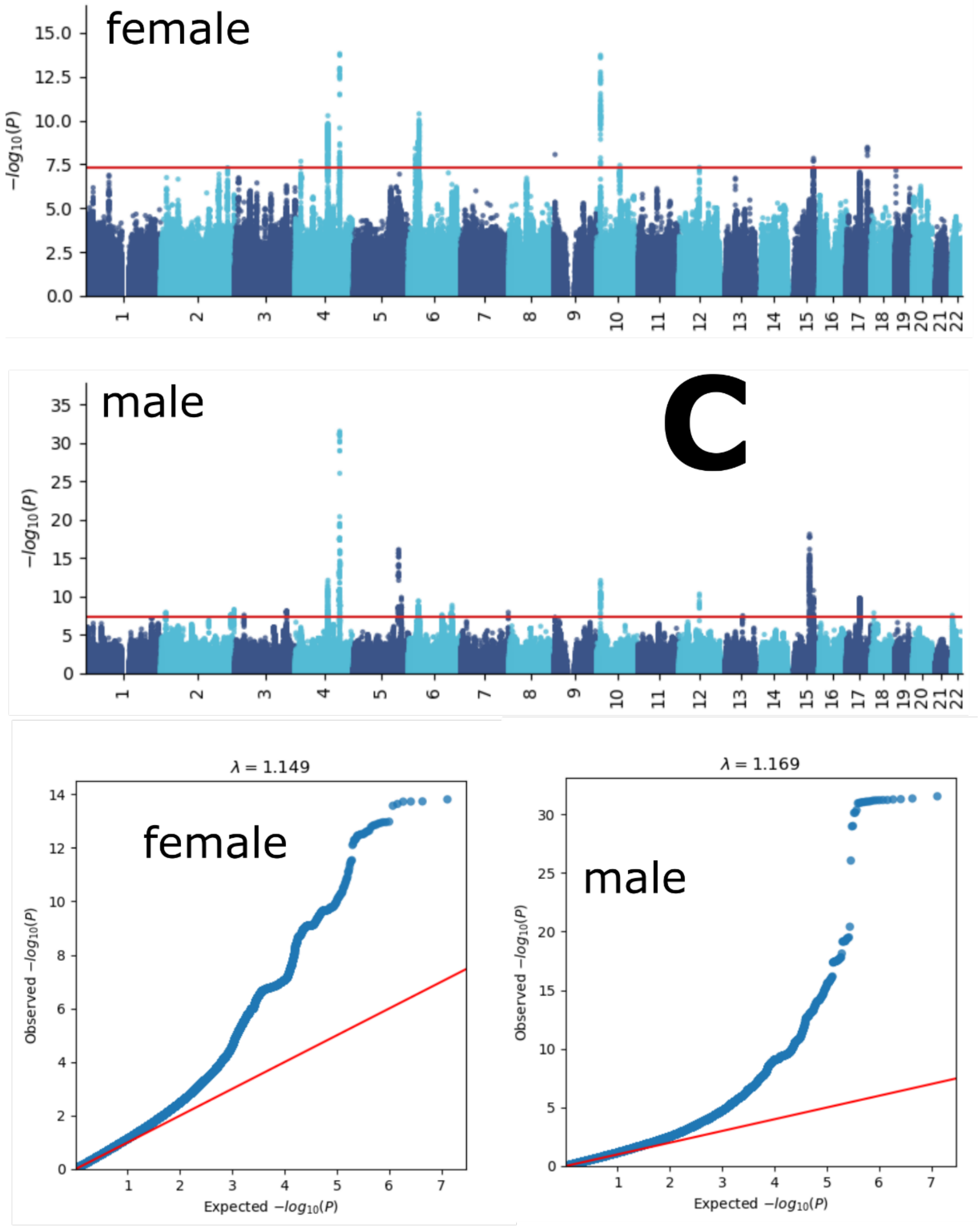
female
15.0
12.5
10.0
7.5
5.0
2.5
0.0
$-log_{10}(P)$
1 2 3 4 5 6 7 8 9 10 11 12 13 14 15 16 17 18 19 20 21 22
male
C
35
30
25
20
15
10
5
0
$-log_{10}(P)$
1 2 3 4 5 6 7 8 9 10 11 12 13 14 15 16 17 18 19 20 21 22
$\lambda = 1.149$
female
Observed $-log_{10}(P)$
Expected $-log_{10}(P)$
$\lambda = 1.169$
male
Observed $-log_{10}(P)$
Expected $-log_{10}(P)$

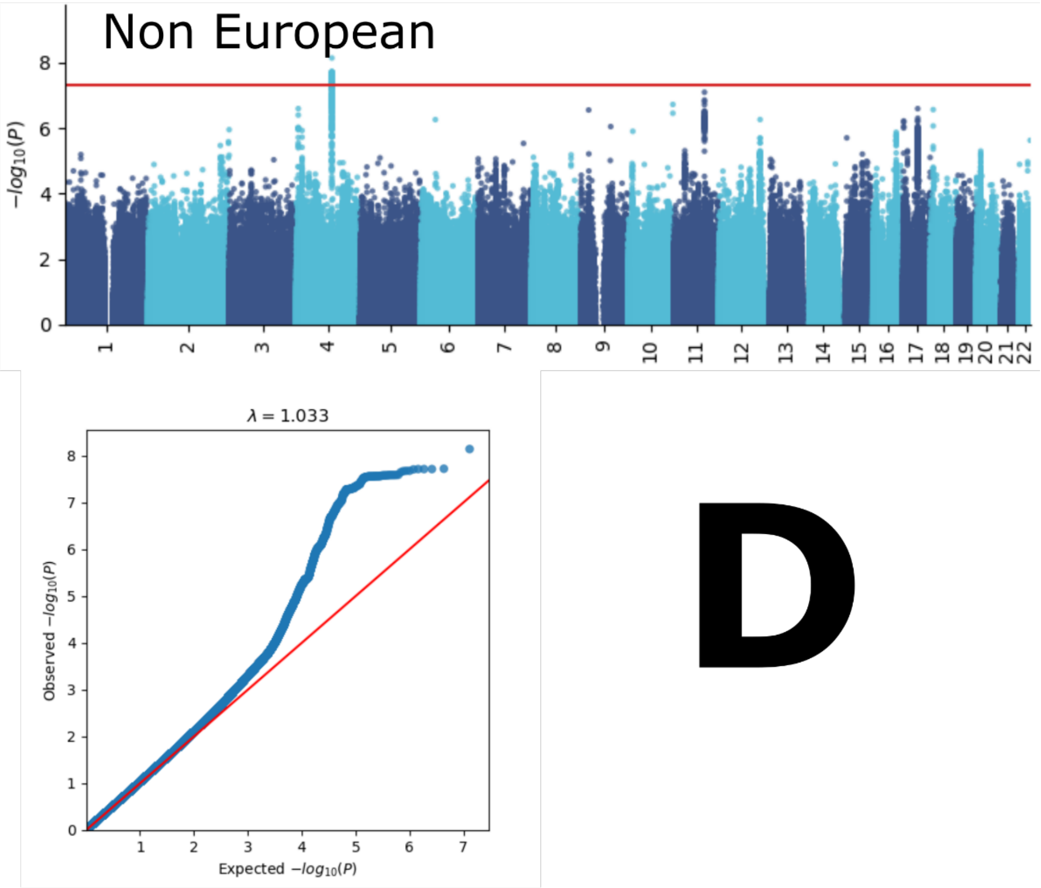

Manhattan and QQ plots, along with genomic inflation factors and LDSC intercepts, are displayed for the primary GWAS conducted on individuals of European ancestry (*N*=111,386) using PLINK and fastGWA (**A**). Additionally, results are presented for split-sample GWAS (split1 and split2, **B**), sex-stratified GWAS (female and male, **C**), and GWAS involving non-European ancestry populations (*N*=20,408, **D**).

**eFigure 9: GWAS Manhattan plots for the renal BAG**

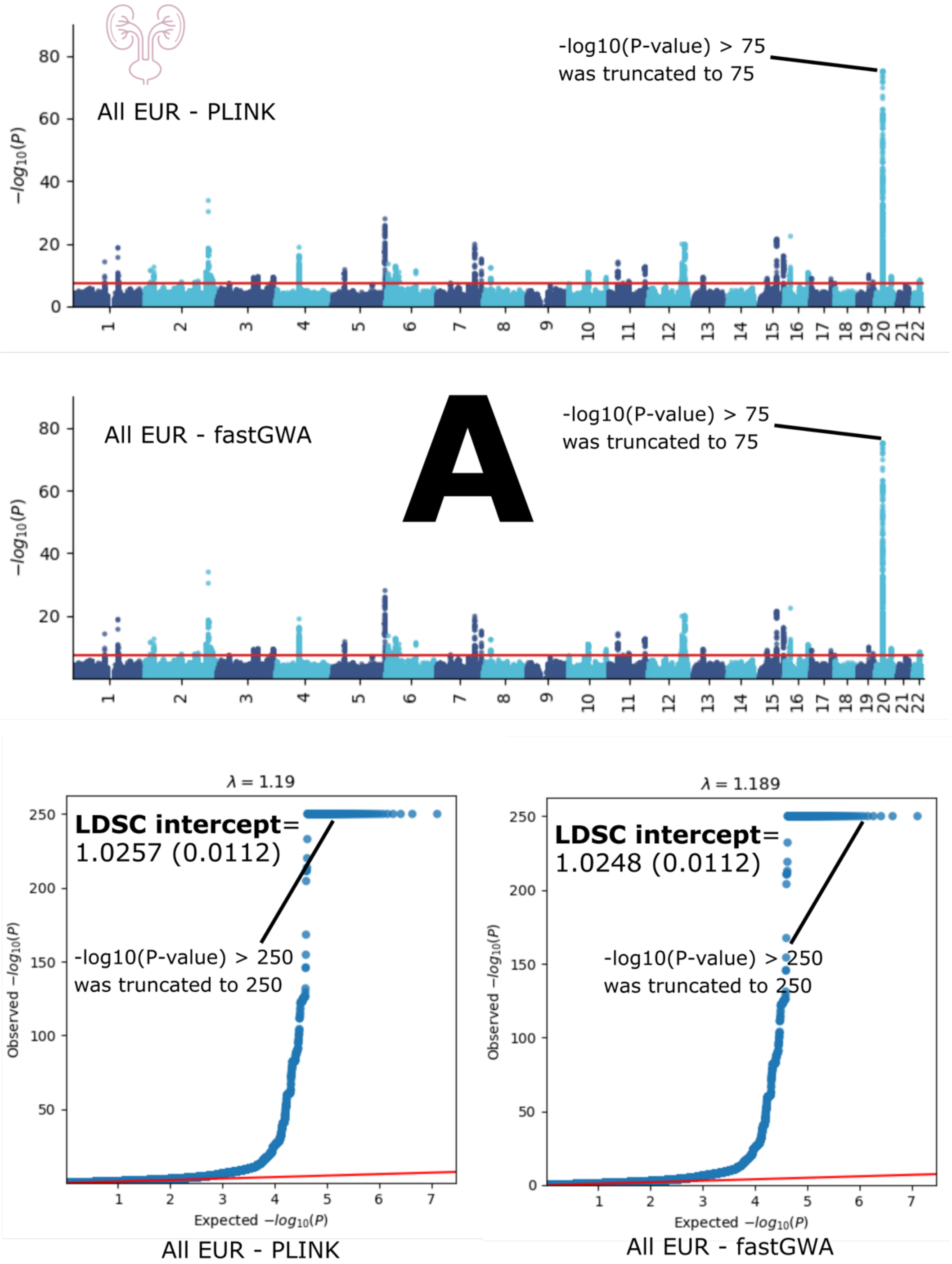

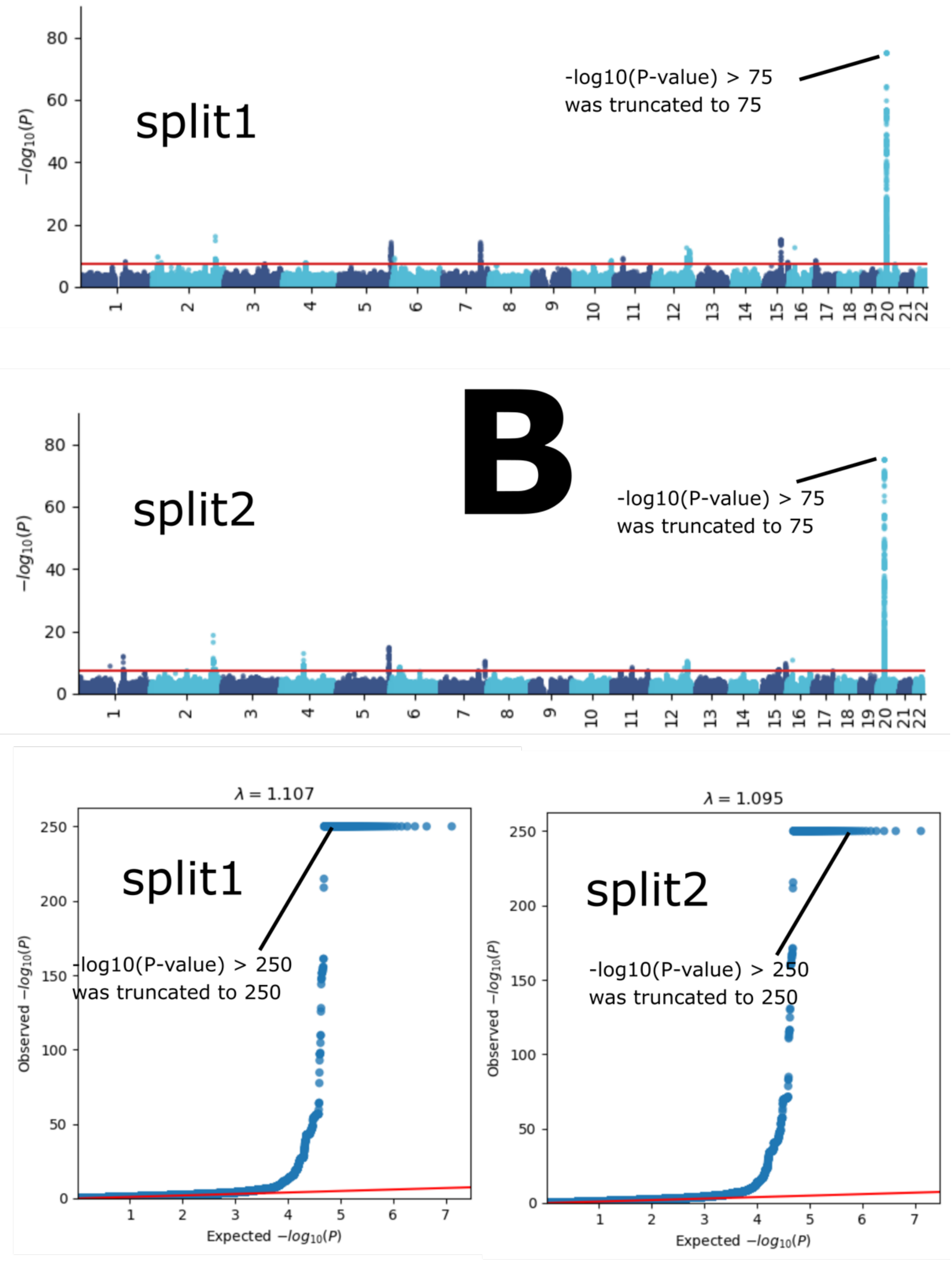
B
split1
-log10(P-value) > 75
was truncated to 75
split2
-log10(P-value) > 75
was truncated to 75
$-log_{10}(P)$
λ = 1.107
split1
-log10(P-value) > 250
was truncated to 250
λ = 1.095
split2
-log10(P-value) > 250
was truncated to 250
Observed $-log_{10}(P)$
Expected $-log_{10}(P)$

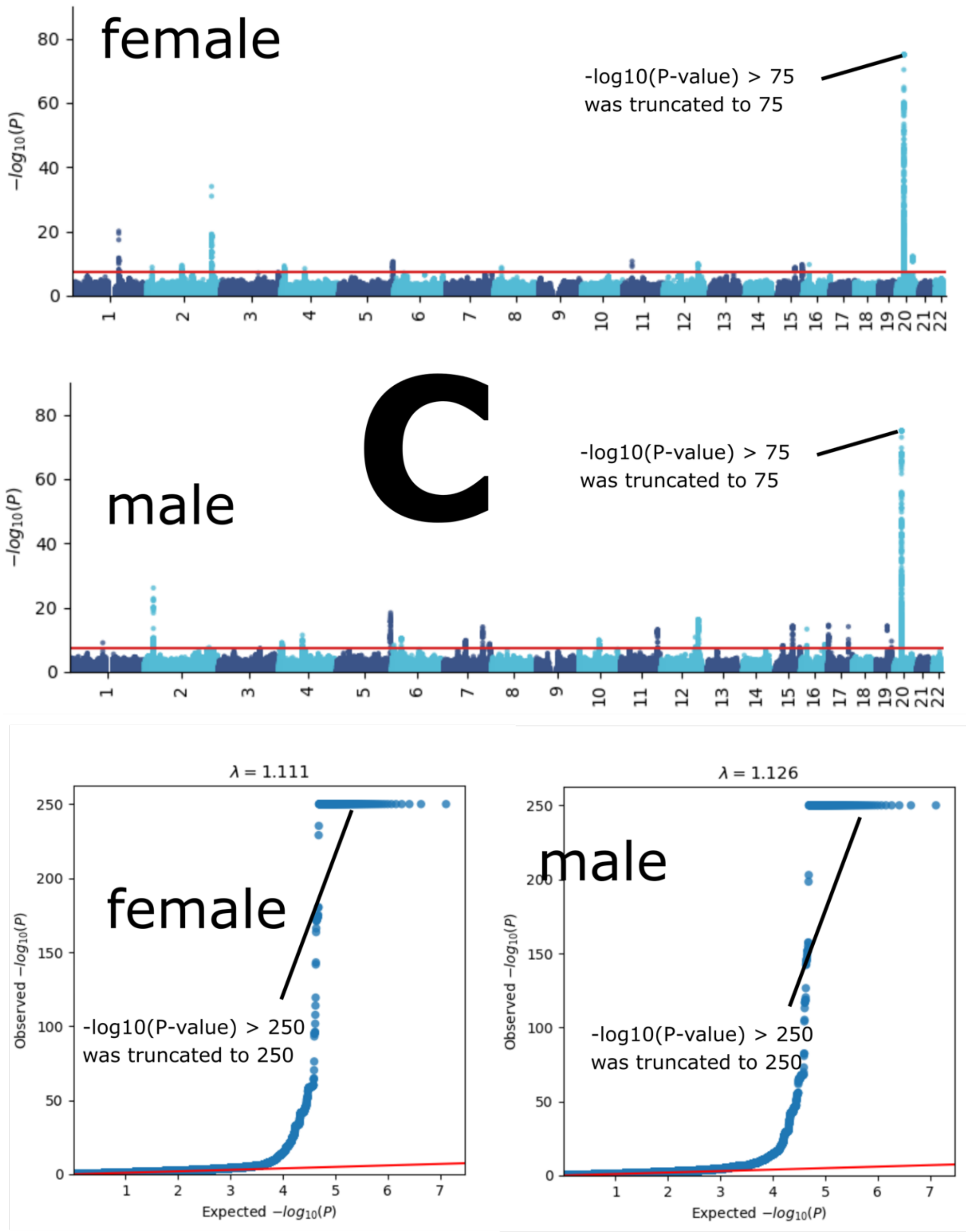
female
-log10(P-value) > 75
was truncated to 75
C
male
-log10(P-value) > 75
was truncated to 75
$-log_{10}(P)$
$\lambda = 1.111$
female
-log10(P-value) > 250
was truncated to 250
$\lambda = 1.126$
male
-log10(P-value) > 250
was truncated to 250
Observed $-log_{10}(P)$
Expected $-log_{10}(P)$

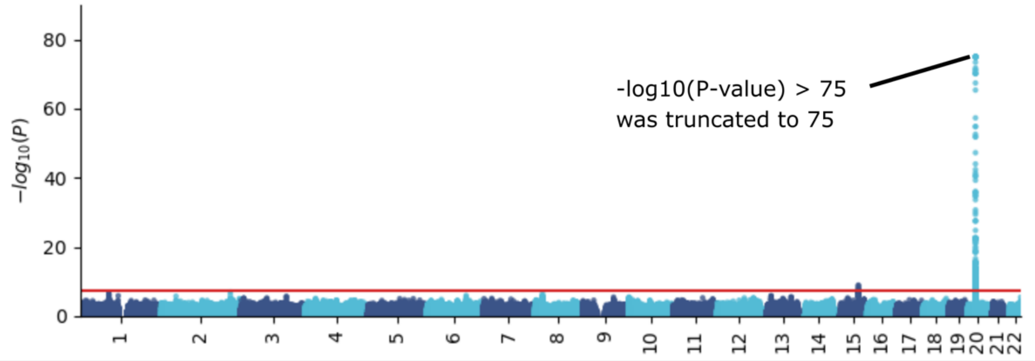

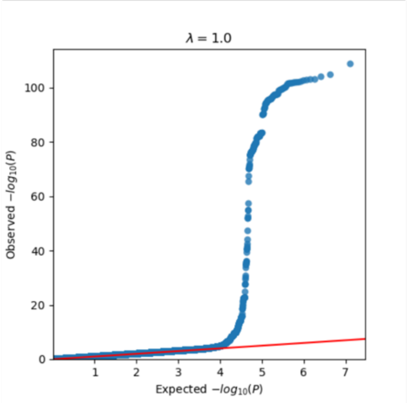

**D**

Manhattan and QQ plots, along with genomic inflation factors and LDSC intercepts, are displayed for the primary GWAS conducted on individuals of European ancestry (*N*=111,386) using PLINK and fastGWA (**A**). Additionally, results are presented for split-sample GWAS (split1 and split2, **B**), sex-stratified GWAS (female and male, **C**), and GWAS involving non-European ancestry populations (*N*=20,408, **D**). For visualization purposes, we chose to truncate the highly significant P-value (P-value<$1x10^{-300}$) to a lower P-value ($1x10^{-75}$ for Manhattan plots and $1x10^{-250}$ for QQ plots).

**eFigure 10: Bayesian colocalization analysis for the locus on chromosome 6 between the hepatic and musculoskeletal BAGs**

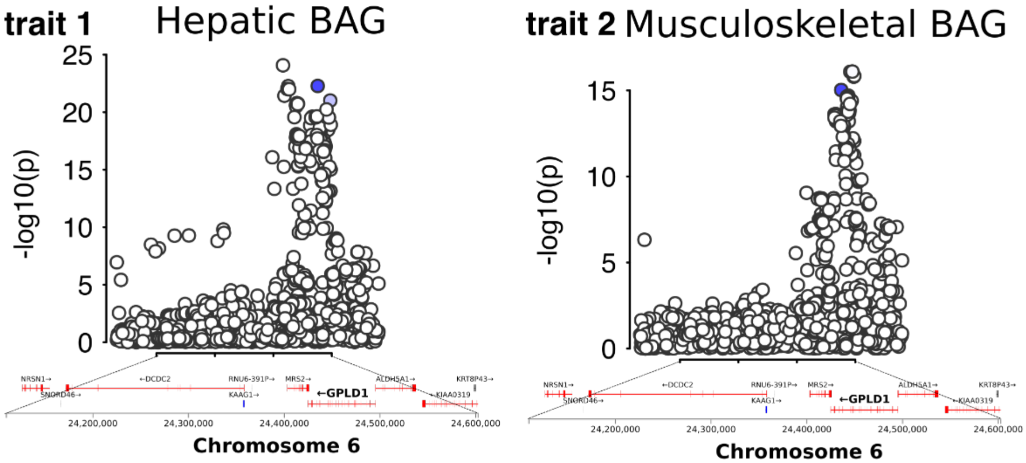

We conducted a Bayesian colocalization analysis using Bayes factors to investigate shared causal variants in a specific locus on chromosome 6 for the hepatic and musculoskeletal BAGs. The analysis tested five hypotheses, denoted by their posterior probabilities: H0 (no association with either trait), H1 (association with trait 1 but not trait 2), H2 (association with trait 2 but not trait 1), H3 (association with both traits but with separate causal variants), and H4 (association with both traits with a shared causal variant). The potential causal variants for both traits are indicated by blue-colored SNPs, assuming each locus contains at most one causal variant. The gene mapped to this locus (*GPLD1*) is shown in bold based on physical positions.

**eFigure 11: Exemplary genomic locus for each BAG in the nine human organ systems**

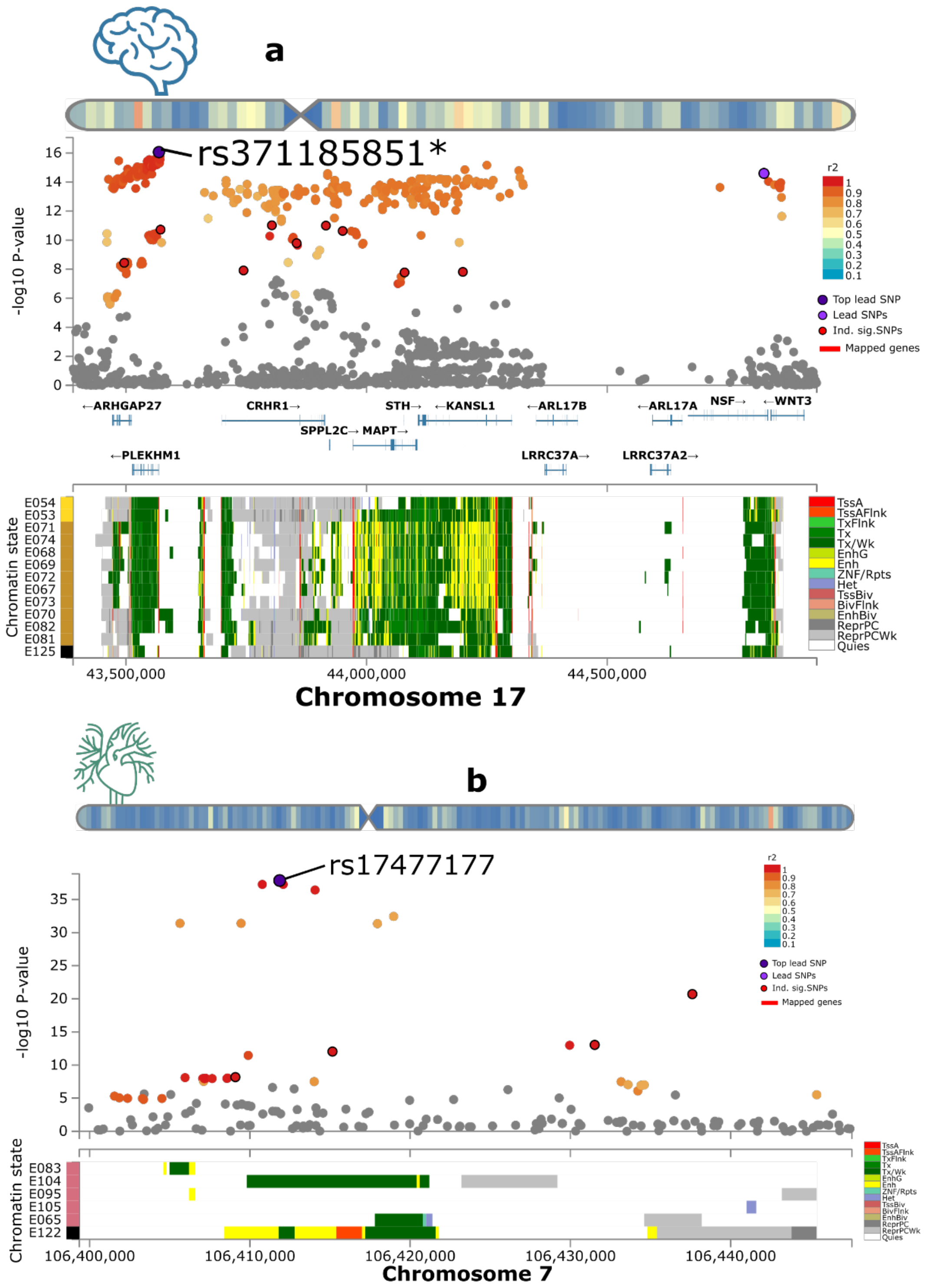

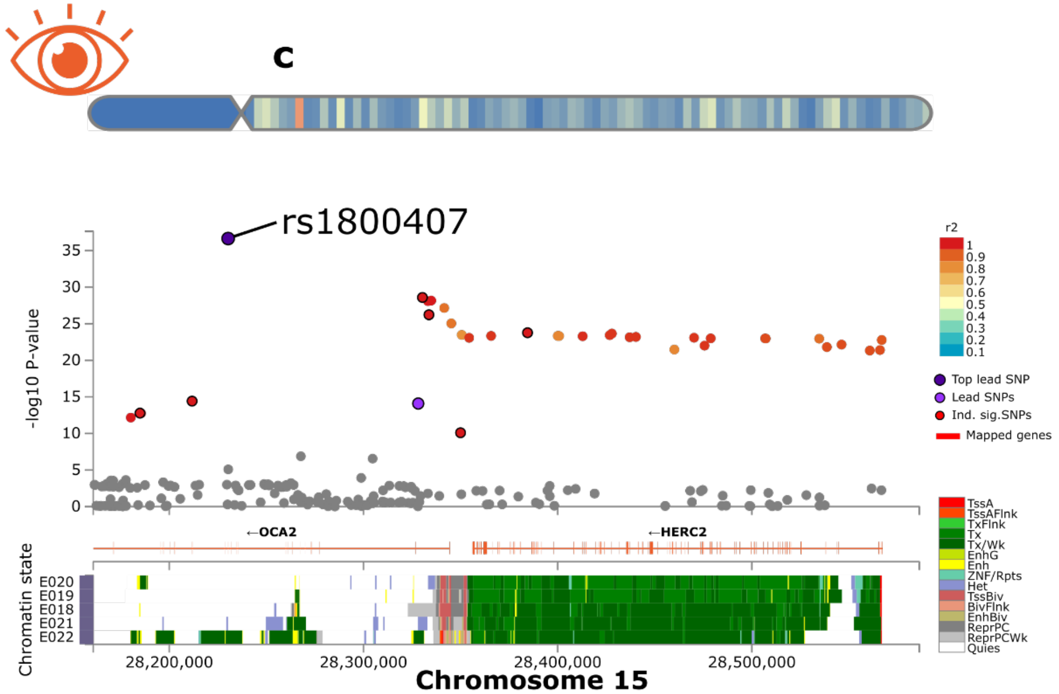
c
rs1800407
-log10 P-value
r2
Top lead SNP
Lead SNPs
Ind. sig.SNPs
Mapped genes
←OCA2
←HERC2
Chromatin state
E020
E019
E018
E021
E022
TssA
TssAFlnk
TxFlnk
Tx
Tx/Wk
EnhG
Enh
ZNF/Rpts
Het
TssBiv
BivFlnk
EnhBiv
ReprPC
ReprPCWk
Quies
28,200,000
28,300,000
28,400,000
28,500,000
Chromosome 15

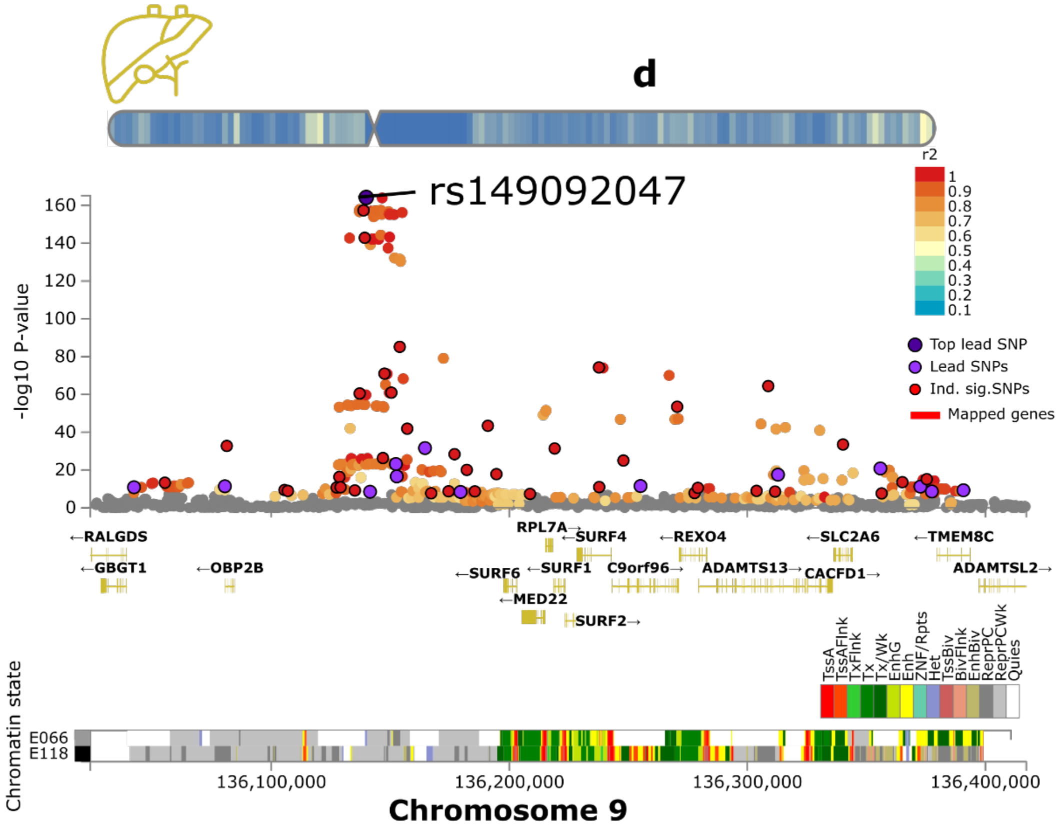
d
rs149092047
-log10 P-value
r2
Top lead SNP
Lead SNPs
Ind. sig.SNPs
Mapped genes
←RALGDS
←GBGT1
←OBP2B
RPL7A→
←SURF4
←REXO4
←SLC2A6
←TMEM8C
←SURF6
←SURF1
C9orf96→
ADAMTS13→
CACFD1→
ADAMTSL2→
←MED22
SURF2→
Chromatin state
E066
E118
TssA
TssAFlnk
TxFlnk
Tx
Tx/Wk
EnhG
Enh
ZNF/Rpts
Het
TssBiv
BivFlnk
EnhBiv
ReprPC
ReprPCWk
Quies
136,100,000
136,200,000
136,300,000
136,400,000
Chromosome 9

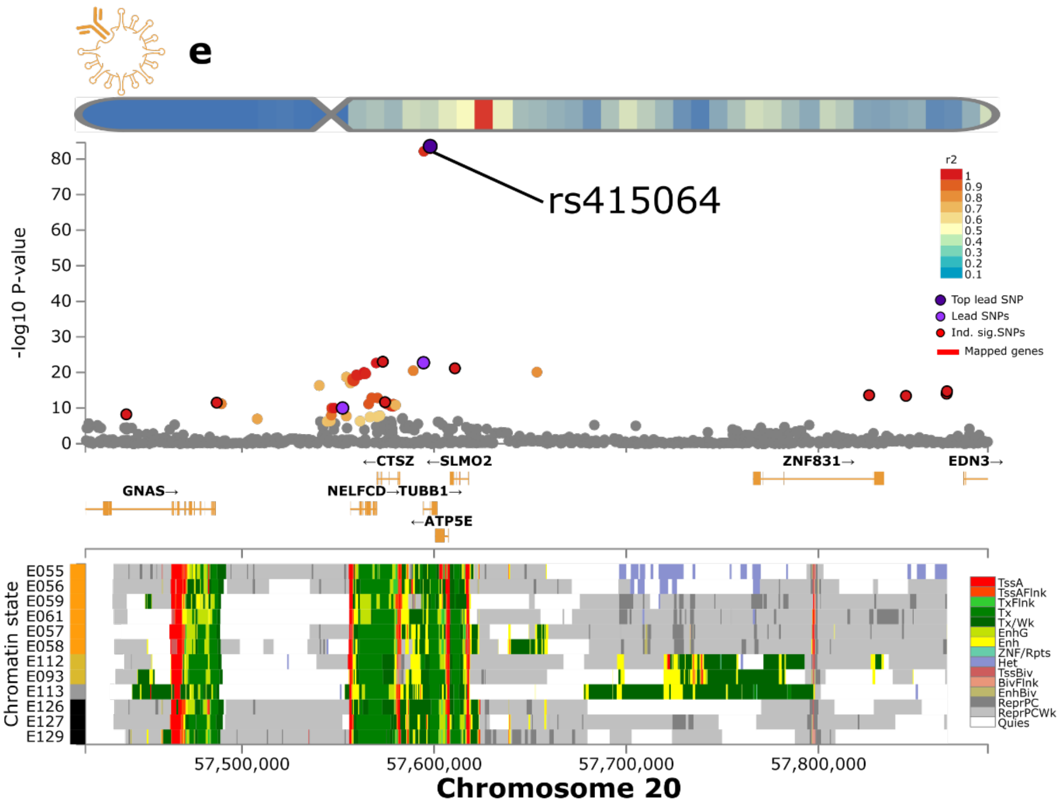
e
rs415064
-log10 P-value
r2
Top lead SNP
Lead SNPs
Ind. sig.SNPs
Mapped genes
←CTSZ
←SLMO2
ZNF831→
EDN3→
GNAS→
NELFCD→TUBB1→
←ATP5E
Chromatin state
E055
E056
E059
E061
E057
E058
E112
E093
E113
E126
E127
E129
TssA
TssAFlnk
TxFlnk
Tx
Tx/Wk
EnhG
Enh
ZNF/Rpts
Het
TssBiv
BivFlnk
EnhBiv
ReprPC
ReprPCWk
Quies
57,500,000
57,600,000
57,700,000
57,800,000
Chromosome 20

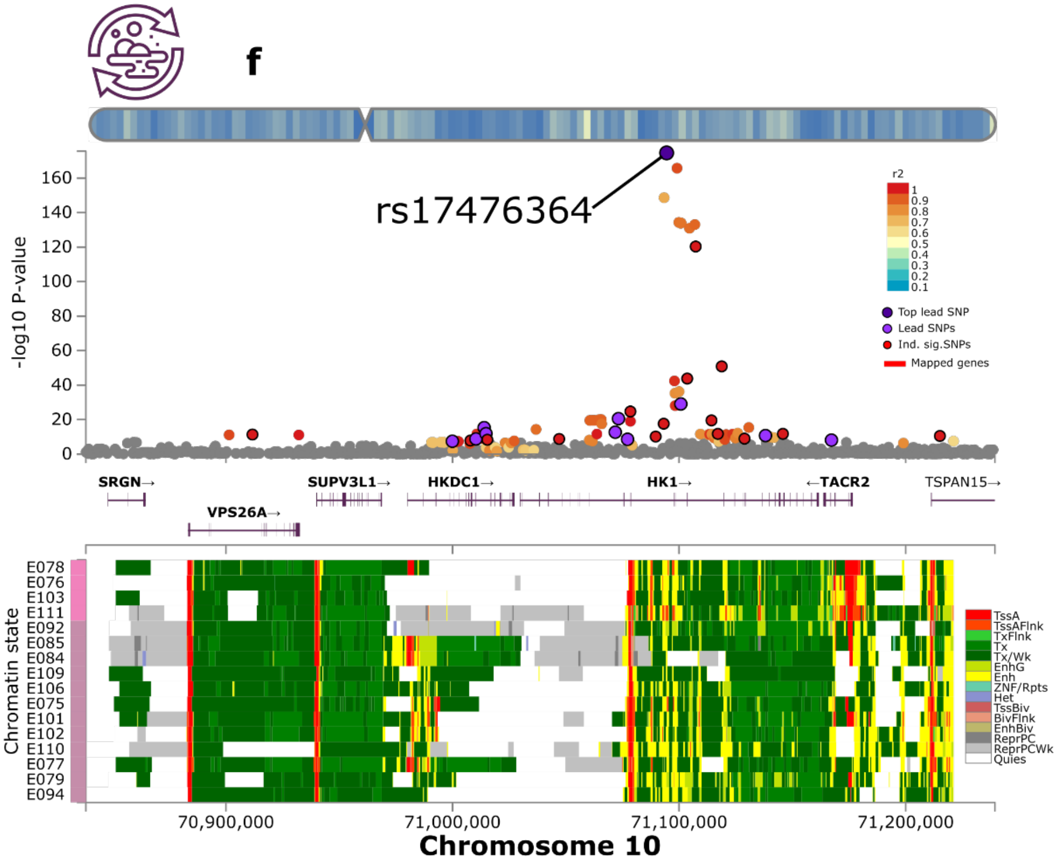
f
rs17476364
-log10 P-value
r2
Top lead SNP
Lead SNPs
Ind. sig.SNPs
Mapped genes
SRGN→
SUPV3L1→
HKDC1→
HK1→
←TACR2
TSPAN15→
VPS26A→
Chromatin state
E078
E076
E103
E111
E092
E085
E084
E109
E106
E075
E101
E102
E110
E077
E079
E094
TssA
TssAFlnk
TxFlnk
Tx
Tx/Wk
EnhG
Enh
ZNF/Rpts
Het
TssBiv
BivFlnk
EnhBiv
ReprPC
ReprPCWk
Quies
70,900,000
71,000,000
71,100,000
71,200,000
Chromosome 10

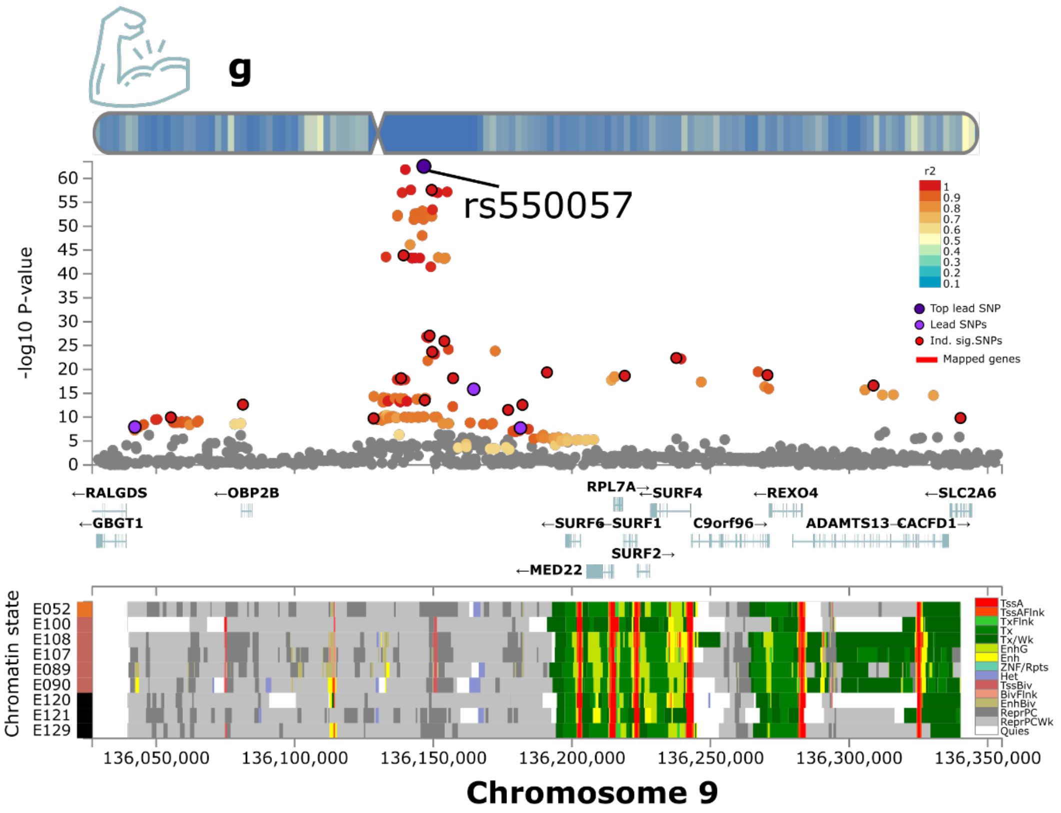
g
rs550057
-log10 P-value
r2
Top lead SNP
Lead SNPs
Ind. sig.SNPs
Mapped genes
←RALGDS
←OBP2B
RPL7A→
←SURF4
←REXO4
←SLC2A6
←GBGT1
←SURF6
←SURF1
C9orf96→
ADAMTS13
←CACFD1→
SURF2→
←MED22
Chromatin state
E052
E100
E108
E107
E089
E090
E120
E121
E129
TssA
TssAFlnk
TxFlnk
Tx
Tx/Wk
EnhG
Enh
ZNF/Rpts
Het
TssBiv
BivFlnk
EnhBiv
ReprPC
ReprPCWk
Quies
136,050,000
136,100,000
136,150,000
136,200,000
136,250,000
136,300,000
136,350,000
Chromosome 9

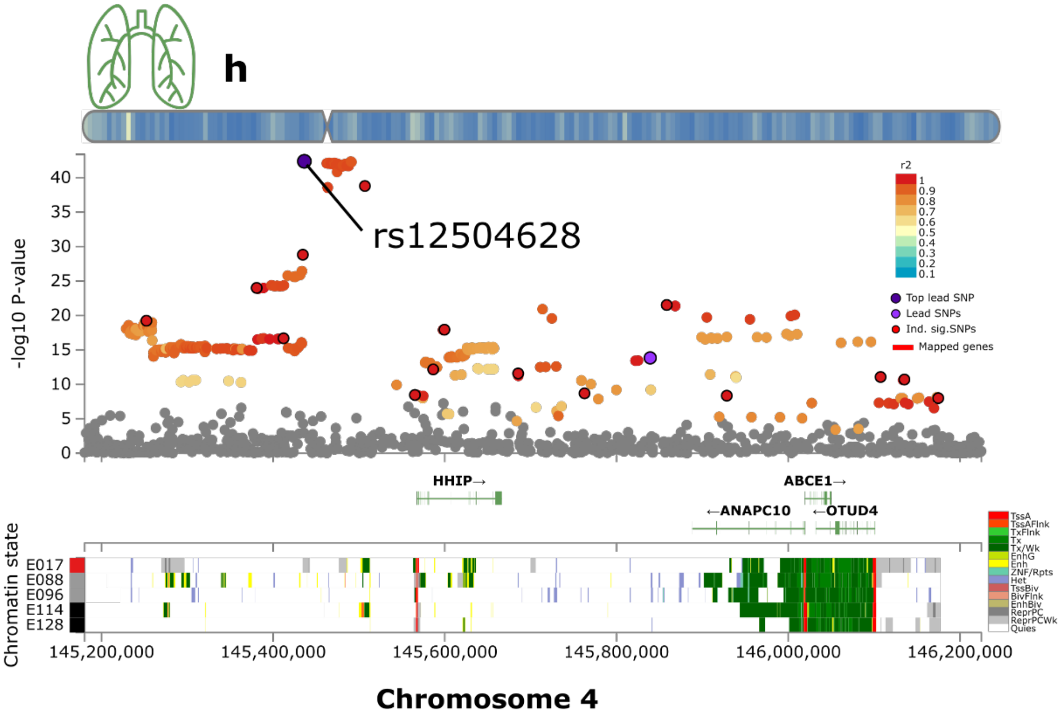
h
rs12504628
-log10 P-value
r2
Top lead SNP
Lead SNPs
Ind. sig.SNPs
Mapped genes
HHIP→
ABCE1→
←ANAPC10
←OTUD4
Chromatin state
E017
E088
E096
E114
E128
TssA
TssAFlnk
TxFlnk
Tx
Tx/Wk
EnhG
Enh
ZNF/Rpts
Het
TssBiv
BivFlnk
EnhBiv
ReprPC
ReprPCWk
Quies
145,200,000
145,400,000
145,600,000
145,800,000
146,000,000
146,200,000
Chromosome 4

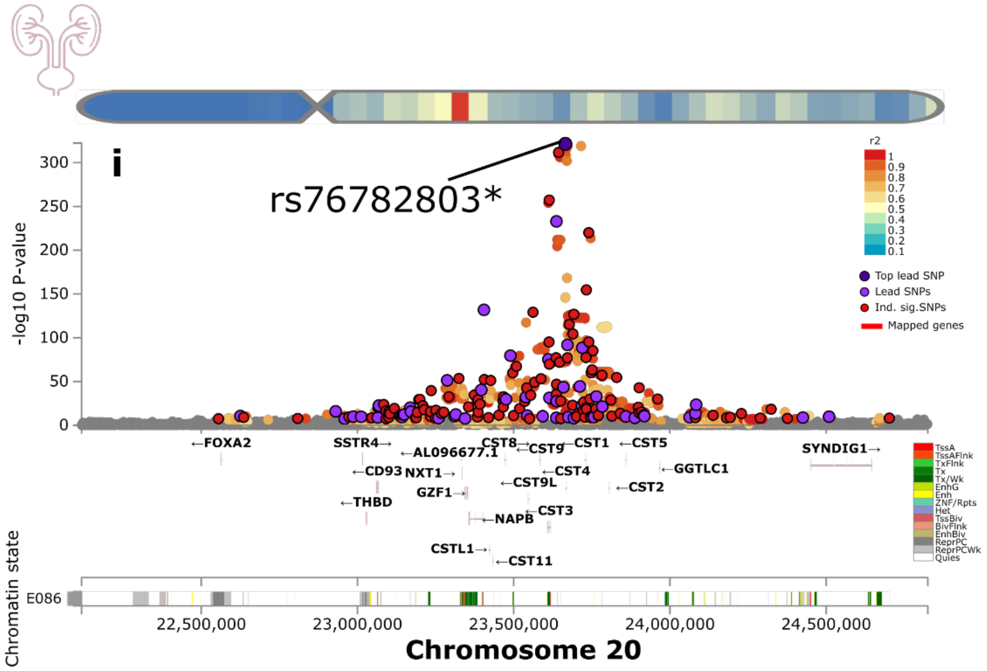

**a-i**) The exemplary genomic locus with the most significant signals for the brain, cardiovascular, eye, hepatic, immune, metabolic, musculoskeletal, pulmonary, and renal BAGs. The top lead SNP, lead SNPs, and independent significant SNPs are annotated within each locus. We mapped the SNPs to the genes and predicted their chromatin states in specific tissues, including the brain for the brain BAG, the heart and vascular tissues for the cardiovascular BAG, the iPSC for the eye BAG, the liver for the hepatic BAG, the spleen, bone, skin, and thymus tissues for the immune BAG, the gastrointestinal tissue for the metabolic BAG, the muscle and bone tissues for the musculoskeletal BAG, the lung tissue for the pulmonary BAG, and the kidney for the renal BAG, respectively.

**eFigure 12: SNP-based heritability, beta coefficients, and alternative allele frequency using the brain-BAG comparable populations and different inclusion criteria for the SNPs**

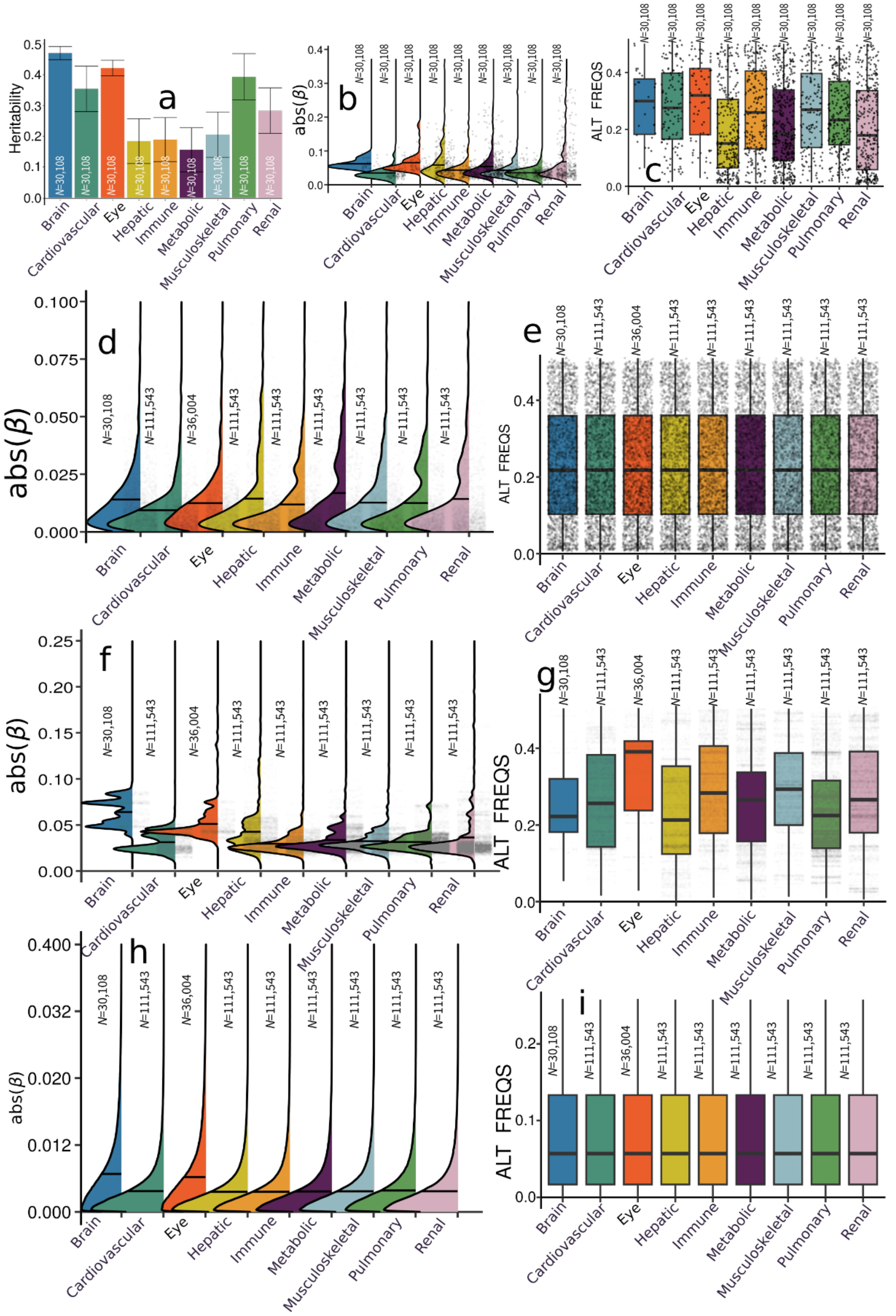

**a**) The SNP-based heritability of the nine BAGs using populations from downsampling to the brain BAG population. Error bars represent the standard error of the estimated parameters. **b**)

The absolute value of the beta coefficients of the independent significant SNPs of the nine BAG GWASs using populations from downsampling to the brain BAG population (*N*=30,108); the independent significant SNPs are shown separately for each BAG. **c**) The alternative (effective) allele frequency of the independent significant SNPs from the nine BAG GWASs using populations from downsampling to the brain BAG population (*N*=30,108). **d**) The beta coefficients of the independent significant SNPs using the original full samples but with all identified independent significant SNPs across the nine BAG GWASs (with the same number of SNPs tested), where we see no difference regarding allele frequency in Figure **e**). **f**) The absolute value of the beta coefficients of the independent significant SNPs plus the candidate SNPs in LD of the nine BAG GWASs using the original full samples; the SNPs are shown separately for each BAG. **g**) The alternative allele frequency for the setting in Figure **f**). **h**) The absolute beta coefficients of the nine BAGs using all genome-wide SNPs (the y-axis was truncated to 0.1 for visualization purposes). **i**) the alternative allele frequency did not differ for Figure h) including all genome-wide SNPs.

**eFigure 13: Trumpet plots of the alternative allele frequency vs. the beta coefficient of the nine BAG GWASs**

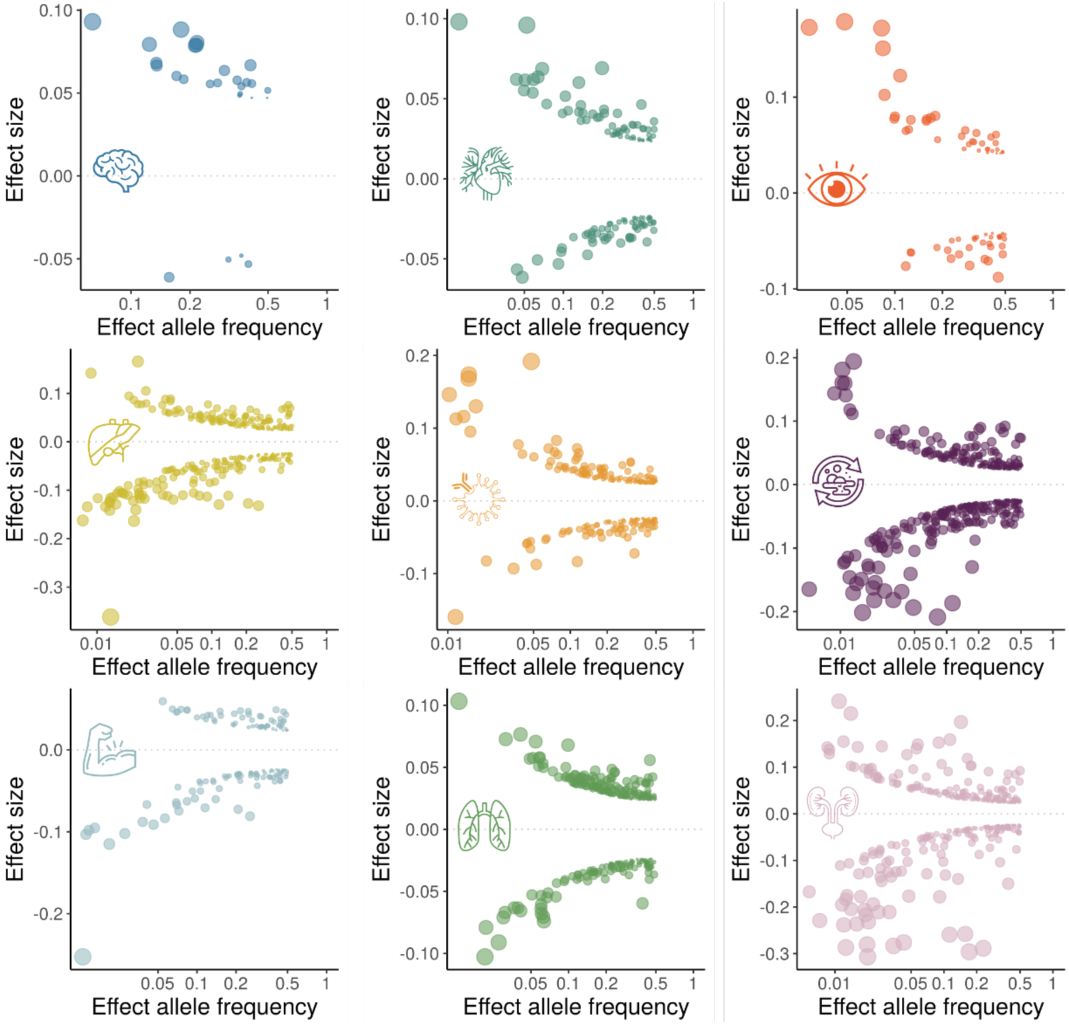

The trumpet plots display the inverse relationship between the alternative (effect) allele frequency and the effect size (beta coefficient) for the brain, cardiovascular, eye, hepatic, immune, metabolic, musculoskeletal, pulmonary, and renal BAGs. Only the independent significant SNPs were considered. The dot size corresponds to the effect size, while the transparency of the dot is proportional to its statistical significance.

**eFigure 14: Manhattan and QQ plots for the four pulmonary features used to compute the pulmonary BAG**

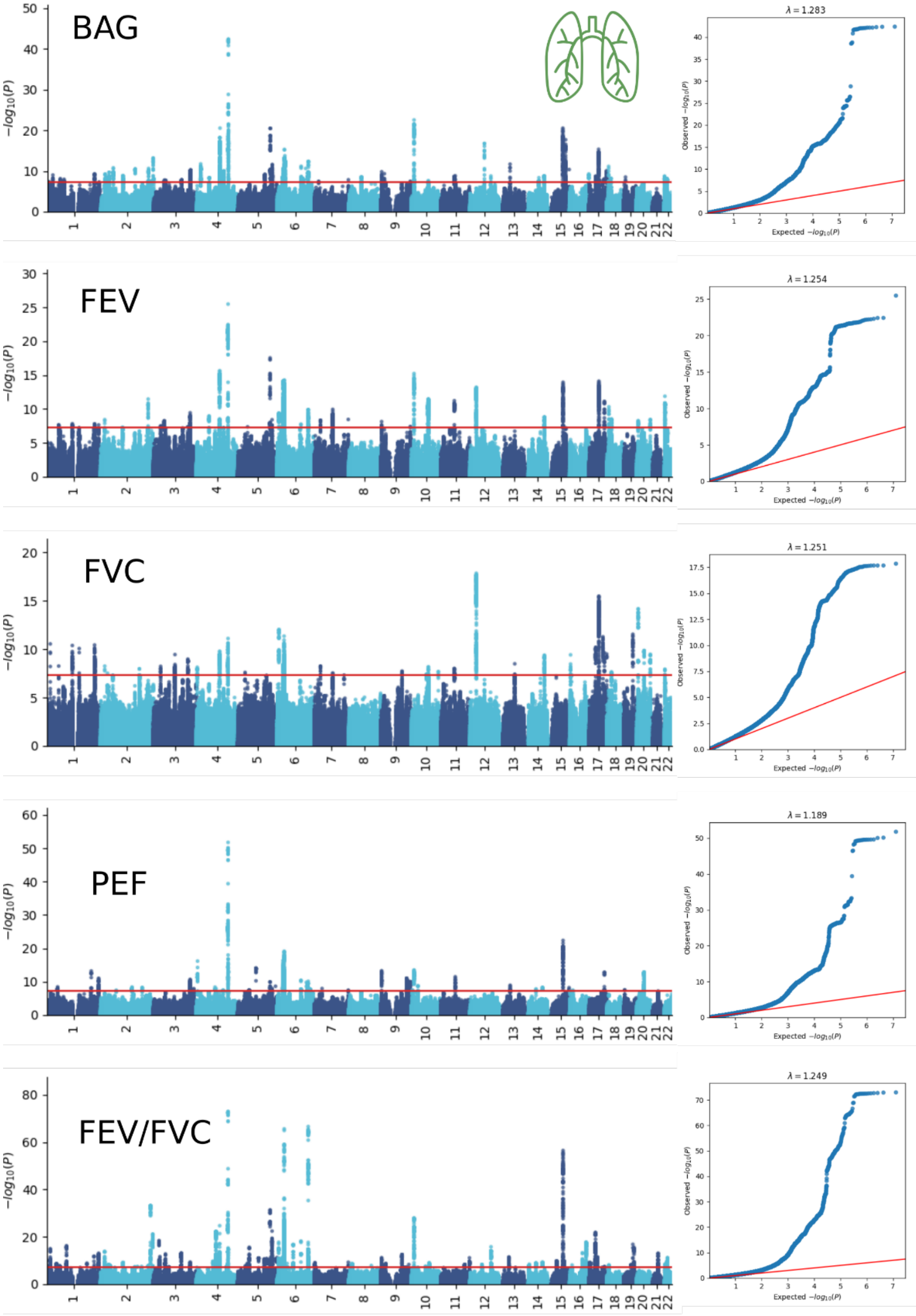

The Manhattan and QQ plots for the pulmonary BAG vs. its four features used to compute the BAG: forced vital capacity (FVC), forced expiratory volume (FEV), peak expiratory flow (PEF), and the ratio of forced expiratory volume to forced vital capacity (FEV/FVC).

**eFigure 15: Bayesian colocalization signal between the pulmonary BAG and FEV/FVC**

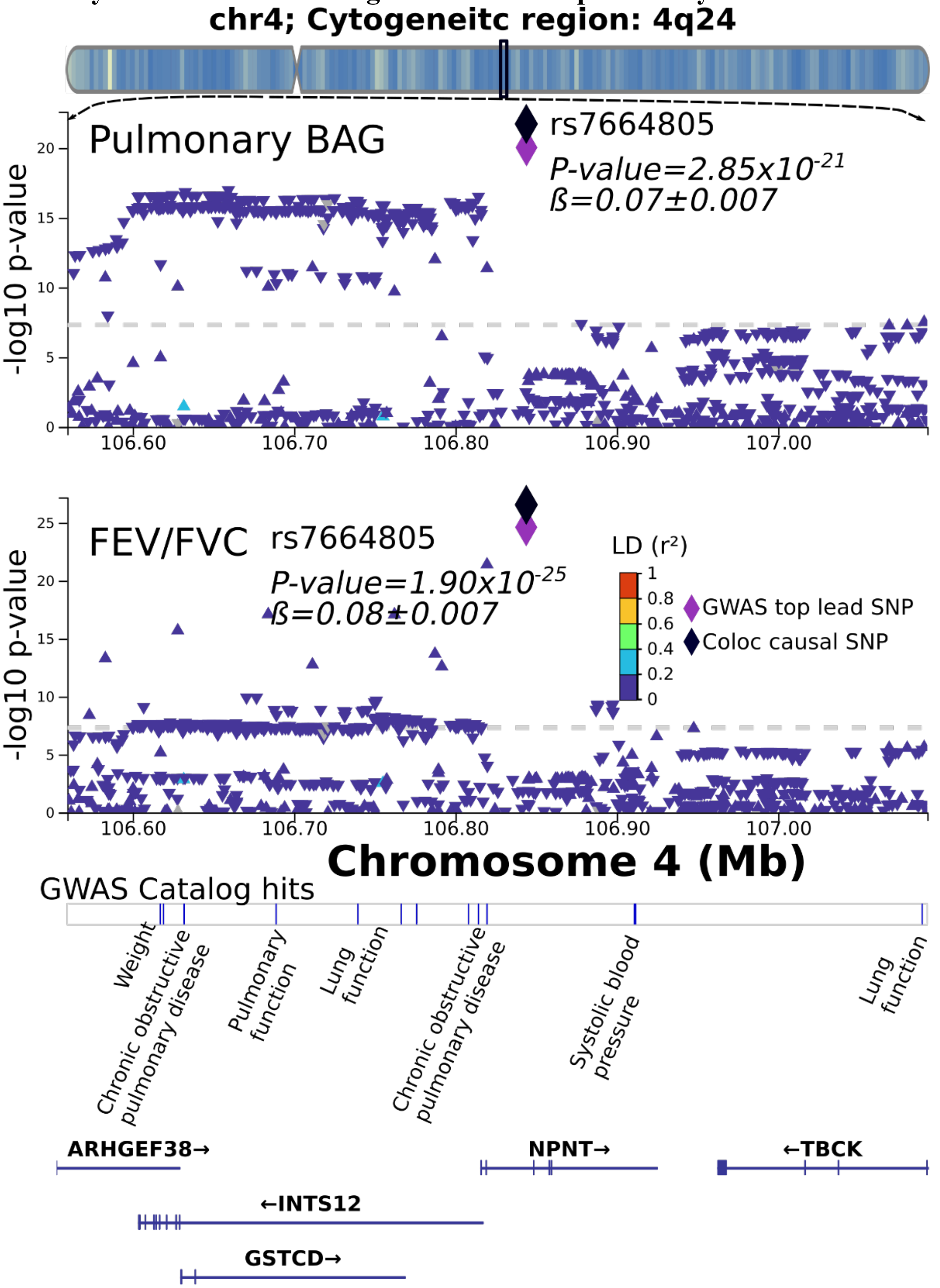

We illustrate here the colocalization signal between the pulmonary BAG and the FEV/FCV feature at the genomic locus: 4q24 with the top lead SNP (causal SNP: rs7664805). Genetic colocalization was evidenced at one locus (4q24) between the pulmonary BAG and the FEV/FCV feature. The signed PP.H4.ABF (0.99) denotes the posterior probability (PP) of hypothesis H4, which suggests that both traits share the same causal SNP (rs7664805).

**eFigure 16: Beta coefficients of the significant colocalization signal between the pulmonary BAG and the four pulmonary features**

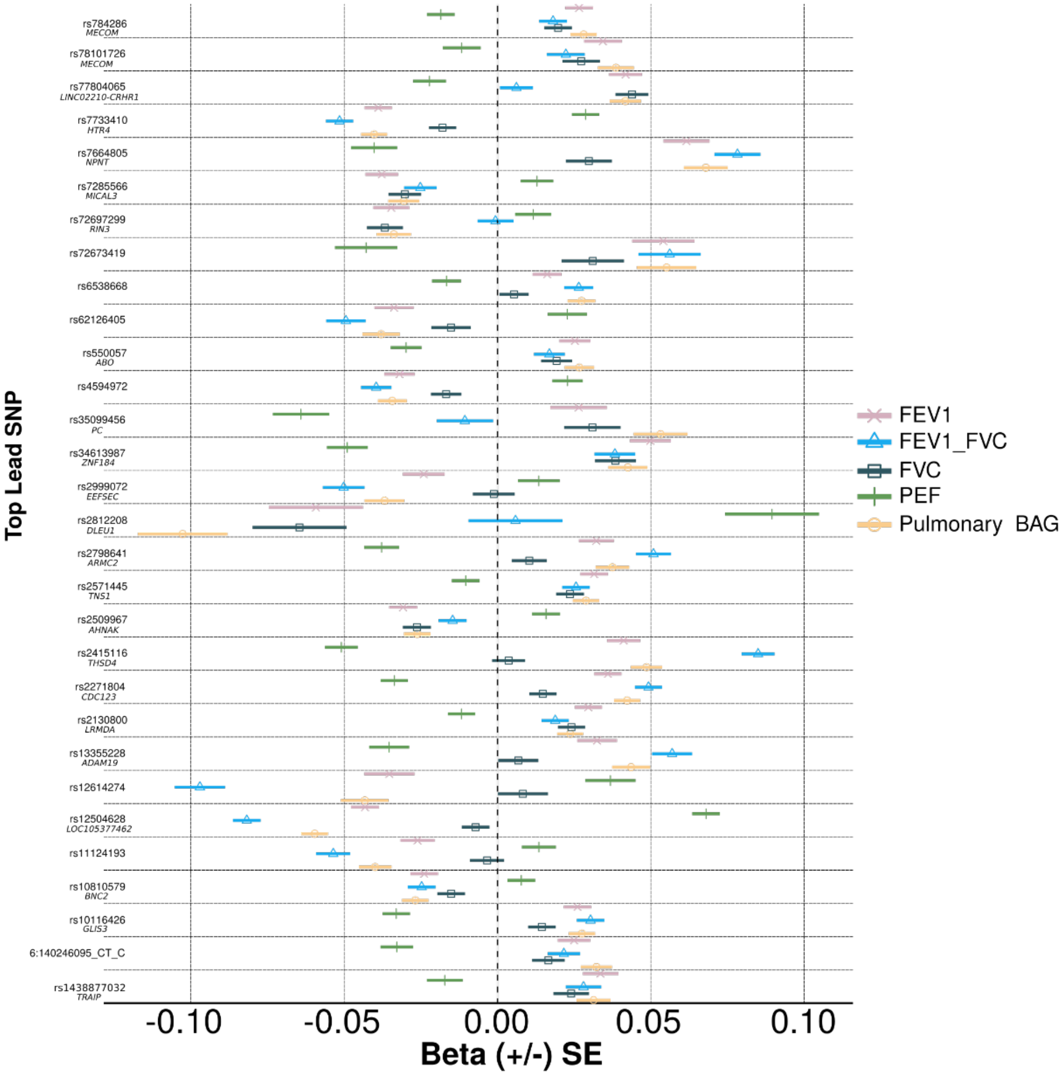

We show the beta coefficients of the significant colocalization signals between the pulmonary BAG and its underlying four pulmonary features. We ensured that at least one of the four pulmonary features achieved the genome-wide P-value threshold, totaling 48 loci (represented by its top lead SNP). We also showed the mapped gene when available.

**eFigure 17: GSEA using sex-stratified GWAS results**

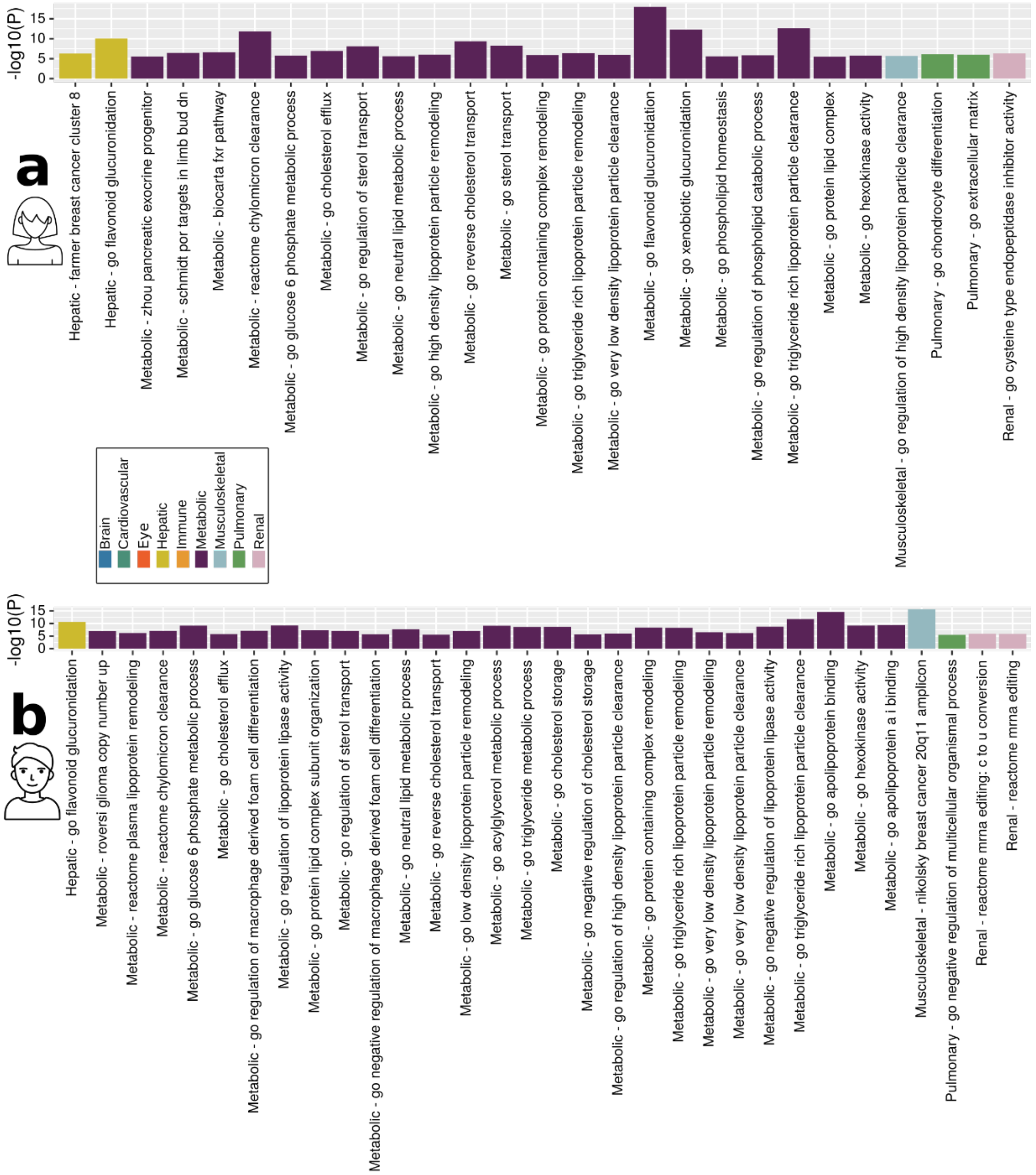

Gene-set enrichment analysis was performed using the GWAS summary statistics specific to females (**a**) and males (**b**).

**eFigure 18: TEA correlations using sex-stratified GWAS results**

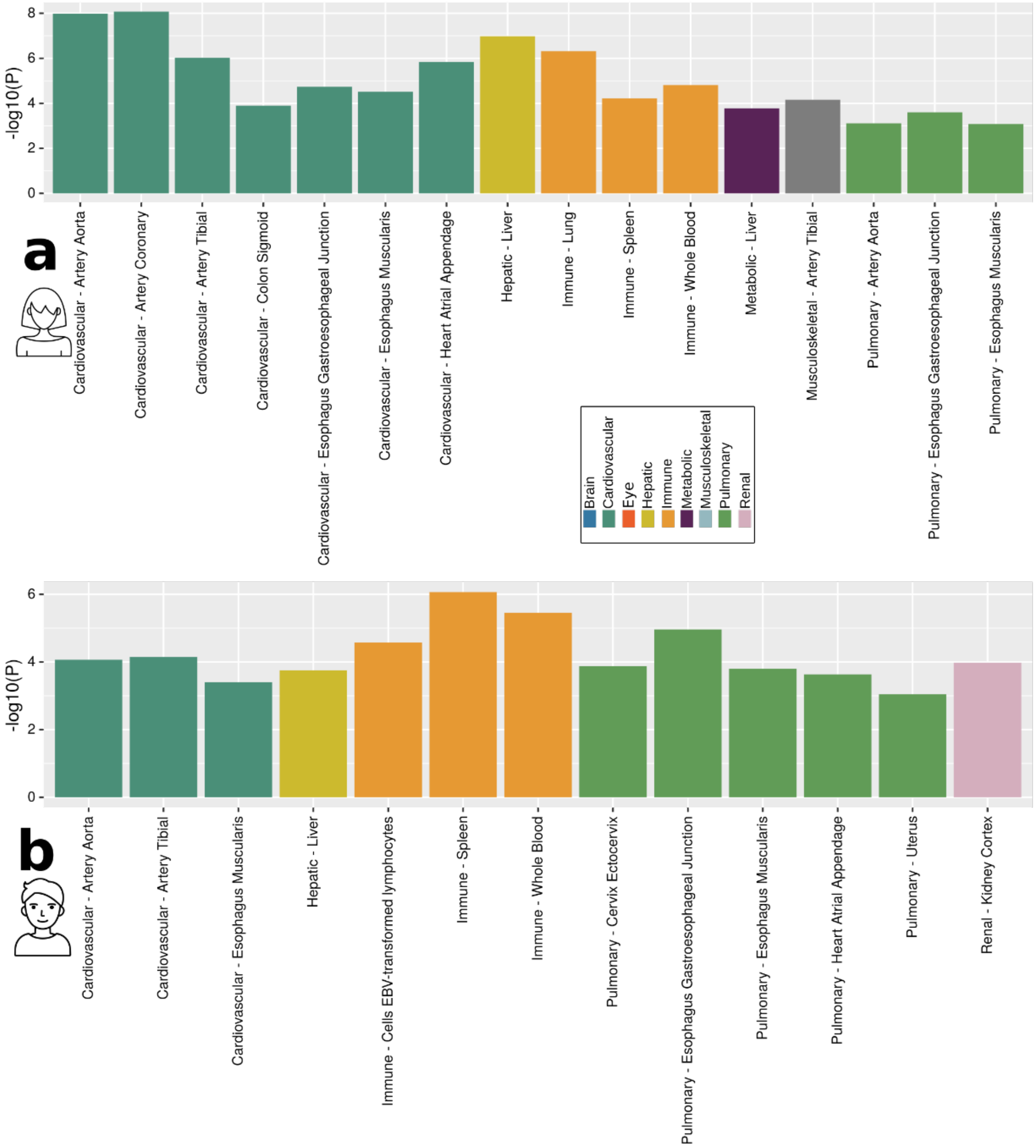

Tissue-specific enrichment analysis was performed using the GWAS summary statistics specific to females (**a**) and males (**b**).

**eFigure 19: Genetic correlations using sex-stratified GWAS results**

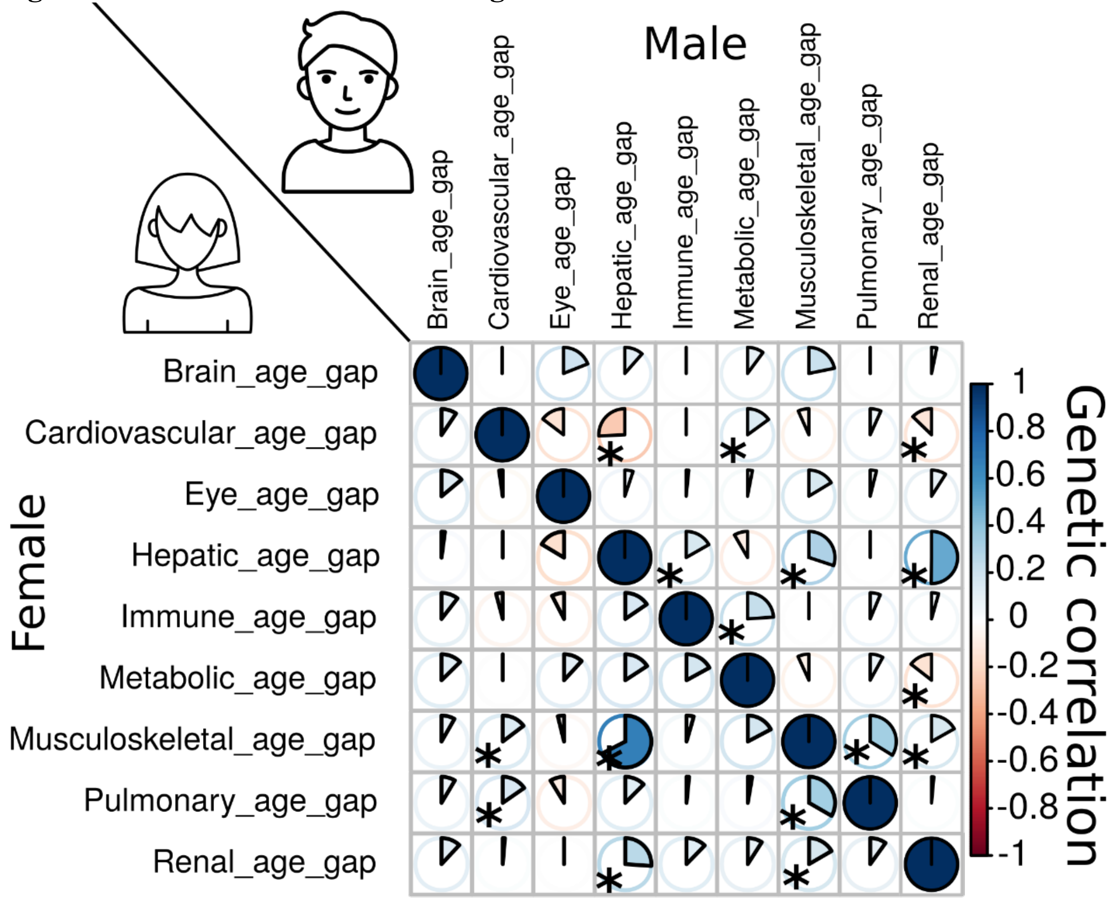

The Genetic correlation between each pair of BAGs using sex-stratified GWAS summary statistics from our analyses. Most of the genetic correlations showed consistency between females and males, albeit sex differences are evident in certain BAGs, particularly in the cardiovascular BAG results. Specifically, males exhibit dominant correlations between cardiovascular BAGs and hepatic and renal BAGs, while females demonstrate specific correlations with musculoskeletal and pulmonary BAGs.

**eFigure 20: Mendelian randomization sensitivity check for the hepatic BAG on the musculoskeletal BAG**

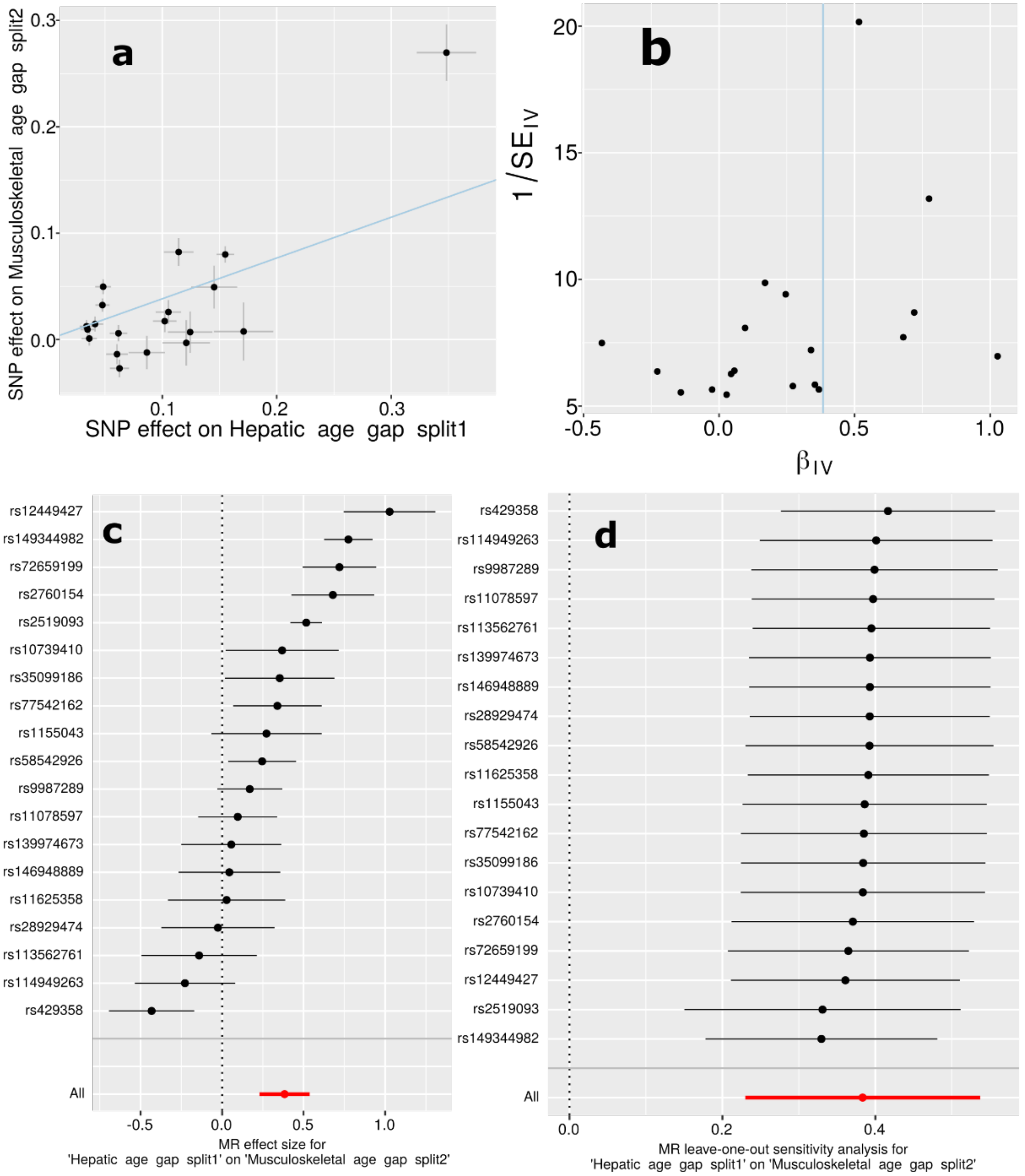

**a**) Scatter plot for the MR effect sizes of the exposure variable (hepatic BAG, *x*-axis, SD units) and the outcome variable (musculoskeletal BAG, *y*-axis, log OR) with standard error bars. The slopes of the regression line correspond to the causal effect sizes estimated by the IVW estimator. **b**) Funnel plot for the relationship between the causal effect of the exposure variable on the outcome variable. Each dot represents MR effect sizes estimated using each SNP as a separate instrument against the inverse of the standard error of the causal estimate. The vertical red line shows the MR estimates using all SNPs. **c**) Forest plot for the single-SNP MR results. Each line represents the MR effect (log OR) for the exposure variable on the outcome variable using only one SNP; the red line shows the MR effect using all SNPs together. **d**) Leave-one-out analysis of the exposure variable on the outcome variable. Each row represents the MR effect

(log OR) and the 95% CI by excluding that SNP from the analysis. The red line depicts the IVW estimator using all SNPs.

**eFigure 21: Mendelian randomization sensitivity check for the musculoskeletal BAG on the hepatic BAG**

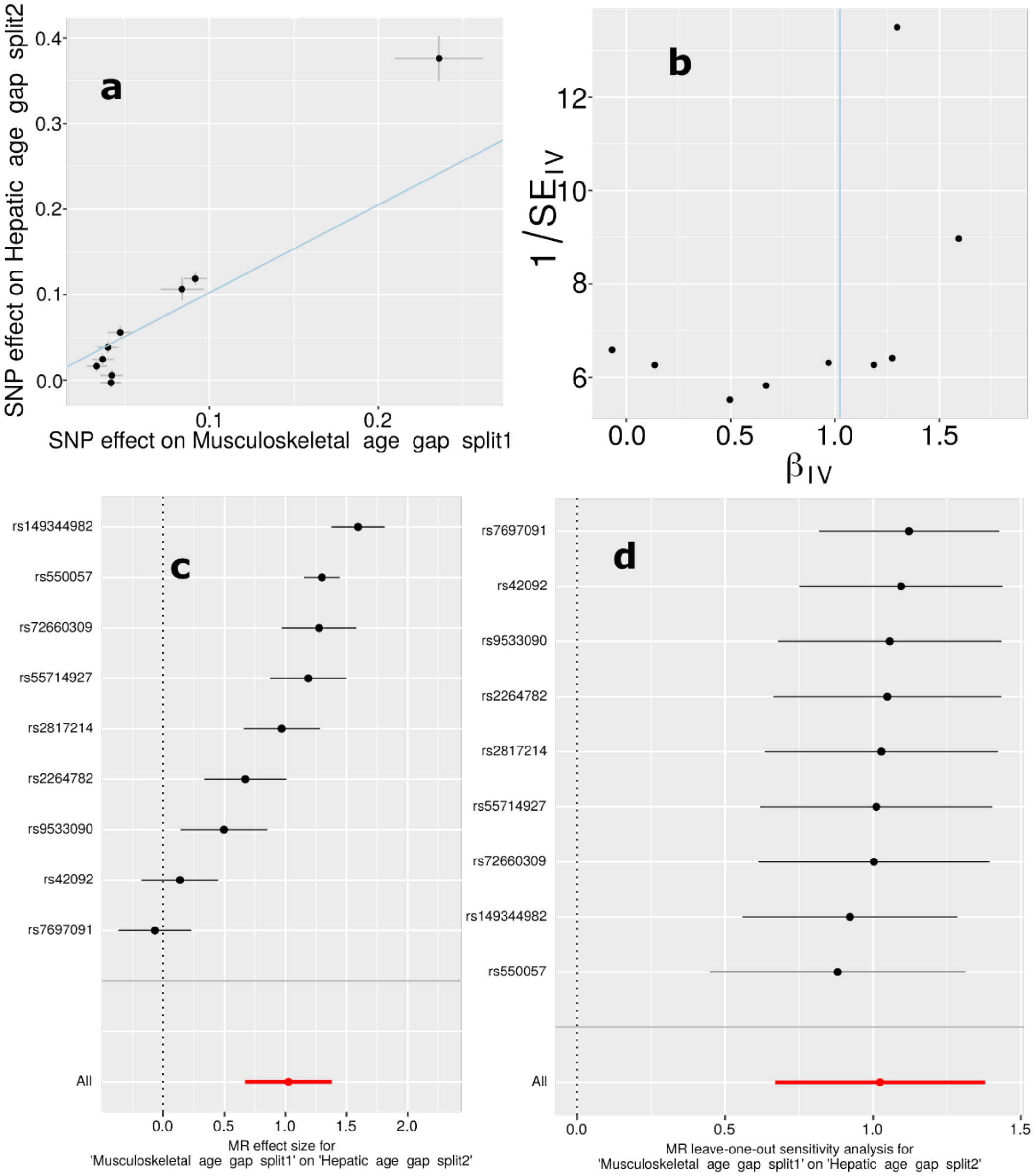

**a**) Scatter plot for the MR effect sizes of the exposure variable (musculoskeletal BAG, *x*-axis, SD units) and the outcome variable (hepatic BAG, *y*-axis, log OR) with standard error bars. The slopes of the regression line correspond to the causal effect sizes estimated by the IVW estimator. **b**) Funnel plot for the relationship between the causal effect of the exposure variable on the outcome variable. Each dot represents MR effect sizes estimated using each SNP as a separate instrument against the inverse of the standard error of the causal estimate. The vertical red line shows the MR estimates using all SNPs. **c**) Forest plot for the single-SNP MR results. Each line represents the MR effect (log OR) for the exposure variable on the outcome variable using only one SNP; the red line shows the MR effect using all SNPs together. **d**) Leave-one-out analysis of the exposure variable on the outcome variable. Each row represents the MR effect

(log OR) and the 95% CI by excluding that SNP from the analysis. The red line depicts the IVW estimator using all SNPs.

**eFigure 22: Mendelian randomization sensitivity check for AD on the brain BAG**

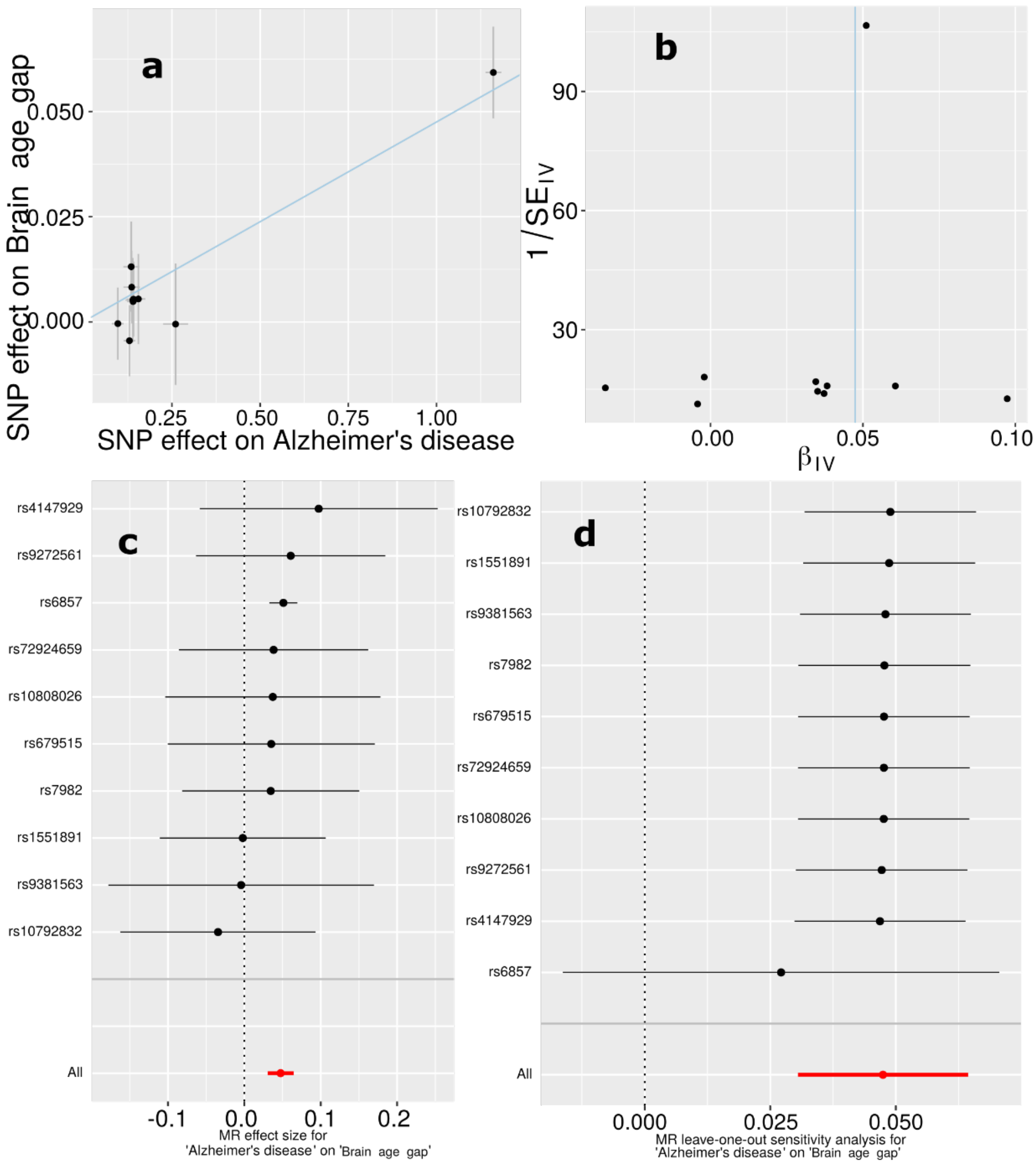

**a**) Scatter plot for the MR effect sizes of the exposure variable (AD, *x*-axis, SD units) and the outcome variable (brain BAG, *y*-axis, log OR) with standard error bars. The slopes of the regression line correspond to the causal effect sizes estimated by the IVW estimator. **b**) Funnel plot for the relationship between the causal effect of the exposure variable on the outcome variable. Each dot represents MR effect sizes estimated using each SNP as a separate instrument against the inverse of the standard error of the causal estimate. The vertical red line shows the MR estimates using all SNPs. **c**) Forest plot for the single-SNP MR results. Each line represents the MR effect (log OR) for the exposure variable on the outcome variable using only one SNP; the red line shows the MR effect using all SNPs together. **d**) Leave-one-out analysis of the exposure variable on the outcome variable. Each row represents the MR effect (log OR) and the 95% CI by excluding that SNP from the analysis. The red line depicts the IVW estimator using all SNPs.

**eFigure 23: Mendelian randomization sensitivity check for AD on the hepatic BAG**

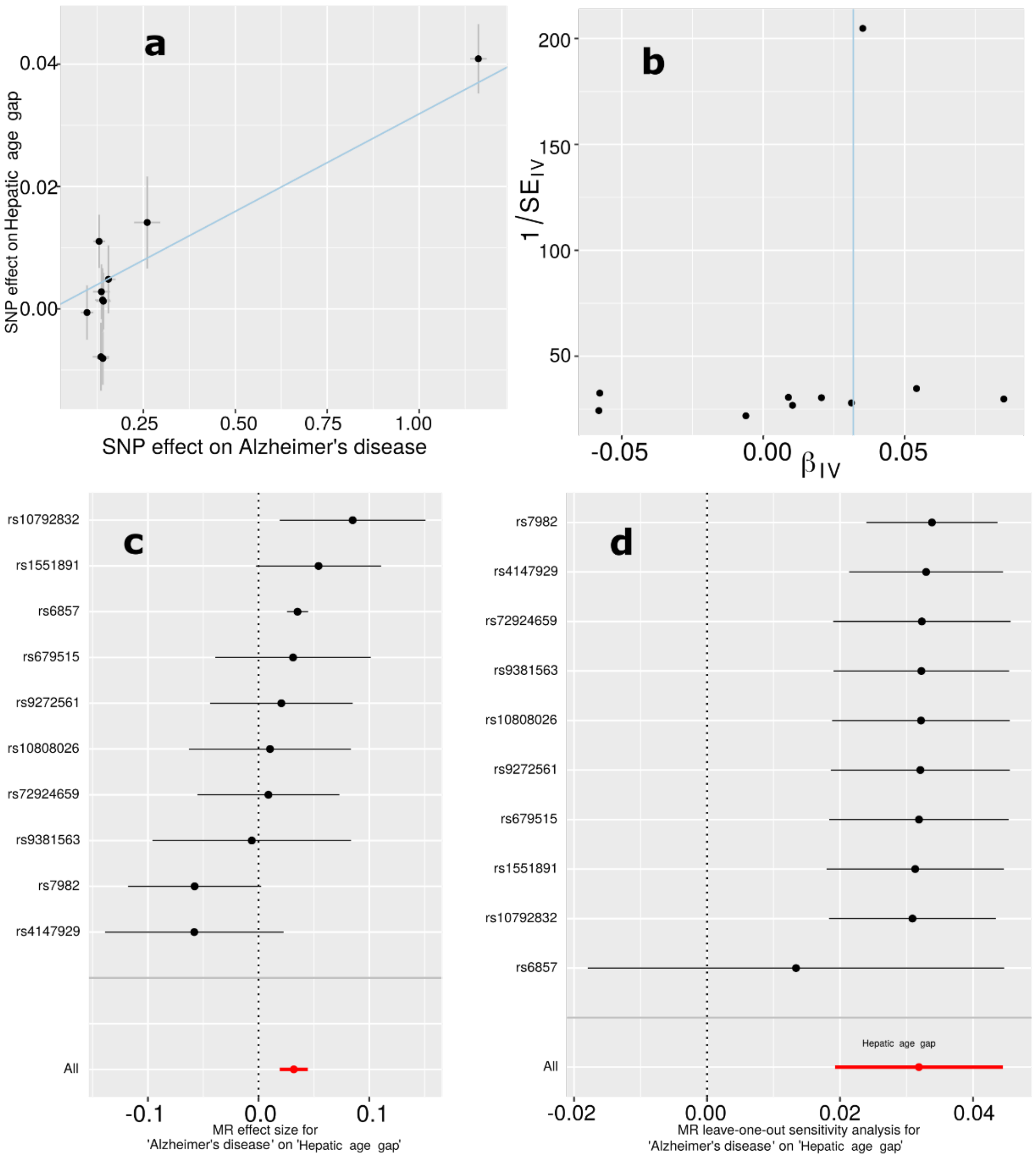

**a**) Scatter plot for the MR effect sizes of the exposure variable (AD, *x*-axis, SD units) and the outcome variable (hepatic BAG, *y*-axis, log OR) with standard error bars. The slopes of the regression line correspond to the causal effect sizes estimated by the IVW estimator. **b**) Funnel plot for the relationship between the causal effect of the exposure variable on the outcome variable. Each dot represents MR effect sizes estimated using each SNP as a separate instrument against the inverse of the standard error of the causal estimate. The vertical red line shows the MR estimates using all SNPs. **c**) Forest plot for the single-SNP MR results. Each line represents the MR effect (log OR) for the exposure variable on the outcome variable using only one SNP; the red line shows the MR effect using all SNPs together. **d**) Leave-one-out analysis of the exposure variable on the outcome variable. Each row represents the MR effect (log OR) and the 95% CI by excluding that SNP from the analysis. The red line depicts the IVW estimator using all SNPs.

539

**eFigure 24: Mendelian randomization sensitivity check for Crohn's disease on the hepatic BAG**

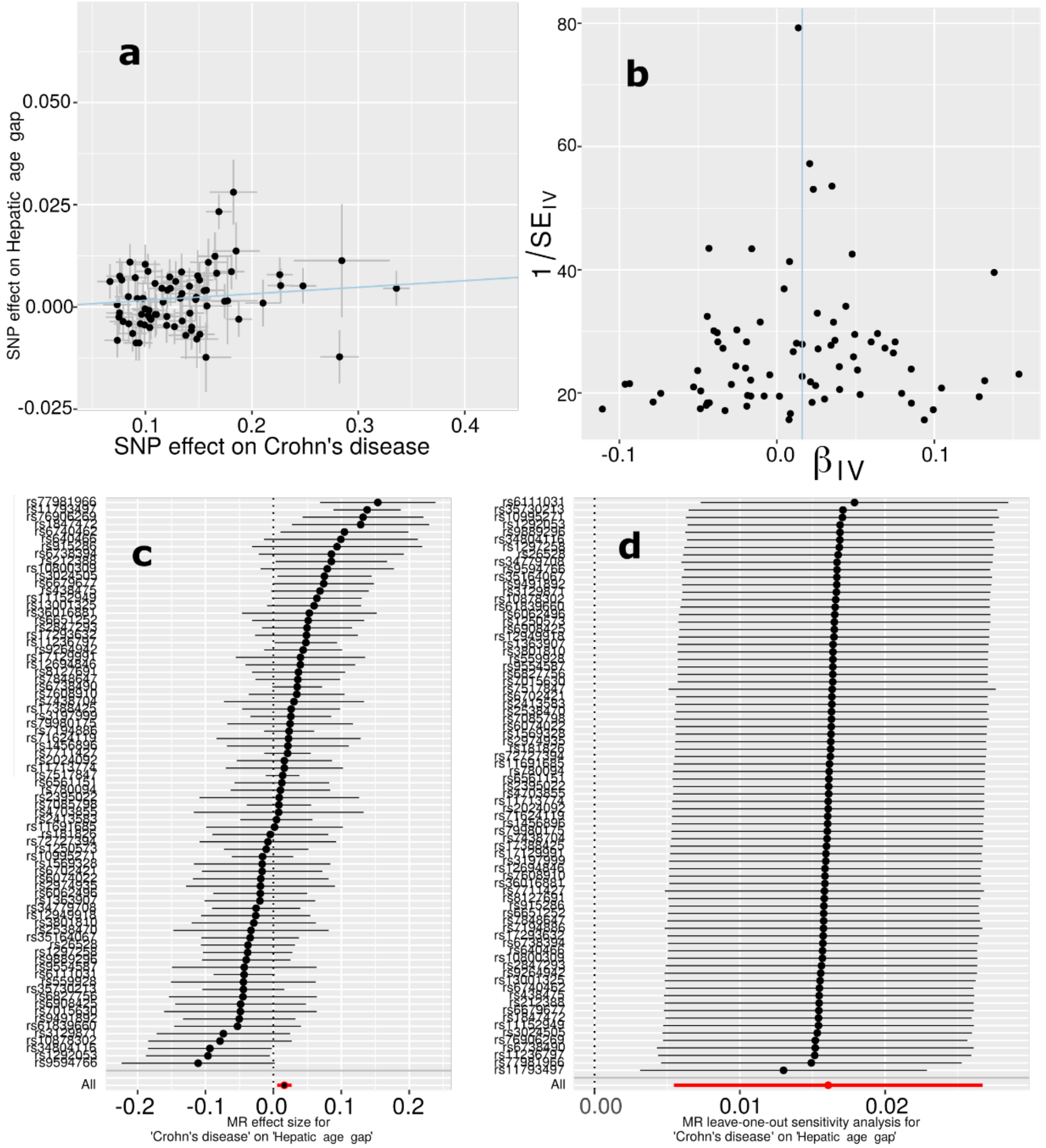

**a**) Scatter plot for the MR effect sizes of the exposure variable (Crohn's disease, *x*-axis, SD units) and the outcome variable (hepatic BAG, *y*-axis, log OR) with standard error bars. The slopes of the regression line correspond to the causal effect sizes estimated by the IVW estimator. **b**) Funnel plot for the relationship between the causal effect of the exposure variable on the outcome variable. Each dot represents MR effect sizes estimated using each SNP as a separate instrument against the inverse of the standard error of the causal estimate. The vertical red line shows the MR estimates using all SNPs. **c**) Forest plot for the single-SNP MR results. Each line represents the MR effect (log OR) for the exposure variable on the outcome variable using only one SNP; the red line shows the MR effect using all SNPs together. **d**) Leave-one-out analysis of the exposure variable on the outcome variable. Each row represents the MR effect (log OR) and

the 95% CI by excluding that SNP from the analysis. The red line depicts the IVW estimator using all SNPs.

**eFigure 25: Mendelian randomization sensitivity check for body weight on the immune BAG**

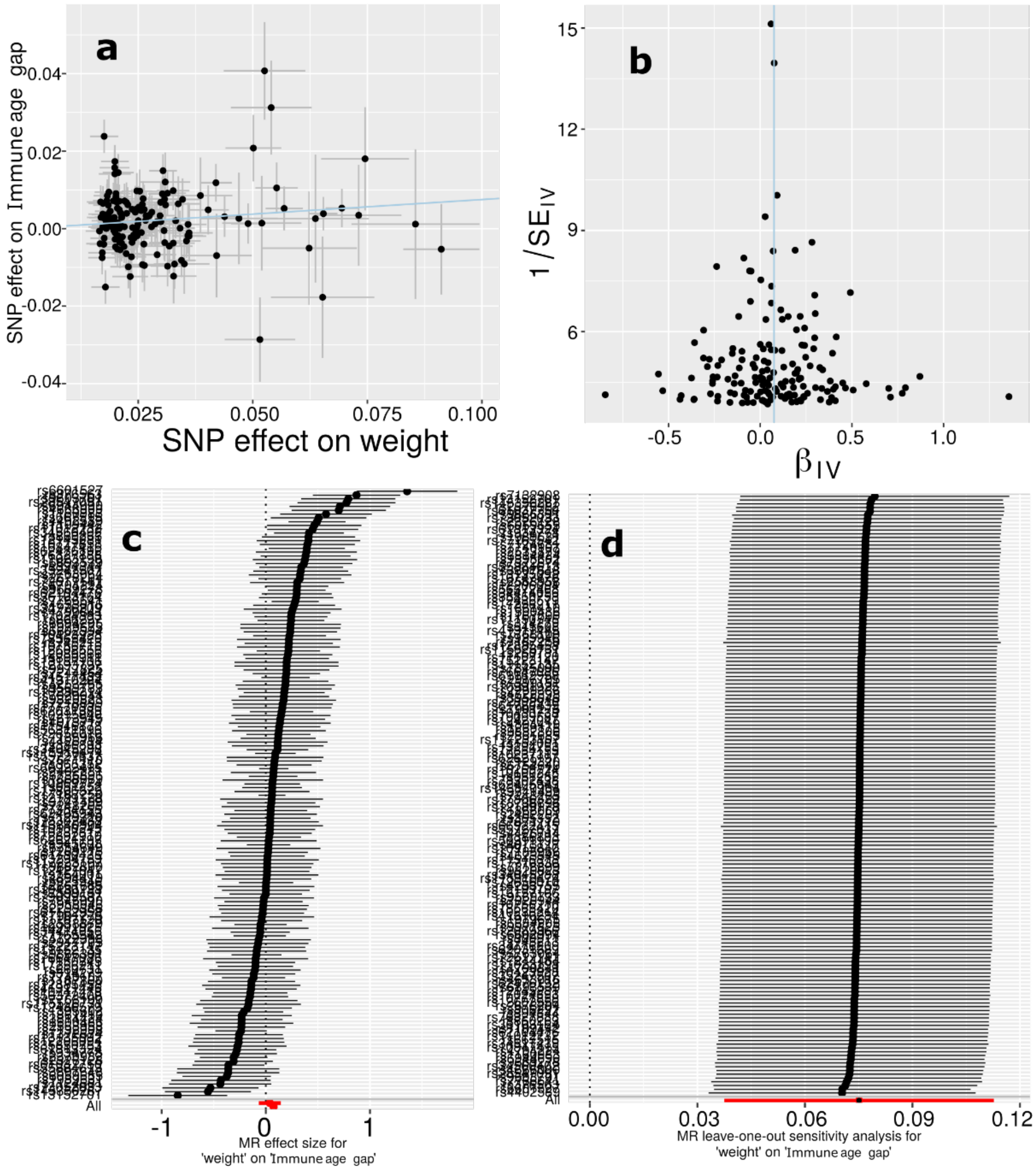

**a**) Scatter plot for the MR effect sizes of the exposure variable (body weight, *x*-axis, SD units) and the outcome variable (immune BAG, *y*-axis, log OR) with standard error bars. The slopes of the regression line correspond to the causal effect sizes estimated by the IVW estimator. **b**) Funnel plot for the relationship between the causal effect of the exposure variable on the outcome variable. Each dot represents MR effect sizes estimated using each SNP as a separate instrument against the inverse of the standard error of the causal estimate. The vertical red line shows the MR estimates using all SNPs. **c**) Forest plot for the single-SNP MR results. Each line represents the MR effect (log OR) for the exposure variable on the outcome variable using only one SNP; the red line shows the MR effect using all SNPs together. **d**) Leave-one-out analysis of the exposure variable on the outcome variable. Each row represents the MR effect (log OR) and

the 95% CI by excluding that SNP from the analysis. The red line depicts the IVW estimator using all SNPs.

**eFigure 26: Mendelian randomization sensitivity check for type 2 diabetes on the metabolic BAG**

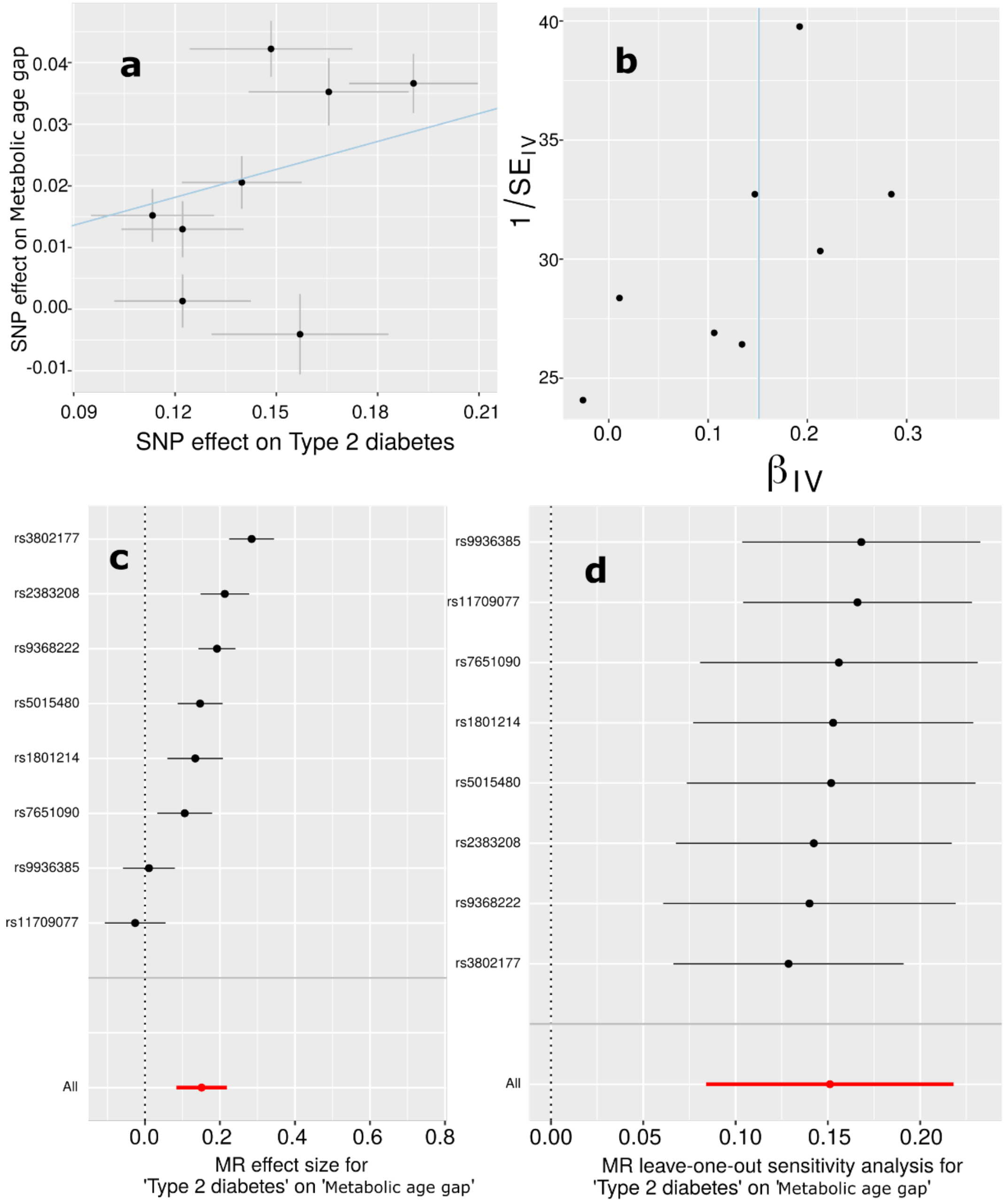

**a**) Scatter plot for the MR effect sizes of the exposure variable (type 2 diabetes, *x*-axis, SD units) and the outcome variable (metabolic BAG, *y*-axis, log OR) with standard error bars. The slopes of the regression line correspond to the causal effect sizes estimated by the IVW estimator. **b**) Funnel plot for the relationship between the causal effect of the exposure variable on the outcome variable. Each dot represents MR effect sizes estimated using each SNP as a separate instrument against the inverse of the standard error of the causal estimate. The vertical red line shows the MR estimates using all SNPs. **c**) Forest plot for the single-SNP MR results. Each line represents the MR effect (log OR) for the exposure variable on the outcome variable using only one SNP; the red line shows the MR effect using all SNPs together. **d**) Leave-one-out analysis of the exposure variable on the outcome variable. Each row represents the MR effect (log OR) and the 95% CI by excluding that SNP from the analysis. The red line depicts the IVW estimator using all SNPs.

**eFigure 27: Mendelian randomization sensitivity check for AD on the musculoskeletal BAG**

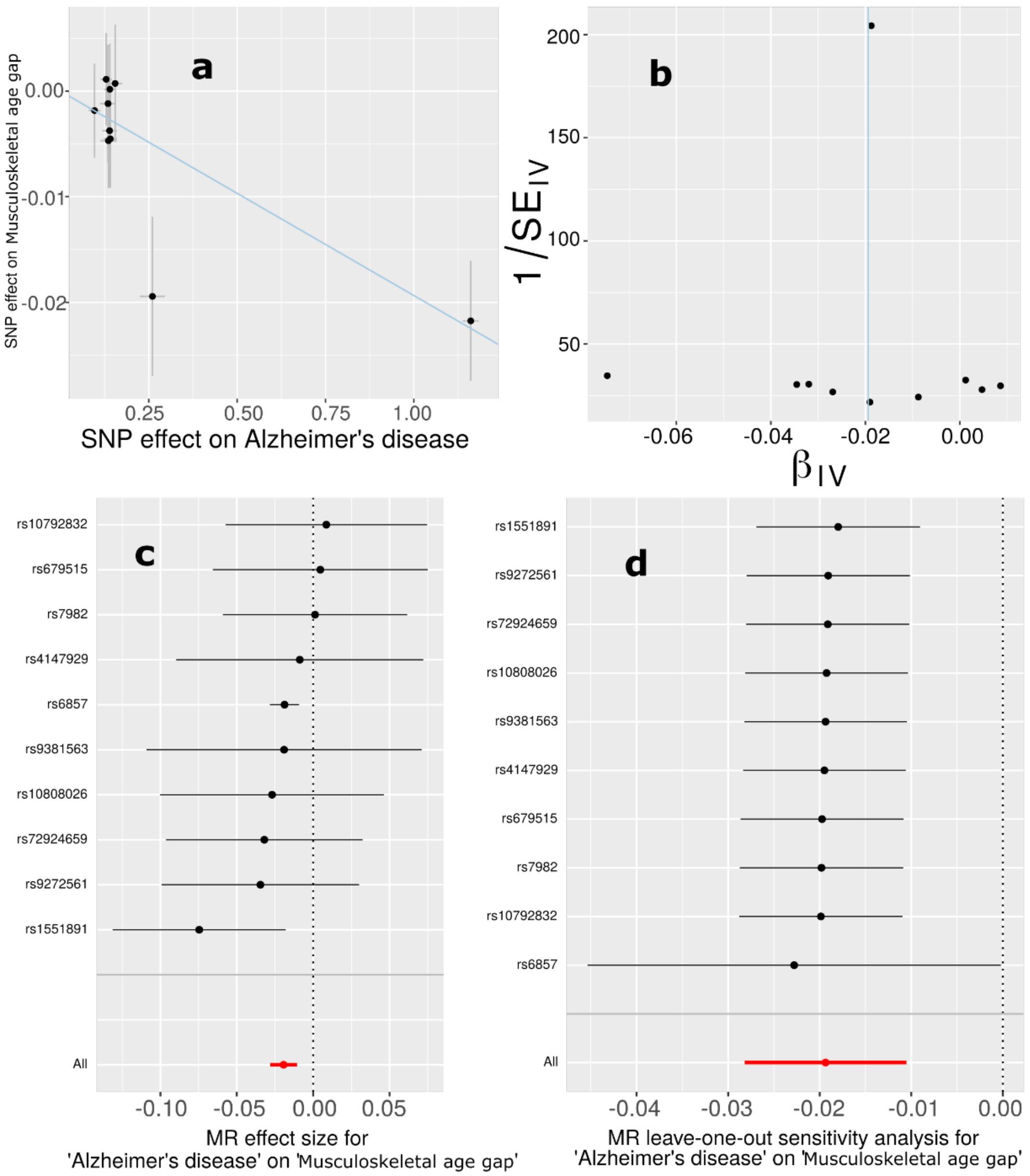

**a**) Scatter plot for the MR effect sizes of the exposure variable (AD, *x*-axis, SD units) and the outcome variable (musculoskeletal BAG, *y*-axis, log OR) with standard error bars. The slopes of the regression line correspond to the causal effect sizes estimated by the IVW estimator. **b**) Funnel plot for the relationship between the causal effect of the exposure variable on the outcome variable. Each dot represents MR effect sizes estimated using each SNP as a separate instrument against the inverse of the standard error of the causal estimate. The vertical red line shows the MR estimates using all SNPs. **c**) Forest plot for the single-SNP MR results. Each line represents the MR effect (log OR) for the exposure variable on the outcome variable using only one SNP; the red line shows the MR effect using all SNPs together. **d**) Leave-one-out analysis of the exposure variable on the outcome variable. Each row represents the MR effect (log OR) and the 95% CI by excluding that SNP from the analysis. The red line depicts the IVW estimator using all SNPs.

602

**eFigure 28: Mendelian randomization sensitivity check for IBD on the musculoskeletal BAG**

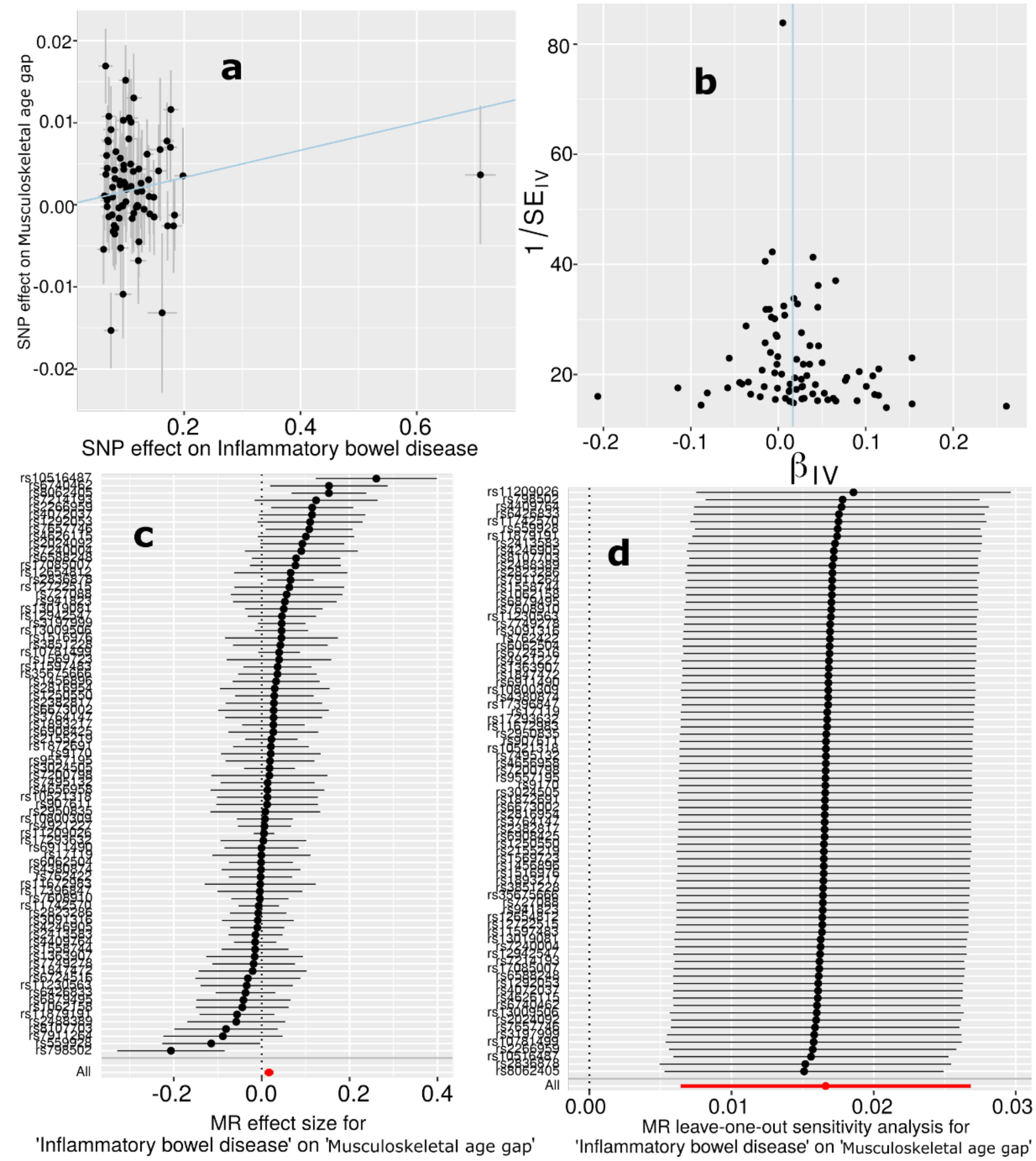

**a**) Scatter plot for the MR effect sizes of the exposure variable (IBD, *x*-axis, SD units) and the outcome variable (musculoskeletal BAG, *y*-axis, log OR) with standard error bars. The slopes of the regression line correspond to the causal effect sizes estimated by the IVW estimator. **b**) Funnel plot for the relationship between the causal effect of the exposure variable on the outcome variable. Each dot represents MR effect sizes estimated using each SNP as a separate instrument against the inverse of the standard error of the causal estimate. The vertical red line shows the MR estimates using all SNPs. **c**) Forest plot for the single-SNP MR results. Each line represents the MR effect (log OR) for the exposure variable on the outcome variable using only one SNP; the red line shows the MR effect using all SNPs together. **d**) Leave-one-out analysis of the exposure variable on the outcome variable. Each row represents the MR effect (log OR) and

the 95% CI by excluding that SNP from the analysis. The red line depicts the IVW estimator using all SNPs.

**eFigure 29: Mendelian randomization sensitivity check for PBC on the musculoskeletal BAG**

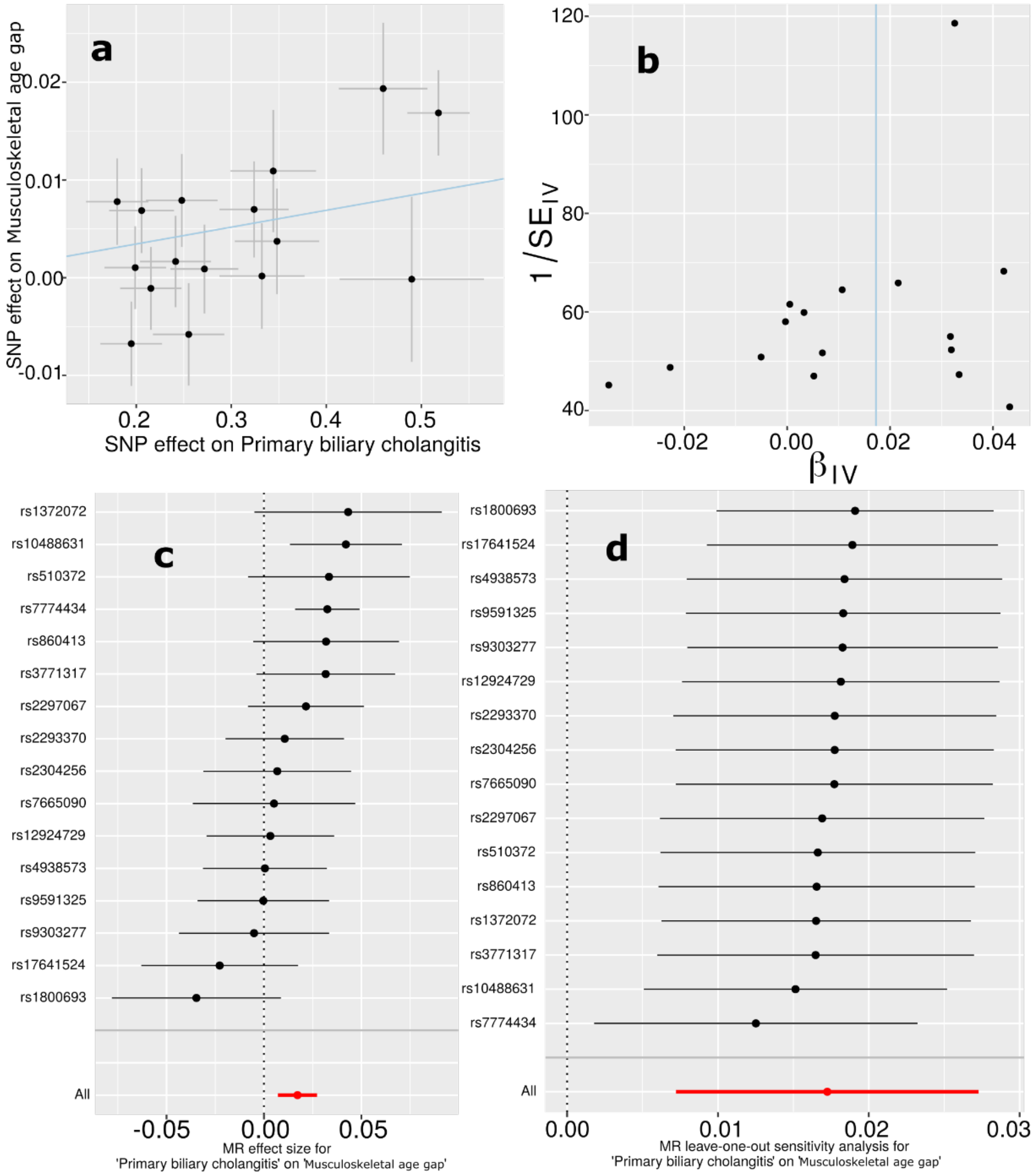

**a**) Scatter plot for the MR effect sizes of the exposure variable (PBC, *x*-axis, SD units) and the outcome variable (musculoskeletal BAG, *y*-axis, log OR) with standard error bars. The slopes of the regression line correspond to the causal effect sizes estimated by the IVW estimator. **b**) Funnel plot for the relationship between the causal effect of the exposure variable on the outcome variable. Each dot represents MR effect sizes estimated using each SNP as a separate instrument against the inverse of the standard error of the causal estimate. The vertical red line shows the MR estimates using all SNPs. **c**) Forest plot for the single-SNP MR results. Each line represents the MR effect (log OR) for the exposure variable on the outcome variable using only one SNP; the red line shows the MR effect using all SNPs together. **d**) Leave-one-out analysis of the exposure variable on the outcome variable. Each row represents the MR effect (log OR) and

the 95% CI by excluding that SNP from the analysis. The red line depicts the IVW estimator using all SNPs.

**eFigure 30: Mendelian randomization sensitivity check for weight on the musculoskeletal BAG**

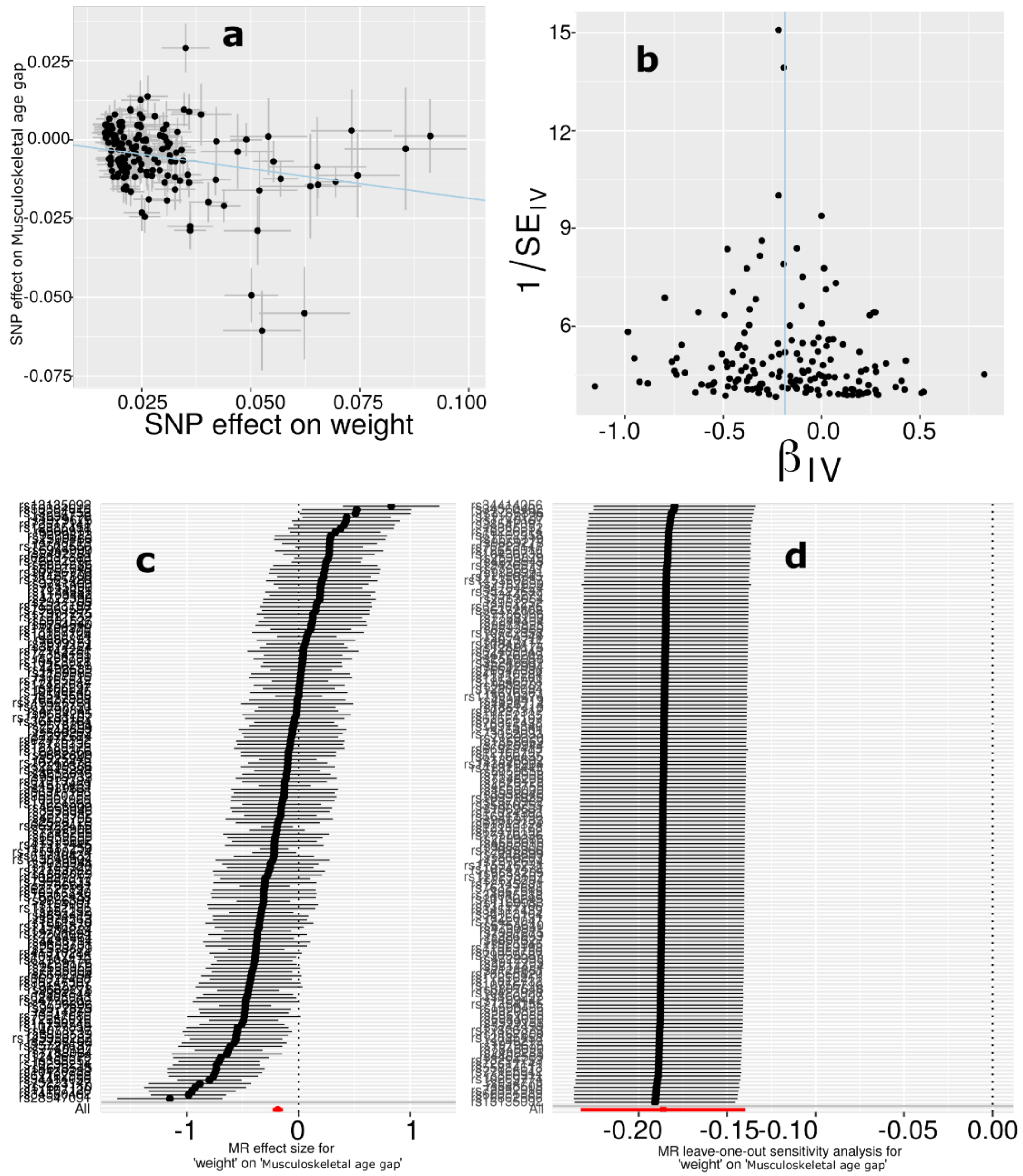

**a**) Scatter plot for the MR effect sizes of the exposure variable (body weight, *x*-axis, SD units) and the outcome variable (musculoskeletal BAG, *y*-axis, log OR) with standard error bars. The slopes of the regression line correspond to the causal effect sizes estimated by the IVW estimator. **b**) Funnel plot for the relationship between the causal effect of the exposure variable on the outcome variable. Each dot represents MR effect sizes estimated using each SNP as a separate instrument against the inverse of the standard error of the causal estimate. The vertical red line shows the MR estimates using all SNPs. **c**) Forest plot for the single-SNP MR results. Each line represents the MR effect (log OR) for the exposure variable on the outcome variable using only one SNP; the red line shows the MR effect using all SNPs together. **d**) Leave-one-out analysis of the exposure variable on the outcome variable. Each row represents the MR effect

(log OR) and the 95% CI by excluding that SNP from the analysis. The red line depicts the IVW estimator using all SNPs.

**eFigure 31: Mendelian randomization sensitivity check for weight on the pulmonary BAG**

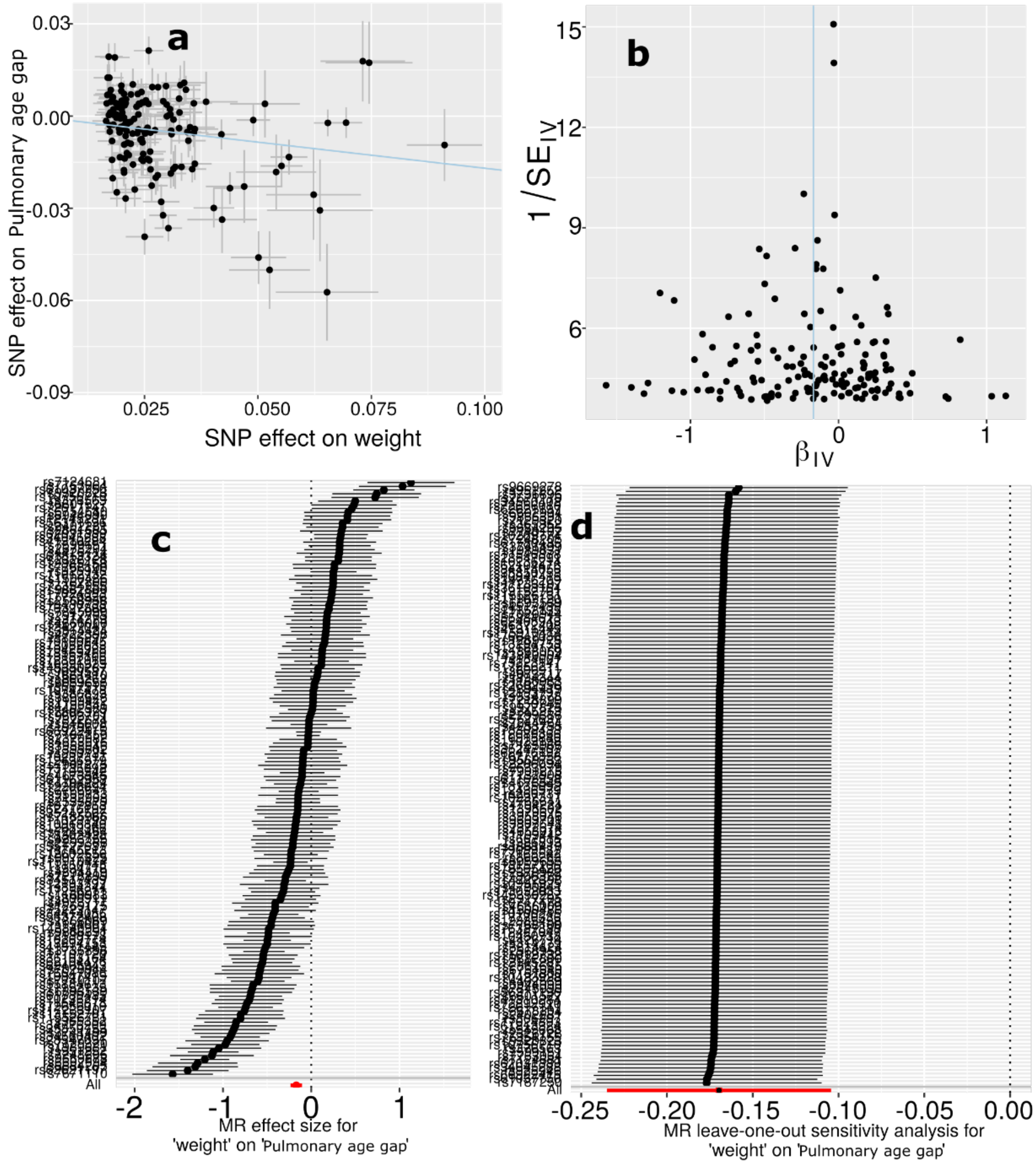

**a**) Scatter plot for the MR effect sizes of the exposure variable (body weight, *x*-axis, SD units) and the outcome variable (pulmonary BAG, *y*-axis, log OR) with standard error bars. The slopes of the regression line correspond to the causal effect sizes estimated by the IVW estimator. **b**) Funnel plot for the relationship between the causal effect of the exposure variable on the outcome variable. Each dot represents MR effect sizes estimated using each SNP as a separate instrument against the inverse of the standard error of the causal estimate. The vertical red line shows the MR estimates using all SNPs. **c**) Forest plot for the single-SNP MR results. Each line represents the MR effect (log OR) for the exposure variable on the outcome variable using only one SNP; the red line shows the MR effect using all SNPs together. **d**) Leave-one-out analysis of the exposure variable on the outcome variable. Each row represents the MR effect (log OR) and the 95% CI by excluding that SNP from the analysis. The red line depicts the IVW estimator using all SNPs.

**eFigure 32: Mendelian randomization sensitivity check for AD on the renal BAG**

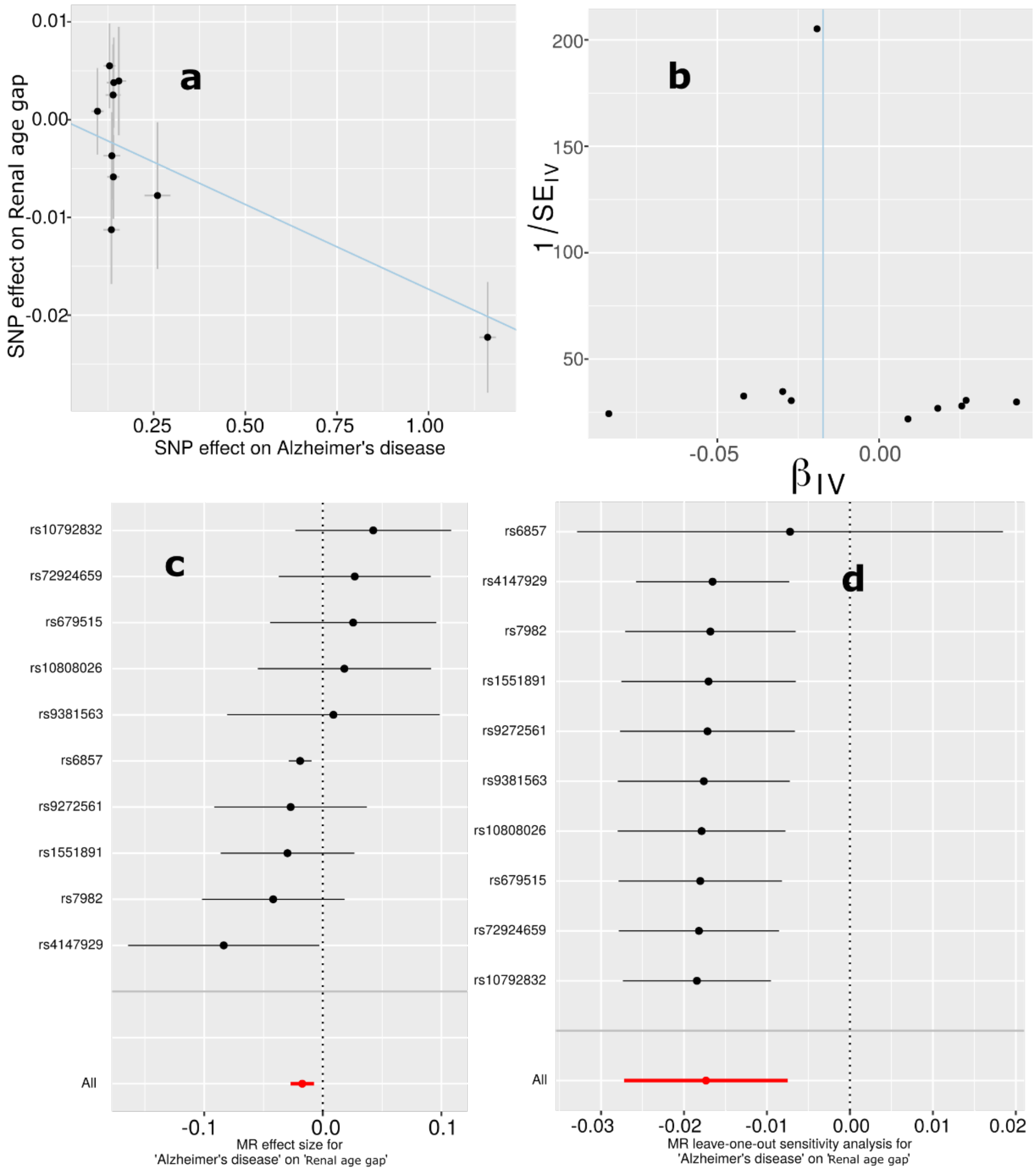

**a**) Scatter plot for the MR effect sizes of the exposure variable (AD, *x*-axis, SD units) and the outcome variable (renal BAG, *y*-axis, log OR) with standard error bars. The slopes of the regression line correspond to the causal effect sizes estimated by the IVW estimator. **b**) Funnel plot for the relationship between the causal effect of the exposure variable on the outcome variable. Each dot represents MR effect sizes estimated using each SNP as a separate instrument against the inverse of the standard error of the causal estimate. The vertical red line shows the MR estimates using all SNPs. **c**) Forest plot for the single-SNP MR results. Each line represents the MR effect (log OR) for the exposure variable on the outcome variable using only one SNP; the red line shows the MR effect using all SNPs together. **d**) Leave-one-out analysis of the exposure variable on the outcome variable. Each row represents the MR effect (log OR) and the 95% CI by excluding that SNP from the analysis. The red line depicts the IVW estimator using all SNPs.

**eFigure 33: Mendelian randomization sensitivity check for weight on the renal BAG**

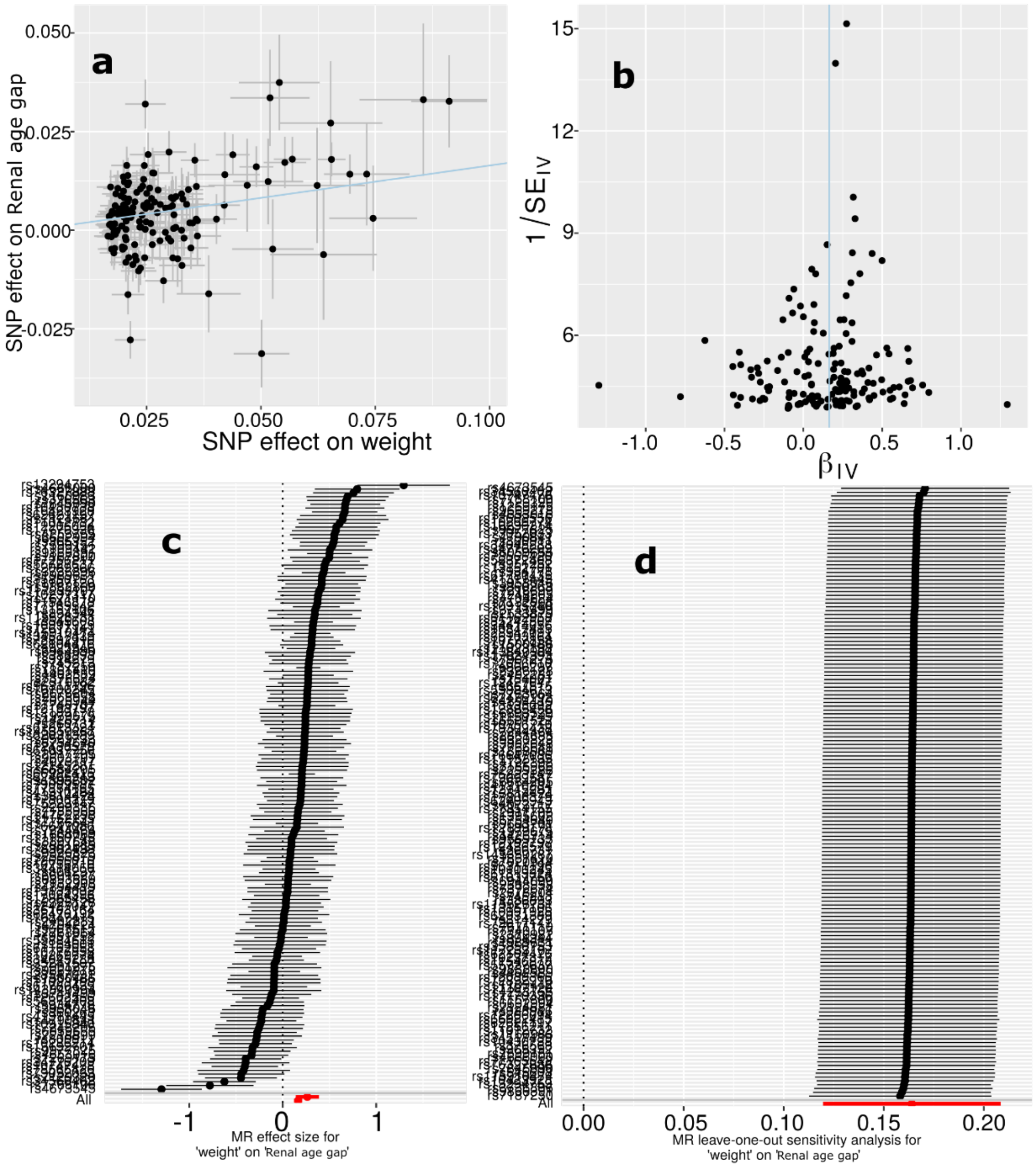

**a**) Scatter plot for the MR effect sizes of the exposure variable (body weight, *x*-axis, SD units) and the outcome variable (renal BAG, *y*-axis, log OR) with standard error bars. The slopes of the regression line correspond to the causal effect sizes estimated by the IVW estimator. **b**) Funnel plot for the relationship between the causal effect of the exposure variable on the outcome variable. Each dot represents MR effect sizes estimated using each SNP as a separate instrument against the inverse of the standard error of the causal estimate. The vertical red line shows the MR estimates using all SNPs. **c**) Forest plot for the single-SNP MR results. Each line represents the MR effect (log OR) for the exposure variable on the outcome variable using only one SNP; the red line shows the MR effect using all SNPs together. **d**) Leave-one-out analysis of the exposure variable on the outcome variable. Each row represents the MR effect (log OR) and the 95% CI by excluding that SNP from the analysis. The red line depicts the IVW estimator using all SNPs.

**eFigure 34: Mendelian randomization sensitivity check for the brain BAG on sleep duration**

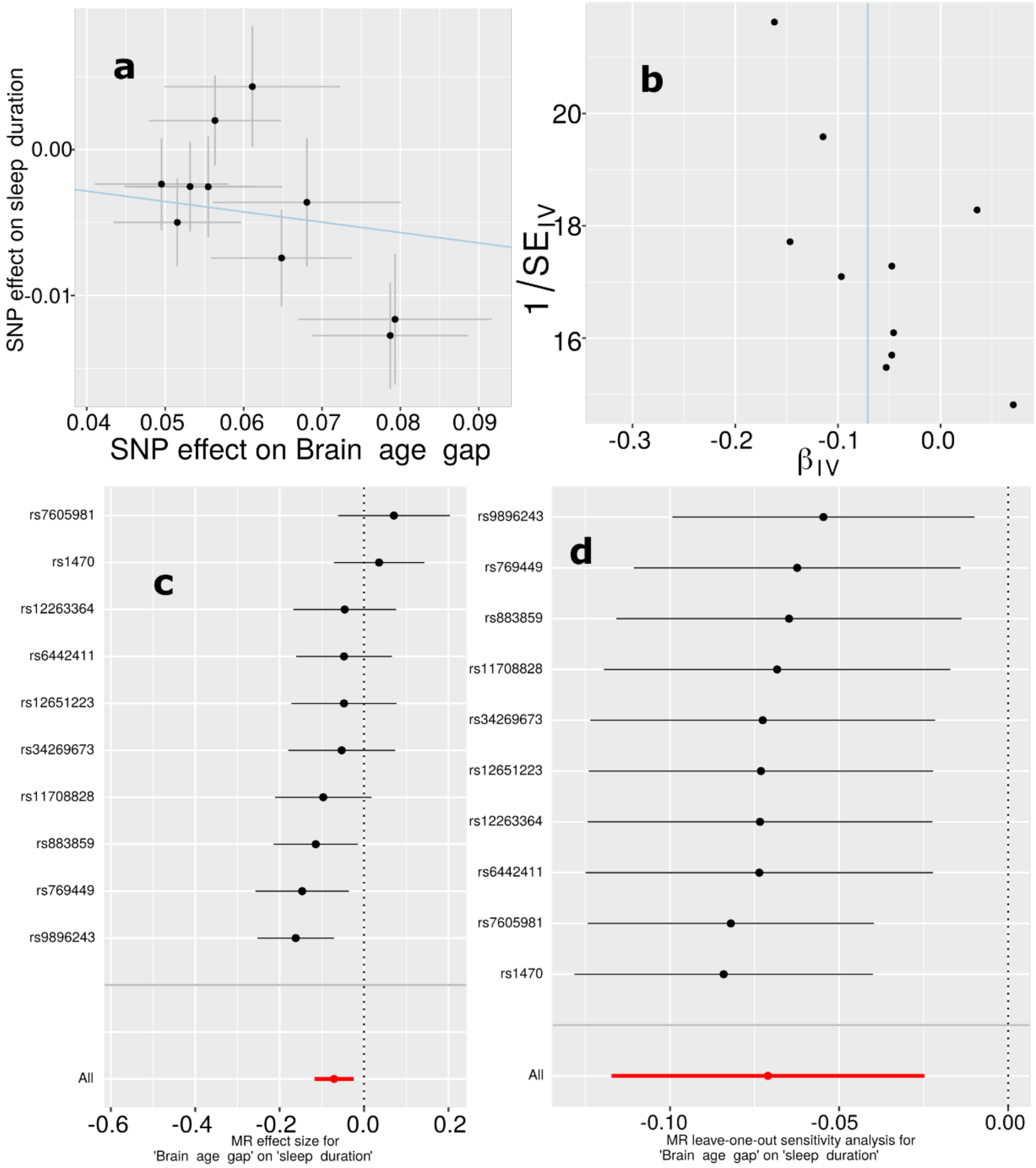

**a**) Scatter plot for the MR effect sizes of the exposure variable (brain BAG, *x*-axis, SD units) and the outcome variable (sleep duration, *y*-axis, log OR) with standard error bars. The slopes of the regression line correspond to the causal effect sizes estimated by the IVW estimator. **b**) Funnel plot for the relationship between the causal effect of the exposure variable on the outcome variable. Each dot represents MR effect sizes estimated using each SNP as a separate instrument against the inverse of the standard error of the causal estimate. The vertical red line shows the MR estimates using all SNPs. **c**) Forest plot for the single-SNP MR results. Each line represents the MR effect (log OR) for the exposure variable on the outcome variable using only one SNP; the red line shows the MR effect using all SNPs together. **d**) Leave-one-out analysis of the exposure variable on the outcome variable. Each row represents the MR effect (log OR) and the

95% CI by excluding that SNP from the analysis. The red line depicts the IVW estimator using all SNPs.

**eFigure 35: Mendelian randomization sensitivity check for the cardiovascular BAG on triglycerides to lipids ratio in very large VLDL**

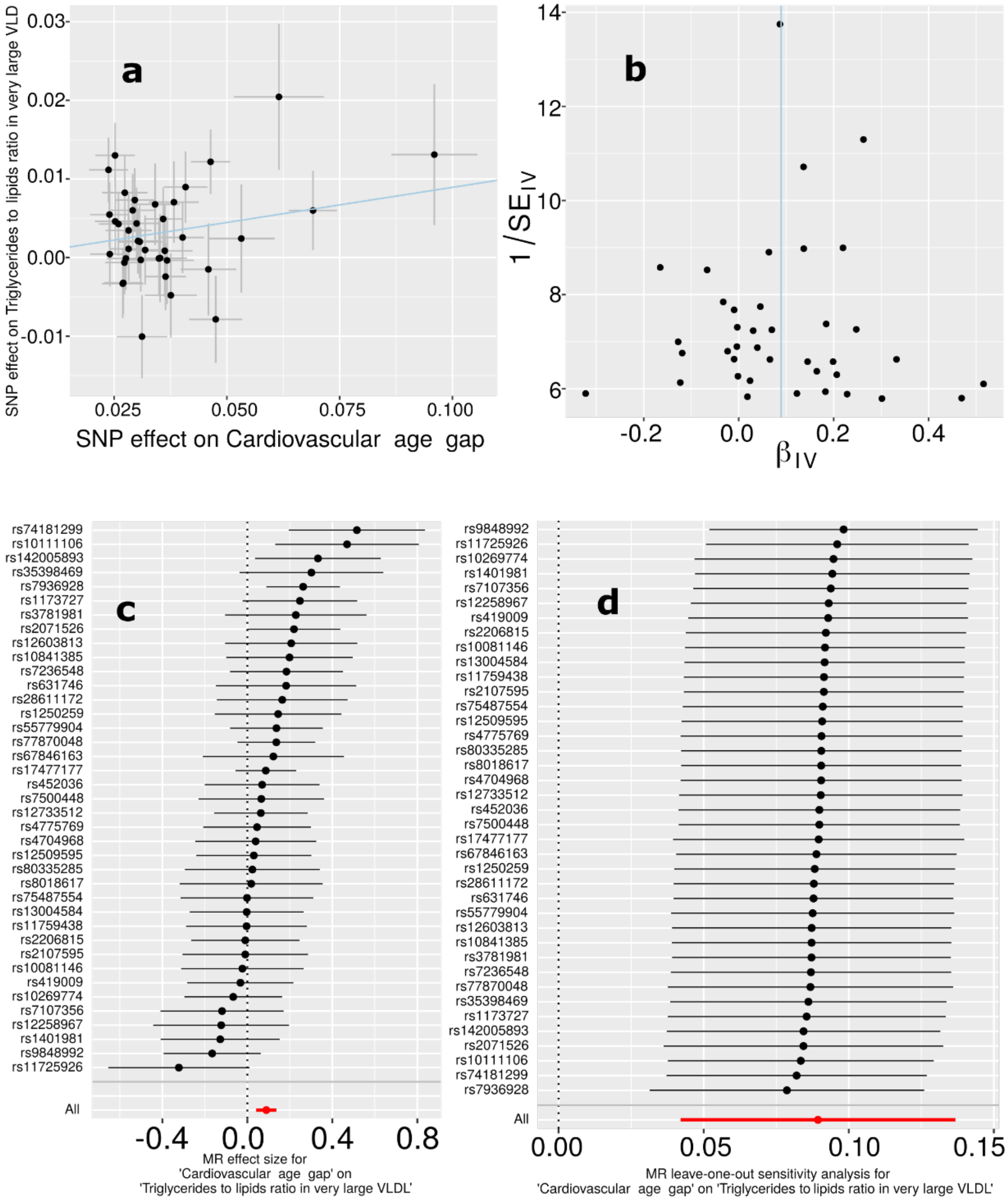

**a**) Scatter plot for the MR effect sizes of the exposure variable (cardiovascular BAG, *x*-axis, SD units) and the outcome variable (triglycerides to lipids ratio in very large VLDL, *y*-axis, log OR) with standard error bars. The slopes of the regression line correspond to the causal effect sizes estimated by the IVW estimator. **b**) Funnel plot for the relationship between the causal effect of the exposure variable on the outcome variable. Each dot represents MR effect sizes estimated using each SNP as a separate instrument against the inverse of the standard error of the causal estimate. The vertical red line shows the MR estimates using all SNPs. **c**) Forest plot for the single-SNP MR results. Each line represents the MR effect (log OR) for the exposure variable on the outcome variable using only one SNP; the red line shows the MR effect using all SNPs together. **d**) Leave-one-out analysis of the exposure variable on the outcome variable. Each row represents the MR effect (log OR) and the 95% CI by excluding that SNP from the analysis. The red line depicts the IVW estimator using all SNPs.

**eFigure 36: Mendelian randomization sensitivity check for the metabolic BAG on weight**

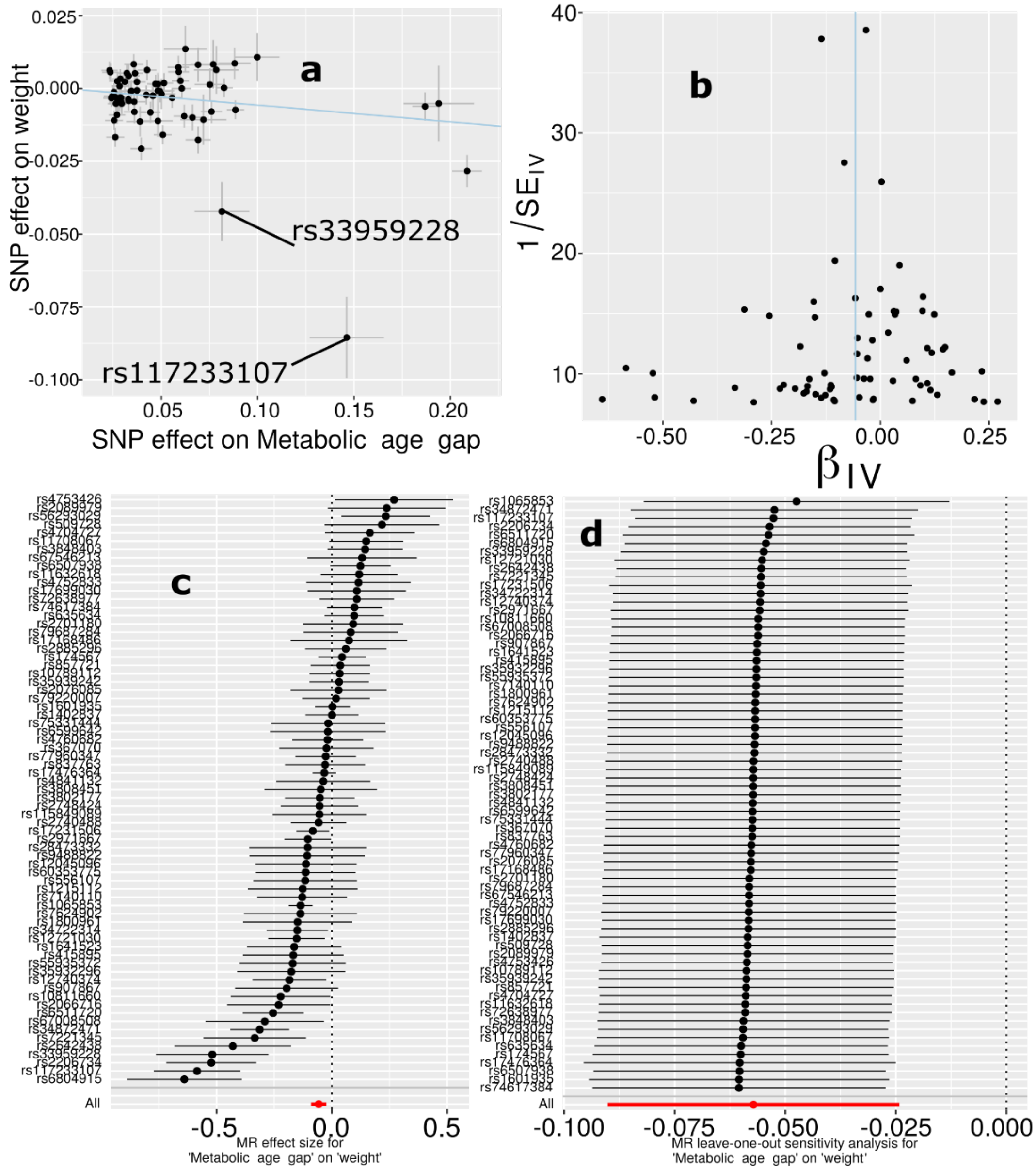

**a**) Scatter plot for the MR effect sizes of the exposure variable (metabolic BAG, *x*-axis, SD units) and the outcome variable (body weight, *y*-axis, log OR) with standard error bars. The slopes of the regression line correspond to the causal effect sizes estimated by the IVW estimator. **b**) Funnel plot for the relationship between the causal effect of the exposure variable on the outcome variable. Each dot represents MR effect sizes estimated using each SNP as a separate instrument against the inverse of the standard error of the causal estimate. The vertical red line shows the MR estimates using all SNPs. **c**) Forest plot for the single-SNP MR results. Each line represents the MR effect (log OR) for the exposure variable on the outcome variable using only one SNP; the red line shows the MR effect using all SNPs together. **d**) Leave-one-out analysis of the exposure variable on the outcome variable. Each row represents the MR effect (log OR) and the 95% CI by excluding that SNP from the analysis. The red line depicts the IVW estimator using all SNPs.

**eFigure 37: Mendelian randomization sensitivity check for the pulmonary BAG on weight**

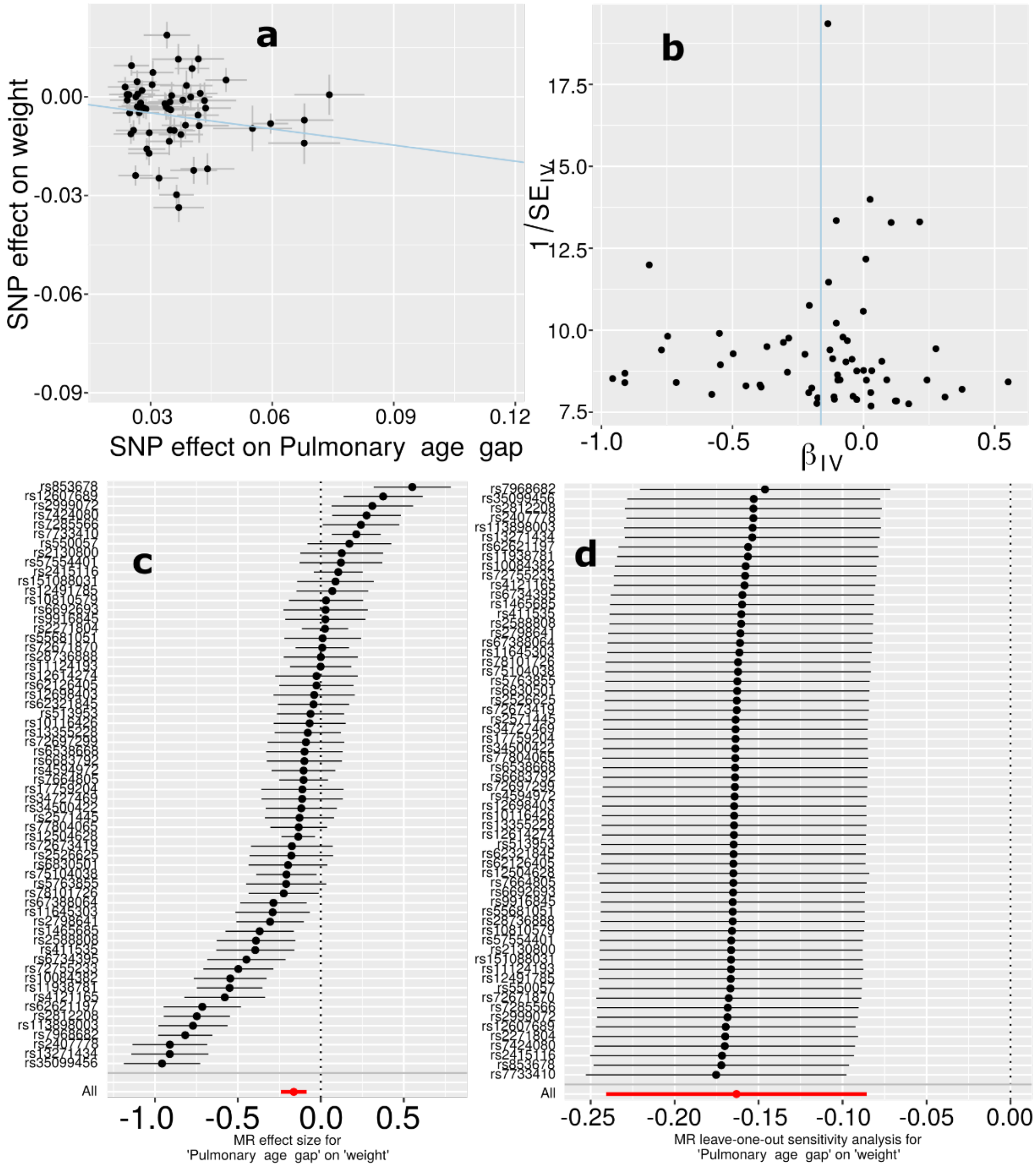

**a**) Scatter plot for the MR effect sizes of the exposure variable (pulmonary BAG, *x*-axis, SD units) and the outcome variable (body weight, *y*-axis, log OR) with standard error bars. The slopes of the regression line correspond to the causal effect sizes estimated by the IVW estimator. **b**) Funnel plot for the relationship between the causal effect of the exposure variable on the outcome variable. Each dot represents MR effect sizes estimated using each SNP as a separate instrument against the inverse of the standard error of the causal estimate. The vertical red line shows the MR estimates using all SNPs. **c**) Forest plot for the single-SNP MR results. Each line represents the MR effect (log OR) for the exposure variable on the outcome variable using only one SNP; the red line shows the MR effect using all SNPs together. **d**) Leave-one-out analysis of the exposure variable on the outcome variable. Each row represents the MR effect (log OR) and the 95% CI by excluding that SNP from the analysis. The red line depicts the IVW estimator using all SNPs.

**eFigure 38: Causal multi-organ network between the nine biological age gaps and 17 clinical traits of chronic diseases, lifestyle factors, and cognition**

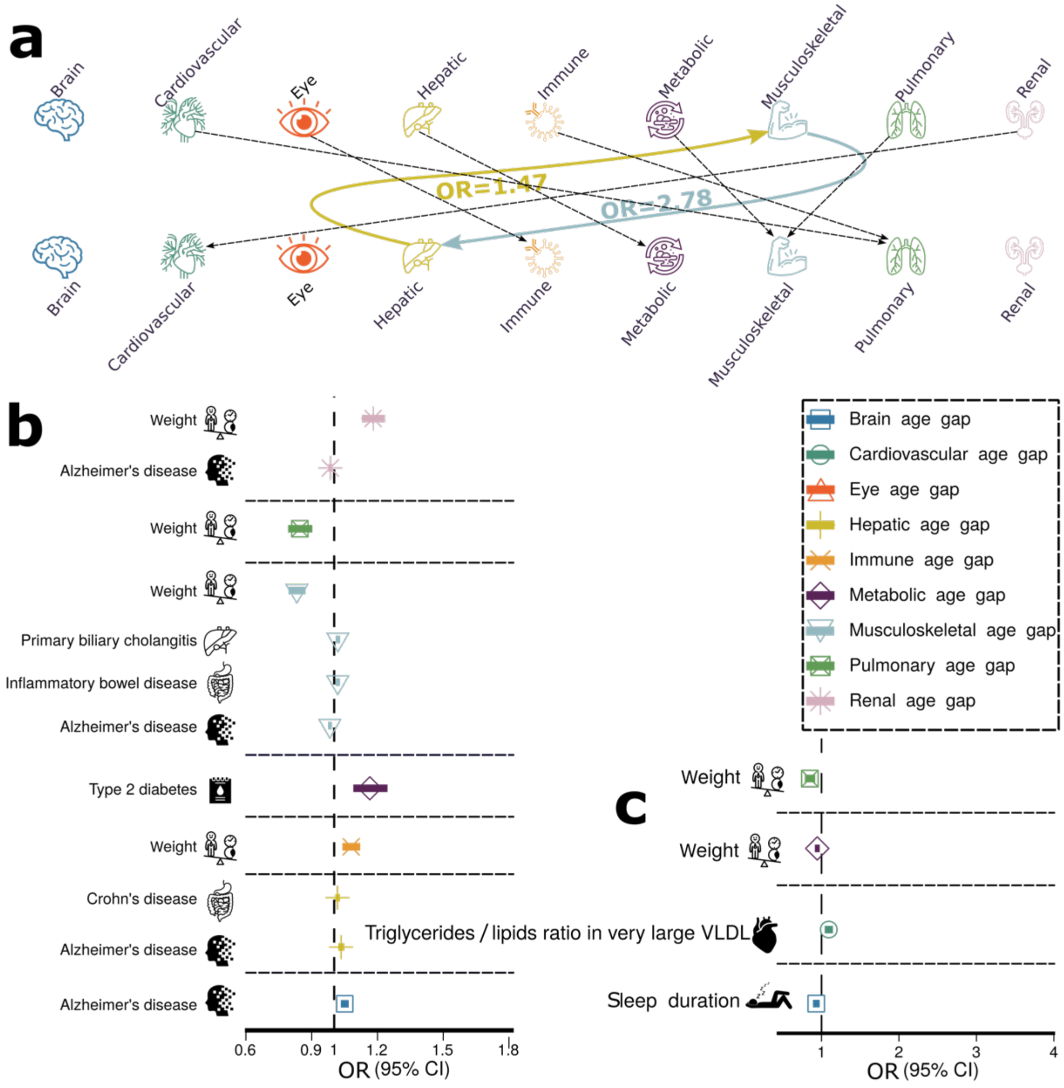

**a**) Causal inference between each pair of BAGs using bi-directional two-sample Mendelian randomization by excluding overlapping populations. The colored lines represent causal effects that survived the correction for multiple comparisons using the Bonferroni method; the dotted lines denote the nominal significant causal effects (P-value < 0.05). **b**) The forward Mendelian randomization investigates the causal inference of 17 unbiasedly selected exposure variables on the nine outcome variables (i.e., the nine BAGs). **c**) The inverse Mendelian randomization examines the causal inference of the 9 BAGs on the 17 clinical traits. We present the tests passing the statistical significance after adjusting for multiple comparisons using the Bonferroni correction. The OR and the 95% confidence interval are presented. Abbreviation: VLDL: very low-density lipoprotein; CI: confidence interval; OR: odds ratio.

**eTable 1: Heritability estimates using the GCTA software**

**A) Original sample sizes.** Original sample sizes were used to estimate the heritability for the nine organ systems.

| **BAG** | $\boldsymbol{h^2}$ | $\boldsymbol{h^2}$ SE | **P-value** | ***N*** |
|---|---|---|---|---|
| Brain | 0.47 | 0.02 | $<1\times10^{-10}$ | 30,108 |
| Cardiovascular | 0.27 | 0.006 | $<1\times10^{-10}$ | 111,543 |
| Eye | 0.38 | 0.02 | $<1\times10^{-10}$ | 36,004 |
| Hepatic | 0.23 | 0.006 | $<1\times10^{-10}$ | 111,543 |
| Immune | 0.20 | 0.004 | $<1\times10^{-10}$ | 111,543 |
| Metabolic | 0.29 | 0.006 | $<1\times10^{-10}$ | 111,543 |
| Musculoskeletal | 0.24 | 0.004 | $<1\times10^{-10}$ | 111,543 |
| Pulmonary | 0.36 | 0.006 | $<1\times10^{-10}$ | 111,543 |
| Renal | 0.30 | 0.006 | $<1\times10^{-10}$ | 111,543 |

**B) Down-sampled sample sizes.** For the eight BAGs except for the brain BAG, we randomly down-sampled the original sample sizes to that of the brain BAG.

| **BAG** | $\boldsymbol{h^2}$ | $\boldsymbol{h^2}$ SE | **P-value** | ***N*** |
|---|---|---|---|---|
| Brain | 0.47 | 0.02 | $<1\times10^{-10}$ | 30,108 |
| Cardiovascular | 0.35 | 0.07 | $<1\times10^{-5}$ | 30,108 |
| Eye | 0.42 | 0.02 | $<1\times10^{-5}$ | 30,108 |
| Hepatic | 0.18 | 0.07 | $<1\times10^{-5}$ | 30,108 |
| Immune | 0.19 | 0.07 | $<1\times10^{-5}$ | 30,108 |
| Metabolic | 0.16 | 0.07 | $<1\times10^{-5}$ | 30,108 |
| Musculoskeletal | 0.21 | 0.07 | $<1\times10^{-5}$ | 30,108 |
| Pulmonary | 0.39 | 0.07 | $<1\times10^{-5}$ | 30,108 |
| Renal | 0.28 | 0.07 | $<1\times10^{-5}$ | 30,108 |

**C) Brain imaging-derived phenotypes vs. 4 pulmonary features.** For the brain imaging phenotypes, we used four sets of features from our previous studies: *i*) 32 pattern of structural coavairance (PSCs) from the data-driven MuSIC atlas using T1-weighted MRI and orthogonal-projective non-negative matrix factorization[3]; *ii*) 101 GM ROIs using the ANTs (https://stnava.github.io/ANTs/) software[4]; *iii*) the 21 WM tracts for fractional anisotropy (FA) mean values[5]; and *iv*) 21 funtional node (FN) measures from resting-state functional MRI[6]. The 4 pulmonary features included forced vital capacity, forced expiratory volume, peak expiratory flow, and the ratio of forced expiratory volume to forced vital capacity. For comparison purposes, we also show the $h^2$estimates for the brain and pulmonary BAGs. The detailed results for all estimates are presented in **Supplementary eFile 22**. The distribution of each phenotype group is shown in the figure below.

| **Organ** | **Phenotype group** | **Phenotype (mean or individual)** | $\boldsymbol{h^2}$ | $\boldsymbol{h^2}$ SE | **P-value** |
|---|---|---|---|---|---|
| Brain | Brain feature | MuSIC[3] | 0.45 | 0.16 | $<1E^{-20}$ |
| | | GM-IDP[4] | 0.39 | 0.16 | $<1E^{-20}$ |
| | | WM-IDP[5] | 0.53 | 0.08 | $<1E^{-20}$ |

| | | | | | |
|---|---|---|---|---|---|
| | | FN-IDP[6] | 0.29 | 0.06 | <1E$^{-20}$ |
| | Brain BAG | Brain BAG | 0.47 | 0.02 | <1E$^{-20}$ |
| Pulmonary | Pulmonary feature | FVC | 0.34 | 0.007 | <1E$^{-20}$ |
| | | FEV/FVC | 0.41 | 0.007 | <1E$^{-20}$ |
| | | PEF | 0.28 | 0.007 | <1E$^{-20}$ |
| | | FEV | 0.35 | 0.007 | <1E$^{-20}$ |
| | Pulmonary BAG | Pulmonary BAG | 0.36 | 0.006 | <1E$^{-20}$ |

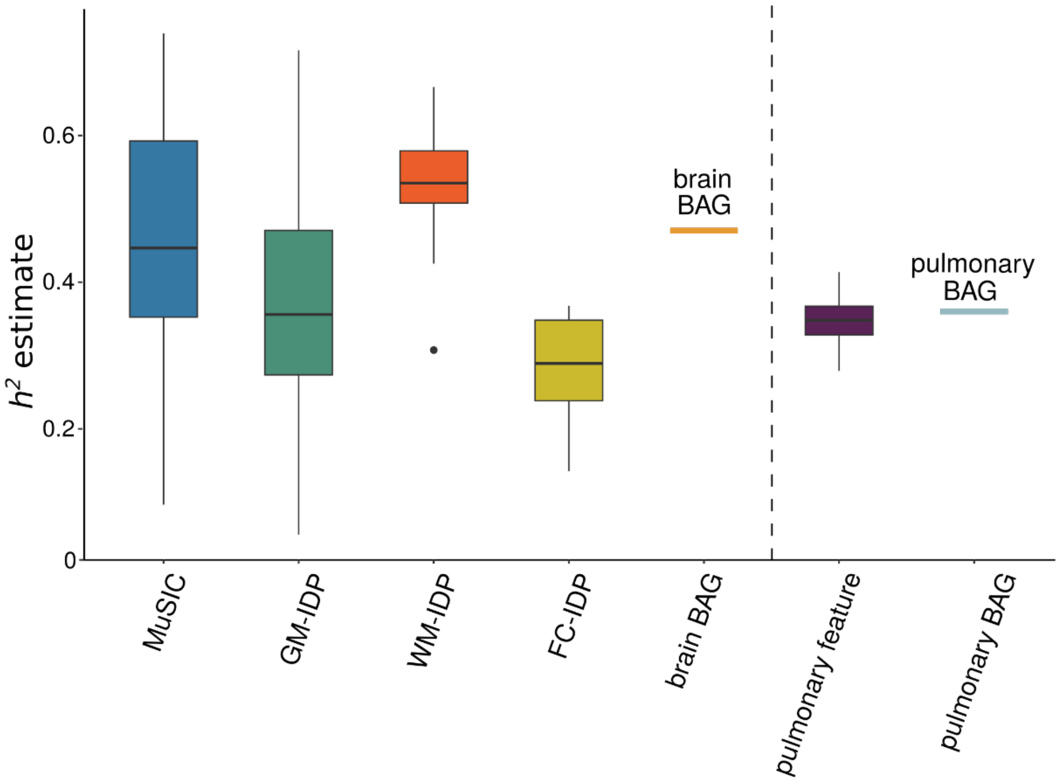

**Figure**. We compared $h^2$ estimates using GCTA between brain features and the brain BAG in contrast to pulmonary features and the pulmonary BAG. In general, our observations indicated that the brain BAG (0.47±0.02) exhibits a higher degree of heritability than the pulmonary BAG (0.36±0.06), and this pattern aligns with the heritability of the underlying features employed in their computation: Brain feature: $h^2$=0.42 across the four sets of brain features vs. pulmonary feature: $h^2$=0.34 across the four pulmonary features.

**eTable 2: The beta coefficient and its SE estimate from the full sample vs. the down-sampled brain BAG comparable sample**

N_ISS: number of independent significant SNPs

| BAG | Mean_beta_downsample | Mean_beta_fullsample | SE_beta_downsample | SE_beta_fullsample | t_beta | p_beta | t_se | p_se | N_ISS |
|---|---|---|---|---|---|---|---|---|---|
| Cardiovascular | 0.034802 | 0.035822 | 0.010533 | 0.005457 | -0.51317 | 0.608293 | 14.0846 | **1.95E-33** | 124 |
| Eye | 0.06527 | 0.064561 | 0.009967 | 0.009128 | 0.136138 | 0.891913 | 1.828485 | **0.069668** | 69 |
| Hepatic | 0.058408 | 0.057479 | 0.014495 | 0.007525 | 0.293471 | 0.769268 | 13.28265 | **2.59E-35** | 289 |
| Immune | 0.043347 | 0.041526 | 0.011454 | 0.005948 | 0.682463 | 0.495312 | 12.78407 | **5.79E-32** | 217 |
| Metabolic | 0.053834 | 0.052587 | 0.013227 | 0.006842 | 0.490113 | 0.624182 | 15.99737 | **1.7E-50** | 422 |
| Musculoskeletal | 0.04263 | 0.041015 | 0.011109 | 0.005817 | 0.520949 | 0.602797 | 11.23119 | **1.44E-24** | 147 |
| Pulmonary | 0.035423 | 0.036056 | 0.010959 | 0.005678 | -0.53629 | 0.591975 | 20.08143 | **1.81E-67** | 272 |
| Renal | 0.067828 | 0.068927 | 0.014536 | 0.007595 | -0.2335 | 0.815446 | 12.87744 | **5.18E-34** | 331 |

**eTable 3: Genetic correlation analyses between the pulmonary BAG and the four features used to derive the BAG**.

| BAG | Pulmonary feature | $g_c$ mean | $g_c$ std | P |
|---|---|---|---|---|
| Pulmonary_age_gap | forced_vital_capacity_fvc_zscore | 0.6409 | 0.0195 | $6.1\text{E-}^{237}$ |
| | fev1_fvc_ratio_zscore | 0.5371 | 0.0316 | $6.47\text{E-}^{65}$ |
| | peak_expiratory_flow_pef | -0.7903 | 0.0175 | $<1\text{E}^{-300}$ |
| | forced_expiratory_volume_in_1second_fev1_zscore | 0.8259 | 0.0111 | $<1\text{E}^{-300}$ |

**eTable 4: Selected 41 clinical traits for genetic correlation analyses**. We selected the candidate studies from the GWAS Catalog for 41 clinical traits, including chronic diseases affecting multiple organ systems, education, and intelligence. To ensure the suitability of the GWAS summary statistics, we first checked that the selected study's population was European ancestry; we then guaranteed a moderate SNP-based heritability $h^2$ estimate and excluded the studies with spurious low $h^2$ (<0.05). Abbreviations are detailed in the main text.

| Primary organ system | Trait | PubMed ID | Sample size |
|---|---|---|---|
| Brain | AD | 30820047 | 63,926 |
| | Smile-GAN-AD1 | NA | 33,540 |
| | SmileGAN-AD2 | NA | 33,540 |
| | SmileGAN-AD3 | NA | 33,540 |
| | SmileGAN-AD4 | NA | 33,540 |
| | SurrealGAN-AD1 | NA | 33,540 |
| | SurrealGAN-AD2 | NA | 33,540 |
| | ADHD | 30478444 | 53,293 |
| | ALS | 27455348 | 36052 |
| | ASD | 30804558 | 46,350 |
| | HYDRA-ASD1 | 37017948 | 14,786 |
| | HYDRA-ASD2 | 37017948 | 14,786 |
| | HYDRA-ASD3 | 37017948 | 14,786 |
| | BIP | 31043756 | 51,710 |
| | MDD | 22472876 | 18,759 |
| | HYDRA-MDD1 | NA | 33,540 |
| | HYDRA-MDD2 | NA | 33,540 |
| | SCZ | 23974872 | 11,244 |
| | HYDRA-SCZ1 | 32103250 | 14,786 |
| | HYDRA-SCZ2 | 32103250 | 14,786 |
| | OCD | 28761083 | 9,725 |
| Cardiovascular | WMH | 31551276 | 11,226 |
| | AF | 30061737 | 1030,836 |
| | Stroke | 29531354 | 446,696 |
| Eye | Glaucoma | 33627673 | 330,905 |
| Hepatic | Liver fat | 34128465 | 32,858 |
| | PBC | 34033851 | 24,510 |
| Immune | SLE | 26502338 | 14,267 |
| | HIV | 34737426 | 208,808 |
| Metabolic | DB | 30054458 | 655,666 |
| | Hyperlipidemia | 34906840 | 349,222 |
| Musculoskeletal | RA | 36333501 | 92,044 |
| Pulmonary | Lung carcinoma | 28604730 | 85,716 |
| Renal | CKD | 31152163 | 625,219 |
| Digestive | CD | 26192919 | 20,883 |
| | IBD | 26192919 | 34652 |
| Breast | Breast cancer | 29059683 | 139,274 |

| | | | |
|---|---|---|---|
| Cognition | Education | 23722424 | 126,559 |
| | Reaction time | 29844566 | 330,069 |
| | Intelligence | 28530673 | 78,308 |
| Lifestyle | Computer use | 32317632 | 408,815 |

**eTable 5: Genetic correlations analyses between the nine BAGs and longevity, household income, and telomere length**. We downloaded the GWAS summary statistics from Deelen et al.[7], which performed two GWASs on longevity based on the 90th survival percentile. For the household income GWAS, we downloaded from Hill et al.[8]. For the telomere length, we used GWAS summary statistics from Codd et al. [9].

| BAG | Trait | $g_c$ mean | $g_c$ std | P | PubMed ID | Sample size |
|---|---|---|---|---|---|---|
| Brain_age_gap | | gc_mean | gc_std | 0.0931 | | |
| Cardiovascular_age_gap | | -0.1588 | 0.0946 | 0.0049 | | |
| Eye_age_gap | | -0.2038 | 0.0725 | 0.0719 | | |
| Hepatic_age_gap | | -0.1657 | 0.0921 | 0.6182 | | |
| Immune_age_gap | Longevity | 0.0495 | 0.0993 | 0.9299 | 31413236 | 36,745 |
| Metabolic_age_gap | | 0.0086 | 0.0979 | 0.7605 | | |
| Musculoskeletal_age_gap | | 0.0328 | 0.1074 | 0.1128 | | |
| Pulmonary_age_gap | | -0.1193 | 0.0752 | 0.0057 | | |
| Renal_age_gap | | -0.197 | 0.0713 | 0.0323 | | |
| Brain_age_gap | | -0.2089 | 0.0403 | $2.2E^{-07}$ | | |
| Cardiovascular_age_gap | | -0.0679 | 0.0356 | 0.0563 | | |
| Eye_age_gap | | -0.066 | 0.0404 | 0.1024 | | |
| Hepatic_age_gap | | -0.1026 | 0.0417 | 0.0138 | | |
| Immune_age_gap | Household income | 0.0028 | 0.0414 | 0.9464 | 31874048 | 286,301 |
| Metabolic_age_gap | | -0.0671 | 0.0389 | 0.0841 | | |
| Musculoskeletal_age_gap | | -0.2867 | 0.0308 | $1.4E^{-20}$ | | |
| Pulmonary_age_gap | | -0.1567 | 0.0286 | $4.4E^{-08}$ | | |
| Renal_age_gap | | -0.0989 | 0.0321 | 0.002 | | |
| Brain_age_gap | | 0.0273 | 0.0506 | 0.5897 | | |
| Cardiovascular_age_gap | | -0.0005 | 0.0038 | 0.9897 | | |
| Eye_age_gap | | -0.0124 | 0.0439 | 0.7769 | | |
| Hepatic_age_gap | | -0.0042 | 0.0306 | 0.9089 | | |
| Immune_age_gap | Telomere length | -0.1338 | 0.0377 | 0.0004 | 34611362 | 472,174 |
| Metabolic_age_gap | | -0.0514 | 0.0393 | 0.1905 | | |
| Musculoskeletal_age_gap | | 0.0045 | 0.0333 | 0.8932 | | |
| Pulmonary_age_gap | | -0.0993 | 0.0331 | 0.0027 | | |
| Renal_age_gap | | -0.029 | 0.0293 | 0.3222 | | |

**eTable 6: Causal analysis using the LCV method**. We performed causal analysis using the LCV method for the bi-directional causality between hepatic and musculoskeletal BAGs, the 9 BAGs and longevity, and the 9 BAGs and telomere length. GCP: genetic causality proportion.

| Trait1 | Trait2 | GCP | GCP_se | P | PubMed ID | Sample size |
|---|---|---|---|---|---|---|
| Musculoskeletal _age_gap | Hepatic_age_gap | -0.75144 | 0.143475 | 9.37E-12 | NA | 111,543 |
| Brain_age_gap | | -0.45597 | 0.208644 | **0.047488** | | |
| Cardiovascular_age_gap | | -0.21694 | 0.395088 | 0.547241 | | |
| Eye_age_gap | | -0.07761 | 0.565366 | 0.639544 | | |
| Hepatic_age_gap | | -0.53253 | 0.321599 | 0.089042 | | |
| Immune_age_gap | Longevity (99$^{th}$ percentile) | -0.15001 | 0.356513 | 0.868225 | 31874048 | 286,301 |
| Musculoskeletal_age_gap | | -0.26633 | 0.440294 | 0.827824 | | |
| Metabolic _age_gap | | -0.3153 | 0.391594 | 0.866896 | | |
| Pulmonary_age_gap | | -0.18056 | 0.375253 | 0.210053 | | |
| Renal_age_gap | | -0.33425 | 0.403767 | 0.573389 | | |
| Brain_age_gap | | -0.05796 | 0.55584 | 0.713688 | | |
| Cardiovascular_age_gap | | -0.32007 | 0.294362 | 0.421771 | | |
| Eye_age_gap | | -0.11877 | 0.49709 | 0.926991 | | |
| Hepatic_age_gap | | -0.00755 | 0.332263 | 0.792948 | | |
| Immune_age_gap | Telomere length | -0.3321 | 0.126005 | **0.002502** | 34611362 | 472,174 |
| Metabolic_age_gap | | -0.07943 | 0.45872 | 0.705827 | | |
| Musculoskeletal_age_gap | | -0.15992 | 0.478106 | 0.821179 | | |
| Pulmonary_age_gap | | -0.67193 | 0.198345 | **3.57E-16** | | |
| Renal_age_gap | | -0.17496 | 0.500093 | 0.6767 | | |

**eTable 7: Selected 17 clinical traits for Mendelian randomization analyses**. We unbiasedly and systematically selected 17 clinical traits, including chronic diseases affecting multiple organ systems, cognition, and lifestyle factors. The selection procedure is detailed in the main text (**Method 2J**).

| Primary organ system | Trait | PubMed ID | IEU-ID (If applicable) | Number of IVs (forward MR) |
|---|---|---|---|---|
| Brain | AD | 24162737 | ebi-a-GCST002245 | 10 |
| | BIP | 31043756 | ieu-a-1126 | 12 |
| Metabolic | Type 2 diabetes | 22885922 | ieu-a-26 | 10 |
| | Triglyceride-to-lipid ratio | 32114887 | met-d-XL_VLDL_TG_pct | 41 |
| Eye | Glaucoma | NA | finn-b-H7_GLAUCOMA | 9 |
| Musculoskeletal | RA | 23143596 | ebi-a-GCST005569 | 11 |
| Hepatic | PBC | 26394269 | ebi-a-GCST003129 | 16 |
| Digestive | CD | 26192919 | ieu-a-12 | 77 |
| | IBD | 23128233 | ieu-a-292 | 81 |
| Breast | Breast cancer | 29059683 | ieu-a-1126 | 86 |
| Cognition | Reaction time | NA | Local-UKBB | 18 |
| Lifestyle | Coffee intake | NA | Local-UKBB | 11 |
| | Fresh fruit | NA | Local-UKBB | 15 |
| | Tea intake | NA | Local-UKBB | 12 |
| | Sleep duration | NA | Local-UKBB | 8 |
| | Summer outdoor activity hour | NA | Local-UKBB | 14 |
| | Body weight | NA | Local-UKBB | 161 |
